# Explaining socioeconomic inequalities in antibiotic prescribing for common infections in English primary care: a population-based study

**DOI:** 10.64898/2026.05.26.26354118

**Authors:** Murong Yang, Nam Vinh Nguyen, A. Sarah Walker, Julie V. Robotham, Edwin van Leeuwen, Gail Hayward, Christopher C. Butler, Koen B. Pouwels

## Abstract

**OBJECTIVES:** To quantify socioeconomic inequalities in antibiotic prescribing for common infections in primary care, and assess whether these inequalities arise from differences in consultation frequency, prescribing behaviour, or variation in vaccination uptake, smoking, and body mass index.

**DESIGN:** Population based cohort study.

**SETTING:** Primary care data from Clinical Practice Research Datalink, England.

**PARTICIPANTS:** 17,195,399 children and adults estimated to have been registered with a general practice in 2019.

**MAIN OUTCOME MEASURES:** Antibiotic prescribing rates (prescriptions per person-year), consultation rates (consultations per person-year), and probability of receiving an antibiotic prescription following consultation.

**RESULTS:** Higher deprivation was associated with higher antibiotic prescribing rates for most respiratory tract indications. In children, prescribing rates were 44.8% (95% confidence interval [CI] 41.9% to 47.7%) higher for upper respiratory tract infections and 47.6% (95% CI 44.2% to 51.3%) higher for lower respiratory tract infections in the most versus least deprived twentile. In adults, prescribing rates for lower respiratory tract infections were 22.7% (95% CI 21.4% to 24.1%) higher in the most deprived twentile. Prescribing rates for other indications showed weak, U-shaped, or negative associations with deprivation. Prescribing inequalities were primarily driven by inequalities in consultation rates rather than probability of receiving antibiotics once consulted. Lower influenza vaccination uptake partly accounted for higher consultation rates for respiratory infections among more deprived children, while smoking prevalence contributed to inequalities among adults.

**CONCLUSIONS:** Socioeconomic inequalities in antibiotic prescribing vary by indication type and are largely explained by consultation frequency. Reducing inequalities may require interventions that decrease the need to consult, e.g. improving influenza vaccination coverage in children and reducing smoking among adults, rather than focussing solely on prescribing behaviour.

Summary boxes

**What is already known on this topic:** Overall antibiotic prescribing rates are higher in more deprived areas in the UK and internationally.

The relationship between deprivation and the probability of receiving antibiotics differs across common infections, many of which infrequently require antibiotics.

It remains unclear whether inequalities reflect differences in consultations, prescribing decisions, or underlying risk factors.

**What this study adds:** This study quantifies deprivation-related inequalities in antibiotic prescribing across 17 common indications and explores underlying mechanisms.

Socioeconomic inequalities in antibiotic prescribing varied by indication: higher deprivation was consistently associated with higher antibiotic prescribing rates (and wider variation in prescribing) for most respiratory tract indications, with rates 4% to 151% higher in the most deprived compared with the least deprived.

Inequalities in prescribing were largely explained by differences in consultation frequency, not by the likelihood of receiving antibiotics once a consultation occurred.

Lower vaccine uptake partly accounted for the higher consultation rates for respiratory indications in deprived children, while higher smoking prevalence contributed to inequalities among adults.

## Introduction

Antimicrobial resistance (AMR) is a major global health threat,^1^ largely driven by the overuse of antibiotics.^2^ Evidence from many countries, including the UK, shows that AMR is linked to socioeconomic deprivation, with higher levels of resistance observed in more deprived areas.^3^ ^4^ Recognising this, the UK government’s AMR National Action Plan highlights tackling health inequalities as a core priority, with commitments to improve data collection, develop toolkits, and implement targeted interventions to reduce infection burden and avoidable antimicrobial exposure across deprived groups. ^5^

In the UK, around 80% of antibiotics are prescribed in primary care.^6^ Several studies have reported higher prescribing rates in more deprived areas,^7–9^ but the underlying reasons remain unclear. Given the need for antibiotics to prevent serious outcomes from conditions such as pneumonia and pyelonephritis, and the clear association between antibiotic use and resistance,^2^ ^10^ it is essential to clarify where and why inequalities in antibiotic prescribing arise in order to inform strategies that effectively reduce disparities in antibiotic exposure and outcomes.

One possibility is that patients from more deprived areas consult more frequently,^11^ potentially due to differences in underlying infection prevalence or health-seeking behaviour across deprivation levels; another is that general practitioners are more likely to prescribe antibiotics to these patients once they consult with an infection.^12^ Understanding the relative contribution of these drivers is important, as they imply different policy responses: higher consultation rates due to underlying infection prevalence point to preventive interventions that reduce infection burden, while variation in prescribing behaviour highlights the need for targeted antibiotic stewardship to prevent unnecessary development of antimicrobial resistance.

To identify where interventions might be most effective, it is also critical to examine the contribution of potentially modifiable health behaviours (such as vaccination uptake and smoking) and health status indicators (such as body mass index [BMI]), which are unequally distributed across socioeconomic groups ^13–15^ and may contribute to inequalities in infections and antibiotic treatment.^16–18^ These factors may influence not only infection risk but also prescribing decisions, as patients with poorer underlying health are often considered at higher risk of complications. However, their role in consultation and prescribing inequalities is unclear.

Another limitation of existing research is that it typically examines overall antibiotic prescribing, without considering whether associations differ across specific clinical indications. A recent UK study found that patients from more deprived areas had higher odds of receiving antibiotics for otitis externa, but not for urinary tract infections.^19^ This underlines the need for infection-specific analyses that can identify where and why inequalities in antibiotic prescribing occur.

This study addresses these gaps by using English primary care data to quantify socioeconomic inequalities in antibiotic prescribing across 17 infection-related indications in children and adults. It examines whether these inequalities arise from differences in consultation frequency or from the probability of receiving an antibiotic following a consultation for each indication, and evaluates how health-related factors such as smoking, BMI, and vaccination uptake contribute to these differences.

## Methods

### Study design, data sources, and population

We conducted a retrospective, population-based cohort study using data from the Clinical Practice Research Datalink (CPRD) Aurum.^20^ CRPD Aurum contains routinely collected, anonymised primary care data from general practices across the UK. The database includes information on symptoms, diagnoses, tests, prescriptions, health-related behaviours, and socio-demographic factors. Patients in the database are broadly representative of the UK population.^20^ ^21^ As of 2022, CPRD Aurum included approximately 40 million patients from around 1,500 general practices in the UK.^22^

For this study we focussed on England only, using CPRD Aurum records linked to small-area deprivation data at the patient postcode level.^23^ The initial patient cohort included patients of all ages who were registered with an English general practice contributing to CPRD Aurum, whose records met CPRD’s research-quality standard, and who had at least one consultation for one of the 17 infections of interest (defined below) during the 4-year period from 1 January 2016 to 31 December 2019. This restriction reflects the scope of the original data extraction, which was limited to patients with recorded consultations for these conditions during the 4-year period. This period was selected to provide a recent pre-pandemic baseline, avoiding potential temporary disruptions to consultation and prescribing patterns caused by COVID-19.^24^

For analysis, the antibiotic prescribing outcome was restricted to the study period from January to December 2019. We defined the *observed population* as patients from the initial cohort who were registered with a general practice in 2019 (flow chart of sample selection is presented in supplementary fig S1). Individuals without consultation records for the indications during 2016 to 2019 did not contribute directly to the analysis prescribing outcomes, but were accounted for in the analysis denominator as follows, so that the total population modelled represents the general population at the CPRD practices. Specifically, to enable population-level inference beyond consulting patients, we estimated the number and characteristics of registered patients who did not consult for the indications of interest during 2016-2019. We used national data on GP registered patients by English Index of Multiple Deprivation (IMD) twentile, age (in years), sex, and ethnicity,^25^ ^26^ which were key variables in our study, as described below. We stratified by IMD × age × sex × ethnicity and estimated, for each stratification level, the national number of registered patients in 2019 using iterative probability fitting, a standard method to estimate joint distributions that matches the marginal distributions of each variable. We then calculated, for each stratification level, the national mean number of registered patients per practice as the total number of registered patients in that stratification level divided by the total number of national practices. Multiplying this mean by the number of CPRD practices in our study gave the expected registered population that CPRD would represent at each stratification level if it were nationally representative. This expected population was hereafter referred to as the *total population*. The estimated number of non-consulting patients was obtained as the difference between the total population and our CPRD observed population with at least one consultation during 2016-2019. These patients were hereafter referred to as the *non-consulting population*.

### Variable definition

#### Outcomes

We included the following common infection-related indications for which an antibiotic can be prescribed: asthma exacerbation, chronic obstructive pulmonary disease (COPD) exacerbation, acute bronchitis, acute cough, acute sore throat, acute otitis media, acute rhinosinusitis, unspecified lower respiratory tract infection, unspecified upper respiratory tract infection, pneumonia, acute gastroenteritis, cellulitis, impetigo, acute lower urinary tract infection (UTI), pyelonephritis, and prostatitis. Each condition was identified using SNOMED CT, Read, and EMIS codes recorded in CPRD Aurum.^27^ For analysis, due to lower numbers and common antibiotic prescribing recommendations we grouped sore throat, rhinosinusitis and undefined upper respiratory tract infections as “all upper respiratory tract infections” (All URTI), and bronchitis, pneumonia, and undefined lower respiratory tract infection as “all lower respiratory tract infections” (All LRTI). Upper and lower respiratory infections were analysed both as grouped “respiratory infections” and individually as upper or lower respiratory infections. The 17 indications included in the analyses are summarised in table 1.

**Table 1.** Indications and inequality contributors among children and adults.

| Indication category | Diagnosis | Contributors |  |
| --- | --- | --- | --- |
|  |  | Children | Adults |
| Respiratory | Chronic obstructive pulmonary disease exacerbation | NA | Influenza and pneumococcal vaccination, smoking, body mass index |
|  | Asthma exacerbation | Influenza and pneumococcal vaccination |  |
|  | Cough |  |  |
|  | Upper respiratory tract infections (sore throat, unspecified upper respiratory tract infection, rhinosinusitis) |  |  |
|  | Lower respiratory tract infections (bronchitis, unspecified lower respiratory tract infection, pneumonia) |  |  |
| Skin | Cellulitis | - | Smoking, body mass index |
|  | Impetigo | - | Smoking, body mass index |
| Ear | Otitis externa | - | Smoking, body mass index |
|  | Acute otitis media | Pneumococcal vaccination | NA |
| Gastrointestinal | Gastroenteritis | - | Smoking, body mass index |
| Urinary | Urinary tract infection (female only) | - | Smoking, body mass index |
|  | Pyelonephritis (female only) | NA | Smoking, body mass index |
|  | Prostatitis (male only) | NA | Smoking, body mass index |

NA indication not explored for children/adults because of low incidence in that group; - no additional contributor explored for the indication (deprivation included in all analyses). The rationale for not including BMI for children is described in supplementary methods S1.1 (Body Mass Index).

We derived three outcomes for each indication. The primary outcome was the total number of systemic antibiotic prescriptions for that indication per patient in 2019. Prescriptions were counted as separate prescriptions only if issued more than 30 days after a previous prescription for the same indication among the 17 indications summarised in table 1, thereby aiming to exclude repeat prescriptions for the same infection episode. The second outcome was the number of general practice consultations for each indication per patient in 2019, applying the same 30-day rule to distinguish indication episodes. The third outcome was whether a patient received a systemic antibiotic prescription during a consultation for the specific indication among the 17 indications or not.

#### Socioeconomic status

Socioeconomic status was measured using the English IMD 2019, calculated at the lower layer super output area level - a geographical unit with an average population of 1,500 residents used in the UK census.^28^ The IMD is a composite measure covering seven domains: income, employment, education, health, crime, housing, and living environment.^29^ IMD is a measure of relative deprivation ranking areas from the least to the most deprived and was linked to CPRD through the patient postcodes.^23^

In our dataset, IMD was provided as a twentile variable, with 1 representing the least deprived and 20 representing the most deprived areas. For descriptive analyses, we grouped the twentiles into quintiles, with the first quintile representing individuals living in the least deprived areas and the fifth quintile representing those in the most deprived areas. For regression analysis, we used the full twentile variable to capture finer differences in deprivation.

#### Other explanatory variables

Model adjustment included age (in years), sex, and ethnicity in all models. We also variably included vaccination status, smoking, and body mass index (BMI) as key explanatory variables depending on indication (table 1) to assess their contribution to socioeconomic inequalities in antibiotic prescribing.

We considered two vaccines relevant to the common infection-related indications studied: seasonal influenza and pneumococcal. Observed patients were defined as vaccinated for influenza if they received the vaccine between July 2018 and strictly before their first indication-related consultation in 2019, or, for those without an indication-related consultation in 2019, if they received the vaccine between July 2018 and December 2019. This definition accounts for the seasonal timing and duration of vaccine protection. Pneumococcal vaccination was defined as receipt of at least one dose of the vaccine at any time before their first indication-related consultation in 2019. For patients without an indication-related consultation in 2019, vaccination status was defined based on any recorded receipt.

Smoking status and BMI were defined using the most recent record strictly before the observed patient’s first indication-related consultation in 2019. For observed patients without an indication-related consultation in 2019, the most recent recorded value was used. Smoking status was classified as current smoker, ex-smoker, or non-smoker. BMI for adults was calculated as weight (kg) divided by height squared (m^2^), using the most recent values. Values below 5 or above 200 were considered outliers and excluded from the analysis.^30^

All explanatory variables were measured strictly before the observed patient’s first indication-related consultation in 2019 where applicable to reduce bias from reverse causality, e.g. changes in health-related behaviours or measurements that may occur after the onset of infection.

For non-consulting patients whose vaccination status, smoking status, and BMI were not observed, these variables were estimated using official national data for England (see supplementary methods S1.1 for data sources and details). Data were selected from the time periods consistent with, or closest available to, the study period. We first estimated the national-level prevalence of influenza vaccine uptake and pneumococcal vaccine coverage by age and IMD; smoking prevalence (current, ex-, and non-smoker) by age, IMD, sex, and ethnicity; and mean BMI values by age, IMD, sex, and ethnicity. We then generated individual-level values for the non-consulting population using a stochastic imputation approach. Vaccination status was imputed using Bernoulli distribution with success probability equalling to the corresponding stratum-specific prevalence. Smoking status was assigned using cumulative stratum-specific probabilities based on a single uniform random draw. BMI values were generated by sampling from a truncated normal distribution, parameterised by stratum-specific means and standard deviations. Random number generation was seeded to ensure reproducibility.

### Statistical analysis

Analyses were conducted separately for children (<18 years) and adults (≥18 years) (table 1) using the total population, which included both observed population and estimated non-consulting population. We first summarised study population characteristics by IMD quintile, reporting medians (interquartile range) for continuous variables and percentages for categorical variables. We then calculated age-standardised antibiotic prescription rates for each infection by IMD quintile within children and adults using age-specific rates and the UK standard population.^31^ For each quintile, rates were calculated by dividing the total number of antibiotic prescriptions by the total person-time at risk. Person-time at risk was defined as the total period each patient was registered with a CPRD practice within the study period, during which individuals were eligible to contribute a new infection episode, excluding the 30-day period following each consultation. Non-consulting patients were assigned person-time equal to one person-year per patient for both antibiotic prescription and consultation. Consulting patients contributed approximately one person-year of time at risk for each outcome (mean 1.0 person-years, SD 0.01), indicating comparable follow-up time between groups.

We used a distributional regression approach within the Generalised Additive Models for Location, Scale, and Shape (GAMLSS) framework to investigate the relationship between deprivation and infection-related outcomes.^32^ ^33^ Conventional regression models examine only the association between deprivation and the mean of an outcome, ignoring potential differences in other aspects of the outcome distribution, such as variability across deprivation areas.^34^ GAMLSS overcomes this by allowing up to four parameters of the outcome distribution - location (mu), scale (sigma), and shape (nu and tau) - to depend on explanatory variables via link functions.^32^

Following the recommendations by Stasinopoulos, et al. ^35^, we specified a negative binomial type I distribution for count outcomes (number of antibiotic prescriptions and consultations), modelling location and scale parameters to estimate the mean and variance of the outcome distribution across IMD levels. All count models included person-time at risk as an offset term. Mean estimates represent the expected annual prescribing rates, while variance estimates describe heterogeneity across patients. More details of the GAMLSS models are provided in the supplementary methods S1.2. For the binary outcome (whether the patient received an antibiotic prescription once consulted), we fitted logistic regression models using all consultations, with standard errors clustered at the patient level to account for repeated consultations within patients. Predicted probability of receiving antibiotics across IMD levels were estimated from the fitted models. To account for potential non-linear relationships between deprivation and outcomes, we used B-spline basis functions for IMD in the model specification.

Our primary analyses examined socioeconomic inequalities in antibiotic prescribing. We first estimated the crude association between IMD and antibiotic prescribing rate. Age, sex, and ethnicity were then added to form the baseline model. We quantified socioeconomic inequalities in antibiotic prescribing by comparing the most (IMD=20) and least deprived areas (IMD=1). We reported two measures of the deprivation gap: the absolute difference (mean prescribing rate in the most minus least deprived) and percentage difference (absolute difference divided by the mean prescribing rate in the least deprived areas). Uncertainty around the deprivation gaps was quantified using posterior simulation (supplementary methods S1.3). We conducted sex-specific analyses to assess if the deprivation gaps in antibiotic prescribing differed by sex.

To explore potential mechanisms underlying observed inequalities, we examined the association between IMD and (i) consultation rates and (ii) whether a patient received antibiotics once consulted, assessing to what extent differences in inequalities in antibiotic prescribing were driven by inequalities in consultation frequency or prescribing behaviour.

We further explored the potential contributing factors that might explain inequalities in consultation rates and the probability of receiving antibiotic prescriptions per consultation. For children, this included vaccine uptake, which is expected to affect specific indications by targeting causative pathogens, such as influenza vaccination for respiratory tract infections^36^ and pneumococcal vaccination for respiratory tract infections and acute otitis media.^37^ ^38^ For adults, we considered smoking status, BMI, and vaccine uptake (table 1). Each factor was added separately to the baseline model to form adjusted models. BMI was modelled using B-spline basis functions to allow for potential non-linear associations with outcomes. A reduction in the effect size (slope) of IMD on outcomes was interpreted as evidence that the factor may partly explain the observed inequalities (see supplementary methods S1.4 for details).

To quantify the contribution of each factor to socioeconomic inequalities, we compared predicted indication outcomes between the most and least deprived (IMD=20 vs IMD=1) under two scenarios. First, we calculated the observed deprivation gap using the actual distributions of the explanatory factors. Second, we generated predictions for the counterfactual scenario where the distribution of the contributor of interest in the most deprived was replaced with that of the least deprived within the same age, sex, and ethnicity strata, while holding other characteristics constant. The absolute contribution was estimated as the difference between the observed and counterfactual deprivation gaps, and the relative contribution as the absolute contribution divided by the observed gap.

Missing data were present in the covariates in the observed population: 22.8% for IMD, 3.5% for ethnicity, 0.03% for smoking status, and 6.4% for BMI (see supplementary table S10 for missingness by children and adults). We applied multiple imputation with chained equations to create 20 imputed datasets. The imputation model included all covariates used in the subsequent analysis, as well as comorbidities as auxiliary variables. Predictive mean matching was used for continuous variables, whereas logistic regression was applied for binary variables and multinomial logistic regression for categorical variables. Covariate distributions were compared between the complete case and imputed dataset.

To assess whether exclusion of patients who never consulted for the 17 infections influenced results, we performed a sensitivity analysis restricting the study population to observed population only. In addition, we conducted a complete case analysis for observed population to assess the impact of missing data on results. All analyses were conducted in R version 4.4.3 and Stata 14 SE. All corresponding code lists were made available at GitHub.^39^

### Patient and public involvement

Patients and public contributors were involved in shaping the research focus. They highlighted that improving antibiotic use and reducing inequalities requires first understanding the current baseline and key drivers of variation. This perspective informed our study design and the emphasis on quantifying baseline inequalities to support evaluation of future interventions. Findings will be disseminated to patients and the public through news media.

## Results

### Participant characteristics

The analysis included a total population of 3,640,884 children and 13,554,515 adults, of whom 54.8% had at least one infection consultation between 2016 and 2019 and were registered with a practice in 2019 (figure S1). Median age and sex distribution were similar across IMD quintiles (table 2). Those from the most deprived quintile had lower influenza vaccination rates for both children and adults, and among adults, they were more likely to be current smokers, and have a higher median BMI compared to those from less deprived areas (table 2).

**Table 2.** Characteristics of total study population by deprivation quintile. Data are median (interquartile range) for continuous variables and % for categorical variables.

| Characteristic | Q1 (least deprived) | Q2 | Q3 | Q4 | Q5 (most deprived) |
| --- | --- | --- | --- | --- | --- |
| **Children** |  |  |  |  |  |
| Age (years) | 8.0 (4.0, 13.0) | 8.0 (4.0, 13.0) | 8.0 (4.0, 13.0) | 8.0 (4.0, 13.0) | 8.0 (4.0, 13.0) |
| Age group |  |  |  |  |  |
| 0-4 | 27.9 | 28.1 | 28.2 | 28.6 | 29.1 |
| 5-11 | 40.3 | 40.3 | 40.5 | 40.7 | 40.0 |
| 12-17 | 31.8 | 31.6 | 31.3 | 30.8 | 30.9 |
| Female | 49.6 | 49.4 | 49.3 | 48.7 | 48.5 |
| Ethnic minority group | 25.6 | 25.9 | 27.2 | 28.8 | 29.2 |
| Influenza vaccination | 29.8 | 27.1 | 24.1 | 21.3 | 20.8 |
| Pneumococcal vaccination | 73.7 | 73.4 | 72.4 | 73.2 | 73.0 |
| N | 687,094 | 696,353 | 714,809 | 754,539 | 788,089 |
| **Adults** |  |  |  |  |  |
| Age (years) | 48.0 (32.0, 63.0) | 47.0 (32.0, 63.0) | 47.0 (32.0, 63.0) | 47.0 (32.0, 62.0) | 46.0 (32.0, 62.0) |
| Age group |  |  |  |  |  |
| 18-64 | 76.5 | 76.9 | 77.5 | 77.9 | 78.2 |
| 65 and above | 23.5 | 23.1 | 22.5 | 22.1 | 21.8 |
| Female | 52.3 | 52.2 | 51.9 | 51.4 | 51.0 |
| Ethnic minority group | 16.1 | 16.4 | 16.9 | 17.6 | 18.0 |
| Influenza vaccination | 25.7 | 25.0 | 24.4 | 24.1 | 23.6 |
| Pneumococcal vaccination | 18.5 | 18.2 | 17.7 | 17.9 | 17.8 |
| Smoking status |  |  |  |  |  |
| Non-smoker | 69.1 | 64.8 | 60.9 | 56.3 | 50.3 |
| Ex-smoker | 17.9 | 19.3 | 20.4 | 20.6 | 20.2 |
| Current smoker | 12.9 | 15.9 | 18.8 | 23.0 | 29.5 |
| Body mass index | 25.9 (23.0, 29.4) | 26.6 (23.3, 30.4) | 26.7 (23.4, 30.5) | 27.1 (23.7, 31.0) | 27.7 (23.9, 32.1) |
| N | 2,603,601 | 2,651,371 | 2,702,466 | 2,801,201 | 2,795,876 |

### Antibiotic prescriptions by deprivation quintile

Among the indications included, the highest age-standardised antibiotic prescribing rates were observed for upper and lower respiratory tract infections, urinary tract infections, and acute cough in both children and adults, and acute otitis media in children (fig 1 and supplementary table S1).

**Fig 1.**
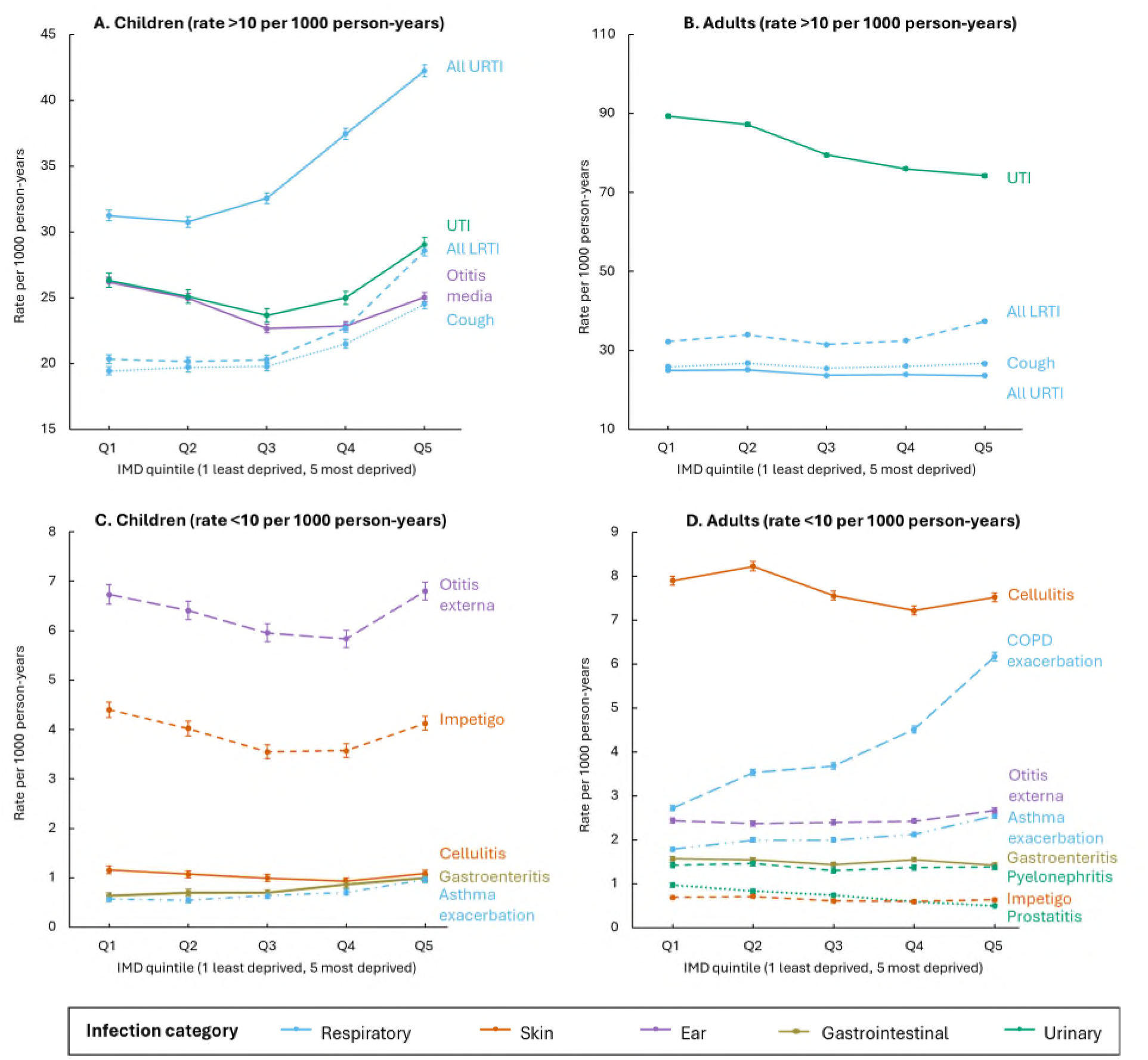
Age-standardised antibiotic prescribing rates (prescriptions per 1000 person-year) for specific infections by deprivation quintile. Error bars represent the 95% confidence intervals. Abbreviations: IMD index of multiple deprivation, URTI upper respiratory tract infections, LRTI lower respiratory tract infections, UTI urinary tract infection, COPD chronic obstructive pulmonary disease. “All URTI” includes sore throat, unspecified upper respiratory tract infection, rhinosinusitis whereas “All LRTI” includes bronchitis, unspecified lower respiratory tract infection, pneumonia. See supplementary fig S2 for prescribing rates of individual respiratory tract infections.

In children, respiratory infections, including upper and lower respiratory tract infections, cough, and asthma exacerbation, showed a clear socioeconomic association, with antibiotic prescribing rates increasing steadily as deprivation level increased (fig 1). This pattern was not observed for urinary tract infections, skin, or ear infections, which had a “U-shaped” association.

Among adults, higher antibiotic prescribing rates in more deprived areas were observed for several respiratory indications, including lower respiratory tract infections, COPD and asthma exacerbations. No clear trend was observed for upper respiratory tract infections, and UTI prescribing was higher in less deprived areas.

### Inequalities in antibiotic prescribing, consultations, and probability of receiving antibiotic prescriptions

In the baseline model adjusted for age, sex, and ethnicity, in children, higher deprivation was associated with higher mean prescribing rates for respiratory indications (excluding pneumonia) and for gastroenteritis (fig 2 and supplementary fig S3). Compared with the least deprived (IMD twentile=1), mean prescribing rates were 44.8% (95% CI 41.9% to 47.7%) higher in the most deprived (IMD twentile=20) for upper respiratory tract infections, 47.6% (95% CI 44.2% to 51.3%) higher for lower respiratory tract infections, and 63.1% (95% CI 43.7% to 82.9%) higher for gastroenteritis (table 3). In contrast, prescribing rates showed a U-shaped relationship with deprivation for impetigo, cellulitis, otitis media, and urinary tract infections, while no significant association was observed for otitis externa.

**Fig 2.**
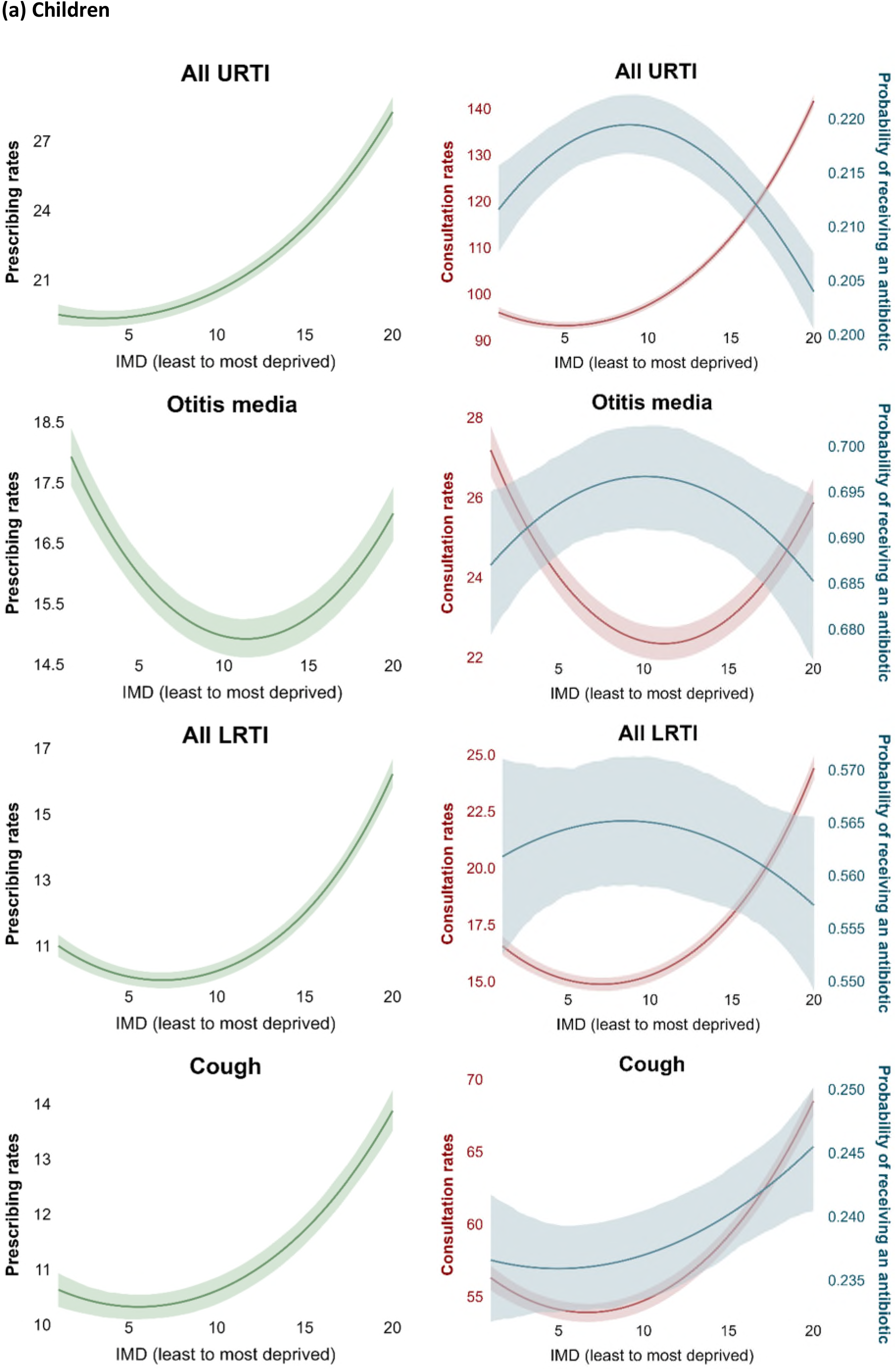
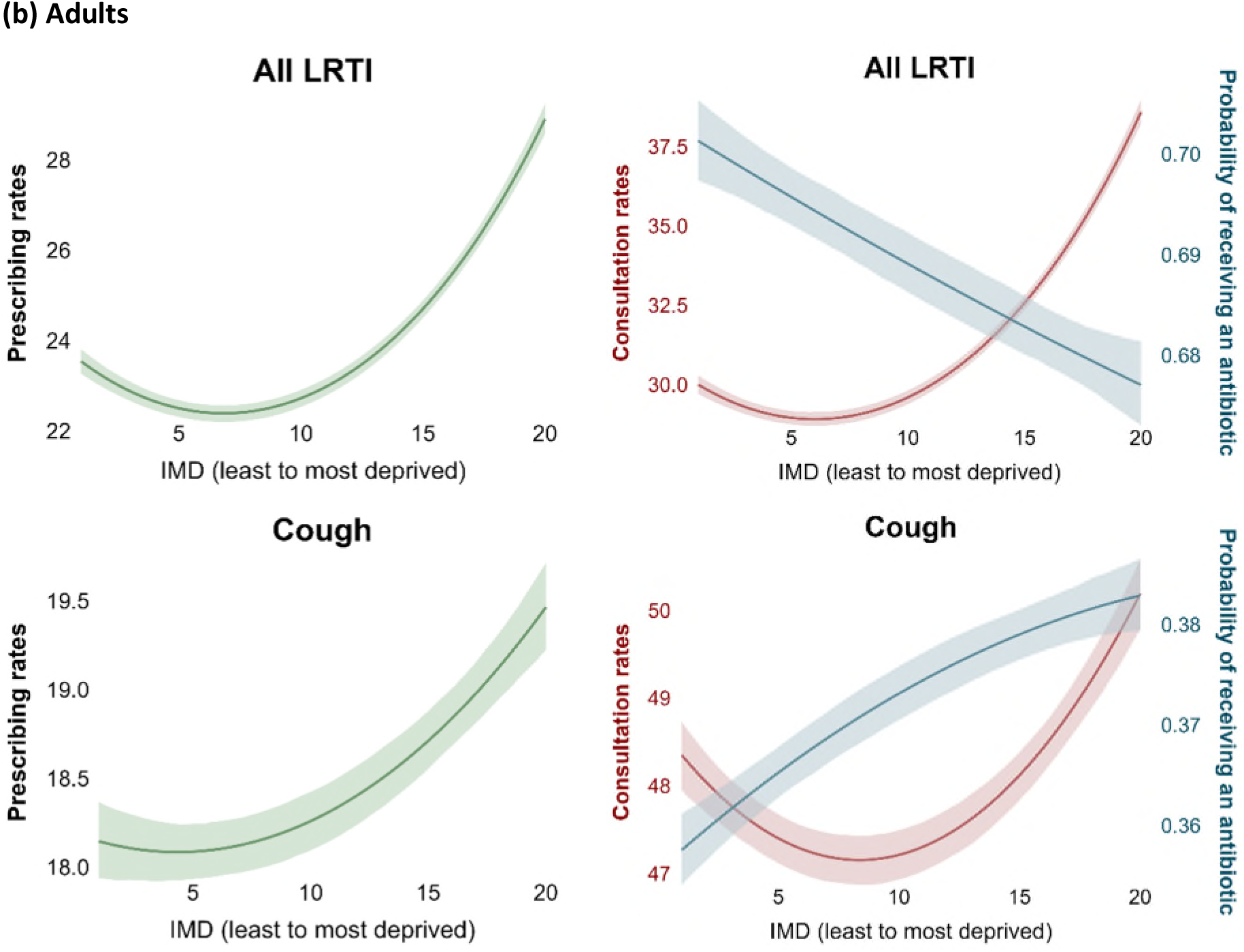
Association between deprivation and antibiotic prescribing rates (prescriptions per 1000 person-years), consultation rates (consultations per 1000 person-years), and probability of receiving antibiotic prescriptions. Estimates represent model-predicted mean measure with shaded 95% confidence intervals. All models were adjusted for age, sex, and ethnicity. Abbreviations: IMD index of multiple deprivation, URTI upper respiratory tract infections, LRTI lower respiratory tract infections. “All URTI” includes sore throat, unspecified upper respiratory tract infection, rhinosinusitis whereas “All LRTI” includes bronchitis, unspecified lower respiratory tract infection, pneumonia.

**Table 3.**
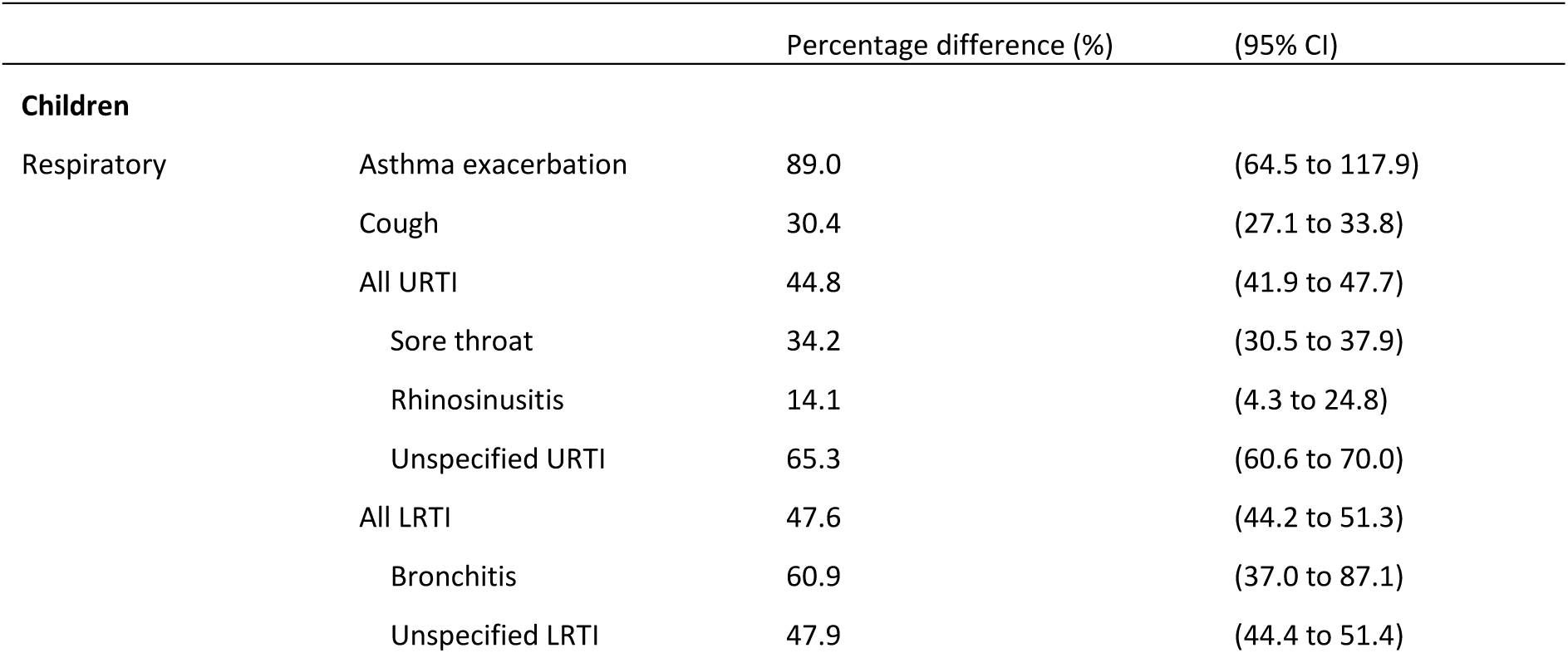
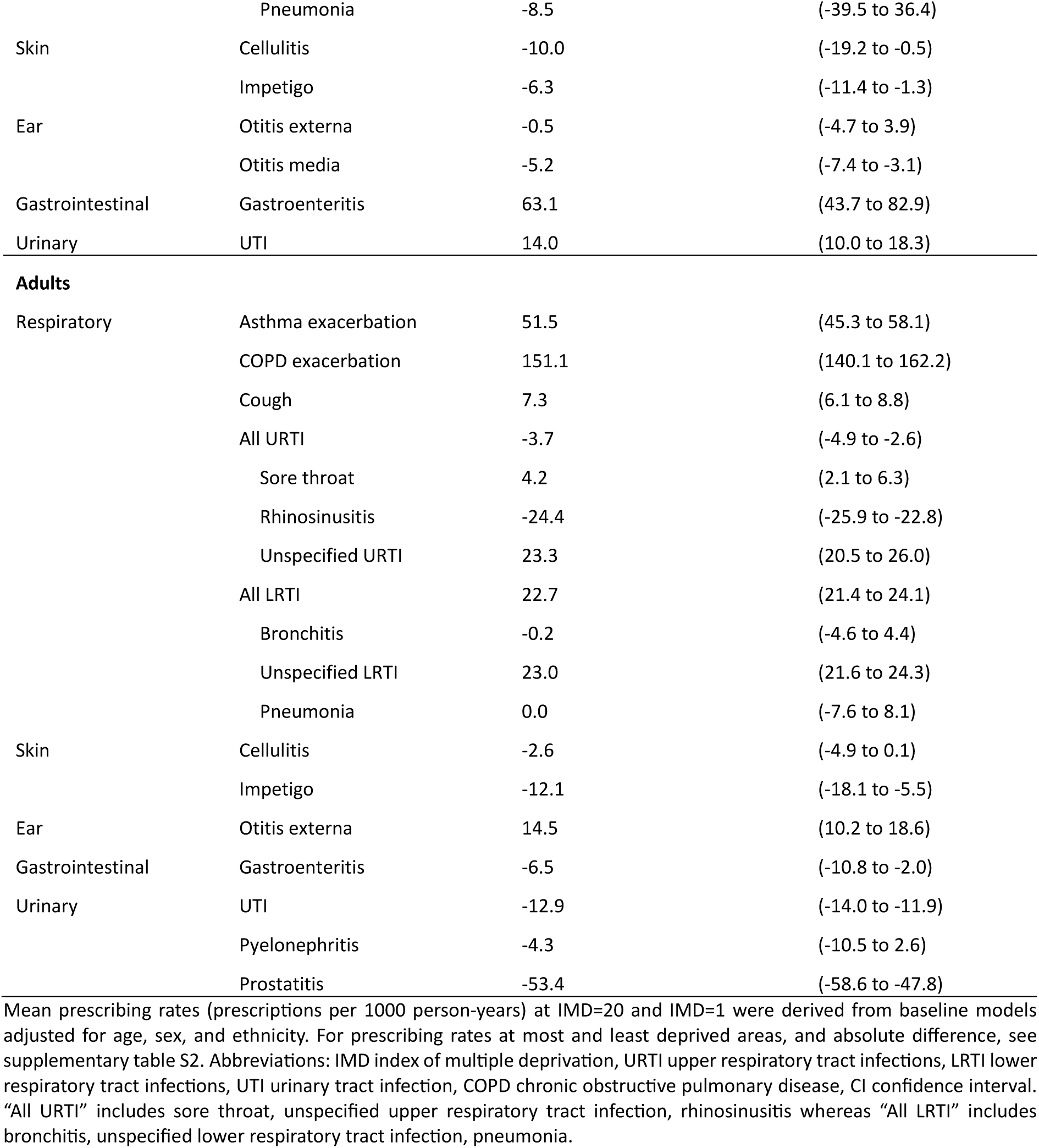
Percentage difference in mean prescribing rates between the most deprived (IMD=20) and least deprived (IMD=1, reference)

Among adults, most respiratory tract indications showed higher prescribing in more deprived areas (fig 2 and supplementary fig S3). Compared with the least deprived, mean prescribing rates in the most deprived were 22.7% (95% CI 21.4% to 24.1%) higher for lower respiratory tract infections and 51.5% (95% CI 45.3% to 58.1%) higher for asthma exacerbation, and notably 151.1% (95% CI 140.1% to 162.2%) higher for COPD exacerbations (table 3). Prescribing for UTI and prostatitis was lower in more deprived areas, while a U-shaped pattern was observed for impetigo and otitis externa. No clear pattern was observed for other infections. Variance estimates followed the same direction as mean estimates, indicating greater heterogeneity in prescribing for infections with higher antibiotic use in more deprived areas (supplementary fig S3). In addition, sex-specific analyses showed that females had consistently larger deprivation gaps than males for most respiratory tract indications, particularly among adults (supplementary tables S3-4).

Patterns of association between deprivation and indication-related outcomes were similar for antibiotic prescribing and consultation rates, but not for the probability of receiving antibiotics once consulted for most indications (fig 2 and supplementary fig S5). For indications with higher prescribing rates that varied by deprivation level (fig 1), higher deprivation was associated with higher consultation rates for upper and lower respiratory tract infections, and cough in children, and for lower respiratory tract infections in adults. These associations were consistent with the patterns observed for antibiotic prescribing rates for the same indications (fig 2). However, the probability of receiving antibiotics once consulted showed either no significant association or an opposite trend (i.e. greater deprivation associated with lower probability of prescribing having attended a consultation), except for cough for both children and adults, where deprivation was positively associated with both prescribing rates and probability, and UTI for adults, where deprivation was negatively associated with both prescribing rates and probability.

### Role of contributors in explaining inequalities

We examined the role of selected contributors in explaining inequalities for infections that had higher prescribing rates in the most deprived compared with the least deprived areas (as shown in Table 3). Among children, we focused on the role of seasonal influenza and pneumococcal vaccination in inequalities for respiratory infections. After adding influenza vaccination and pneumococcal vaccination separately to the baseline model, the association between deprivation and consultation rates became weaker for all respiratory infections, particularly after adjustment for influenza vaccination (supplementary fig S6). This indicates that these factors explained some of the variation attributed to deprivation in the models excluding them, although adjustment for influenza vaccination did not attenuate inequalities in asthma exacerbations.

Compared with the observed deprivation gap based on the actual vaccination distribution, if children in the most deprived areas had the same influenza vaccination coverage as those in the least deprived areas, the deprivation gap in consultation rates would be reduced by 7.1% (95% CI 6.5% to 7.6%) for lower respiratory tract infections, 10.0% (95% CI 9.6% to 10.3%) for upper respiratory tract infections, and 9.5% (95% CI 8.7% to 10.4%) for cough (table 4). Pneumococcal vaccination explained a smaller proportion of inequalities in most respiratory tract indications compared to influenza vaccination (table 4 and supplementary fig S6).

**Table 4.** Contribution of modifiable health factors to consultation rate differences between the most and least deprived.

|  | Δ Observed<br>(95% CI) |  | Δ Counterfactual<br>(95% CI) |  | Relative contribution (%)<br>(95% CI) |  | Δ Observed<br>(95% CI) |  | Δ Counterfactual<br>(95% CI) |  | Relative contribution (%)<br>(95% CI) |  |
| --- | --- | --- | --- | --- | --- | --- | --- | --- | --- | --- | --- | --- |
| Children | Influenza vaccination |  |  |  |  |  | Pneumococcal vaccination |  |  |  |  |  |
| Asthma exacerbation | 1.9 | (1.7 to 2.2) | 2.3 | (2.0 to 2.6) | -19.0 | (-21.6 to -16.8) | 1.9 | (1.7 to 2.2) | 1.9 | (1.6 to 2.1) | 1.6 | (1.3 to 2.0) |
| Cough | 19.4 | (18.1 to 20.6) | 17.5 | (16.3 to 18.8) | 9.5 | (8.7 to 10.4) | 19.6 | (18.4 to 20.8) | 18.9 | (17.7 to 20.1) | 3.7 | (3.3 to 4.1) |
| All URTI | 84.2 | (82.3 to 86.2) | 75.8 | (74.0 to 77.9) | 10.0 | (9.6 to 10.3) | 85.3 | (83.2 to 87.3) | 83.2 | (81.1 to 85.2) | 2.4 | (2.2 to 2.7) |
| Sore throat | 15.4 | (14.5 to 16.3) | 14.1 | (13.3 to 15.0) | 8.3 | (7.7 to 9.0) | 15.6 | (14.7 to 16.5) | 15.0 | (14.1 to 16.0) | 3.8 | (3.4 to 4.1) |
| Rhinosinusitis | 4.1 | (3.7 to 4.5) | 4.0 | (3.6 to 4.3) | 3.7 | (2.9 to 4.5) | 4.2 | (3.9 to 4.6) | 4.0 | (3.6 to 4.3) | 6.7 | (5.9 to 7.5) |
| Unspecified URTI | 70.9 | (69.1 to 72.6) | 63.3 | (61.5 to 65.0) | 10.8 | (10.4 to 11.1) | 72.2 | (70.5 to 74.0) | 70.8 | (69.1 to 72.5) | 2.0 | (1.7 to 2.3) |
| All LRTI | 19.8 | (18.8 to 20.8) | 18.4 | (17.4 to 19.3) | 7.1 | (6.5 to 7.6) | 20.0 | (19.0 to 21.0) | 19.4 | (18.4 to 20.4) | 3.0 | (2.7 to 3.4) |
| Bronchitis | 0.7 | (0.6 to 0.9) | 0.7 | (0.5 to 0.8) | 7.1 | (4.5 to 9.9) | 0.7 | (0.6 to 0.9) | 0.7 | (0.5 to 0.8) | 4.2 | (3.2 to 5.5) |
| Unspecified LRTI | 19.8 | (18.9 to 20.8) | 18.4 | (17.5 to 19.4) | 7.1 | (6.5 to 7.6) | 20.0 | (19.1 to 21.0) | 19.4 | (18.5 to 20.4) | 3.0 | (2.7 to 3.4) |
| Adults | Influenza vaccination |  |  |  |  |  | Pneumococcal vaccination |  |  |  |  |  |
| Asthma exacerbation | 1.8 | (1.7 to 1.9) | 1.8 | (1.7 to 2.0) | -2.9 | (-3.2 to -2.6) | 1.7 | (1.5 to 1.8) | 1.7 | (1.5 to 1.8) | 1.3 | (1.1 to 1.5) |
| COPD exacerbation | 5.8 | (5.6 to 6.1) | 5.7 | (5.4 to 5.9) | 2.7 | (2.5 to 3.0) | 6.1 | (5.8 to 6.3) | 6.0 | (5.8 to 6.3) | 0.4 | (0.3 to 0.5) |
| Cough | 0.1 | (-0.4 to 0.6) | -0.4 | (-0.9 to 0.1) | 167.8 | (-2278.0 to 2787.8) | 0.3 | (-0.2 to 0.9) | 0.3 | (-0.3 to 0.8) | -19.6 | (-226.7 to 214.6) |
| Sore throat | 1.4 | (1.1 to 1.7) | 1.4 | (1.1 to 1.7) | 4.2 | (3.4 to 5.3) | 1.4 | (1.2 to 1.8) | 1.4 | (1.2 to 1.8) | 0.0 | (-0.2 to 0.1) |
| Unspecified URTI | 5.5 | (5.2 to 5.9) | 5.4 | (5.1 to 5.7) | 2.1 | (1.9 to 2.3) | 5.6 | (5.2 to 5.9) | 5.6 | (5.3 to 5.9) | -0.4 | (-0.5 to -0.4) |
| All LRTI | 9.2 | (8.7 to 9.6) | 8.6 | (8.1 to 9.0) | 6.4 | (5.9 to 6.9) | 9.1 | (8.6 to 9.6) | 9.0 | (8.5 to 9.5) | 1.6 | (1.1 to 2.1) |
| Unspecified LRTI | 8.3 | (7.9 to 8.8) | 7.9 | (7.4 to 8.3) | 5.6 | (5.1 to 6.0) | 8.3 | (7.9 to 8.8) | 8.2 | (7.8 to 8.7) | 1.4 | (0.9 to 1.8) |
| Adults | Smoking status |  |  |  |  |  | Body mass index |  |  |  |  |  |
| Asthma exacerbation | 1.8 | (1.6 to 1.9) | 0.7 | (0.5 to 0.8) | 62.6 | (58.5 to 67.4) | 1.7 | (1.6 to 1.9) | 1.2 | (1.0 to 1.3) | 31.8 | (26.0 to 35.1) |
| COPD exacerbation | 7.2 | (6.9 to 7.5) | 1.9 | (1.7 to 2.1) | 73.2 | (71.5 to 75.0) | 6.6 | (6.2 to 7.2) | 5.8 | (5.5 to 6.1) | 12.8 | (10.4 to 18.6) |
| Cough | -0.1 | (-0.6 to 0.4) | -8.3 | (-8.8 to -7.7) | 3605.5 | (-31940.0 to 62234.3) | 0.3 | (-0.3 to 0.9) | -0.1 | (-0.6 to 0.5) | 13.0 | (-1163.9 to 1296.4) |
| Sore throat | 1.5 | (1.2 to 1.8) | 0.5 | (0.2 to 0.8) | 65.0 | (53.5 to 80.3) | 1.4 | (1.1 to 1.7) | 2.1 | (1.8 to 2.4) | -52.2 | (-67.0 to -31.8) |
| Unspecified URTI | 5.7 | (5.3 to 6.0) | 4.0 | (3.7 to 4.3) | 28.7 | (27.1 to 30.7) | 5.5 | (5.2 to 5.9) | 6.1 | (5.7 to 6.4) | -10.2 | (-11.8 to -5.6) |
| All LRTI | 9.4 | (8.9 to 9.9) | 0.6 | (0.2 to 1.1) | 93.5 | (88.8 to 98.0) | 9.7 | (9.2 to 10.2) | 8.3 | (7.8 to 8.8) | 15.0 | (13.3 to 16.4) |
| Unspecified LRTI | 8.6 | (8.2 to 9.0) | 0.6 | (0.2 to 1.1) | 92.7 | (88.0 to 97.1) | 8.8 | (8.3 to 9.3) | 7.5 | (7.1 to 8.0) | 14.1 | (12.8 to 15.2) |
| Otitis externa | 0.3 | (0.1 to 0.5) | -0.4 | (-0.6 to -0.2) | 271.7 | (133.2 to 659.0) | 0.2 | (0.0 to 0.4) | 0.4 | (0.2 to 0.6) | -49.4 | (-203.4 to -11.5) |

Notes: Δ represents the absolute difference in mean consultation rates (consultations per 1000 person-year) between the most deprived (IMD=20) and least deprived (IMD=1, reference), adjusted for age, sex, ethnicity, and the contributor of interest. Δ observed uses the actual contributor distributions, and Δ counterfactual replaces the contributor distribution of the most deprived with that of the least deprived. Relative contribution is calculated as the difference between Δ observed and Δ counterfactual divided by Δ observed (with Δ observed as the reference). Positive values indicate the contributor reduces the deprivation gap, and negative values indicate that the contributor increases the gap. For results on absolute contribution, see supplementary table S5. Abbreviations: IMD index of multiple deprivation, URTI upper respiratory tract infections, LRTI lower respiratory tract infections, UTI urinary tract infection, COPD chronic obstructive pulmonary disease, CI confidence interval. "All URTI" includes sore throat, unspecified upper respiratory tract infection, rhinosinusitis whereas "All LRTI" includes bronchitis, unspecified lower respiratory tract infection, pneumonia.

In adults, smoking status substantially attenuated inequalities in consultations for respiratory infections and otitis externa (supplementary fig S6). Compared with the observed deprivation gap based on actual smoking patterns, if patients in the most deprived areas had the same smoking distribution as those in the least deprived areas, the deprivation gap in consultation rates would be reduced by 93.5% (95% CI 88.8% to 98.0%) for lower respiratory tract infections, 73.2% (95% CI 71.5% to 75.0%) for COPD exacerbations, and 62.6% (95% CI 58.5% to 67.4%) for asthma exacerbations (table 4). Notably, under the observed scenario, which was based on a model further adjusted for smoking status in addition to the baseline model, there was no statistically significant deprivation gap for cough, whereas a deprivation gap was present in the baseline model (supplementary fig S4), indicating that differences in smoking prevalence substantially influence the observed associations with deprivation.

Adding BMI to the baseline model weakened the association between deprivation and consultations in cough and asthma exacerbations (supplementary fig S6). In the counterfactual scenario where BMI distributions in the most deprived areas matched those in the least deprived areas, the deprivation gap would be reduced by 31.8% (95% CI 26.0% to 35.1%) for asthma exacerbations (table 4). However, BMI explained a smaller proportion of inequalities in consultations for other infections than smoking.

## Discussion

### Principal findings

In primary care in England, higher deprivation was associated with higher mean antibiotic prescribing rates for most respiratory tract indications in both children and adults, and for gastroenteritis in children. In contrast, prescribing rates were lower in more deprived areas for UTI and prostatitis in adults, while other indications showed absent or U-shaped associations with deprivation. The variance in prescribing across deprivation followed a similar pattern to the mean, suggesting higher variation in antibiotic prescribing for respiratory tract indications in more deprived areas.

Socioeconomic inequalities in antibiotic prescribing were largely driven by differences in consultations, where patients from more (or less for some infections) deprived areas consulted more often, rather than being more likely to receive antibiotics when consulting. Lower uptake of seasonal influenza and pneumococcal vaccination partly accounted for inequalities in consultation rates for respiratory indications in children, while smoking contributed to inequalities for respiratory indications in adults, with BMI explained them to a lesser extent, thereby driving socioeconomic inequalities in antibiotic prescribing.

### Comparison with other studies

Unlike previous studies that assessed the inequalities in overall antibiotic prescribing,^7–9^ our study extended the evidence by examining inequalities for specific indications, finding substantial heterogeneity in associations between socioeconomic inequalities and antibiotic prescribing across indications, suggesting previous studies may have attenuated or averaged out important effects. Specifically, while most respiratory tract indications consistently showed higher antibiotic prescribing rates in more deprived areas, particularly among females compared with males, other infections showed U-shaped, no associations, or even negative associations.

Furthermore, previous studies focused only on mean prescribing rates, while we also examined variance using a distributional regression approach. For respiratory tract indications, we found that individuals in less deprived areas had lower prescribing levels and less variation, while those from more deprived areas had both higher mean prescribing and higher variation. This provides a fuller picture of inequality, showing that prescribing in deprived areas is not only higher on average but also less consistent. Although our primary focus was on relatively rare outcomes, where most patients had no prescriptions or consultations, resulting in the variance approximating the mean (supplementary figs S3-4), the same GAMLSS-based framework can be applied to more common outcomes. In such contexts, inequalities in the mean and in the variance may diverge. Incorporating variance into the framework could help understand whether deprivation affects all patients similarly or whether it disproportionately impacts a particularly disadvantaged subset.

Published studies using UK primary care data observed that overall antibiotic prescribing rates are higher in more deprived areas, but they did not investigate the underlying reasons for these inequalities. One proposed explanation is differences in prescribing behaviour, with general practitioners more likely to prescribe antibiotics to patients from more deprived areas due to concerns about the increased risk of complications,^12^ or shorter consultation times in more deprived areas, which may lower the threshold for prescribing.^9^ Another possibility is that patients from more deprived areas consult more frequently, as they might have more health needs,^40^ or they have easier access to general practices.^41^ Our study provides empirical evidence supporting the hypothesis that differences in consultation frequency between less and more deprived areas are the main driver of inequalities in prescribing. In contrast, we did not observe that patients from more deprived areas were more likely to receive an antibiotic prescription after having consulted for several indications including LRTI, URTI, and otitis media. In fact, the opposite pattern was observed for several infections. This aligns with a previous study using CPRD data, which found lower odds of receiving an antibiotic for these indications for more deprived areas, conditional on the consultation having happened.^19^ One possible explanation is that patients who require antibiotics may access healthcare through routes other than general practice, such as emergency departments, out-of-hours services, or walk-in centres, which are not captured through current analysis.

Therefore, tackling inequalities in consultations may be a key pathway to addressing inequalities in antibiotic prescribing in primary care, and this requires identifying effective, targeted interventions. Our study found that vaccination uptake, particularly seasonal influenza vaccination, partly explained inequalities in consultations for respiratory infections among children. This may be because influenza vaccination reduced children’s infection risks,^18^ while vaccination coverage is lower in more deprived areas.^14^ Among adults, smoking appeared to play a more important role in inequalities in respiratory infections. This finding aligns with a recent UK study showing that smoking mediated socioeconomic inequalities in respiratory infectious diseases,^42^ which can be explained by the higher risk of respiratory infections among smokers,^16^ ^43^ and the higher prevalence of smoking in lower socioeconomic background.^13^ ^44^ Other factors, including BMI and vaccination, played a less important role in adults.

### Limitations

The composite index of multiple deprivation used in our study includes a health domain capturing morbidity, disability and premature mortality,^26^ which may partly overlap with our outcome measures. This may lead to overestimation of the association between deprivation and infection-related outcomes. However, the health domain accounts for a small proportion (13.5%) of the overall index,^26^ and is therefore unlikely to introduce substantial bias into our findings. In addition, missingness in the observed population was primarily driven by IMD (supplementary table S10). We conducted multiple imputation to address missing data. The imputed covariates showed similar distributions to the complete case (supplementary fig S15 and table S8), and estimates of differences in antibiotic prescribing between the least and most deprived areas were consistent across the complete case and imputed analyses (supplementary tables S11-12). These findings suggest that missing data are unlikely to have substantially influenced the observed results.

We cannot disentangle differences in the underlying incidence of symptomatic indication from differences in consultation propensity across deprivation groups. Examining whether indication-related hospital admissions vary by deprivation could help clarify this issue; however, this was beyond the scope of current study. The attenuation of inequalities after accounting for vaccination uptake suggests that our findings do not solely reflect differences in consultation behaviour, but at least partly reflect genuine differences in indication incidence.

We assessed whether certain factors explained inequalities in indication-related outcomes, but some contributors could not be included due to the data limitations, such as passive smoking in children, and other household factors, including overcrowding. As a result, the relative importance of vaccination in children and smoking in adults may be overestimated. Potential unmeasured factors such as occupational exposures, may also influence the observed association between smoking and consultation frequency. Failure to account for these factors may bias the estimated contribution of smoking to socioeconomic inequalities in either direction. Furthermore, as contributions were estimated using separate regression models rather than a fully joint model, they should be interpreted as model-based explanatory components rather than causal mediation effects. Nevertheless, analyses accounted for potential key confounders, including age, sex, and ethnicity, which may partly reduce residual confounding.

Prescribing behaviour varies by general practice.^9^ We did not account for clustering at the general practice level due to the complexity of incorporating multilevel structures within the GAMLSS framework. This may affect the observed prescribing patterns across deprivation levels. However, our analyses focused on individual-level association between deprivation and common indications, and the large, nationally representative sample provided robust evidence of population-level inequalities.

We divided the study population into children and adults, while incidence of infection-related indications varies across finer age groups, such as infants and young children (<4 years) compared with primary school-aged children, and working-age adults compared with older adults (≥65 years). However, this approach allowed us to provide a clear picture of inequalities across 17 common infections. Future studies could explore these patterns in more age-specific detail.

Our initial cohort was restricted to individuals who had at least one consultation for one of the 17 included indications during 2016 to 2019 due to the limitation of the original data extraction. As a result, explanatory variables for individuals who did not consult were estimated rather than directly observed. The sensitivity analysis restricted to the observed population estimated deprivation gaps in antibiotic prescribing that were smaller for children and larger for adults for most indications, while the direction of the association remained unchanged (supplementary table S8). For adult cellulitis and gastroenteritis, negative associations in the main analysis became positive (supplementary results). However, these changes did not affect the overall interpretation that inequalities in prescribing are primarily driven by differences in consultation frequency rather than prescribing behaviour once patients consult (supplementary fig S13). Additionally, restricting the study period to before the COVID-19 may not capture the potential changes in health-seeking behaviour and prescribing patterns in the post pandemic era. However, this also allows our findings to reflect consultation and prescribing patterns unaffected by the disruptions associated with the pandemic.^45^

### Implications and conclusions

Although previous work has highlighted socioeconomic variation in antibiotic prescribing, it has been unclear what drives these inequalities, and antibiotic stewardship interventions have often been proposed as the solution. Our study shows instead that deprivation-related inequalities are largely explained by higher consultation rates for common indications, particularly respiratory tract infections. This implies that reducing inequalities will require interventions that lessen the need to consult, rather than change prescribing behaviour once patients present, which our study found was at most weakly associated with deprivation.

Our findings suggest that increasing seasonal influenza and pneumococcal vaccination uptake among deprived children and expanding smoking cessation programmes for deprived adults may be more effective at narrowing these inequalities than targeted stewardship initiatives alone. Individual-level smoking cessation interventions have been shown to be effective among socioeconomically disadvantaged populations in the UK.^46^ While specific approaches vary, previous studies have highlighted the value of targeting smoking cessation support for lower socioeconomic groups in the UK.^47^ ^48^ Our findings support this principle, suggesting that expanding cessation support in primary care and community pharmacies in deprived areas may be a practical way to reduce the burden of respiratory infections and consequently reduce inequalities in antibiotic prescribing. However, smoking may also act as a proxy for other unmeasured health risks and comorbidities; therefore, the extent to which smoking cessation alone would reduce inequalities in antibiotic prescribing remains uncertain.

Additionally, large inequalities remain even after accounting for vaccination and smoking, and further work is needed to identify other modifiable drivers of excess consultations. More broadly, stewardship efforts remain essential across all levels of deprivation given high prescribing rates for several conditions that infrequently require antibiotics.

In contrast to the consistently observed deprivation association for respiratory tract indications, we found opposite pattern for UTI in adults, with higher prescribing rates in less deprived areas (table 3 and supplementary fig S3). This differs from the strong association between higher deprivation and increased rates of UTI-related hospital admissions and community-acquired bacteraemias caused by *Escherichia coli*,^6^ ^49^ ^50^ suggesting that inequalities in severe outcomes may arise from differences in healthcare access, health-seeking behaviour, or delayed presentation. Although we could not directly assess timing of presentation, these findings raise the possibility that earlier recognition and treatment of urinary tract infections in more deprived areas could represent a modifiable pathway of reducing Gram-negative bloodstream infections. Further work is needed to examine consultation patterns and pathways to admission in relation to socioeconomic status.

Overall, reducing socioeconomic inequalities in antibiotic prescribing will require interventions that go beyond stewardship to address upstream determinants of indication and consultation.

## Ethical approval

This study was approved by the Clinical Practice Research Datalink’s (CPRD) independent scientific advisory committee (protocol 23_003072). CPRD also has ethical approval from the Health Research Authority to support research using anonymised patient data (research ethics committee reference 21/EM/0265). Individual patient consent was not required as all data were deidentified.

## Data availability statement

Electronic health records are, by definition, considered sensitive data in the UK by the Data Protection Act and cannot be shared via public deposition because of information governance restriction in place to protect patient confidentiality. Access to Clinical Practice Research Datalink (CPRD) data is subject to protocol approval via CPRD’s research data governance process. For more information see https://cprd.com/data-access. Linked index of multiple deprivation data can also be requested from CPRD.

## Contributors

KBP conceived the project and wrote the protocol with MY. NVN and MY prepared the data for analysis. MY conducted the statistical analyses and wrote the first draft of the manuscript under the supervision of KBP. All authors contributed to the interpretation of the data, critically revised the manuscript and approved the final version. MY and KBP act as guarantors. The corresponding author attests that all listed authors meet authorship criteria and that no others meeting the criteria have been omitted.

## Transparency

The corresponding author affirms that the manuscript is an honest, accurate, and transparent account of the study being reported; that no important aspects of the study have been omitted; and that any discrepancies from the study as planned and registered have been explained.

## Copyright

The Corresponding Author has the right to grant on behalf of all authors and does grant on behalf of all authors, a worldwide licence to the Publishers and its licensees in perpetuity, in all forms, formats and media (whether known now or created in the future), to i) publish, reproduce, distribute, display and store the Contribution, ii) translate the Contribution into other languages, create adaptations, reprints, include within collections and create summaries, extracts and/or, abstracts of the Contribution, iii) create any other derivative work(s) based on the Contribution, iv) to exploit all subsidiary rights in the Contribution, v) the inclusion of electronic links from the Contribution to third party material where-ever it may be located; and, vi) licence any third party to do any or all of the above.

