## supplementary materials for "Explaining socioeconomic inequalities in antibiotic prescribing for common infections in English primary care: a population-based study"

### Supplementary files

### Supplementary file 1: supplementary methods

##### S1.1 Vaccination status, smoking status, and body mass index (BMI) for non-consulting population

**Influenza vaccination**

a. Age-specific uptake rates

Age-group-specific influenza vaccination uptake (<2, 2-4 years, 5-15 years, 16-64 years, 65+) was obtained from Public Health England for the 2018/19 winter season.[1] As these data were only available for broad age bands, we derived single-year age-specific vaccination patterns using the Clinical Practice Research Datalink (CPRD) observed population by calculating age-specific vaccination prevalence based on vaccinated counts and population denominators within each age. These CPRD-derived age patterns were then scaled to match the national age-group-level uptake rates using England population counts, producing estimated national vaccinated counts and prevalence for each single year of age. This approach assumes that the within-age group pattern of uptake by single year of age observed in CPRD is representative of the national age association.

b. Index of Multiple Deprivation (IMD)-specific uptake rates

IMD-specific influenza uptake was drawn from the Fingertips public health data platform, which provides routinely collected population-level indicators for England. Data were extracted using the *fingertipsR* package, which provides programmatic access to the Fingertips application programming interface (API).[2] The uptake was drawn for the 2019/20 season, the closest available data to the study period.[3-5] Uptake was reported by IMD decile, ranked from least deprived to most deprived. These data were available for children aged 2-3 years, 4-11 years, and adults aged 65 years and over. In the absence of IMD-specific uptake data for adults aged 16-64 years, we approximated the IMD distribution by averaging the available IMD-specific vaccinated counts and population denominators across the 2-3, 4-11, and 65+ groups with available data. Within each age group, we used the England population counts to scale IMD-specific uptakes to produce national vaccinated counts by IMD decile.

c. Single age-IMD-specific uptake rates

For each broad age group (<2, 2-4 years, 5-15 years, 16-64 years, 65+), IMD-specific uptake distributions were assigned as follows: the 2-3 years IMD distribution was applied to ages <2 and 2-4 years, the 4-11-year IMD distribution to ages 5-15 years, the average IMD distribution to ages 16-64 years, and the 65+ IMD distribution to ages 65 years and over. We applied iterative proportional fitting separately within each broad age group to jointly distribute both population denominators and vaccinated counts across age × IMD decile cells, ensuring that the marginal totals matched observed age and IMD constraints, yielding a consistent prevalence matrix by single year of age and IMD decile. Age-specific uptakes by IMD decile were expanded to single year of age and IMD twentile (1 least deprived, 20 most deprived) by assigning the corresponding IMD decile value to each pair of IMD twentile (e.g. decile 1 values to twentiles 1-2; decile 10 values to twentiles 19-20). This approach assumes that uptake rates are similar within each decile.

**Pneumococcal vaccination**

*Pneumococcal conjugate vaccine (PCV)*

a. Age-specific coverage rates

The PCV immunisation programme was introduced into the UK in 2006.[6] Therefore, individuals aged 13 years and above in 2019 (i.e. born in 2006 or earlier) were assumed to have not received PCV. For children aged 12 and below, single-year age-specific PCV coverage was obtained from the Fingertips for ages 9 and below,[7] and from NHS Digital for children aged 10 to 12.[8] Coverage data were available for both the primary PCV dose (PCV1, administered at 1 year of age) and the PCV booster (administered at 2 years of age). We used PCV1 coverage in the analysis.

b. IMD-specific coverage rates

IMD-specific PCV coverage was drawn from the Fingertips API.[7] IMD distributions by single year of age were available for children aged 3 and below (i.e. <1, 1, 2, and 3 years), with coverage reported by IMD decile. For ages where IMD-specific data were unavailable, we constructed an average IMD distribution by averaging vaccinated counts and population denominators across the four available age groups (<1, 1, 2, and 3 years) and recalculating prevalence. These averaged IMD-specific vaccinated counts and denominators were then scaled to match the corresponding age-specific totals.

c. Age-IMD-specific coverage rates

For children aged 3 years and below, age-specific IMD coverage distributions were applied directly. For children aged 4-12, the average IMD distribution illustrated in section (b) was assigned. We applied iterative proportional fitting to jointly distribute both population denominators and vaccinated counts across age × IMD decile cells, ensuring that the marginal totals matched observed age and IMD constraints, yielding a consistent prevalence matrix by single year of age and IMD decile. Age-specific coverage rates by IMD decile were expanded to single year of age and IMD twentile by assigning the corresponding IMD decile value to each pair of IMD twentile (e.g. decile 1 values to twentiles 1-2).

*Pneumococcal Polysaccharide Vaccine (PPV)*

a. Age-specific coverage rates

PPV is recommended for adults aged 65 years and over.[9] Age-specific PPV coverage for adults aged 65, 66, 67, 68, 69, 70-74, and 75+ was obtained from Public Health England for the 2018/19 season,[10] with both vaccinated counts and population denominator available.

b. IMD-specific coverage rates

As IMD-specific PPV coverage was not available for earlier years, we used data from the Fingertips API for the 2022/23 and 2024/25 seasons,[11] assuming that the deprivation association in uptake remained stable over time. Coverage was reported by IMD decile. To approximate the IMD distribution for 2019, we constructed an average IMD distribution by averaging vaccinated counts and population denominators across the two seasons. We then scaled the averaged vaccinated counts and denominators to match the age-specific total vaccinated counts and population denominators.

c. Age-IMD-specific coverage rates

We applied iterative proportional fitting to jointly distribute both population denominators and vaccinated counts across age × IMD decile cells, ensuring that the marginal totals matched observed age and IMD constraints, yielding a coherent prevalence matrix by age and IMD decile. Age-specific coverage rates by IMD decile were expanded to single year of age and IMD twentile by assigning the corresponding IMD decile value to each pair of IMD twentile (e.g. decile 1 values to twentiles 1-2).

**Smoking status**

Age-IMD-sex-ethnicity-specific smoking prevalence

Smoking status was included as a potential contributor among adults aged 18 years and over only. Smoking prevalence (current smoker, ex-smoker, non-smoker) for this population was obtained from the Fingertips API.[12-14] Prevalence was reported separately by IMD decile, age group (four-year age bands), sex, and ethnicity. IMD-specific prevalence was collected from data aggregated over the period 2019-2021, the closest available time window to the study period, while prevalence by age group, sex, and ethnicity was obtained from data reported in 2018. For each smoking status category, we applied iterative proportional fitting to jointly distribute both population denominators and status-specific counts across age × IMD decile × sex × ethnicity cells, ensuring that the marginal totals matched observed age, IMD, sex, and ethnicity constraints, yielding a consistent prevalence matrix by age, IMD decile, sex, and ethnicity. Smoking prevalence estimates were uniformly assigned to each single year of age within each age group and to the corresponding IMD twentile within each IMD decile (e.g. decile 1 values to twentiles 1-2, decile 5 values to twentile 19-20). This approach assumes that smoking prevalence is similar within each age group and IMD decile.

**Body Mass Index (BMI)**

Age-IMD-sex-ethnicity-specific BMI values

BMI values for adults aged 18 years and over were obtained from Health Survey for England 2018, which included a nationally representative sample of 10,250 individuals in England with information on age (approximately five-year age bands: 18-19, 20-24, 25-29, …, 85-89, ≥90 years), IMD quintile, sex, and ethnicity.[15] For each age group × IMD quintile × sex × ethnicity stratum, we calculated the mean BMI value and the corresponding standard deviation. The estimated mean BMI values were uniformly assigned to each single year of age within each age group and to the corresponding IMD twentile within each IMD quintile (e.g. quintile 1 values to twentiles 1-4, quintile 5 values to twentile 17-20). This approach assumes that the mean BMI values are similar within each age group and IMD quintile.

BMI values for children aged under 18 years were also available in Health Survey for England 2018 dataset in five-year age bands. However, the recommend approach for analysing BMI in children is to use BMI-for-age, which requires the child's exact age to calculate age-specific z-scores.[16] As the dataset did not provide age at this level of precision, BMI-for-age could not be calculated reliably for non-consulting patients. Therefore, due to the data limitations, BMI was not included in the analysis for children.

**S1.2 Generalised Additive Models for Location, Scale, and Shape (GAMLSS)**

GAMLSS models assume that a health outcome can be described by a parametric distribution characterised by multiple parameters, each of which can be conditional on explanatory variables. Each parameter is linked to a regression predictor through a specific link function (such as a log or logit), and the model coefficients are estimated by maximising the likelihood function.[17]

An appropriate distribution was chosen based on the nature of the outcome variable, i.e. whether it is continuous, discrete, or mixed.[18] For count outcomes (number of antibiotic prescriptions and number of consultations), we applied the negative binomial type I distribution (NBI) distribution with a log link and included person-time as a log offset to model rates. The NBI distribution has two parameters, location ($\mu$) and scale ($\sigma$), both modelled using log links. After fitting the model, parameter estimates on the original scale were obtained by exponentiating the corresponding linear predictors. The mean of the outcome distribution equals $\mu$, while the variance is calculated by $\mu + \sigma\mu^2$.[18]

To quantify uncertainty, we constructed 95% confidence intervals for the mean and variance using a simulation-based approach. We drew 1000 sets of regression coefficients from the estimated sampling distribution of the model coefficients (defined by the estimated coefficients and variance-covariance matrix from the regression model), then used these to generate values of the mean and variance across IMD. This approach was chosen because closed-form standard errors are not available for these derived quantities, and bootstrapping was not computationally feasible given the size of the dataset.

**S1.3 Estimation of uncertainty (95% confidence interval) for the deprivation gap**

To facilitate interpretation, we quantified socioeconomic inequalities in prescribing and consultation rates by comparing differences between the most (IMD=20) and least deprived levels (IMD=1), as shown in Tables 3 & 4 and Figure 3 of the main text. We also quantified the extent to which these inequalities were explained by each contributor, using absolute and percentage variation in the mean explained. Similar to the approach described for mean and variance measures (supplementary method S1.2), uncertainty in these analyses was captured by constructing 95% confidence intervals using posterior simulation. We simulated model coefficients using the estimated coefficients and variance-covariance matrix of the corresponding model and repeated the procedure for 1000 draws.

For analyses comparing the differences in mean prescribing rates between the most deprived (IMD=20) and least deprived (IMD=1, reference), the variance-covariance matrix was obtained from the baseline model for the association between IMD and number of prescriptions, which adjusted for age, sex, and ethnicity, and included time at risk as an offset.

For analyses quantifying the contribution of each factor to socioeconomic inequalities in consultation rates, and for analyses comparing the differences in mean consultation rates between the most deprived (IMD=20) and least deprived (IMD=1, reference) within each contributor category, the variance-covariance matrix was derived from the adjusted model for the association between IMD and number of consultations, adjusting for age, sex, ethnicity, and the contributor of interest, and including time at risk as an offset.

**S1.4 Change of IMD effect size with contributors added**

Each contributor was added separately to the baseline model (i.e. model adjusting for age, sex, ethnicity, and including time at risk as an offset) to form the adjusted models. To examine whether contributors explain inequalities, we compared the isolated IMD association (effect size of IMD) across the baseline and adjusted models using the delta method. This allowed us to assess how the estimated association between IMD and the outcome varied when additionally adjusting for the contributor of interest. Specifically, to obtain this isolated IMD association, we extracted the model coefficients corresponding to the IMD spline terms together with their variance-covariance matrix, and used these to calculate the combined IMD association and its uncertainty for each model. A reduction in the effect size (slope) of IMD on outcomes was interpreted as evidence that the contributor may partly explain the observed inequalities.

### Supplementary file 2: supplementary results

**Comparison of characteristics between observed and total population**

The sensitivity analysis restricted to the observed population included 9,426,835 individuals from the initial CPRD cohort who had at least one consultation for any of the 17 infections of interest between 2016 and 2019 and were registered with a general practice in 2019 (2,323,455 children and 7,103,380 adults). This accounted for 54.8% of the total population (observed and estimated non-consulting population) used in the main analysis (fig S1).

Compared with the total population which reflected the distribution of IMD, age group, sex, and ethnicity for patients registered with GP practices in England in 2019, the CPRD observed population included proportionally fewer children from less deprived areas and more children from more deprived areas. In contrast, among adults, the observed population included fewer patients from more deprived and more patients from less deprived areas. The observed population also included proportionally more younger children and older adults, more adult females and more individuals of White ethnicity (figs S7-8).

Among the observed population, individuals from the most deprived quintile had lower vaccination rates among both children and adults, and among adults, they were more likely to be current smokers, and to have a higher median BMI compared to those from less deprived areas (table S6). These patterns were consistent with those observed in the total population.

Compared with the total population (table 2), the observed population had higher vaccination rates and higher proportion of current smokers across all deprivation quintiles. These patterns suggest that observed populations may include individuals with greater healthcare needs that the total registered population.

**Antibiotic prescriptions by deprivation quintile**

Prescribing rates were consistently higher in the observed population than in the total population. This reflects the different target populations: the total population analysis estimates prescribing rates among the GP-registered population overall, whereas the observed population analysis estimates rates among individuals with a history of consultation for the infections of interest during 2016–2019 (tables S1 and S7). Consistent with the main analysis, among the observed population the highest age-standardised antibiotic prescribing rates were observed for upper and lower respiratory tract infections, urinary tract infections, and acute cough in both children and adults, and acute otitis media in children.

Similar to the main analysis using the total population, a positive association between higher deprived quintile and higher prescribing rates was observed for most respiratory infections in the observed population (figs 1 and S9). Notably, a clear positive association for acute cough among adults was observed in the observed population, whereas no such association was observed based on the total population.

**Inequalities in antibiotic prescribing, consultations, and probability of receiving antibiotic prescriptions**

In the baseline model adjusted for age, sex, and ethnicity, the direction of the association between deprivation and prescribing rates was consistent for most infections between the main analysis and sensitivity analysis (figs S3 and S11), although the magnitude of the associations differed. These included all respiratory infections (except rhinosinusitis), impetigo, otitis media, gastroenteritis, and UTI among children, and all respiratory infections (except bronchitis, pneumonia, and sore throat), otitis externa, pyelonephritis, and prostatitis among adults.

For these infections in which prescribing rates were higher in the most deprived areas (IMD twentile=20) than in the least deprived areas (IMD twentile=1), deprivation gaps were consistently smaller in sensitivity analysis restricted to the observed population than in the main analysis using the total population for children, whereas the opposite pattern was observed for adults (table S8 and table 3). For example, among children, compared with the least deprived, mean prescribing rates were 24.6% (95% CI 21.7% to 27.7%) higher in the most deprived for lower respiratory tract infections when restricted to the observed population, compared with 47.6% (95% CI 44.2% to 51.3%) in the total population. In contrast, among adults, the deprivation gap for lower respiratory tract infections was larger in observed population (83.6%, 95% CI 82.7% to 84.5%) than in the total population (22.7%, 95% CI 21.4% to 24.1%).

These differences in deprivation gaps likely reflect the differing deprivation distributions of the two study populations. The observed population included proportionally more deprived children than the total population, which may lead to underestimation of deprivation gaps for infections with higher prescribing rates in more deprived areas. Conversely, the observed population included proportionally fewer deprived adults, which may result in overestimation of deprivation gaps for infections with higher prescribing rates in more deprived areas among adults.

Other infections did not show a consistent direction of association between the observed and total population. For several infections, deprivation gaps between the most and least deprived areas changed from statistically significant in the total population to no clear pattern in the observed population (rhinosinusitis among children; sore throat, impetigo, and UTI among adults), or vice versa (otitis externa among children; bronchitis and pneumonia among adults). In addition, for gastroenteritis among adults, deprivation gaps changed direction, from negative in the total population to positive in the observed population.

Consistent with the main analysis, patterns of association between deprivation and indication-related outcomes were similar for antibiotic prescribing and consultation rates, but not for the probability of receiving antibiotics once consulted in the sensitivity analysis (fig S13).

**Role of contributors in explaining inequalities**

After adding the contributors to the baseline model, sensitivity analyses restricted to the observed population showed that influenza and pneumococcal vaccination both explained inequalities in consultation rates for most respiratory infections among children, and smoking contributed to inequalities in respiratory infections among adults (fig S14 and table S9). These findings were consistent with those of the main analysis.

Adding BMI to the baseline model also weakened the association for consultations in cellulitis in the observed population (fig S14). In the counterfactual scenario where BMI distributions in the most deprived areas matched those in the least deprived, the deprivation gap would be reduced by 93.6% (95% CI 81.7% to 106.0%) (table S9). However, because the observed deprivation gap for cellulitis was not statistically significant in the main analysis (table 3), the contribution of BMI and other factors to inequalities in cellulitis was not examined further in the main analysis.

### Supplementary file 3: supplementary figures

#### S3.1 Study population

##### Figure S1 Flow chart of patients included in the analysis

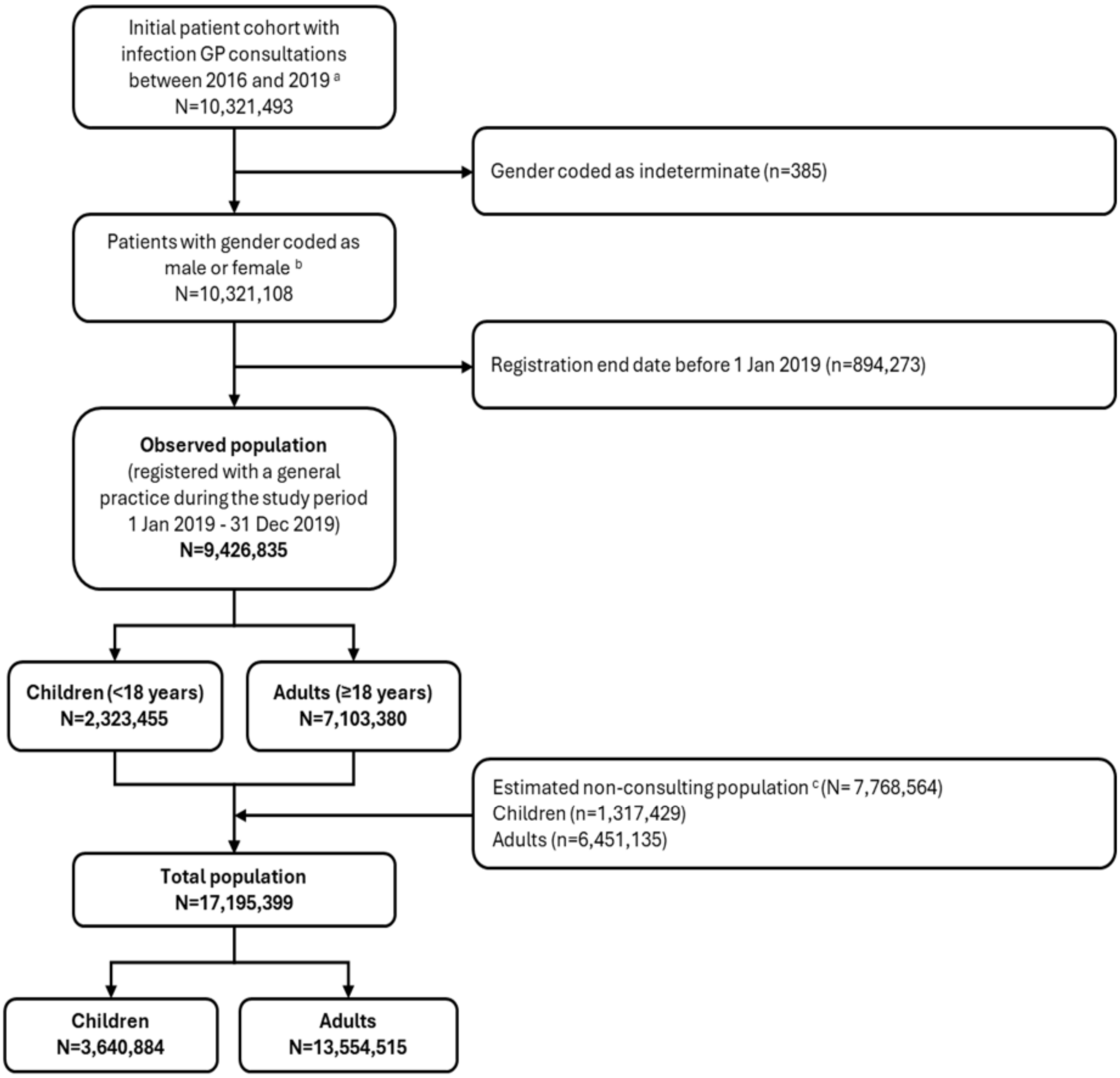


[a] Initial cohort included patients of all ages who were registered with an English general practice contributing to CPRD Aurum, whose records met CPRD's research-quality standard, and who had at least one consultation for one of the 17 infections of interest (defined in the main text *Variable definition*) during the 4-year period from 1 January 2016 to 31 December 2019. Practices likely to have merged with other contributing practices [19] and those closed before 1 Jan 2019 were excluded during data preprocessing and are not shown in the flow diagram. [b] Patients with gender coded as indeterminate were excluded due to the small number and because sex-specific analyses were conducted for certain outcomes (e.g., urinary tract infection in females and prostatitis in males).[c] Estimated non-consulting population included patients who did not consult for any of the 17 infections included in this study over 2016-2019 and were registered with a practice in 2019. This definition was used rather than restricting to non-consultation in 2019 alone, as patients who did not consult in 2019 were still included in the observed population if they consulted in earlier years (2016-2018). Number of non-consulting patients were estimated based on NHS England GP registration data. Details see main text Methods section.

#### S3.2 Main analysis using total population

##### S3.2.1 Prescribing rates by deprivation quintile

*Figure S2 Age-standardised antibiotic prescribing rates (prescriptions per 1000 person-years) for specific respiratory tract infections by IMD quintile in the total population*

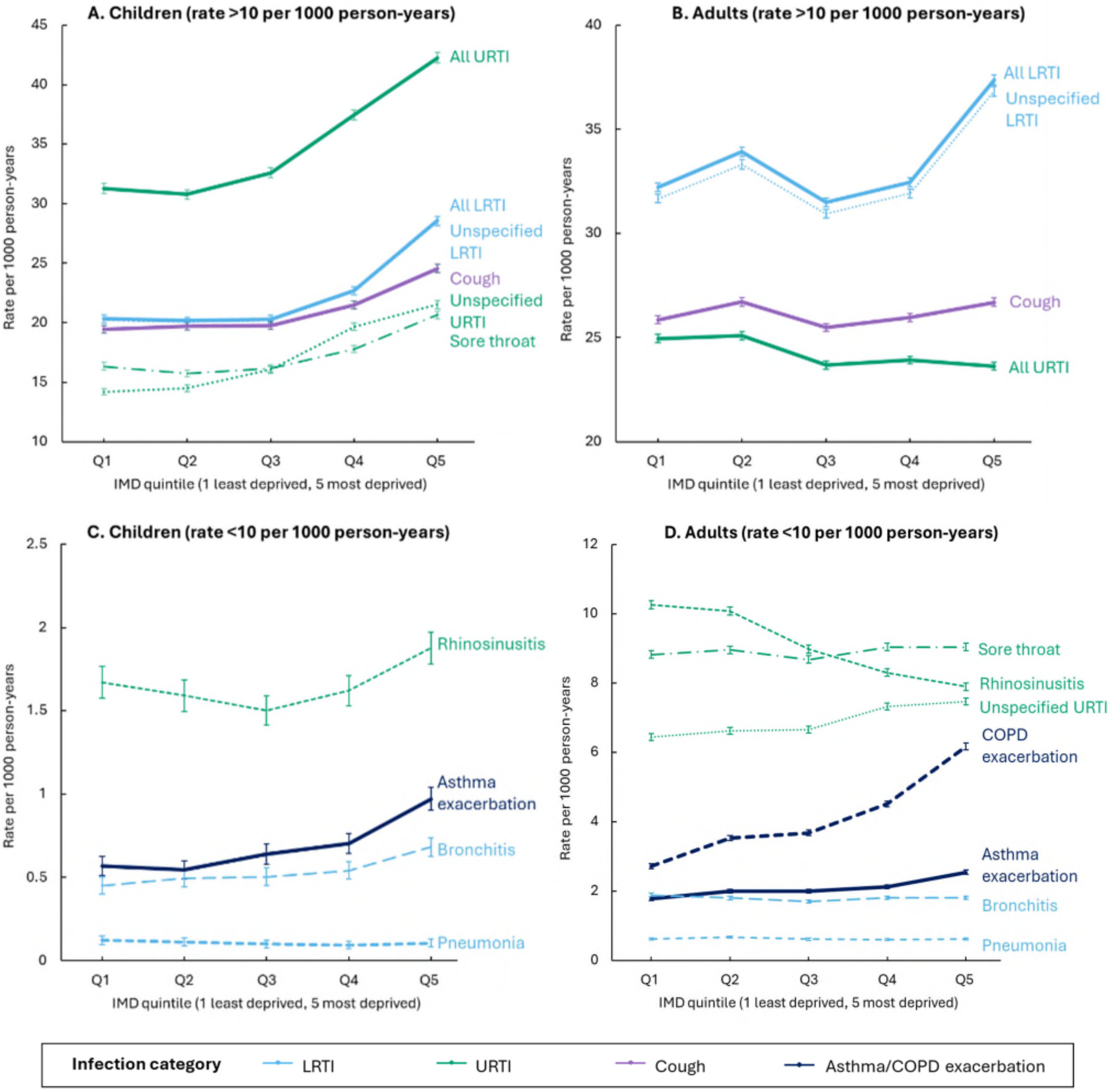


Notes: Error bars represent the 95% confidence intervals. Abbreviations: IMD index of multiple deprivation, URTI upper respiratory tract infections, LRTI lower respiratory tract infections, UTI urinary tract infection, COPD chronic obstructive pulmonary disease. “All URTI” includes sore throat, unspecified upper respiratory tract infection, rhinosinusitis whereas “All LRTI” includes bronchitis, unspecified lower respiratory tract infection, pneumonia.

##### S3.2.2 Association between deprivation and indication-related outcomes

*Figure S3 Association between IMD and antibiotic prescribing rates (prescriptions per 1000 person-years) - mean and variance measure with shaded 95% confidence intervals reported*

Panel (a) Children

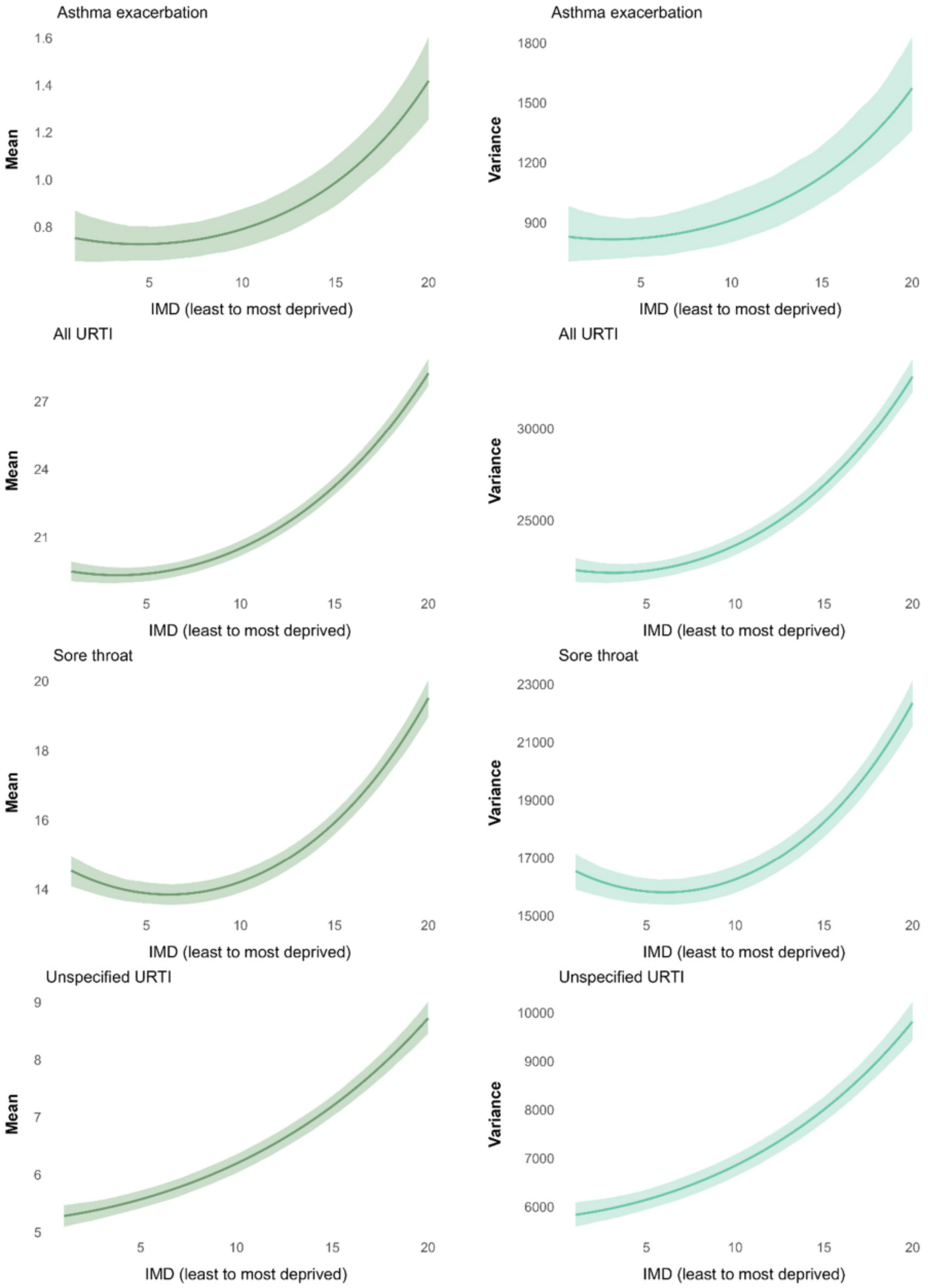

*Figure S3 (a). Prescribing rate (per 1000 person-years) in children - mean and variance (continued)*

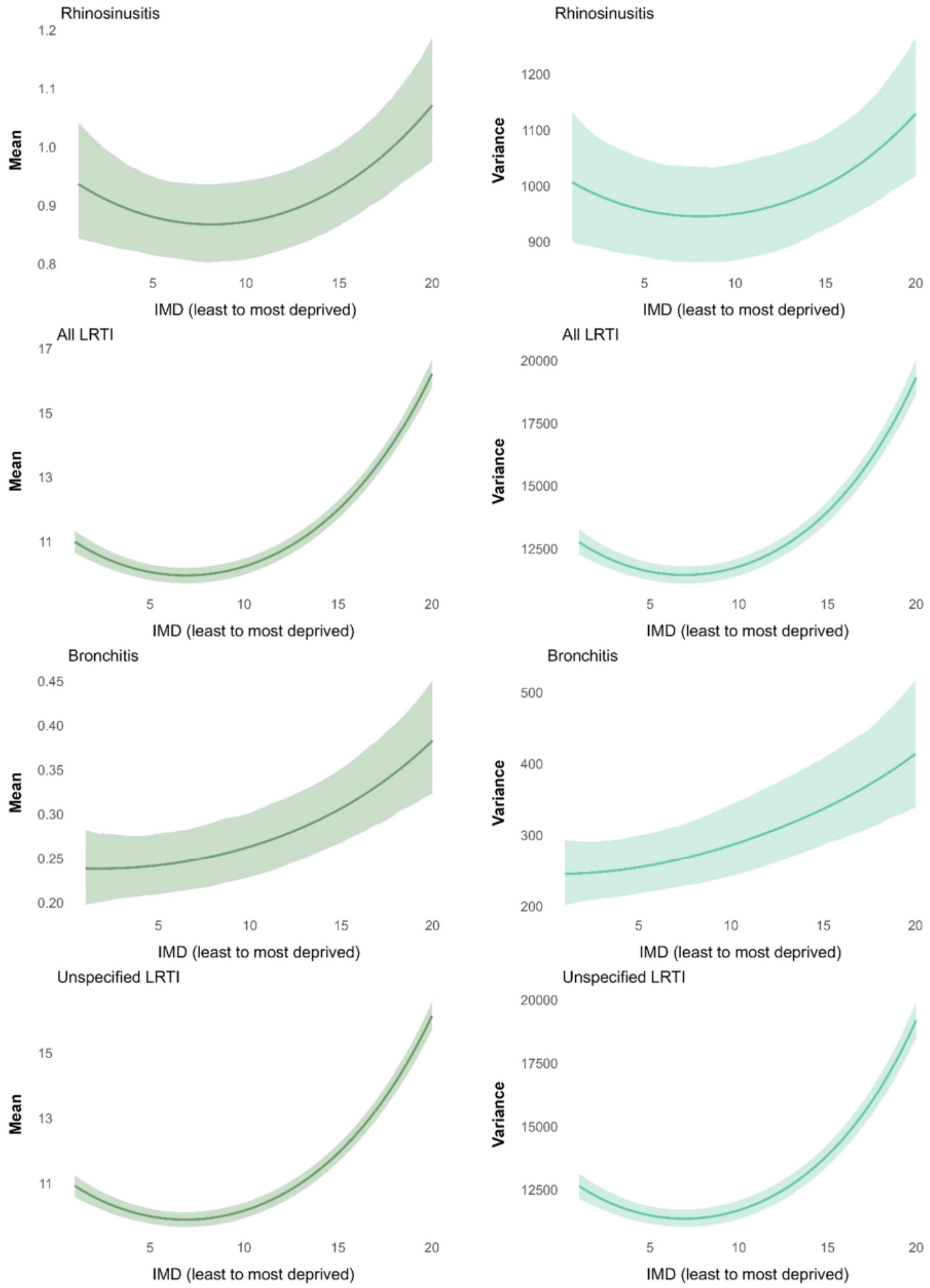

*Figure S3 (a). Prescribing rate (per 1000 person-years) in children - mean and variance (continued)*

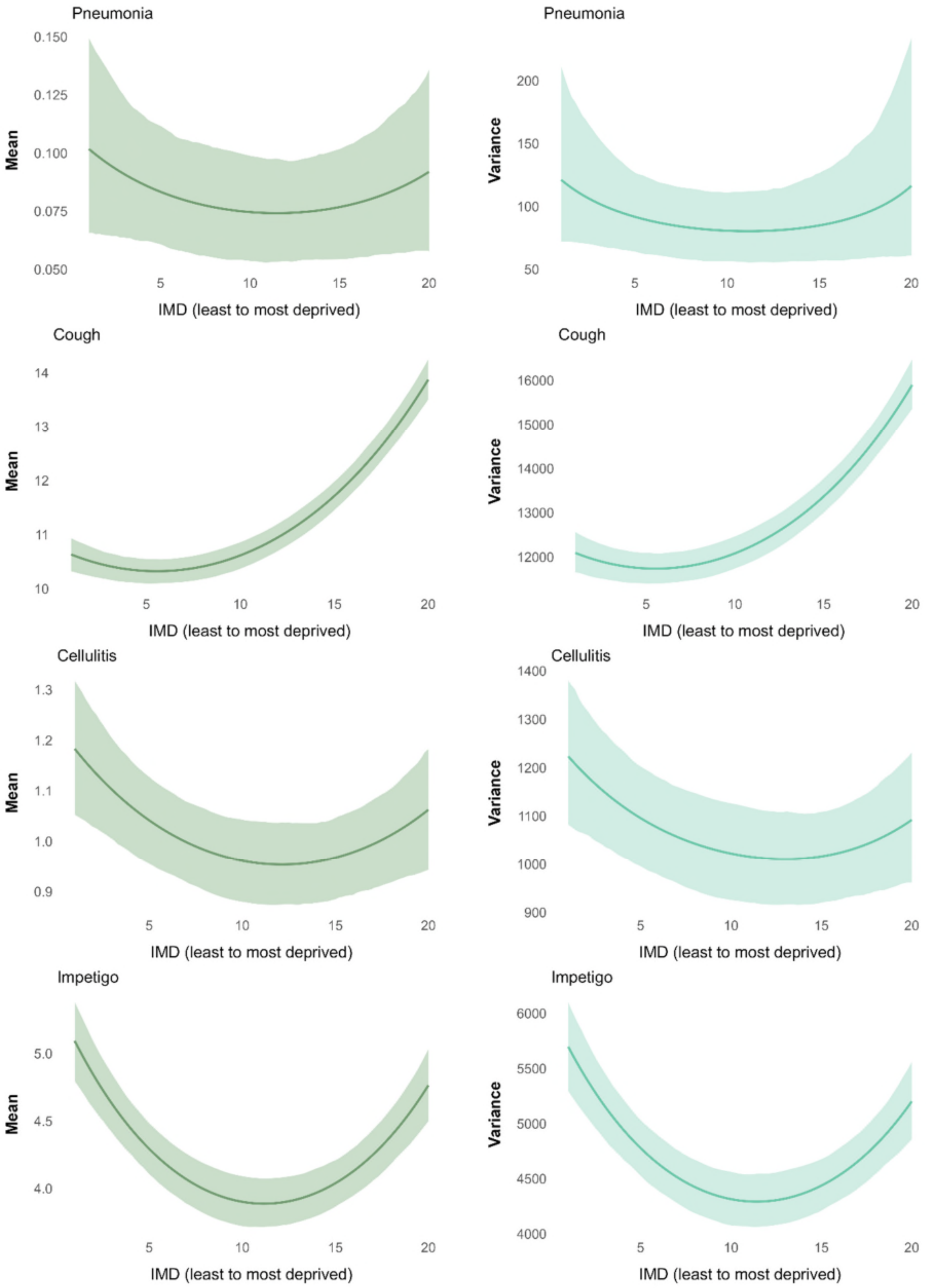

*Figure S3 (a). Prescribing rate (per 1000 person-years) in children - mean and variance (continued)*

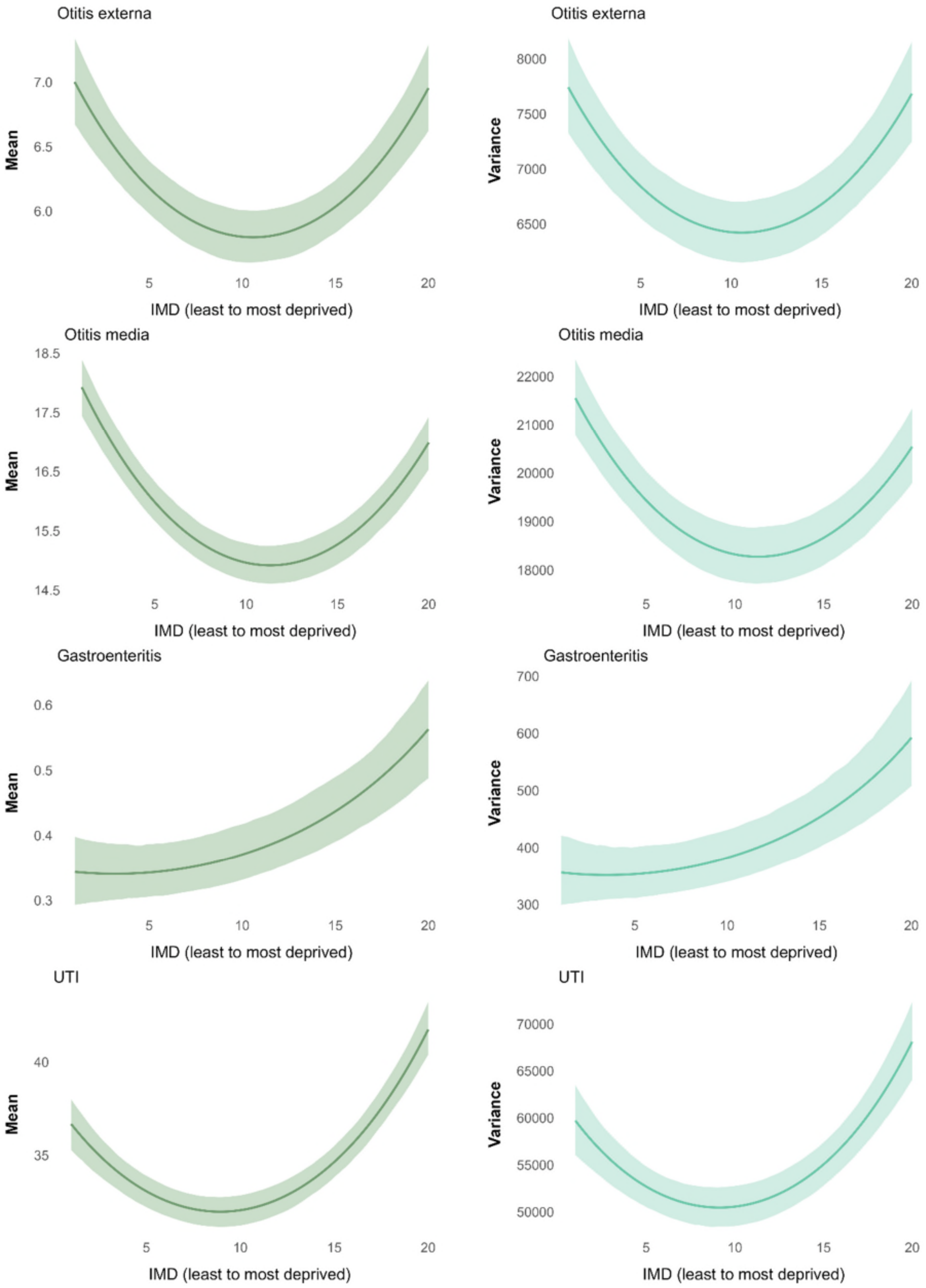

Panel (b) Adults

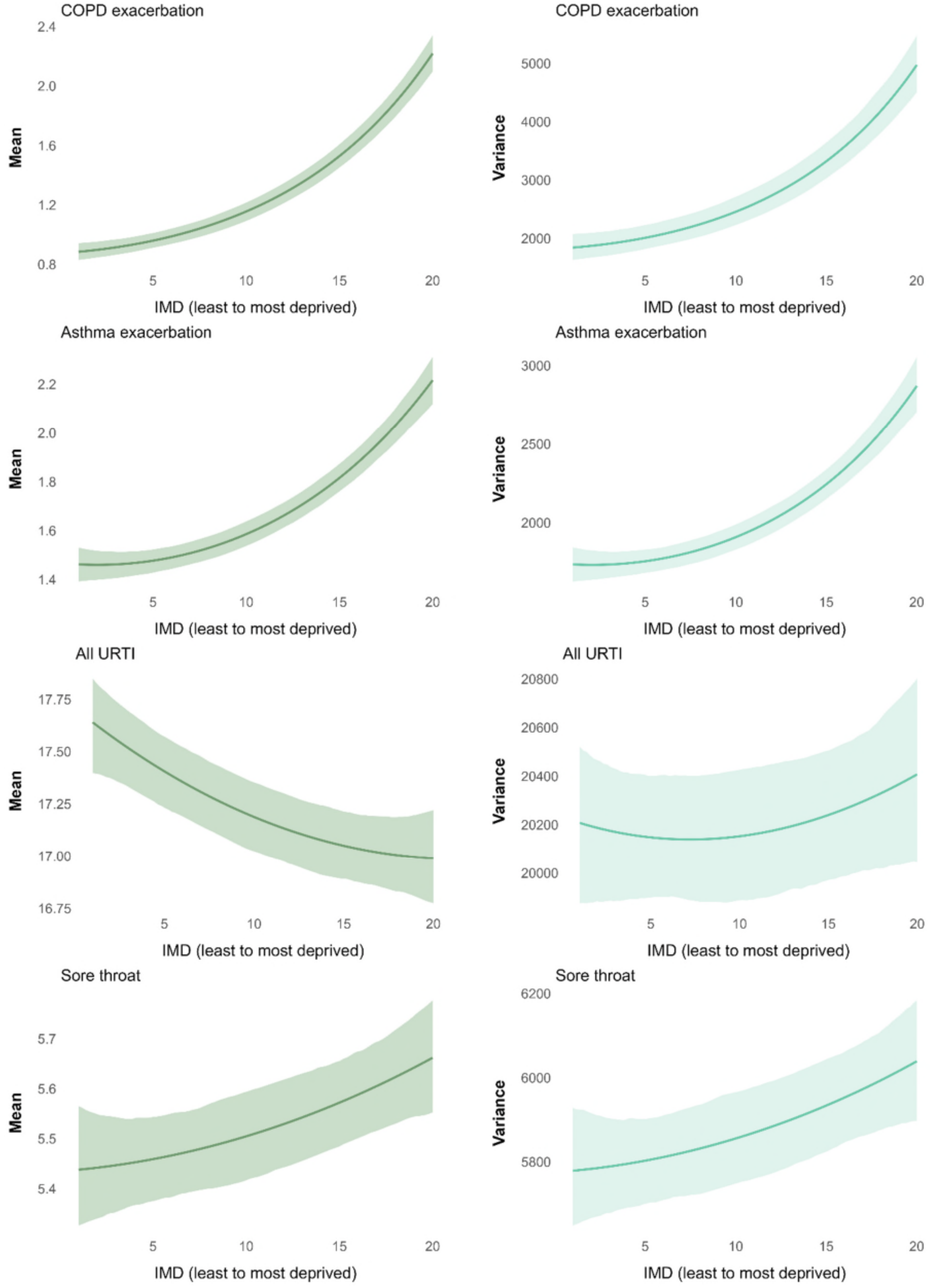

*Figure S3 (b). Prescribing rate (per 1000 person-years) in adults - mean and variance (continued)*

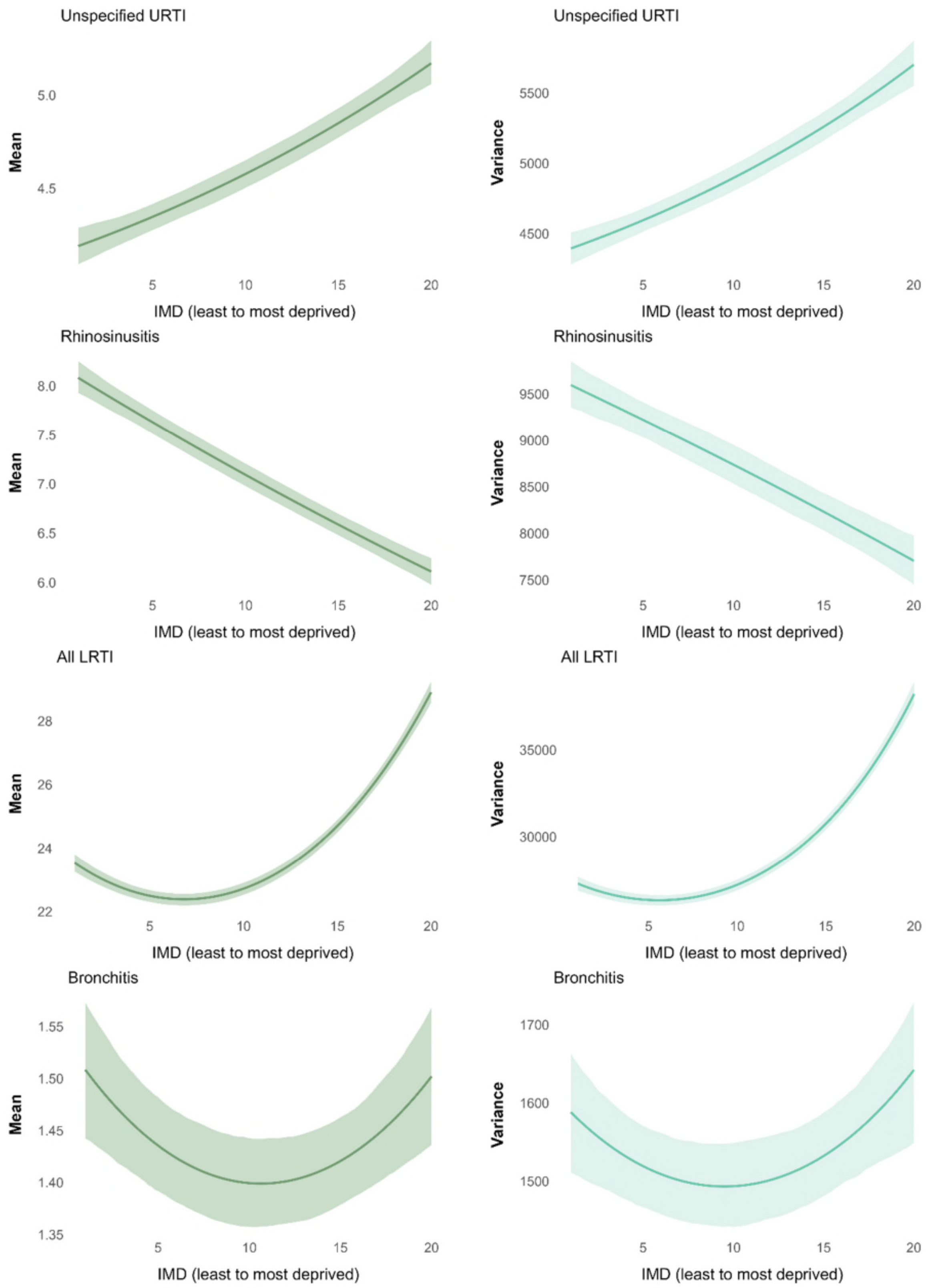

*Figure S3 (b). Prescribing rate (per 1000 person-years) in adults - mean and variance (continued)*

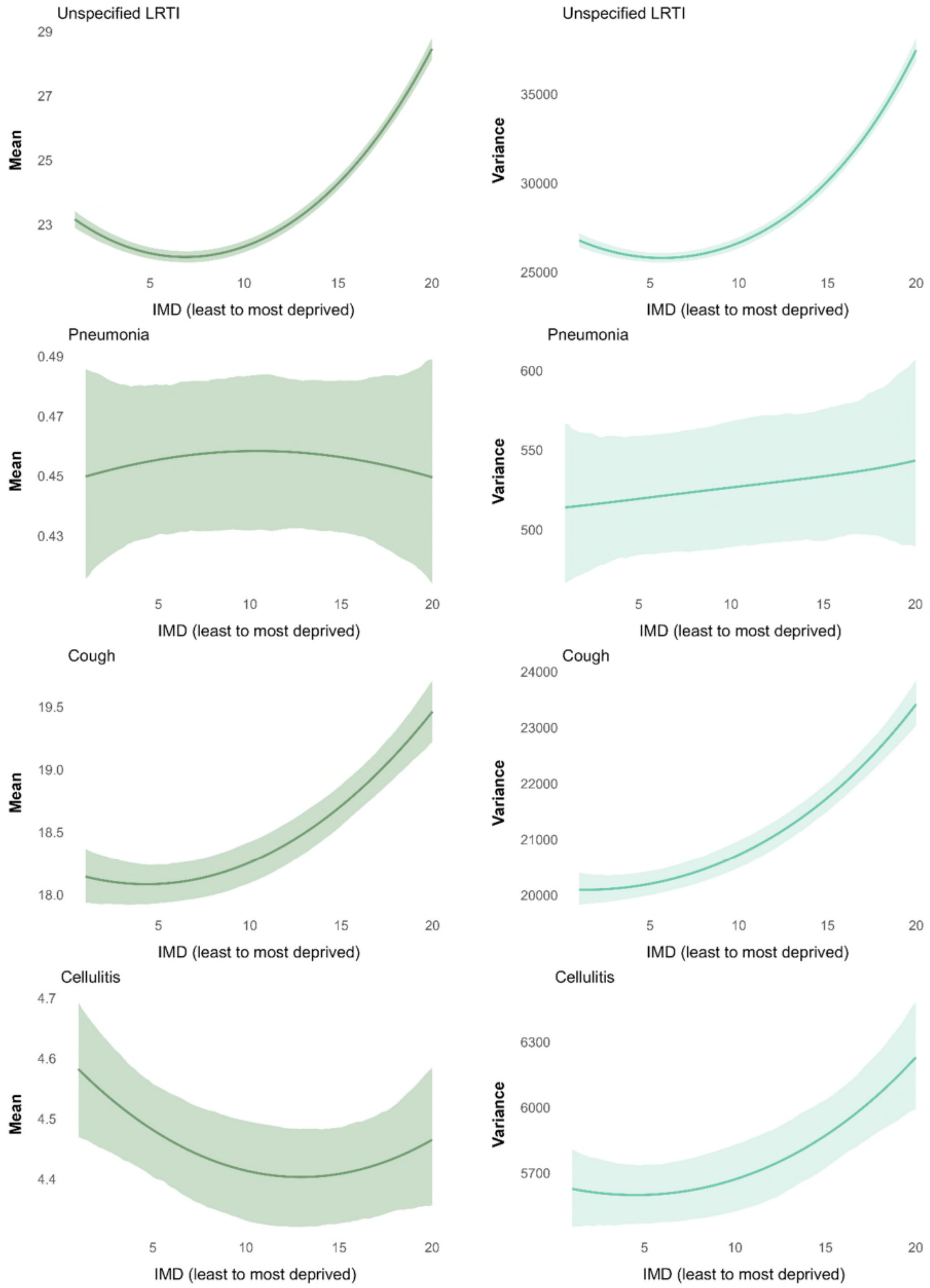

*Figure S3 (b). Prescribing rate (per 1000 person-years) in adults - mean and variance (continued)*

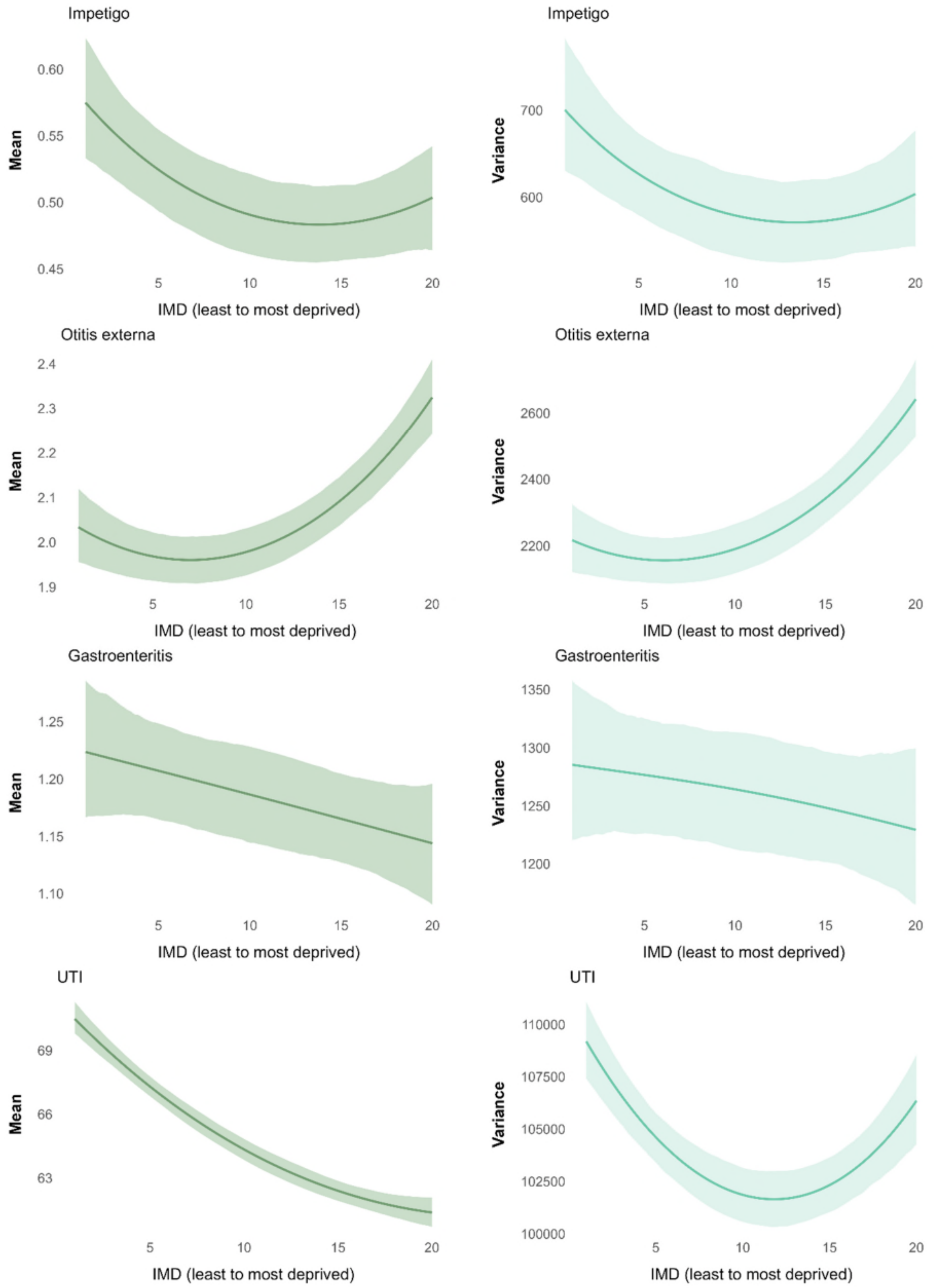

*Figure S3 (b). Prescribing rate (per 1000 person-years) in adults - mean and variance (continued)*

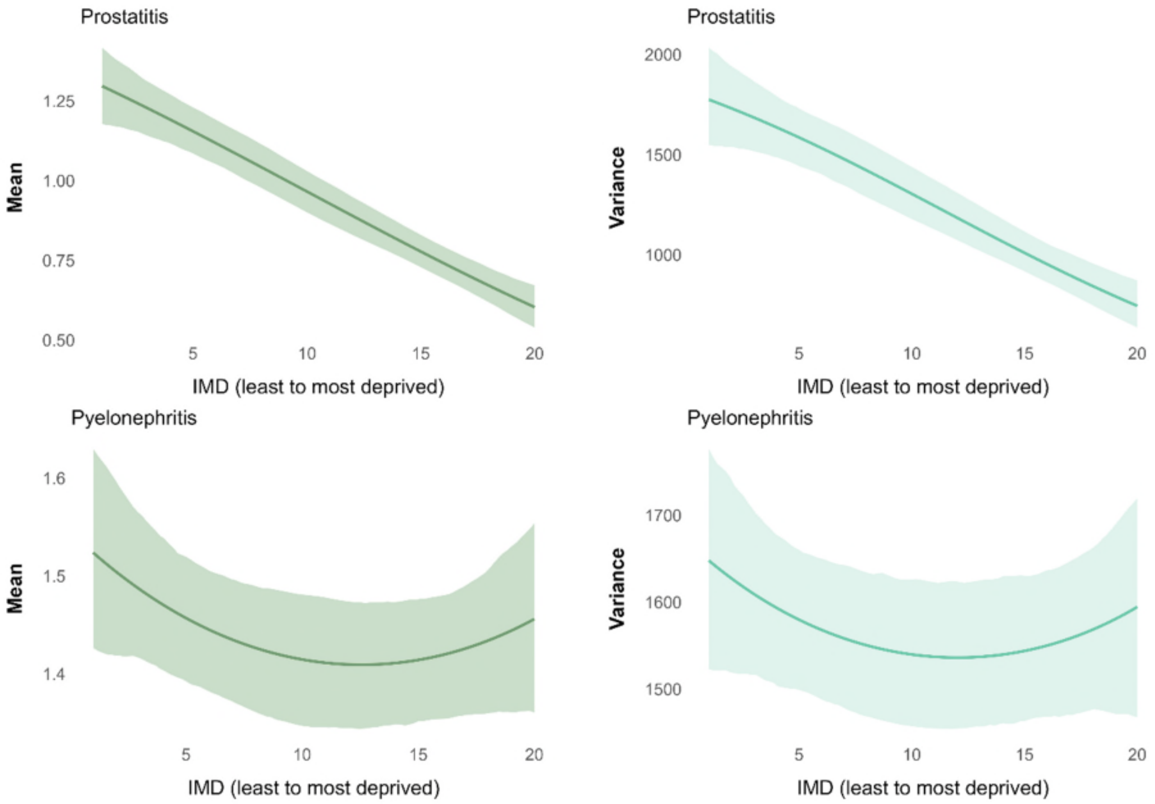


Notes: All models were adjusted for age, sex, and ethnicity. Abbreviations: IMD index of multiple deprivation, COPD chronic obstructive pulmonary disease, LRTI lower respiratory tract infections, URTI upper respiratory tract infections, UTI urinary tract infections. "All URTI" includes sore throat, unspecified upper respiratory tract infection, rhinosinusitis whereas "All LRTI" includes bronchitis, unspecified lower respiratory tract infection, pneumonia.

*Figure S4 Association between IMD and consultation rates (consultations per 1000 person-years) - mean and variance measure with shaded 95% confidence intervals reported*

Panel (a) Children

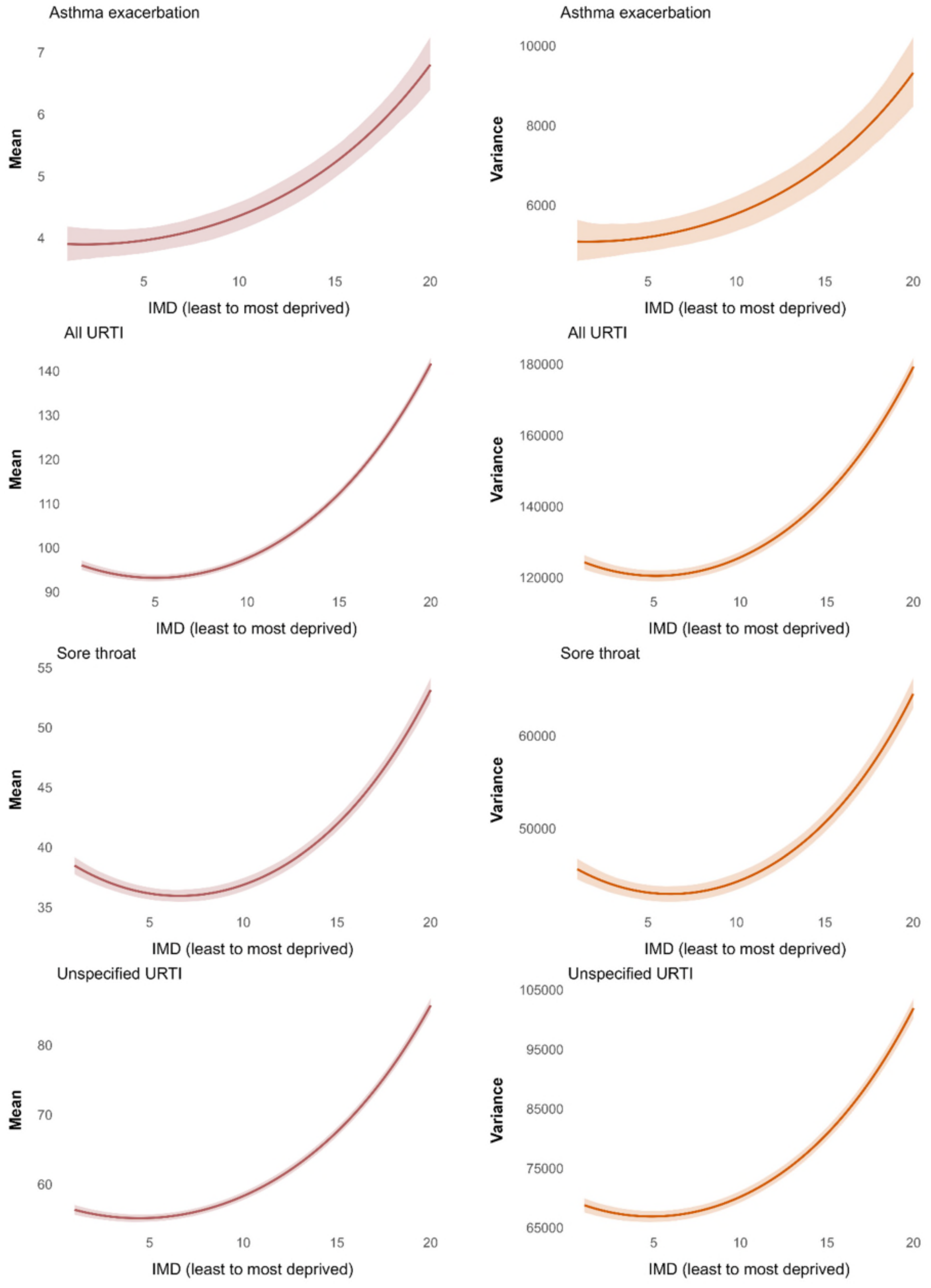

*Figure S4 (a). Consultation rate (per 1000 person-years) in children - mean and variance (continued)*

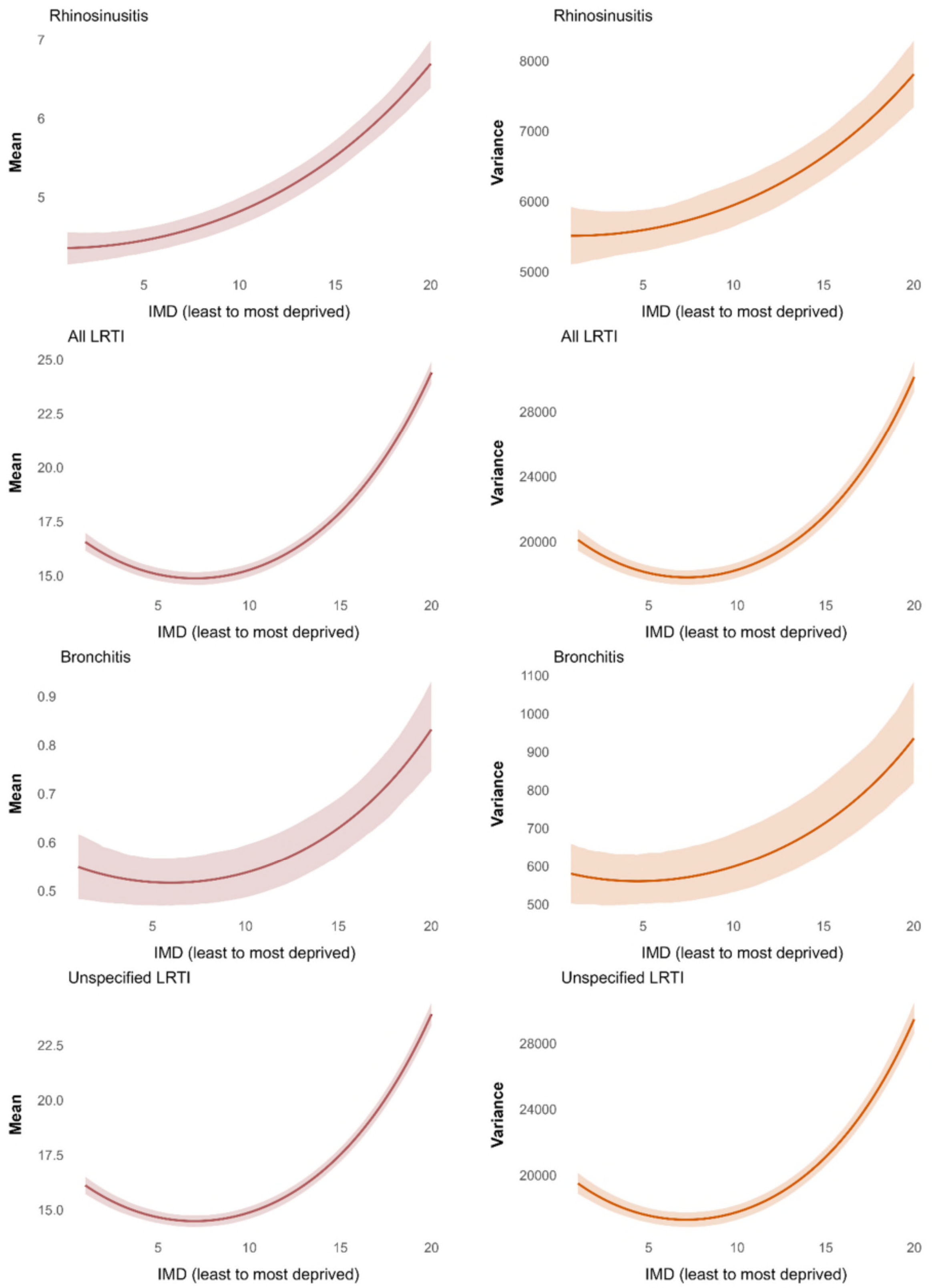

*Figure S4 (a). Consultation rate (per 1000 person-years) in children - mean and variance (continued)*

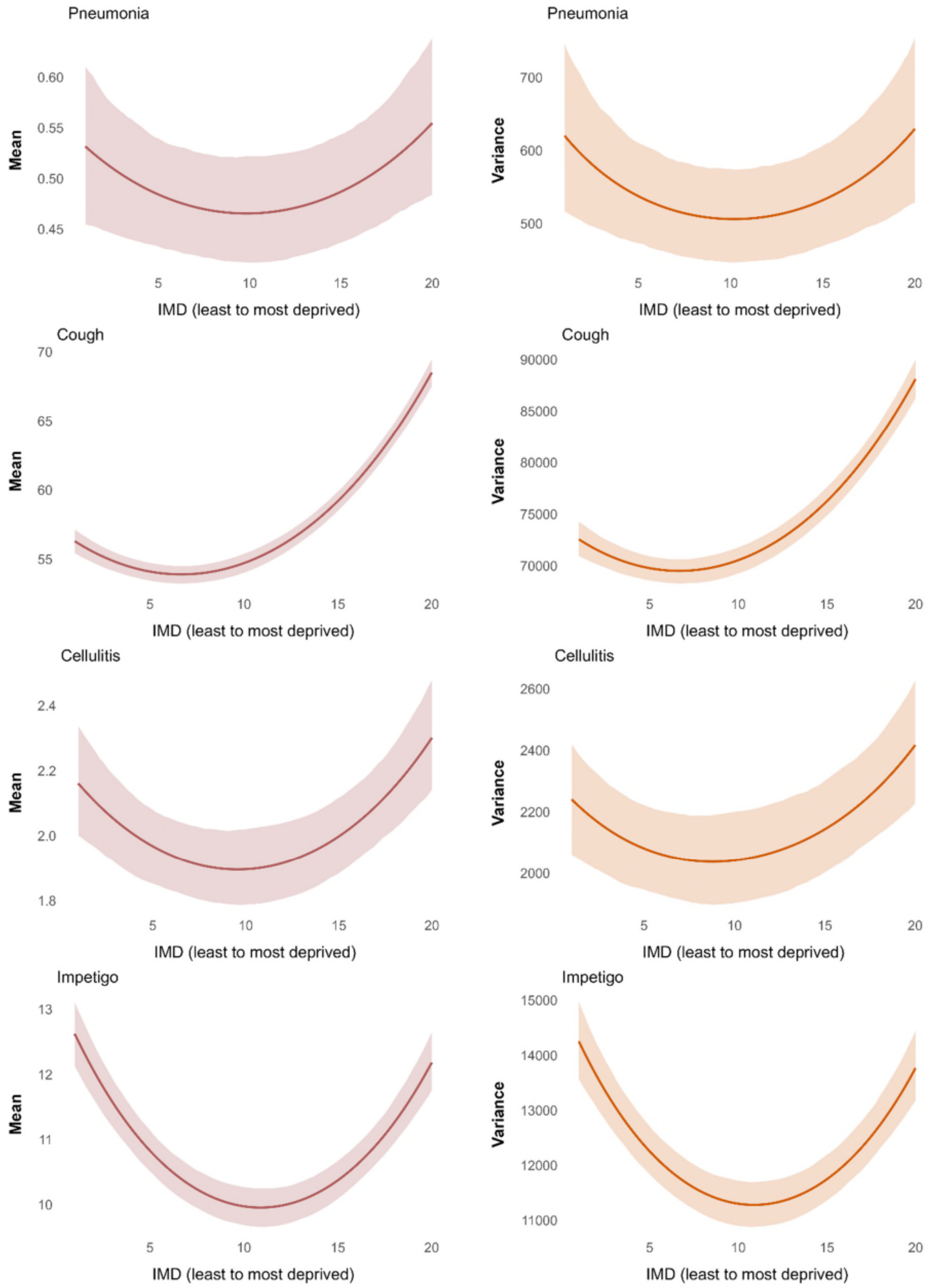

*Figure S4 (a). Consultation rate (per 1000 person-years) in children - mean and variance (continued)*

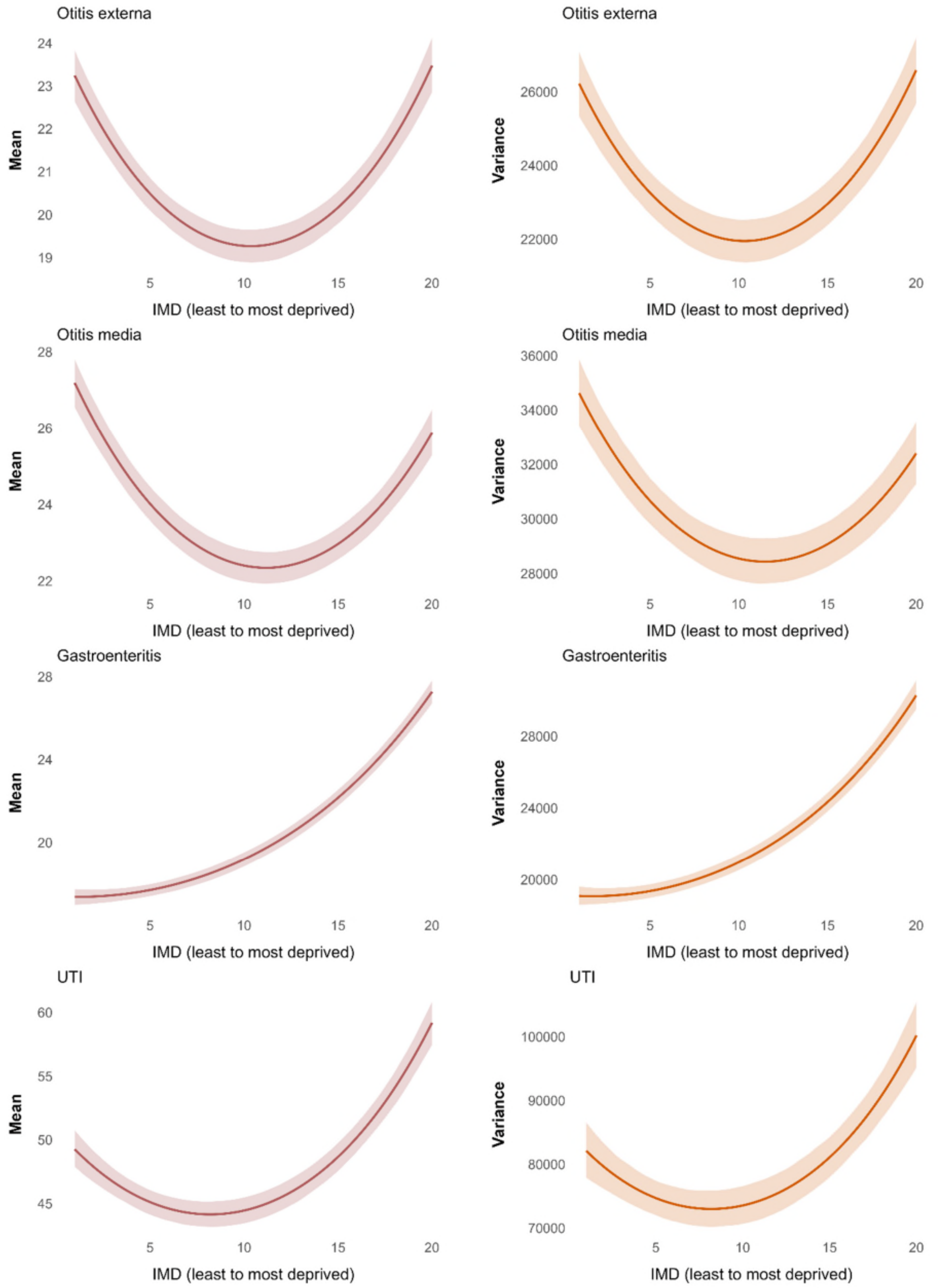

Panel (b) Adults

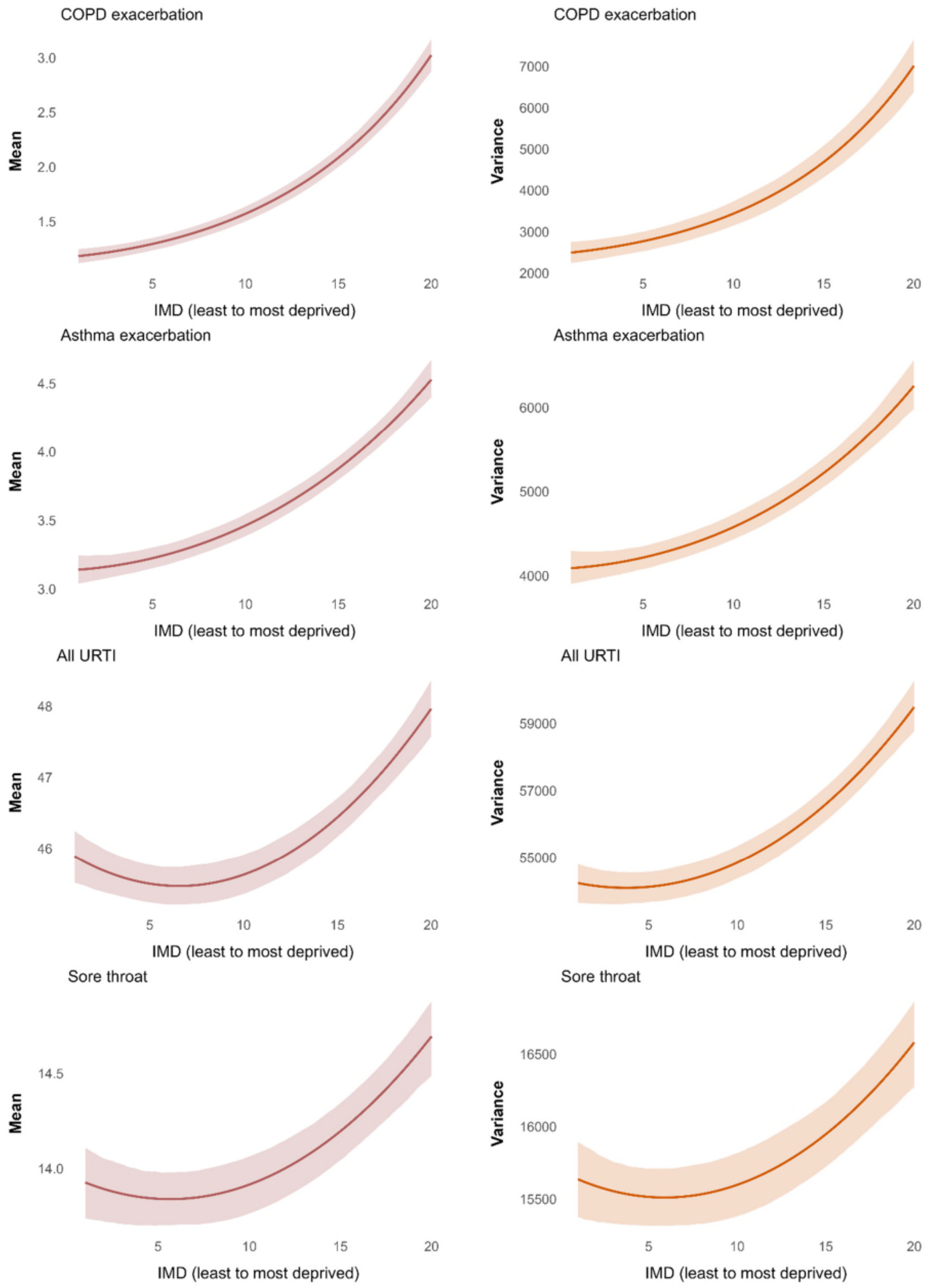

*Figure S4 (b). Consultation rate (per 1000 person-years) in adults - mean and variance (continued)*

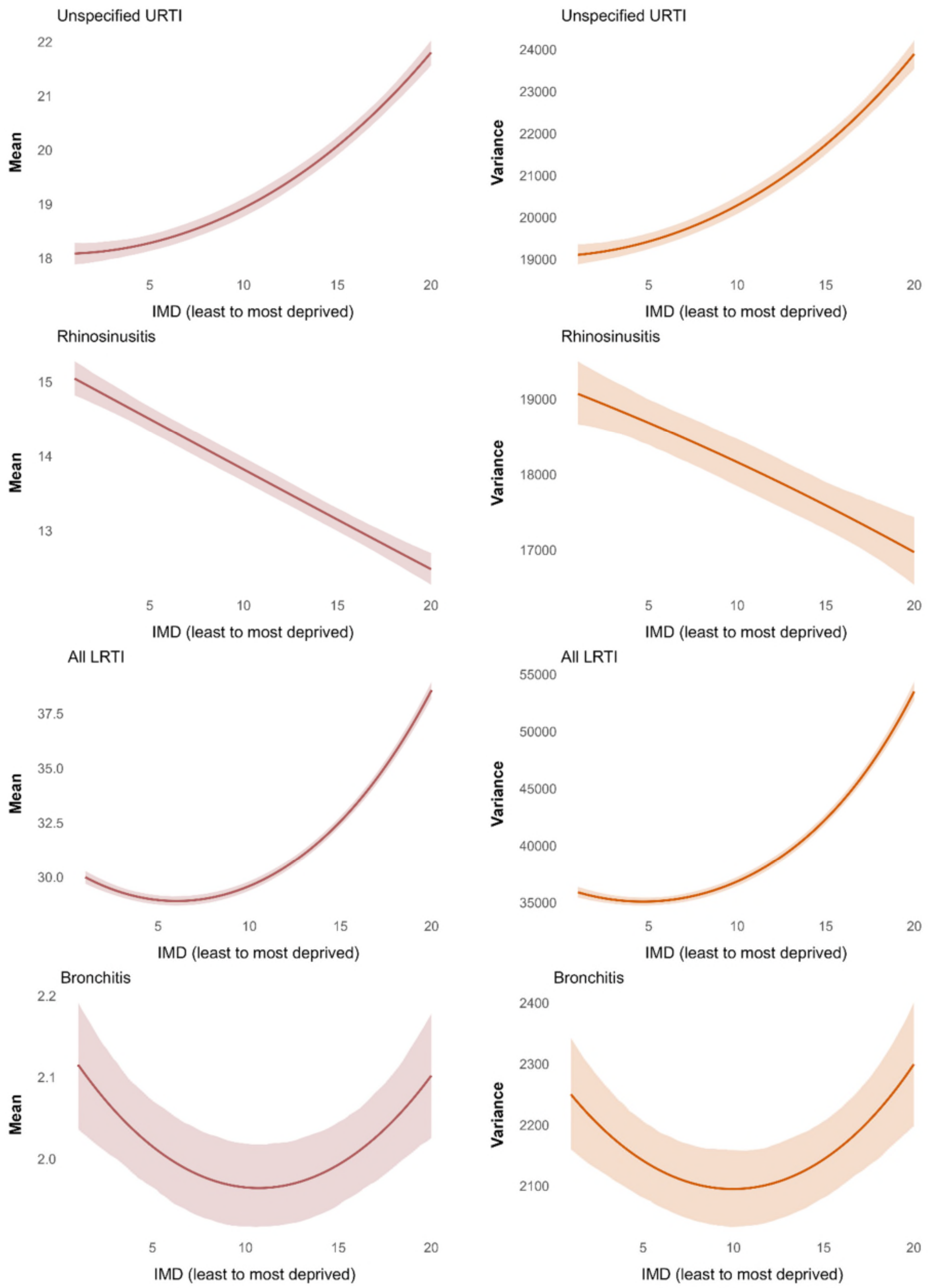

*Figure S4 (b). Consultation rate (per 1000 person-years) in adults - mean and variance (continued)*

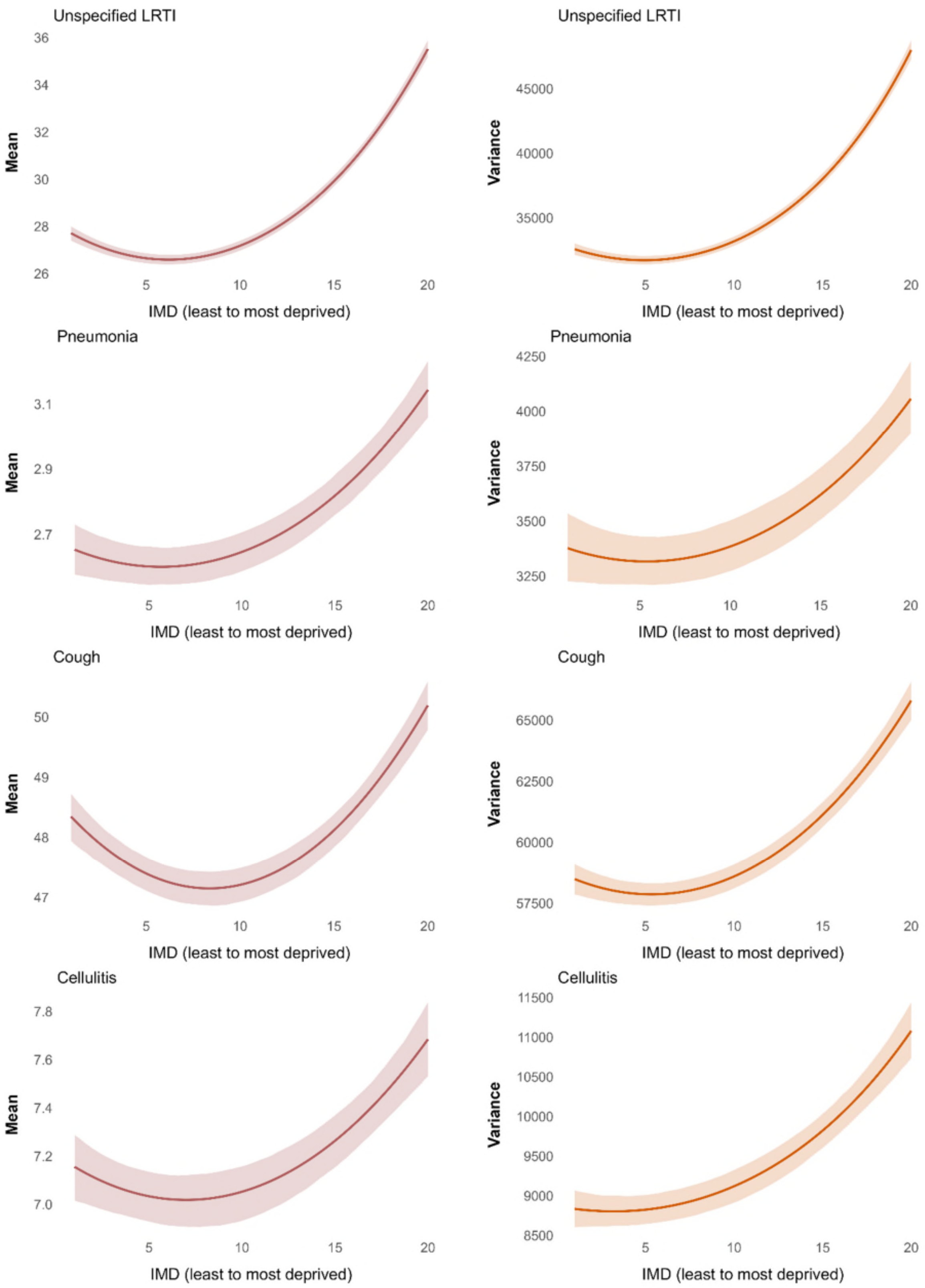

*Figure S4 (b). Consultation rate (per 1000 person-years) in adults - mean and variance (continued)*

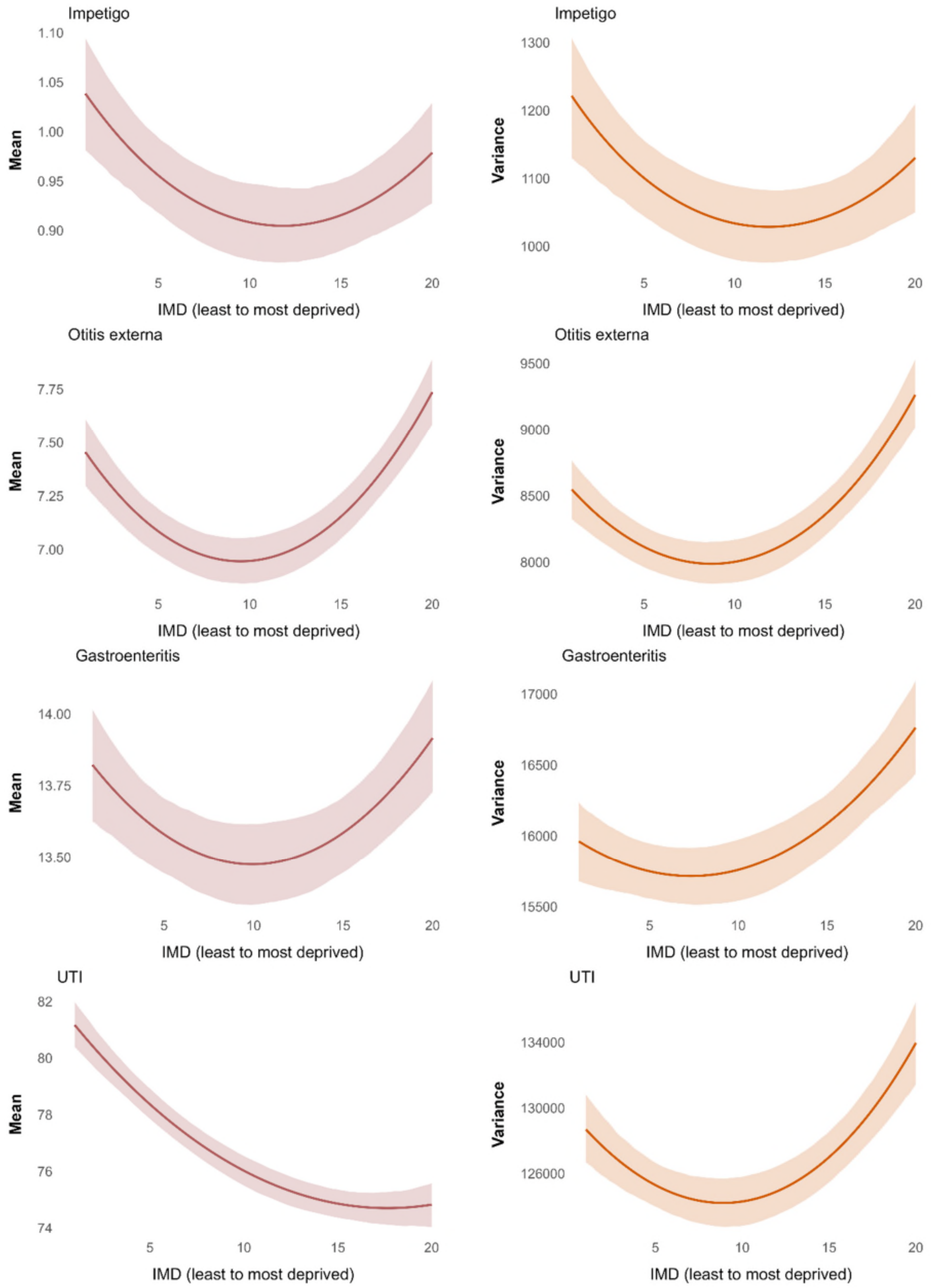

*Figure S4 (b). Consultation rate (per 1000 person-years) in adults - mean and variance (continued)*

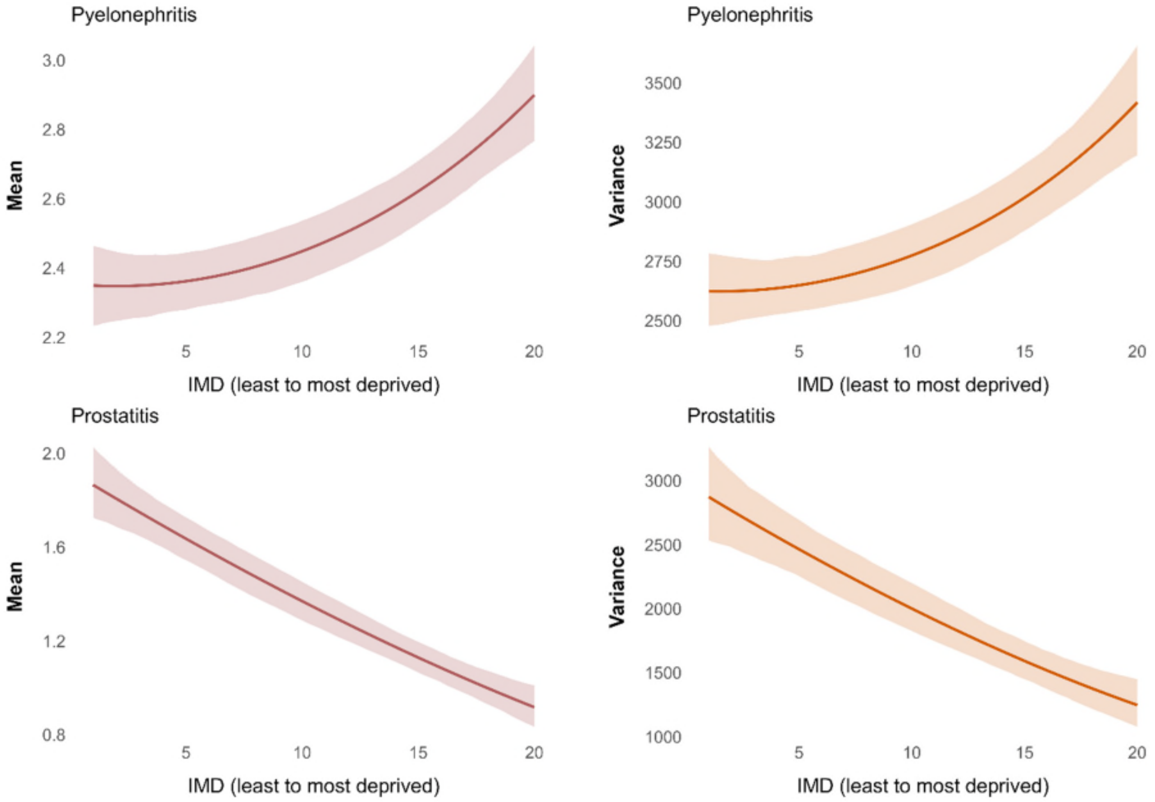


Notes: All models were adjusted for age, sex, and ethnicity. Abbreviations: IMD index of multiple deprivation, COPD chronic obstructive pulmonary disease, LRTI lower respiratory tract infections, URTI upper respiratory tract infections, UTI urinary tract infections. “All URTI” includes sore throat, unspecified upper respiratory tract infection, rhinosinusitis whereas “All LRTI” includes bronchitis, unspecified lower respiratory tract infection, pneumonia.

*Figure S5 Association between IMD and prescribing rates, consultation rates, and probability of receiving antibiotics once consulted (mean measure with shaded 95% confidence intervals reported).*

Panel (a) Children

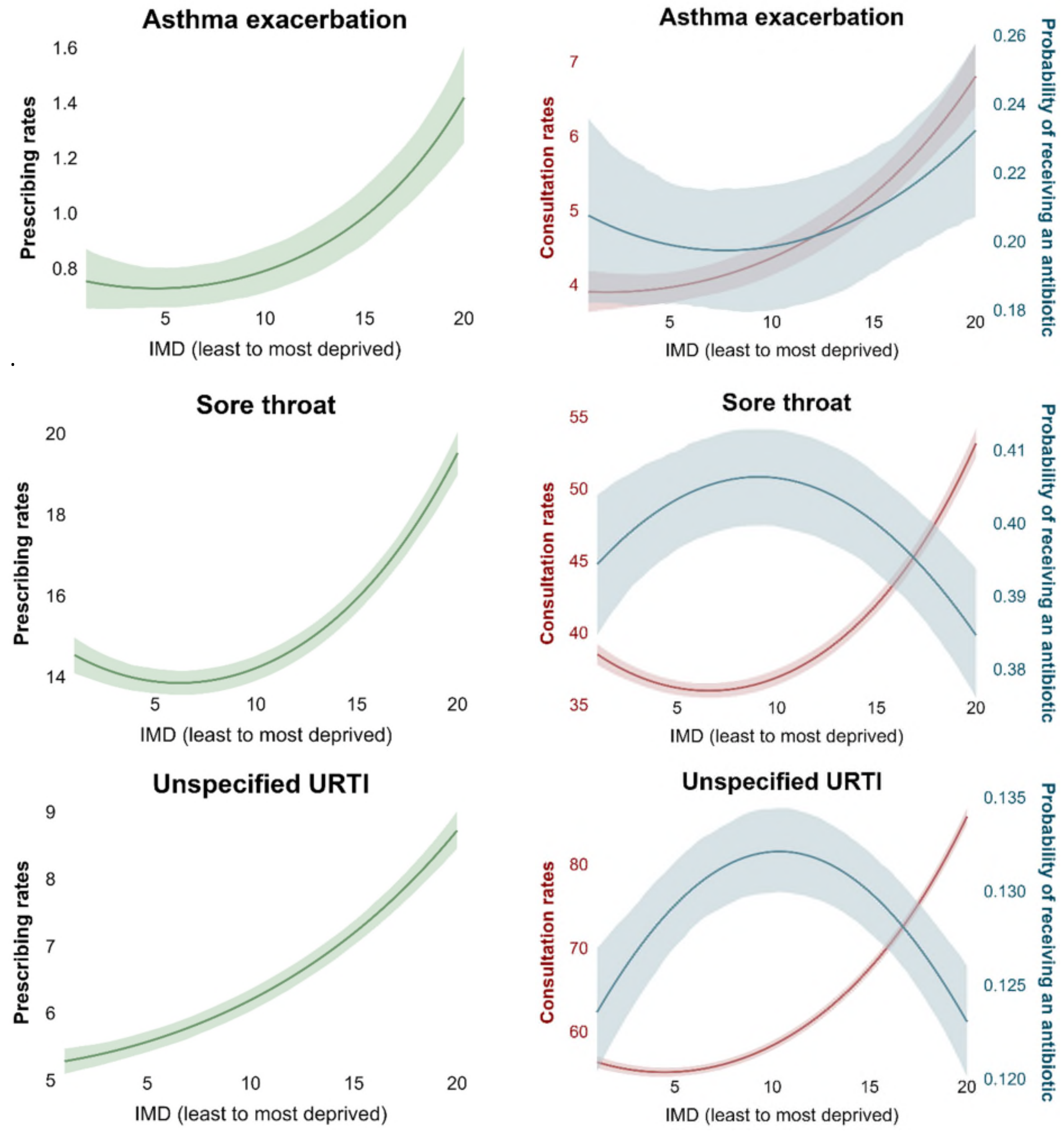

*Figure S5 (a). Mean prescribing rates, consultation rates, and probability of receiving antibiotics once consulted in children (continued)*

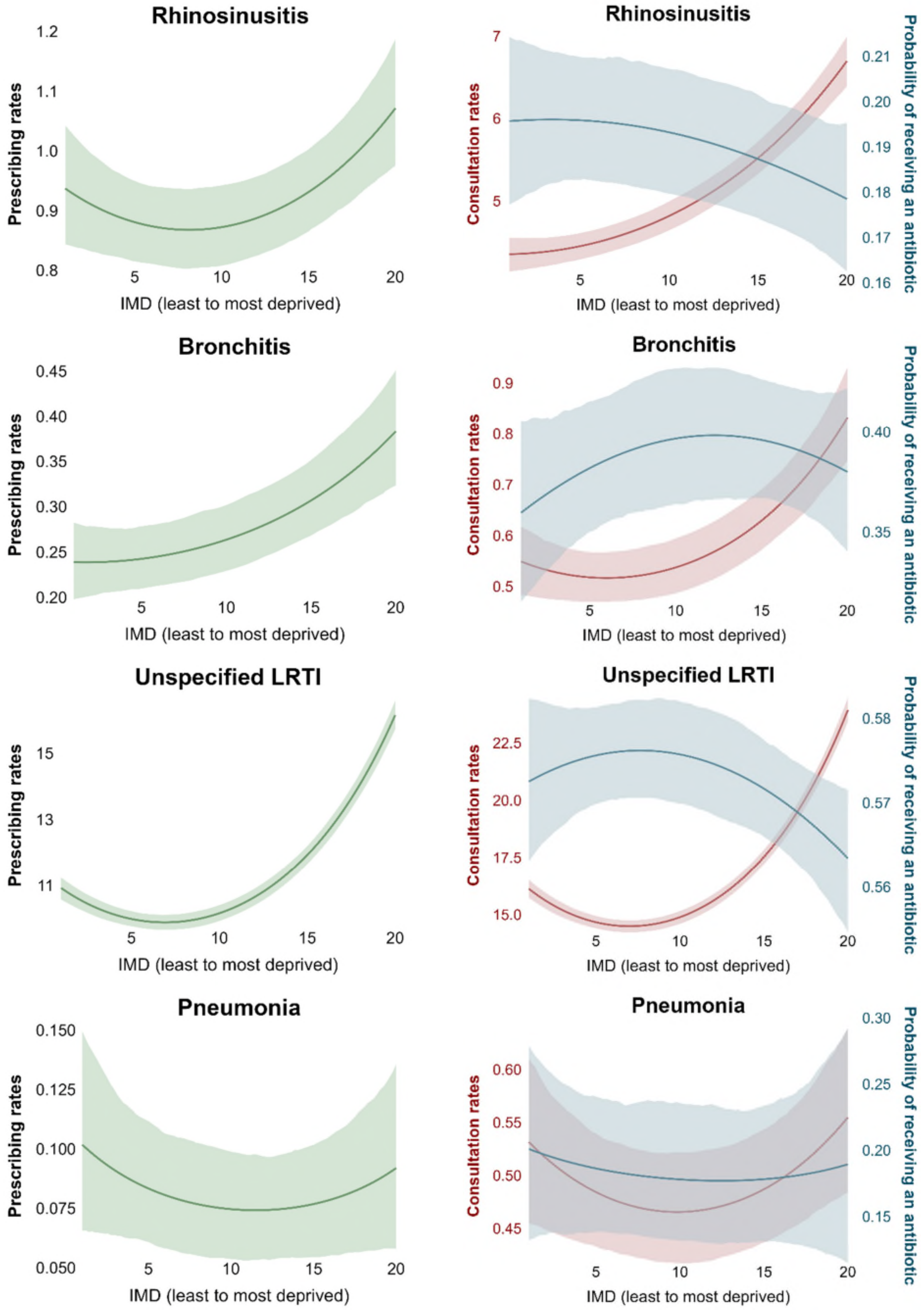

*Figure S5 (a). Mean prescribing rates, consultation rates, and probability of receiving antibiotics once consulted in children (continued)*

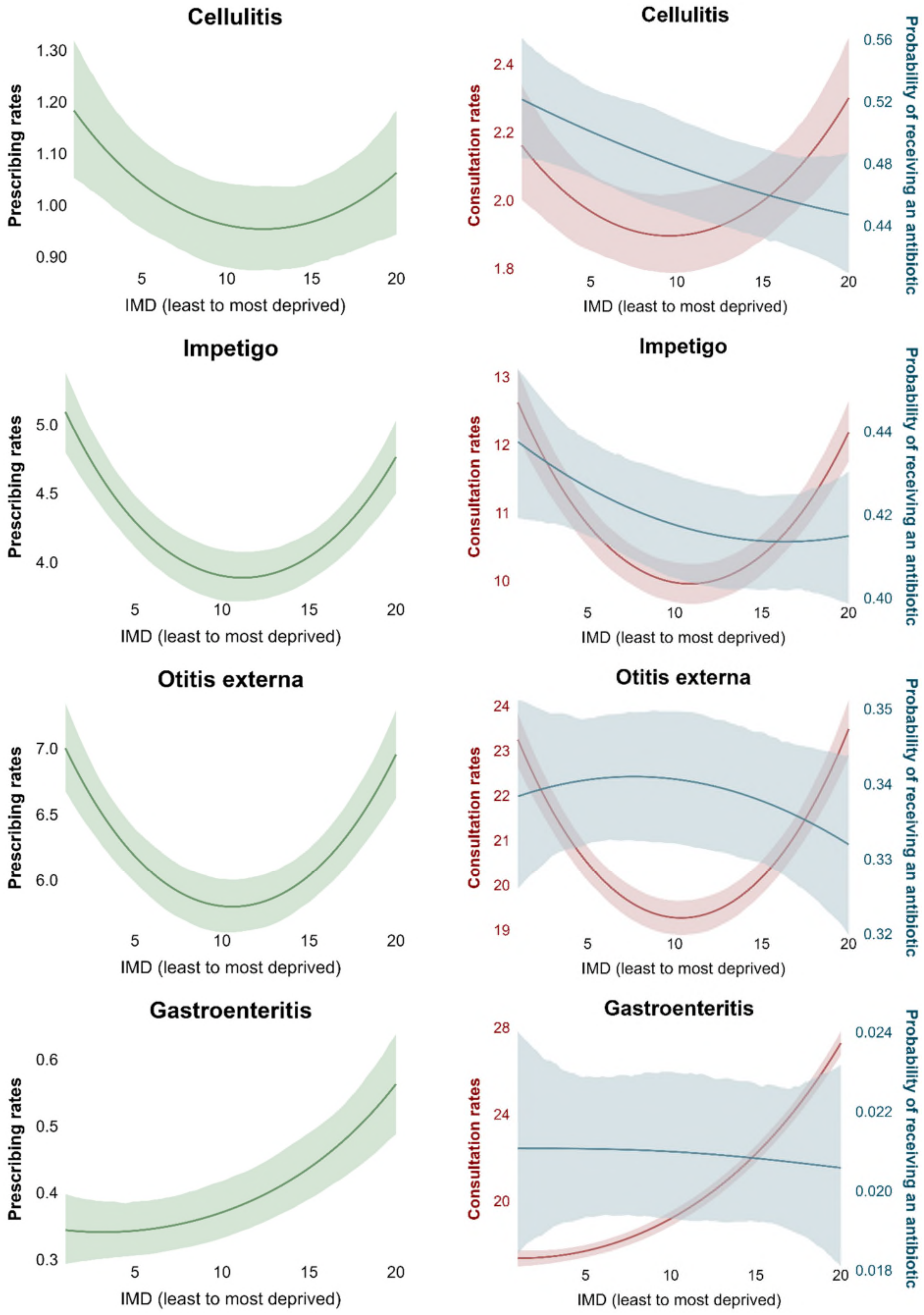

*Figure S5 (a). Mean prescribing rates, consultation rates, and probability of receiving antibiotics once consulted in children (continued)*

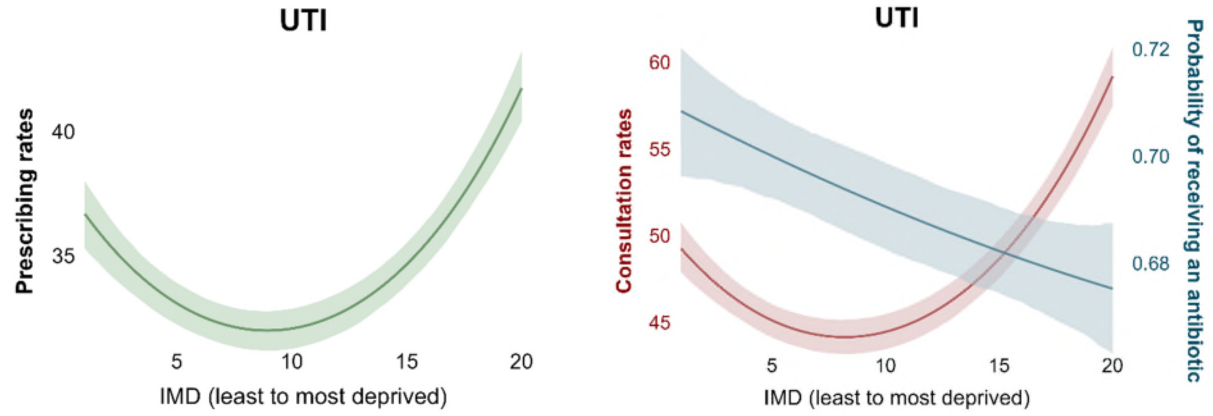


Panel (b) Adults

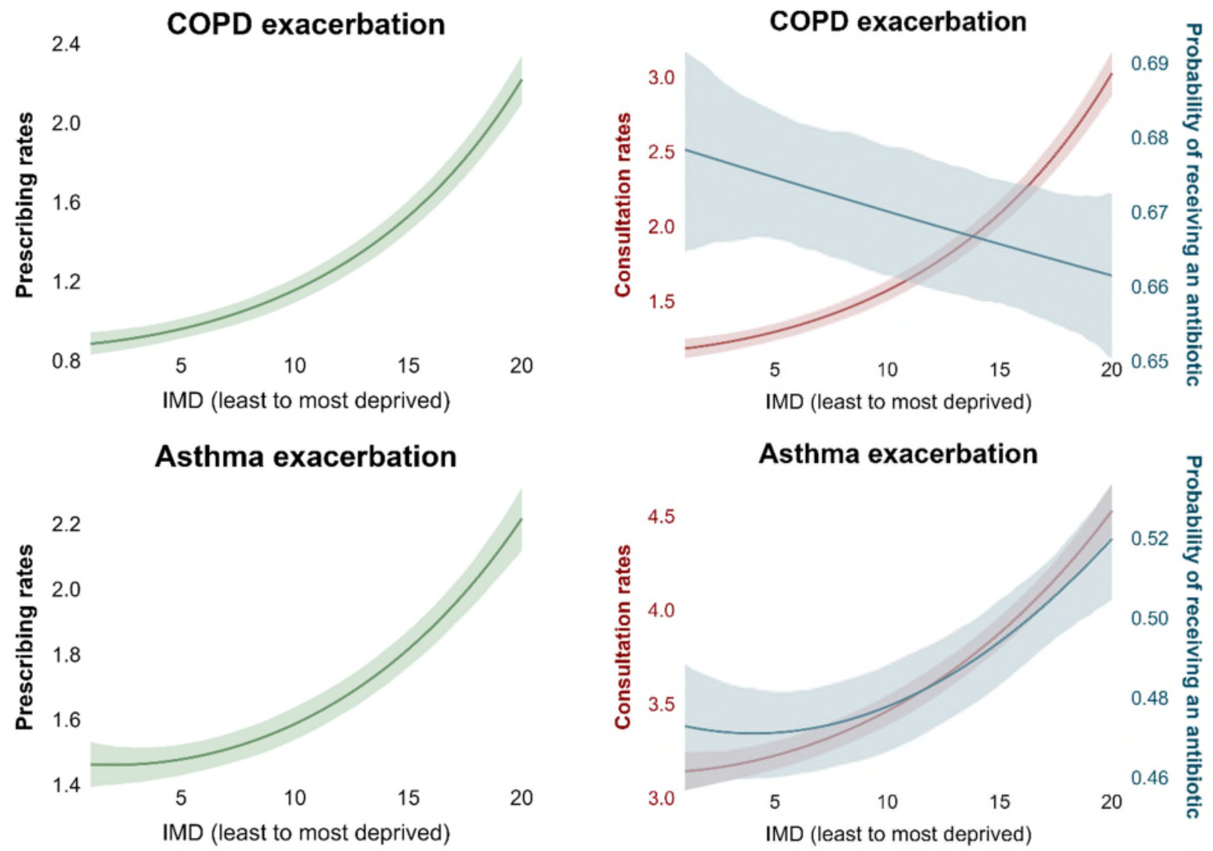

*Figure S5 (b). Mean prescribing rates, consultation rates, and probability of receiving antibiotics once consulted in adults (continued)*

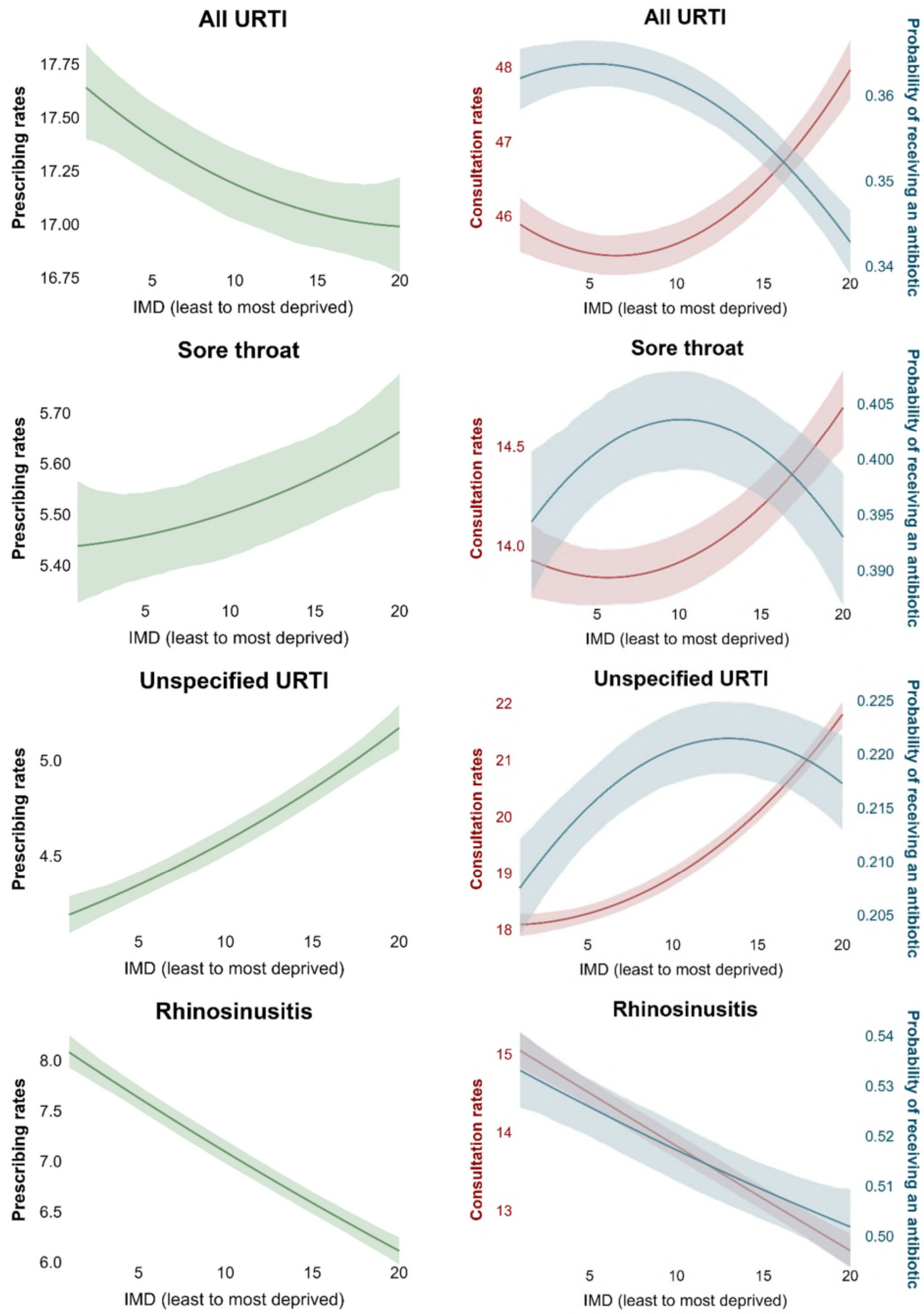

*Figure S5 (b). Mean prescribing rates, consultation rates, and probability of receiving antibiotics once consulted in adults (continued)*

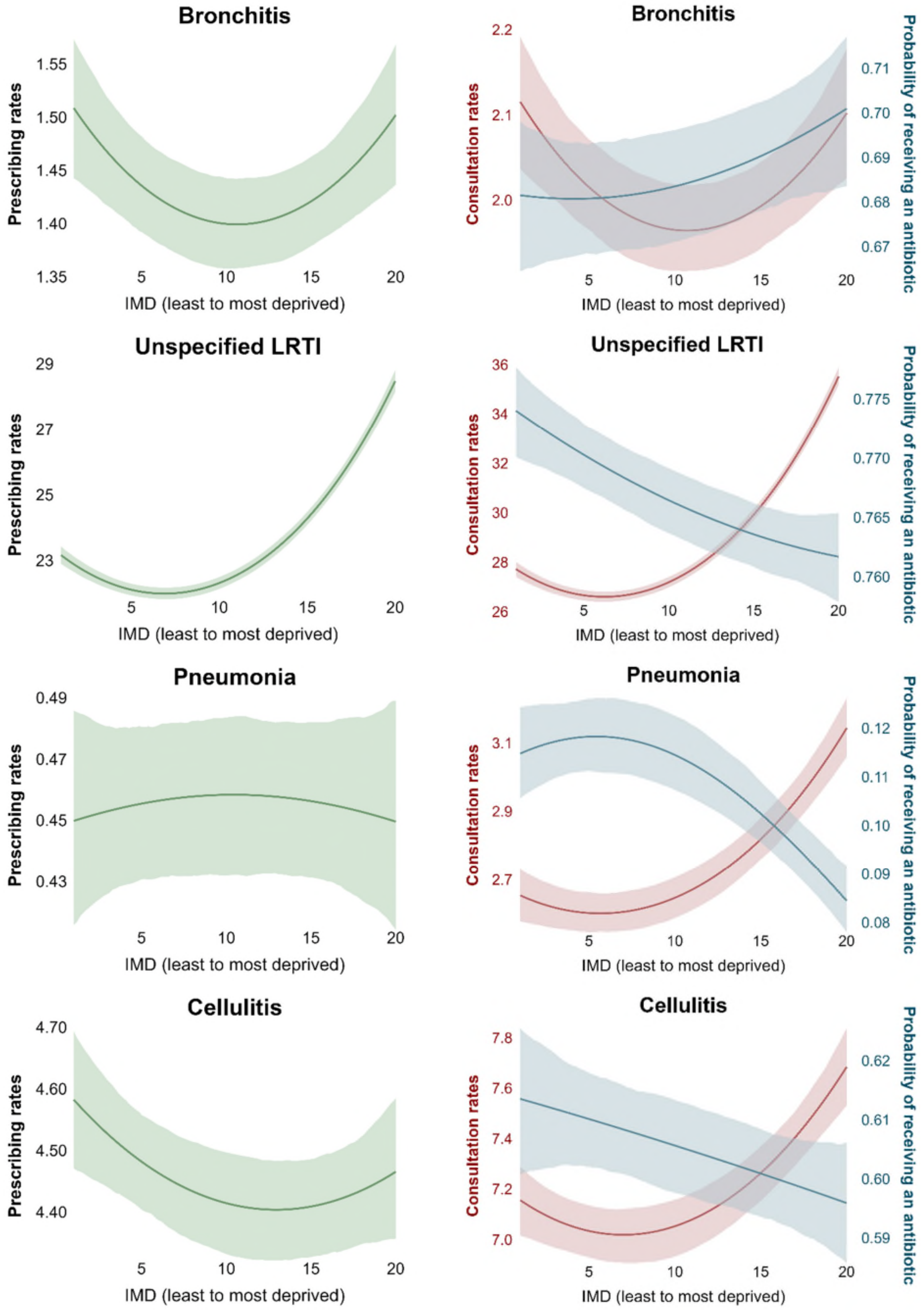

*Figure S5 (b). Mean prescribing rates, consultation rates, and probability of receiving antibiotics once consulted in adults (continued)*

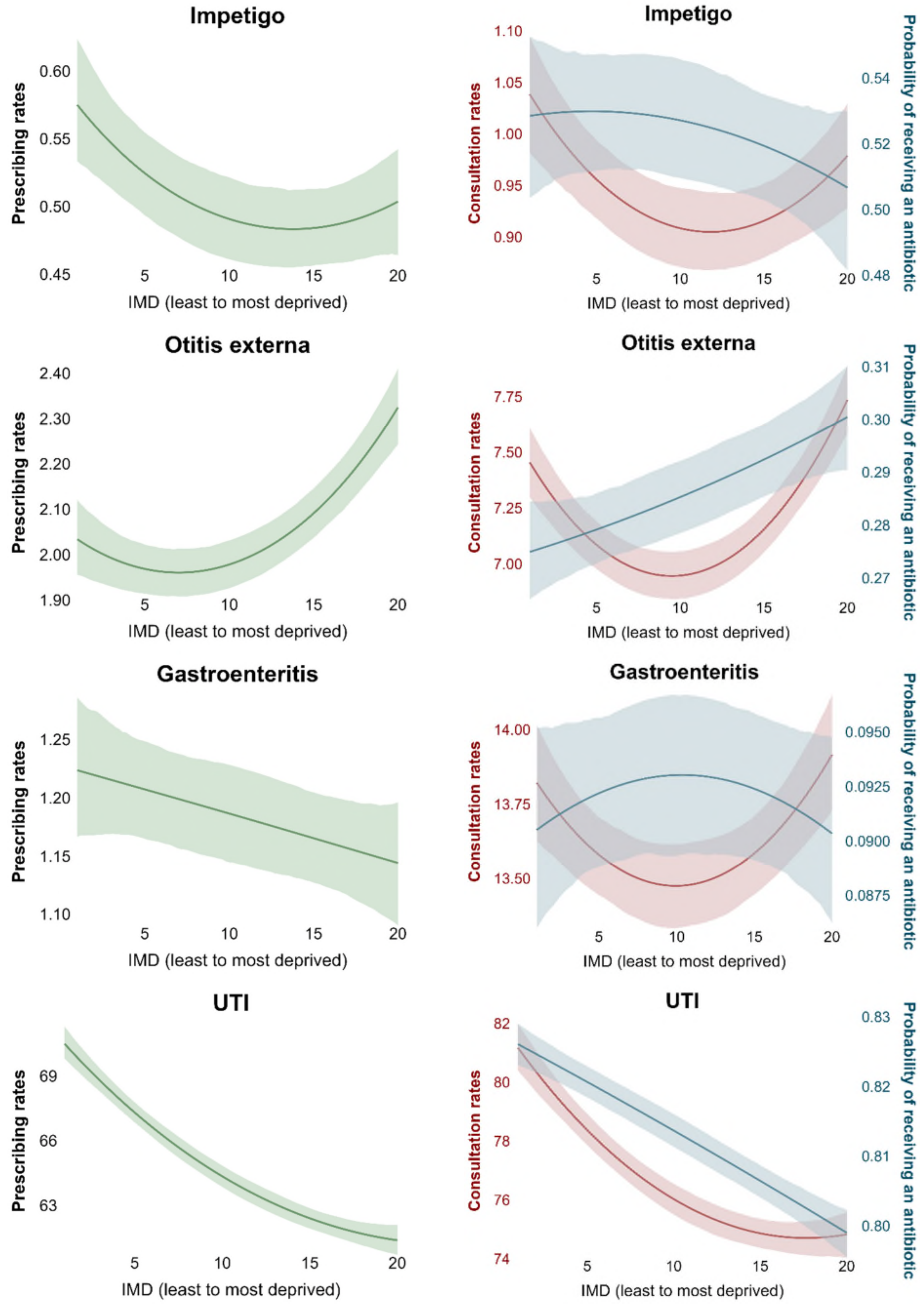

*Figure S5 (b). Mean prescribing rates, consultation rates, and probability of receiving antibiotics once consulted in adults (continued)*

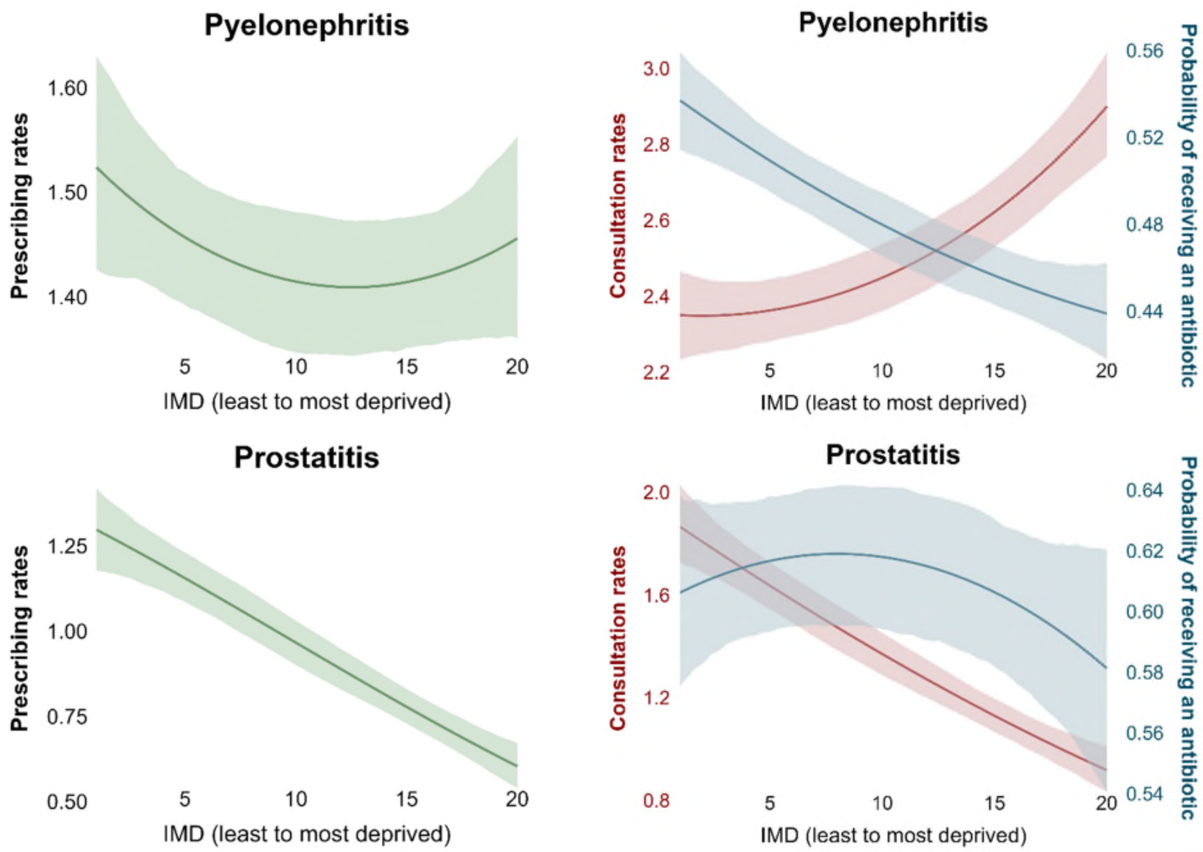


Notes: All models were adjusted for age, sex, and ethnicity. Abbreviations: IMD index of multiple deprivation, COPD chronic obstructive pulmonary disease, LRTI lower respiratory tract infections, URTI upper respiratory tract infections, UTI urinary tract infections. "All URTI" includes sore throat, unspecified upper respiratory tract infection, rhinosinusitis whereas "All LRTI" includes bronchitis, unspecified lower respiratory tract infection, pneumonia.

##### S3.2.3 Role of contributors in explaining inequalities

*Figure S6 Role of contributors in explaining inequalities in consultations*

Panel (a) Children

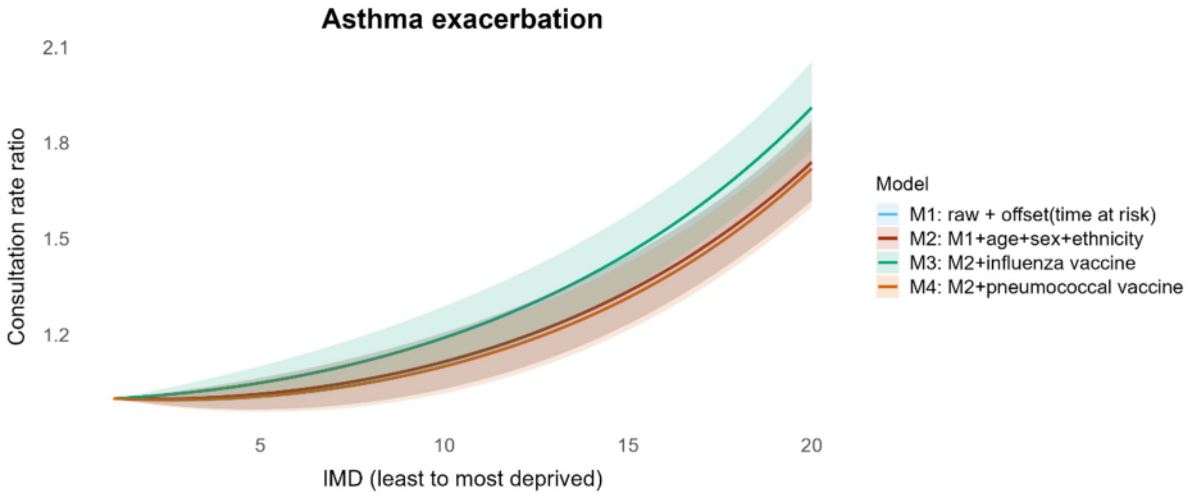


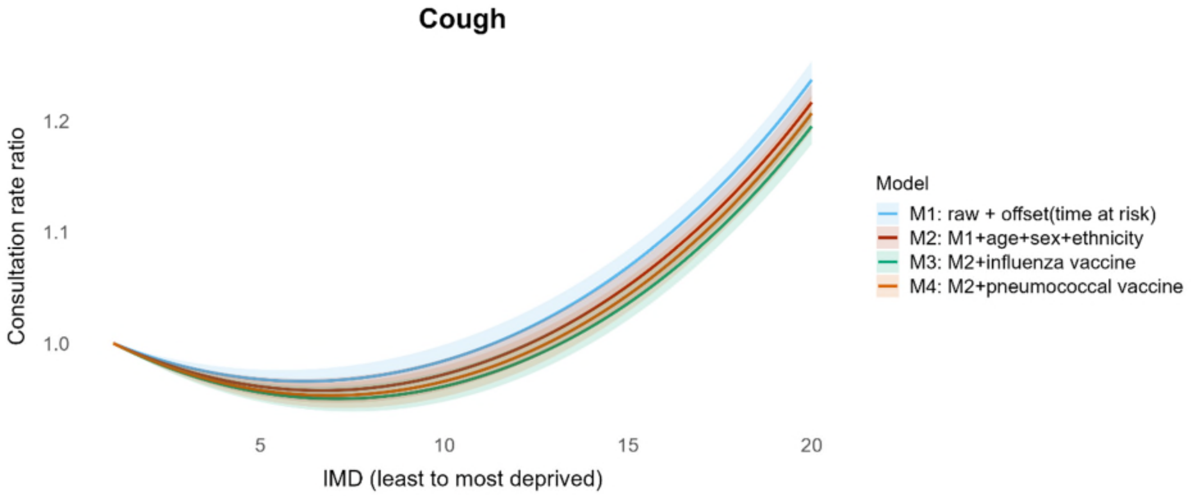


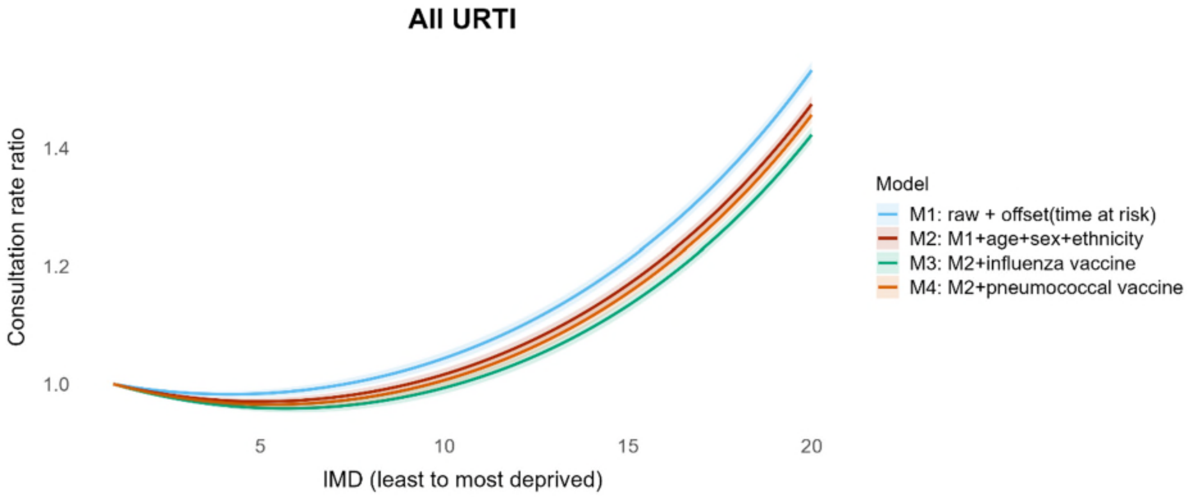

*Figure S6 (a).* Role of contributors in explaining inequalities in consultations *in children (continued)*

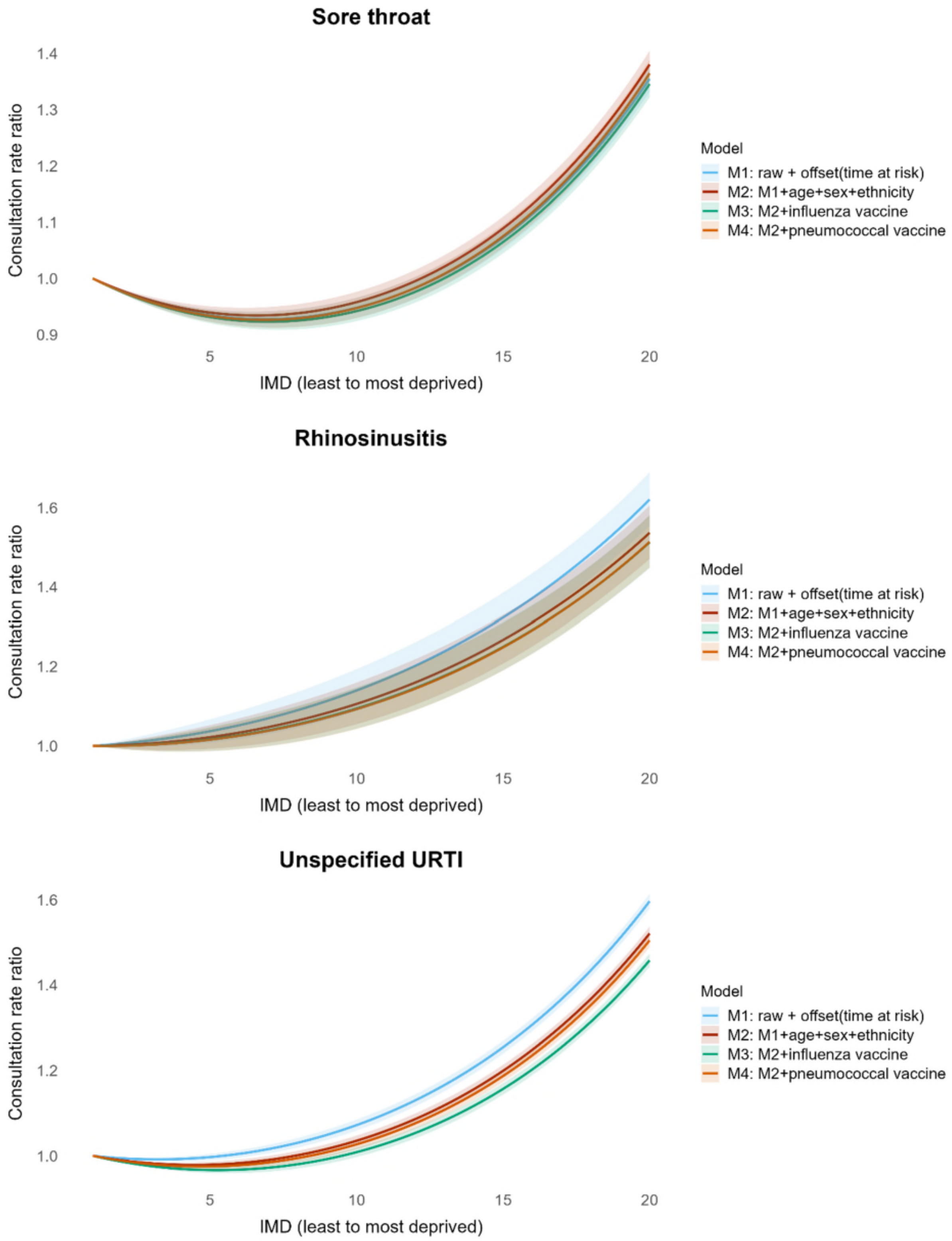

*Figure S6 (a).* Role of contributors in explaining inequalities in consultations *in children (continued)*

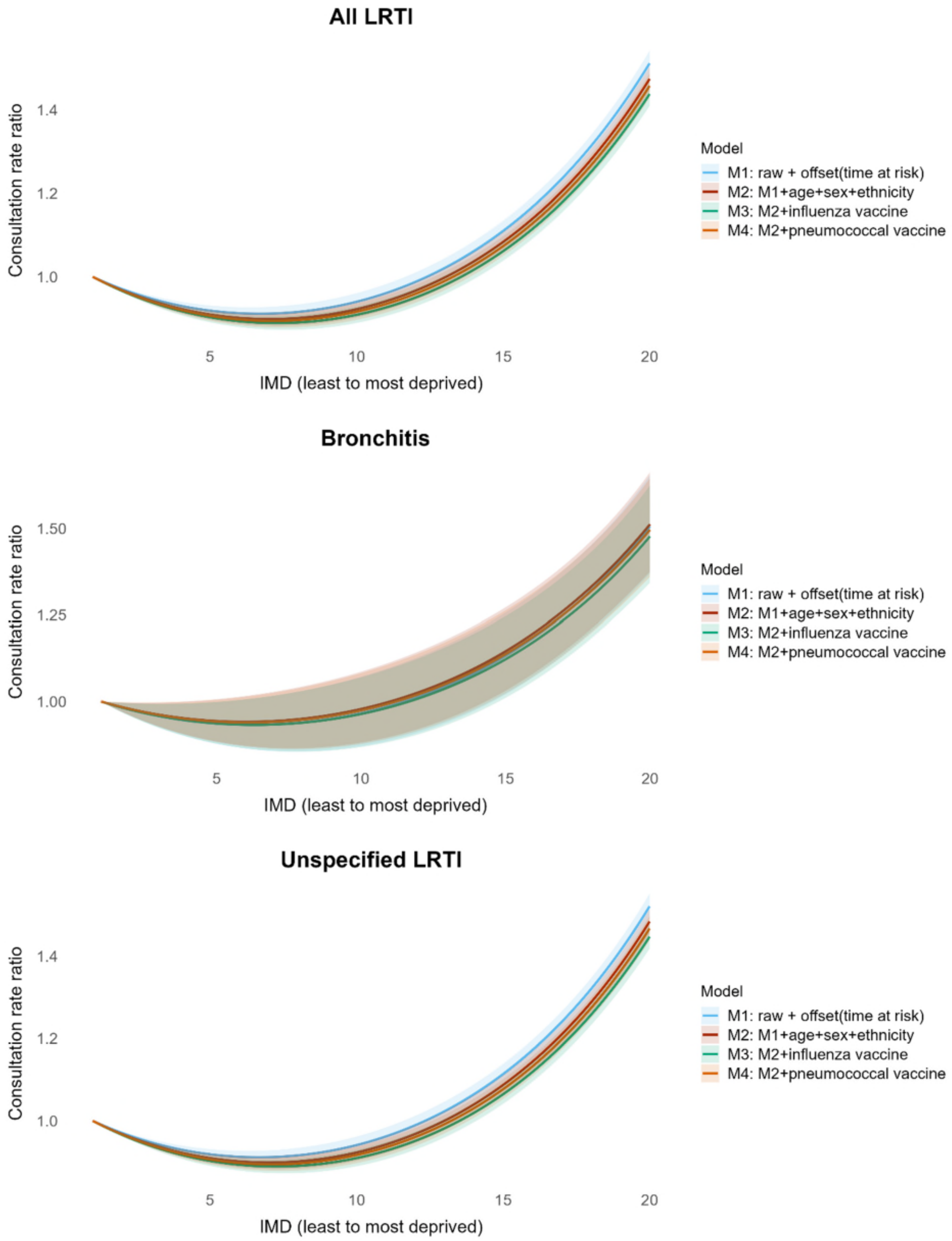

Panel (b) Adults

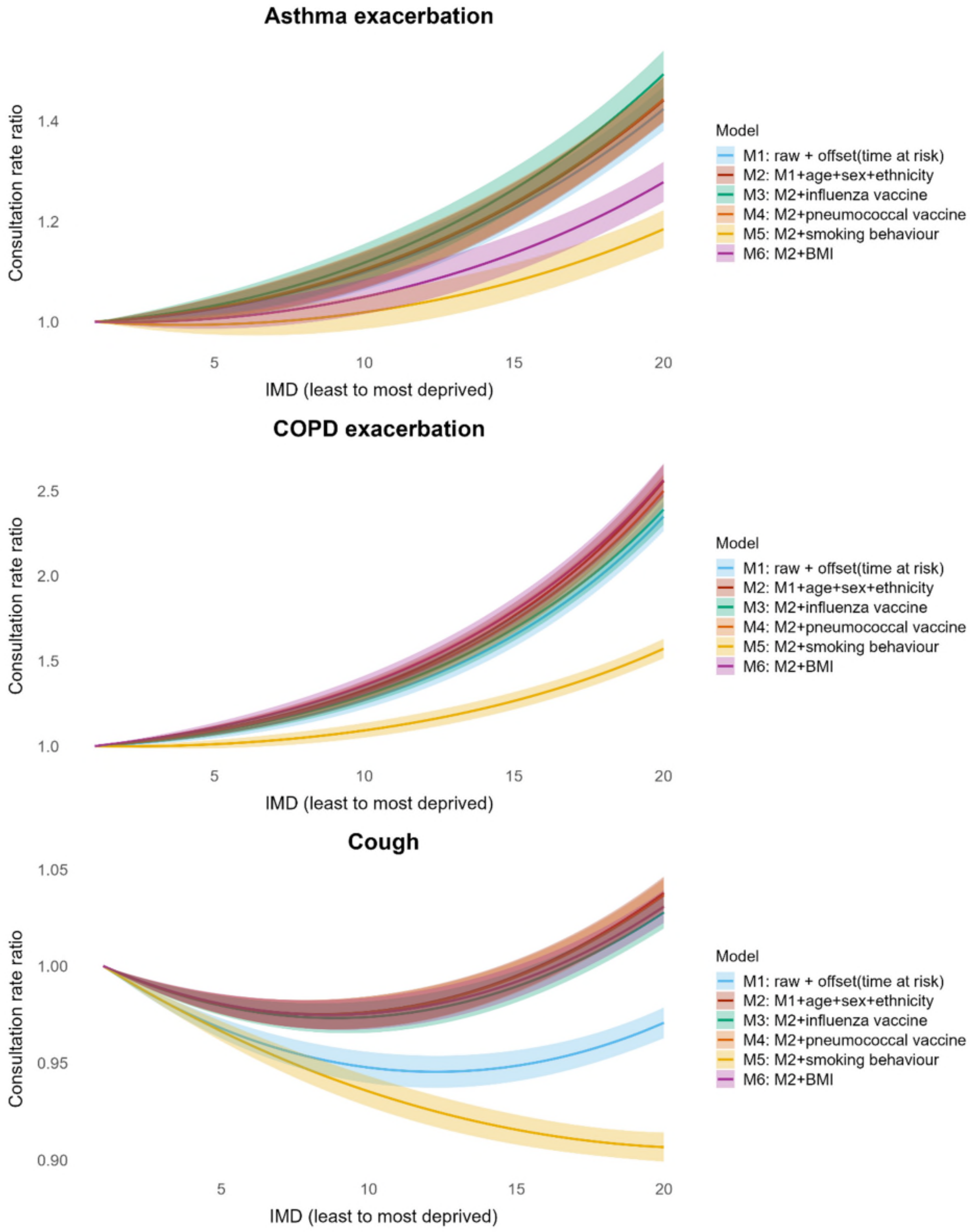

*Figure S6 (b).* Role of contributors in explaining inequalities in consultations *in adults (continued)*

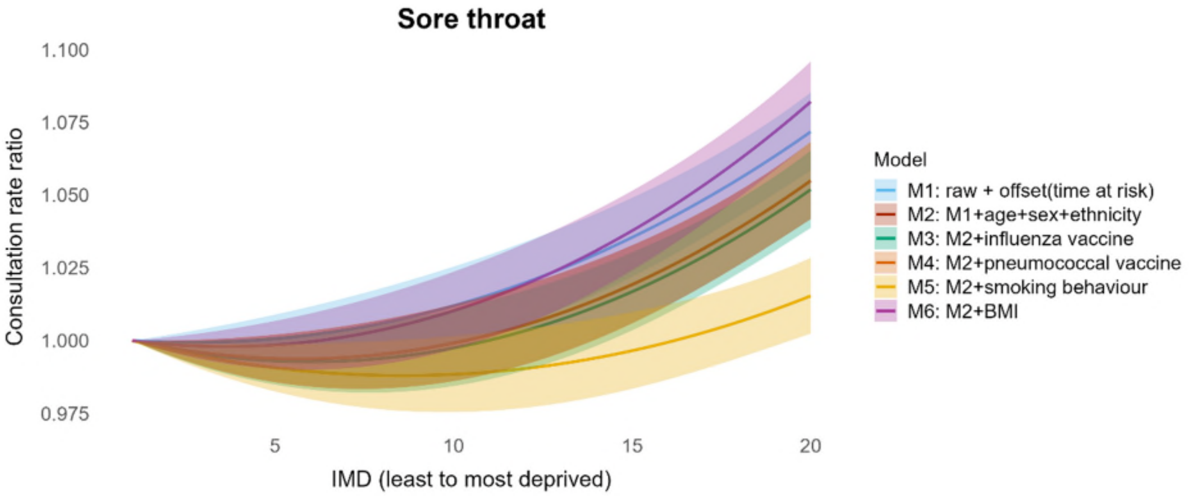


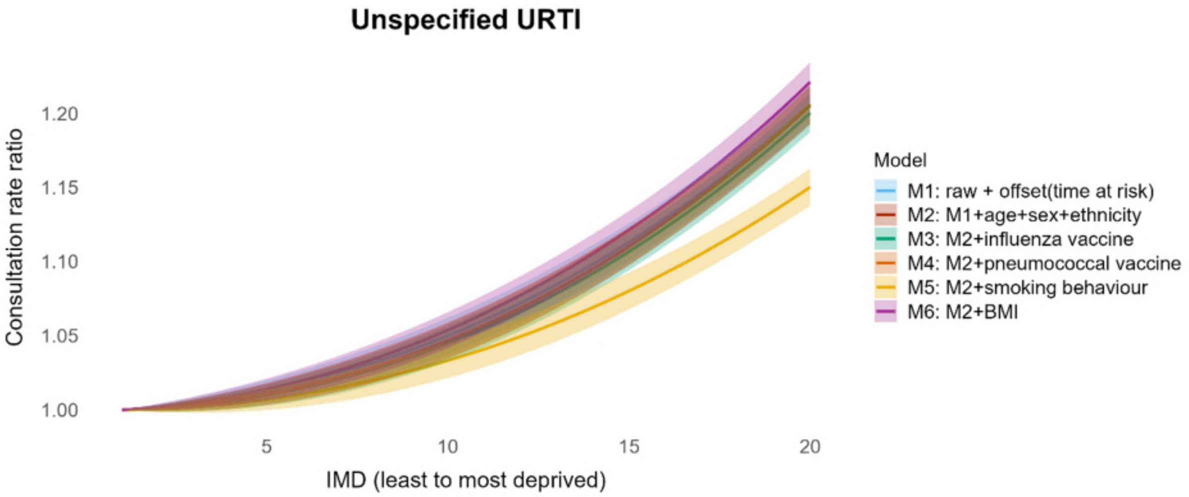


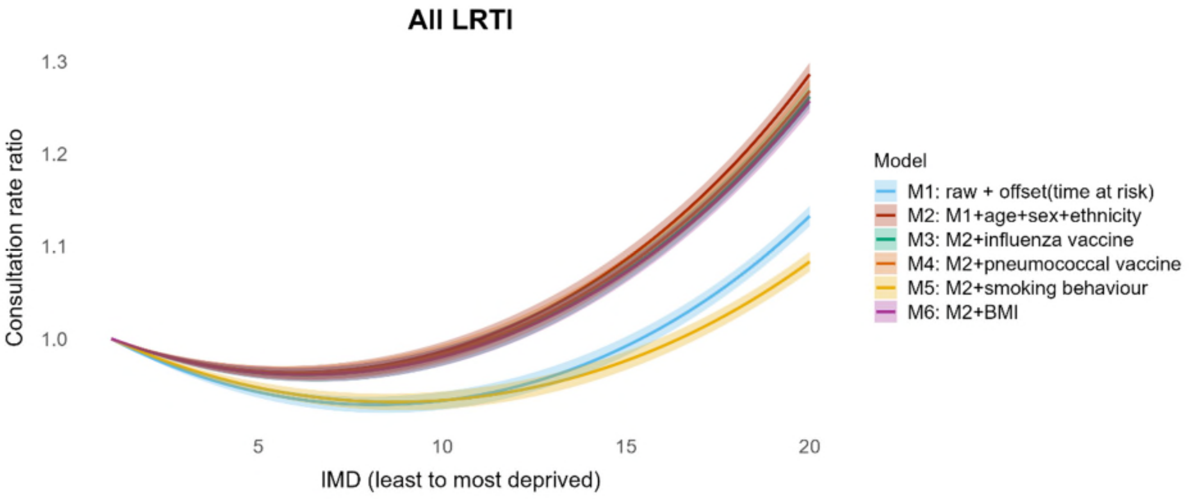

*Figure S6 (b).* Role of contributors in explaining inequalities in consultations *in adults (continued)*

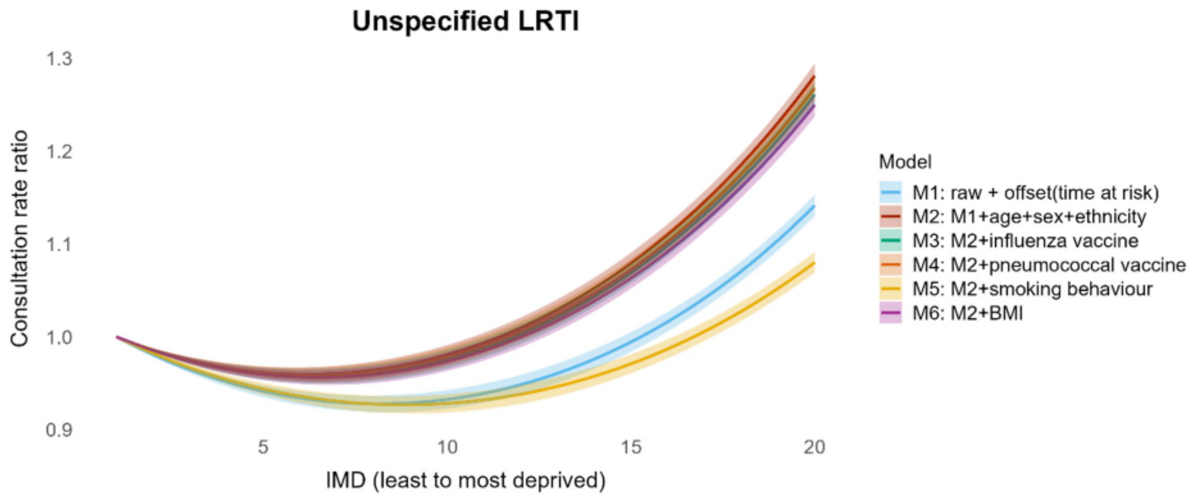


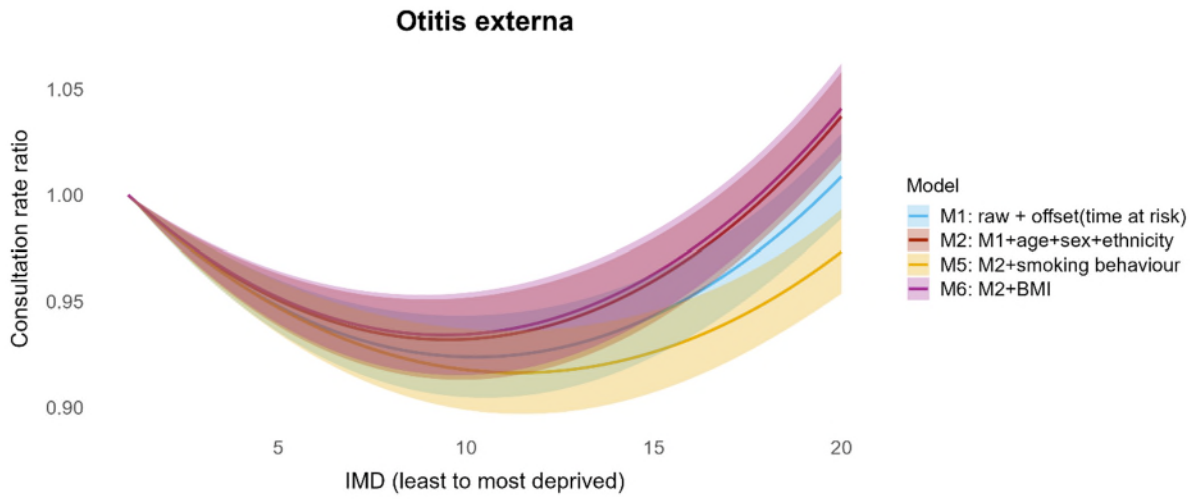


Notes: Figure S6 reports the isolated IMD association and consultation rates, interpreted as the rate ratio of mean consultation rates (per person-day) across IMD, relative to the least deprived level (IMD=1). Shaded areas indicate 95% confidence intervals. This ratio-based scale arises because in the negative binomial models fitted with GAMLSS, the mean parameter is linked to IMD via a log link. Exponentiating the model coefficients gives relative differences (rate ratios) rather than absolute rates, using the least deprived level as the reference. A description of the corresponding statistical method is provided in supplementary methods S1.4. Compared with baseline model controlling for age, sex, and ethnicity (M2), a flatter slope after adjusting for the contributor indicates that this contributor partly explained inequalities in the consultation rates. The crude model (M1) should be interpreted with caution, as differences in observed inequalities between the crude and baseline model may reflect differences in population structure across deprivation groups rather than deprivation itself.

#### S3.3 Sensitivity analysis: repeating analyses using observed population

##### S3.3.1 Comparison between observed and total population

*Figure S7 Proportion of patients by IMD twentile, age group, sex, and ethnicity for children*

Panel (a) IMD twentile

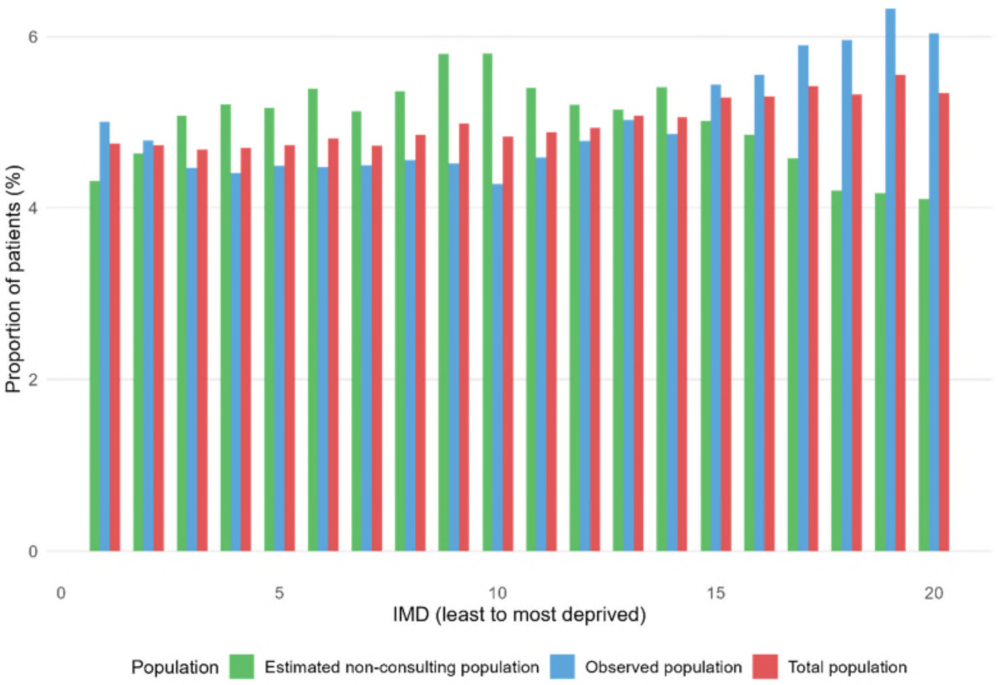


Panel (b) Age group

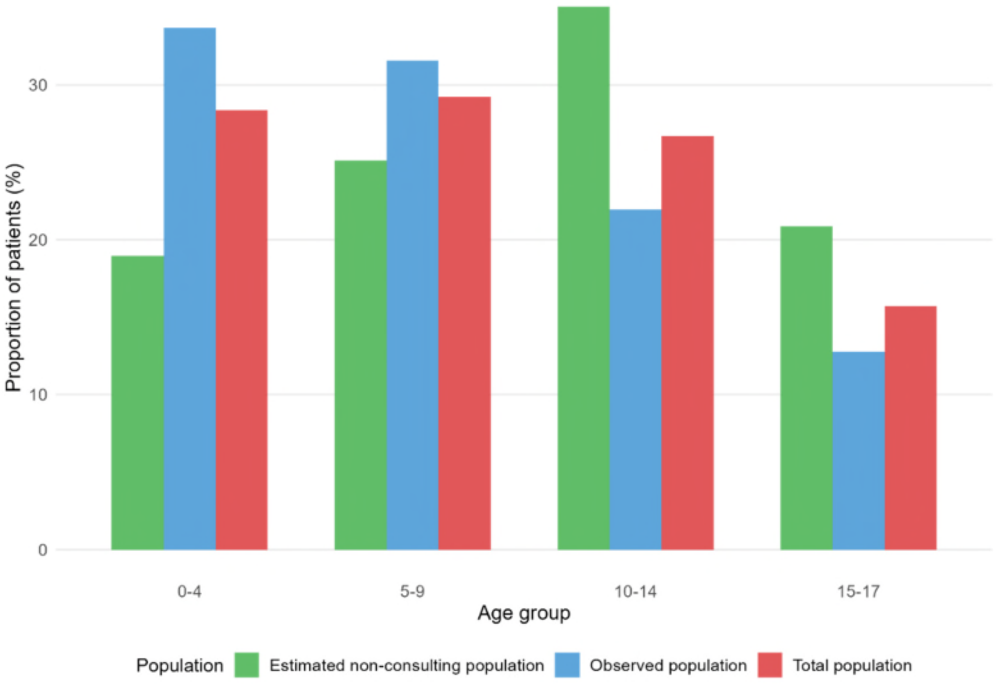

Panel (c) Sex

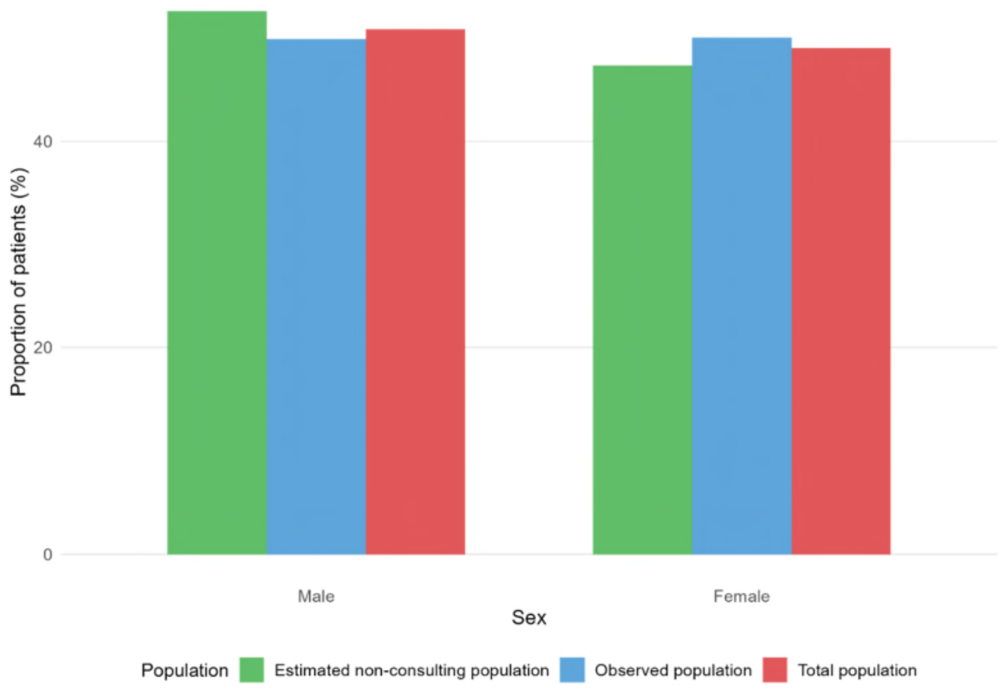


Panel (d) Ethnicity

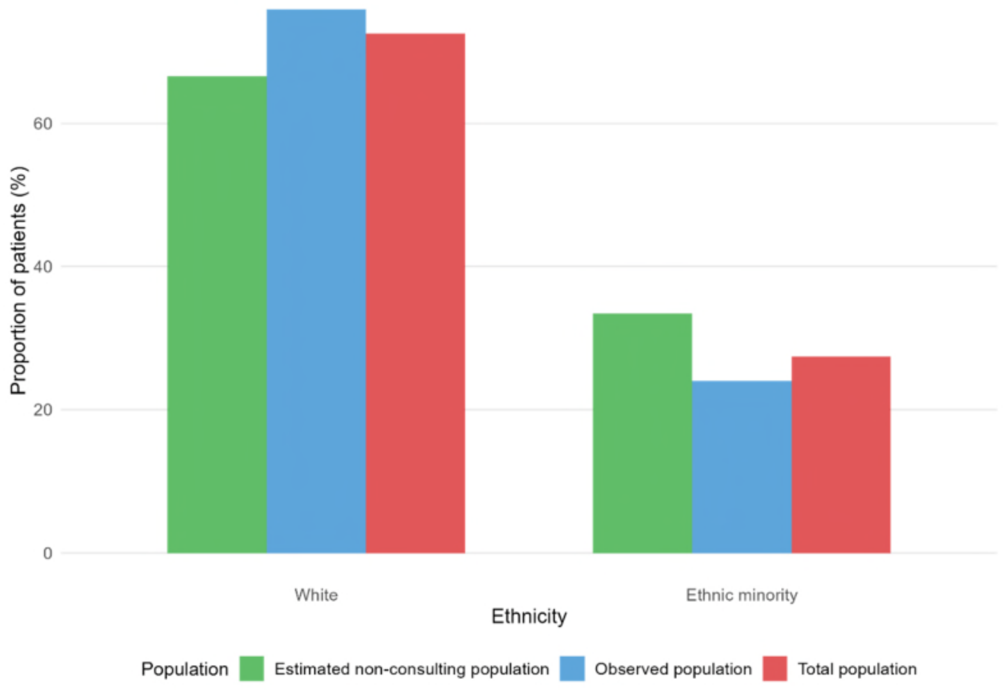

*Figure S8 Proportion of patients by IMD twentile, age group, sex, and ethnicity for adults*

Panel (a) IMD twentile

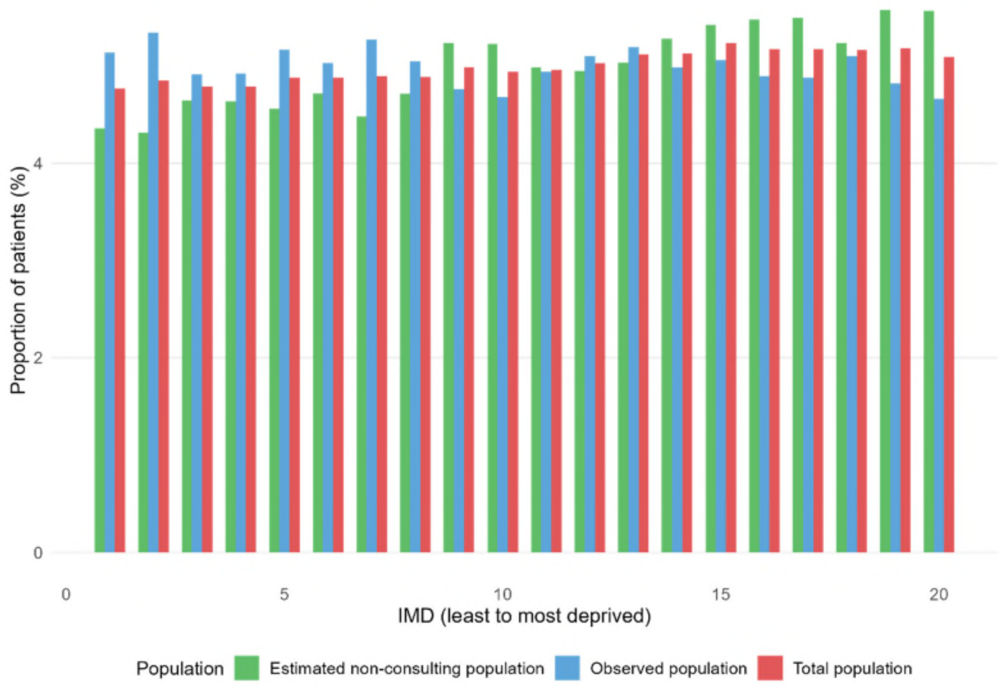


Panel (b) Age group

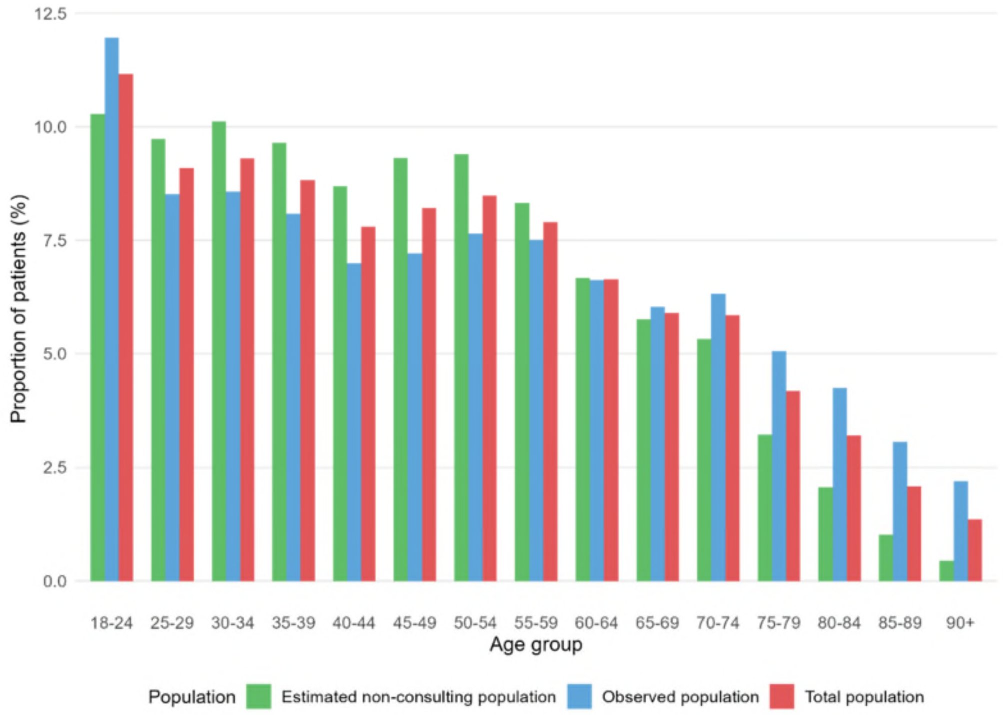

Panel (c) Sex

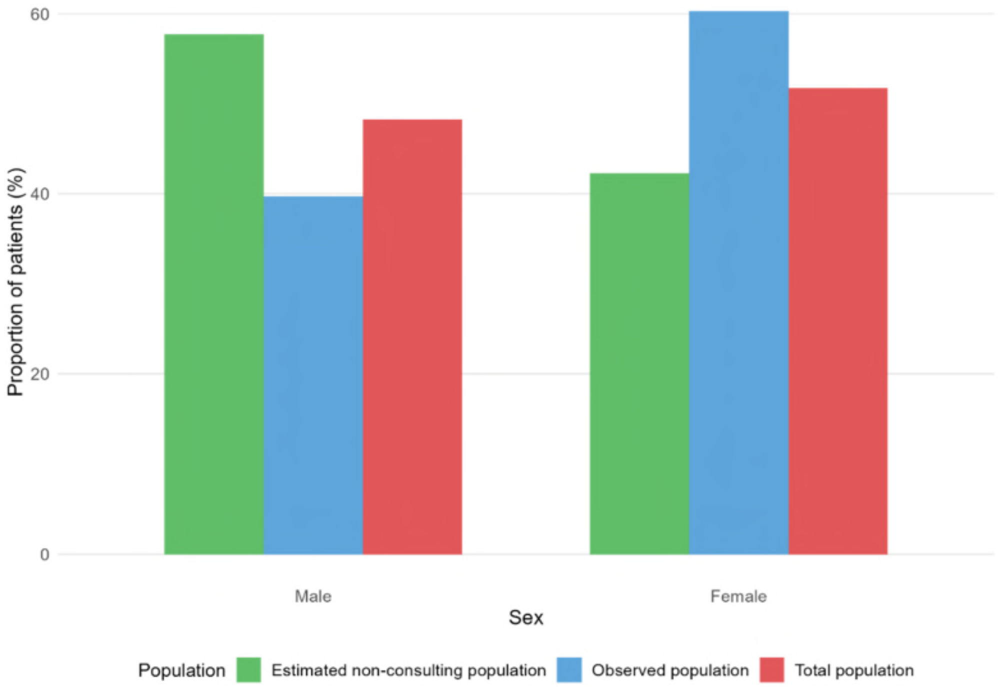


Panel (d) Ethnicity

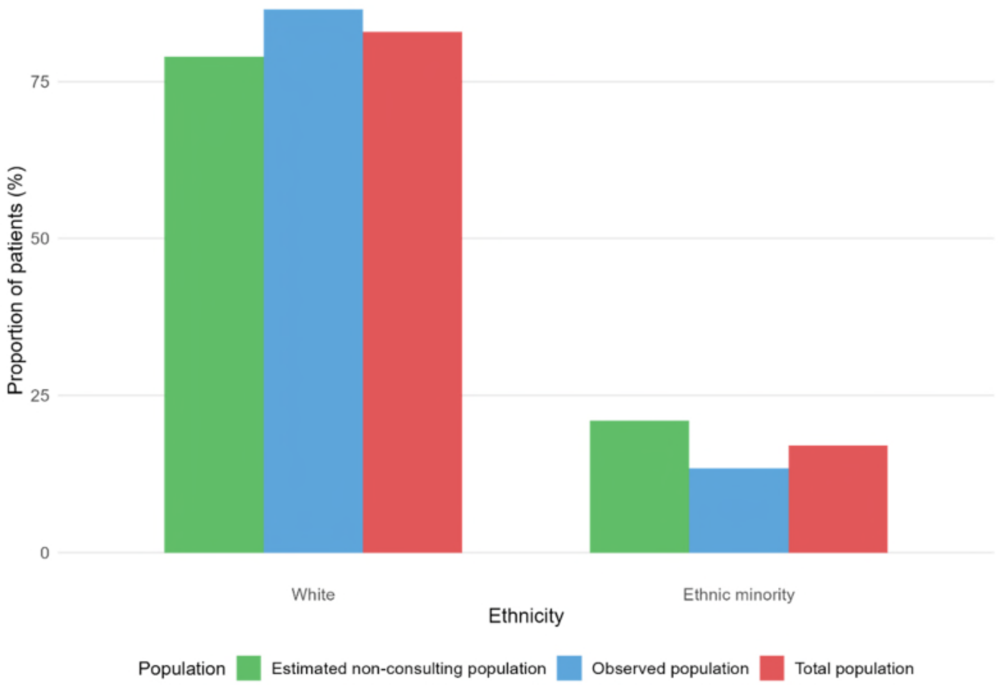

##### S3.3.2 Prescribing rates by deprivation quintile

*Figure S9 Age-standardised antibiotic prescribing rates (prescriptions per 1000 person-years) for specific infections by deprivation quintile in the observed population*

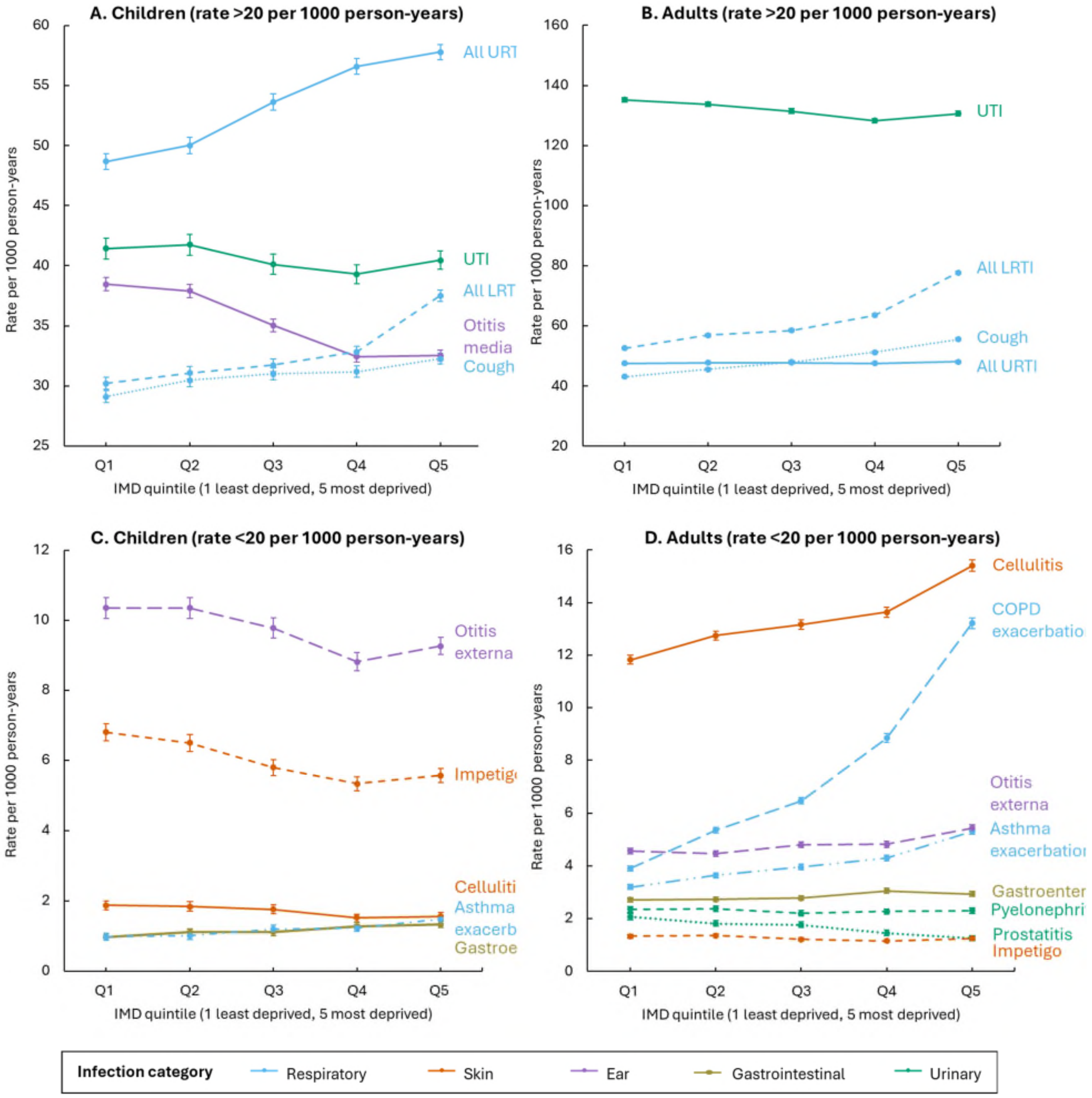


Notes: Error bars represent the 95% confidence intervals. Abbreviations: IMD index of multiple deprivation, URTI upper respiratory tract infections, LRTI lower respiratory tract infections, UTI urinary tract infection, COPD chronic obstructive pulmonary disease. "All URTI" includes sore throat, unspecified upper respiratory tract infection, rhinosinusitis whereas "All LRTI" includes bronchitis, unspecified lower respiratory tract infection, pneumonia. See fig S10 for prescribing rates of individual respiratory tract infections.

*Figure S10 Age-standardised antibiotic prescribing rates (prescriptions per 1000 person-years) for respiratory tract infections by IMD quintile in the observed population*

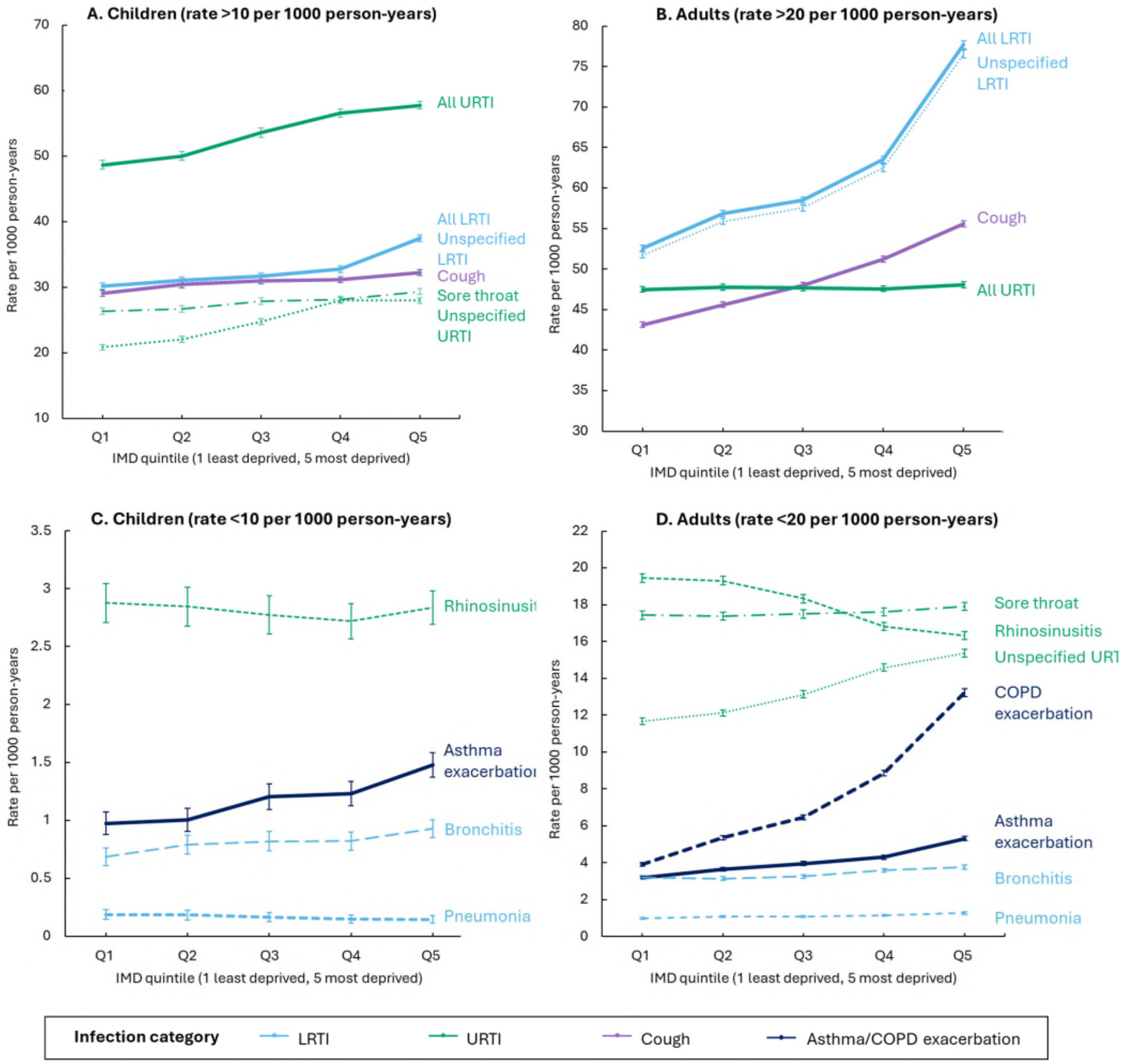


Notes: Error bars represent the 95% confidence intervals. Abbreviations: IMD index of multiple deprivation, URTI upper respiratory tract infections, LRTI lower respiratory tract infections, UTI urinary tract infection, COPD chronic obstructive pulmonary disease. "All URTI" includes sore throat, unspecified upper respiratory tract infection, rhinosinusitis whereas "All LRTI" includes bronchitis, unspecified lower respiratory tract infection, pneumonia.

##### S3.3.3 Association between deprivation and indication-related outcomes

*Figure S11 Association between IMD and antibiotic prescribing rates (prescriptions per 1000 person-years) in the observed population - mean and variance measure with shaded 95% confidence intervals reported*

Panel (a) Children

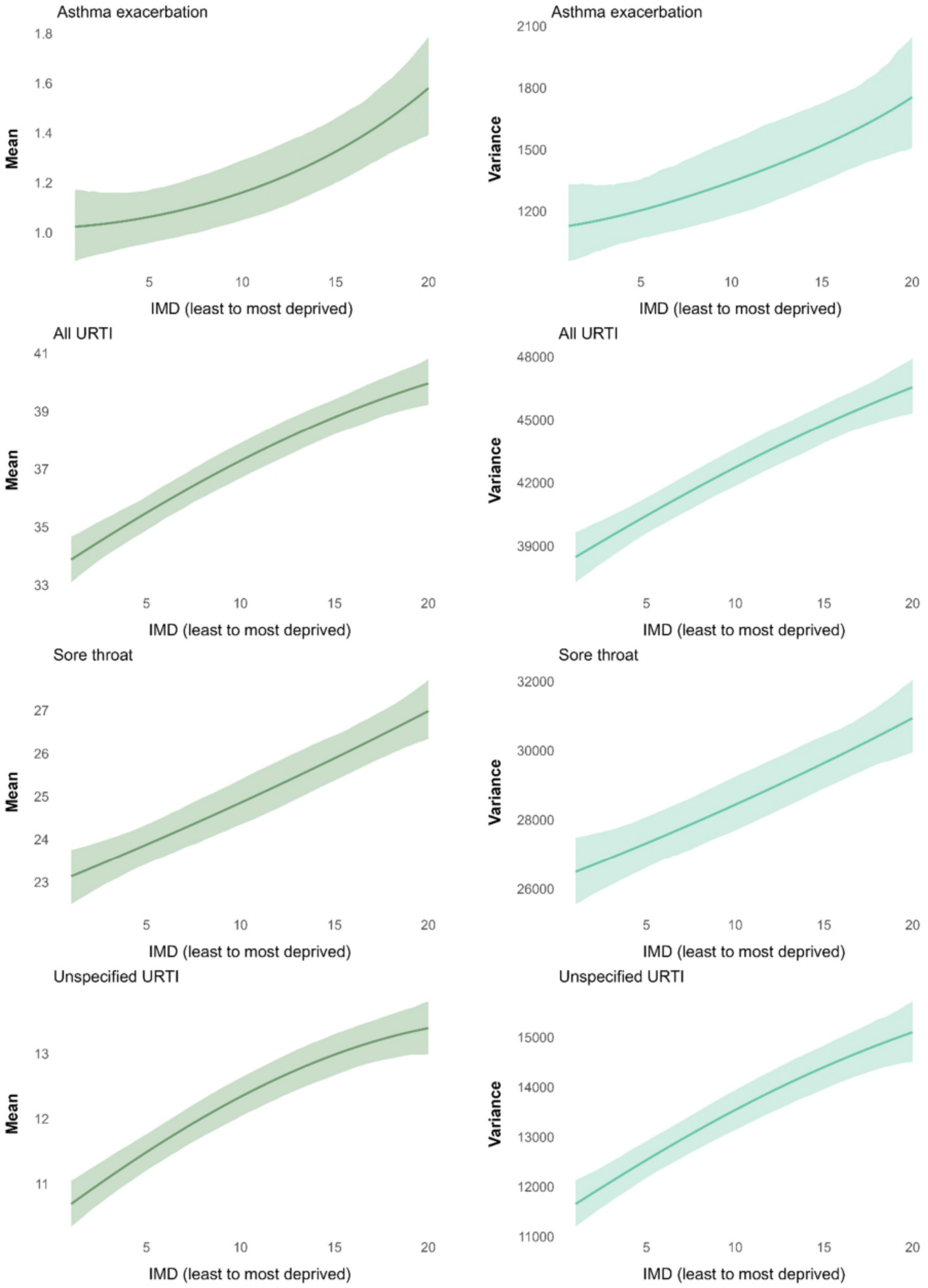

*Figure S11 (a). Prescribing rate (per 1000 person-years) in children - mean and variance (continued)*

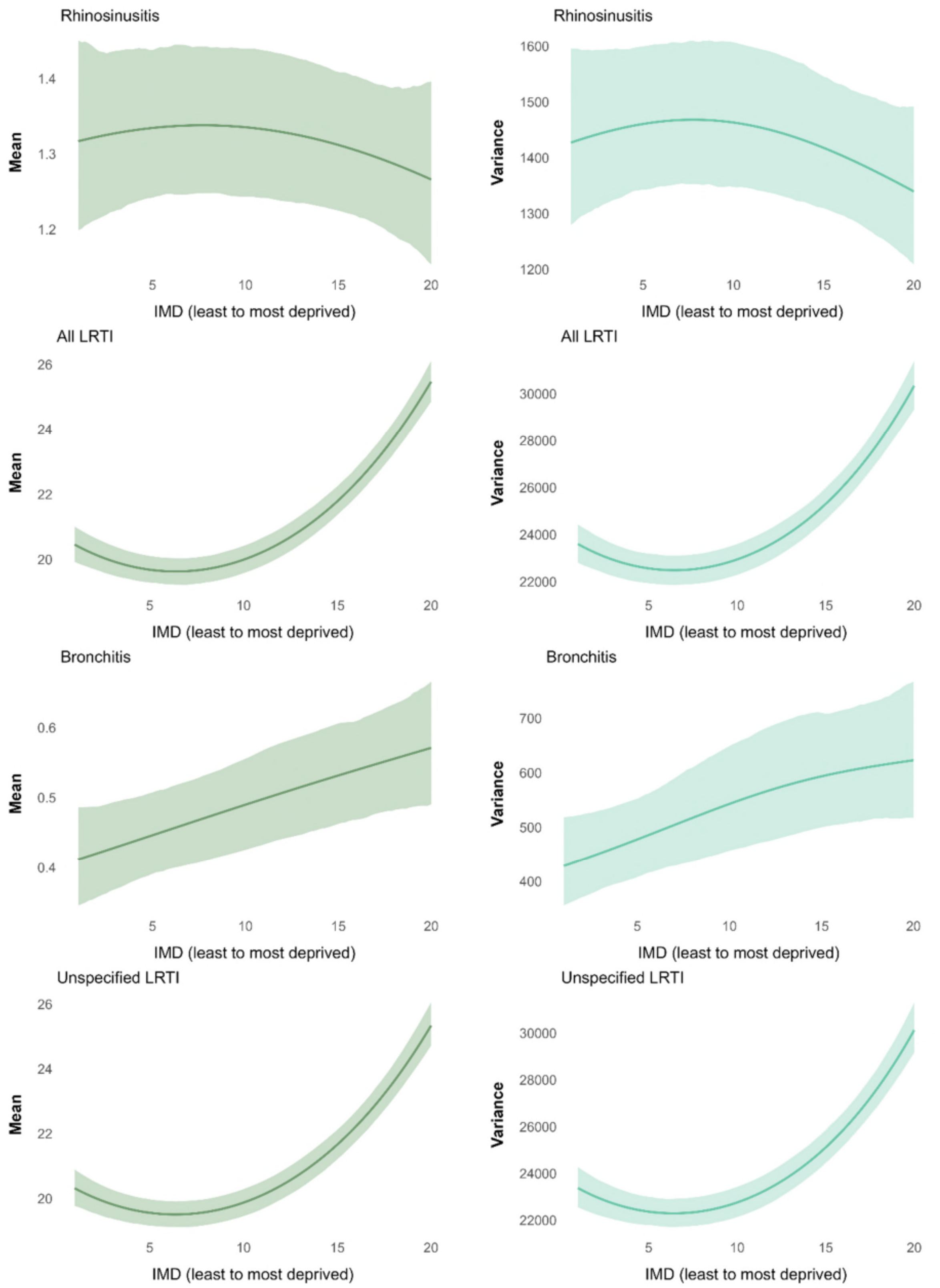

*Figure S11 (a). Prescribing rate (per 1000 person-years) in children - mean and variance (continued)*

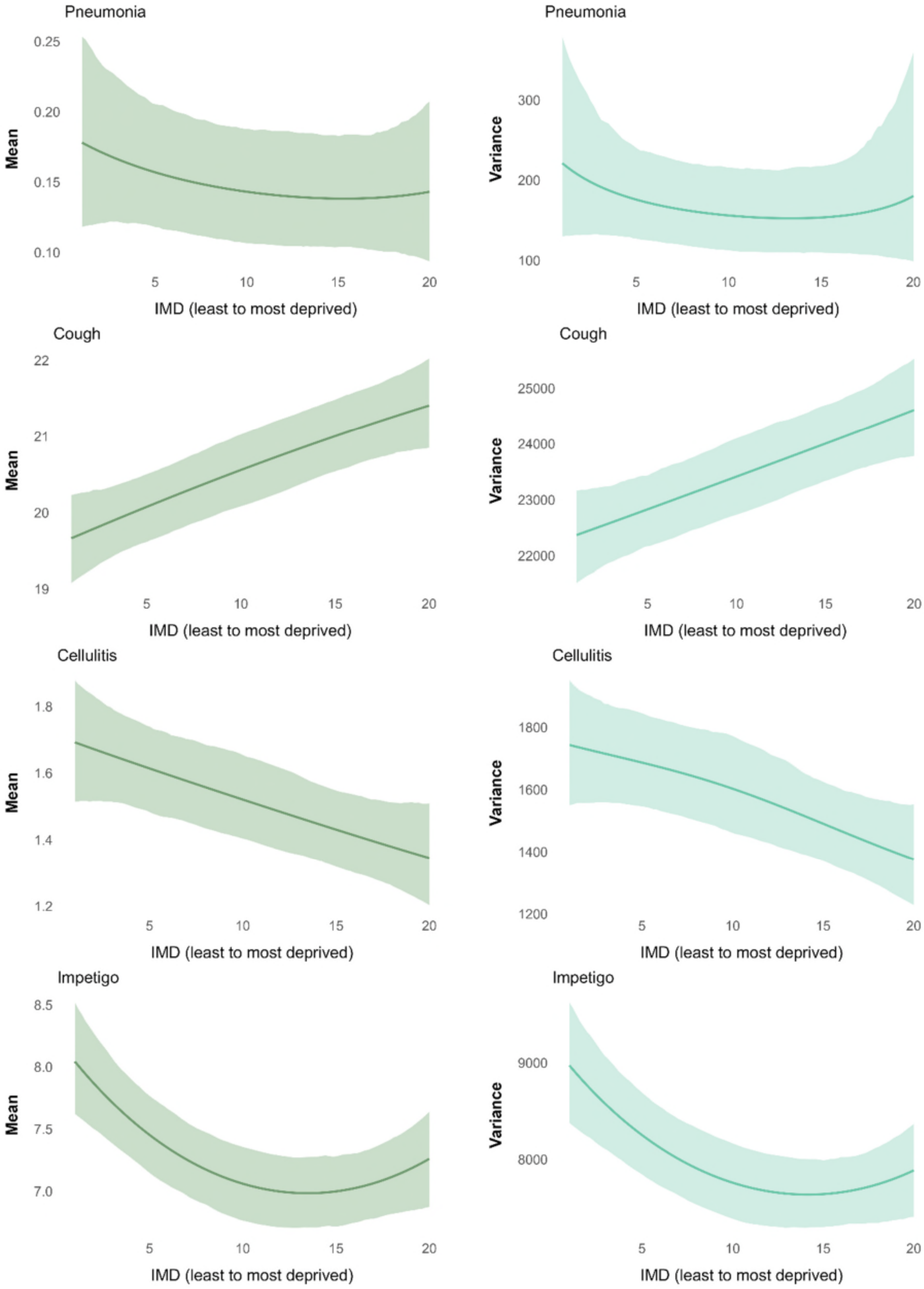

*Figure S11 (a). Prescribing rate (per 1000 person-years) in children - mean and variance (continued)*

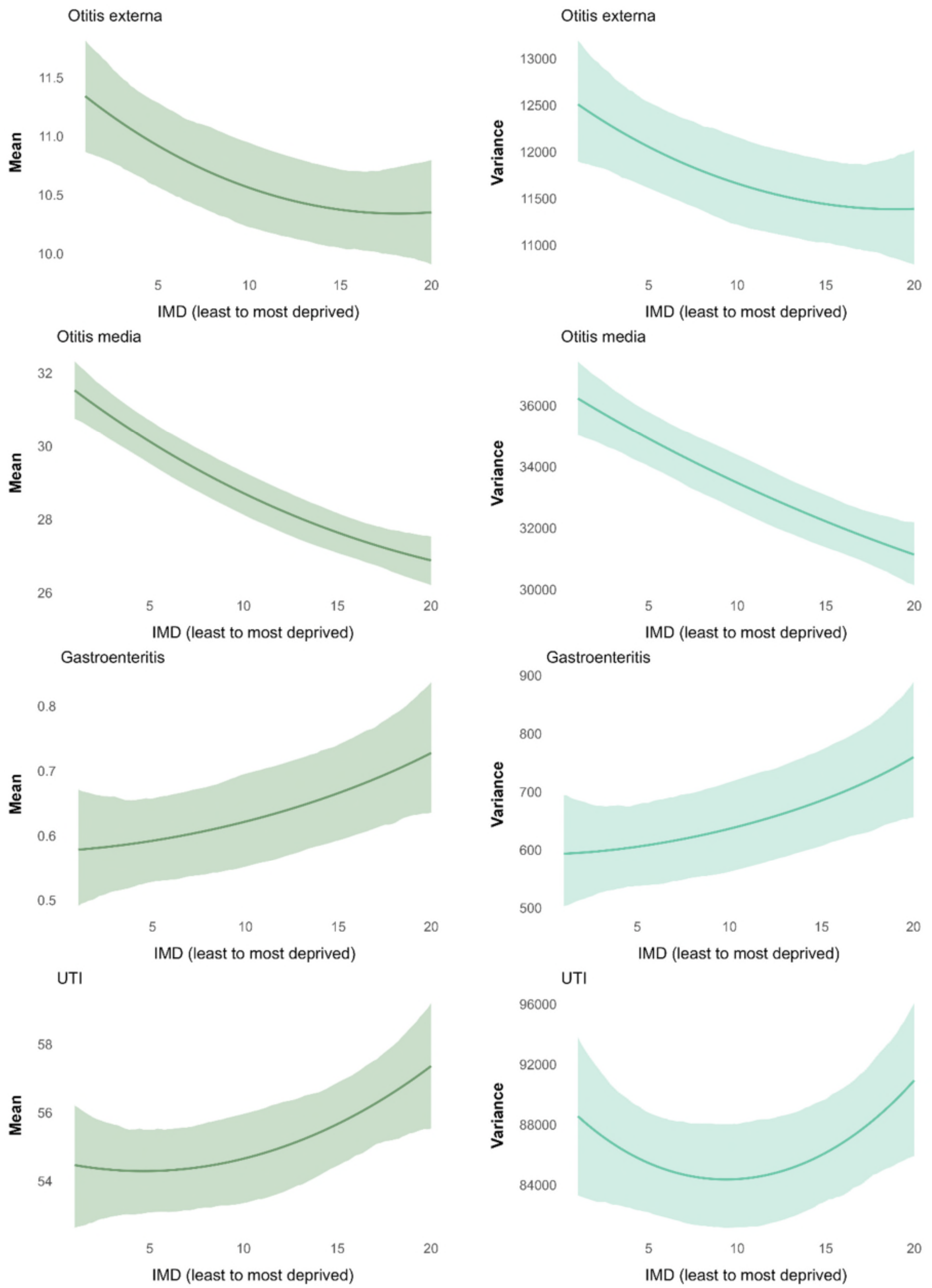


Notes: All models were adjusted for age, sex, and ethnicity. Abbreviations: IMD index of multiple deprivation, COPD chronic obstructive pulmonary disease, LRTI lower respiratory tract infections, URTI upper respiratory tract infections, UTI urinary tract infections.

Panel (b) Adults

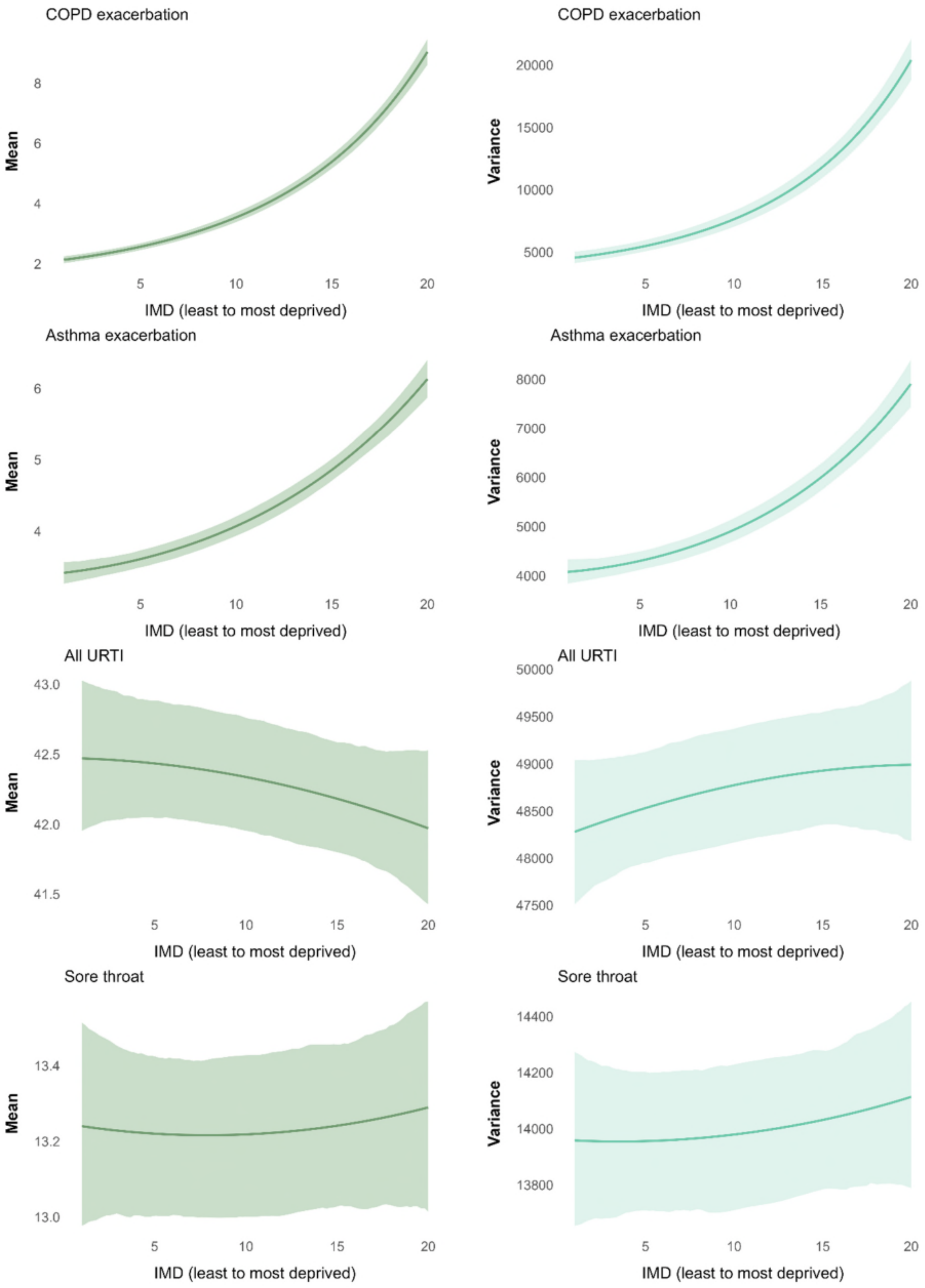

*Figure S11 (b). Prescribing rate (per 1000 person-years) in adults - mean and variance (continued)*

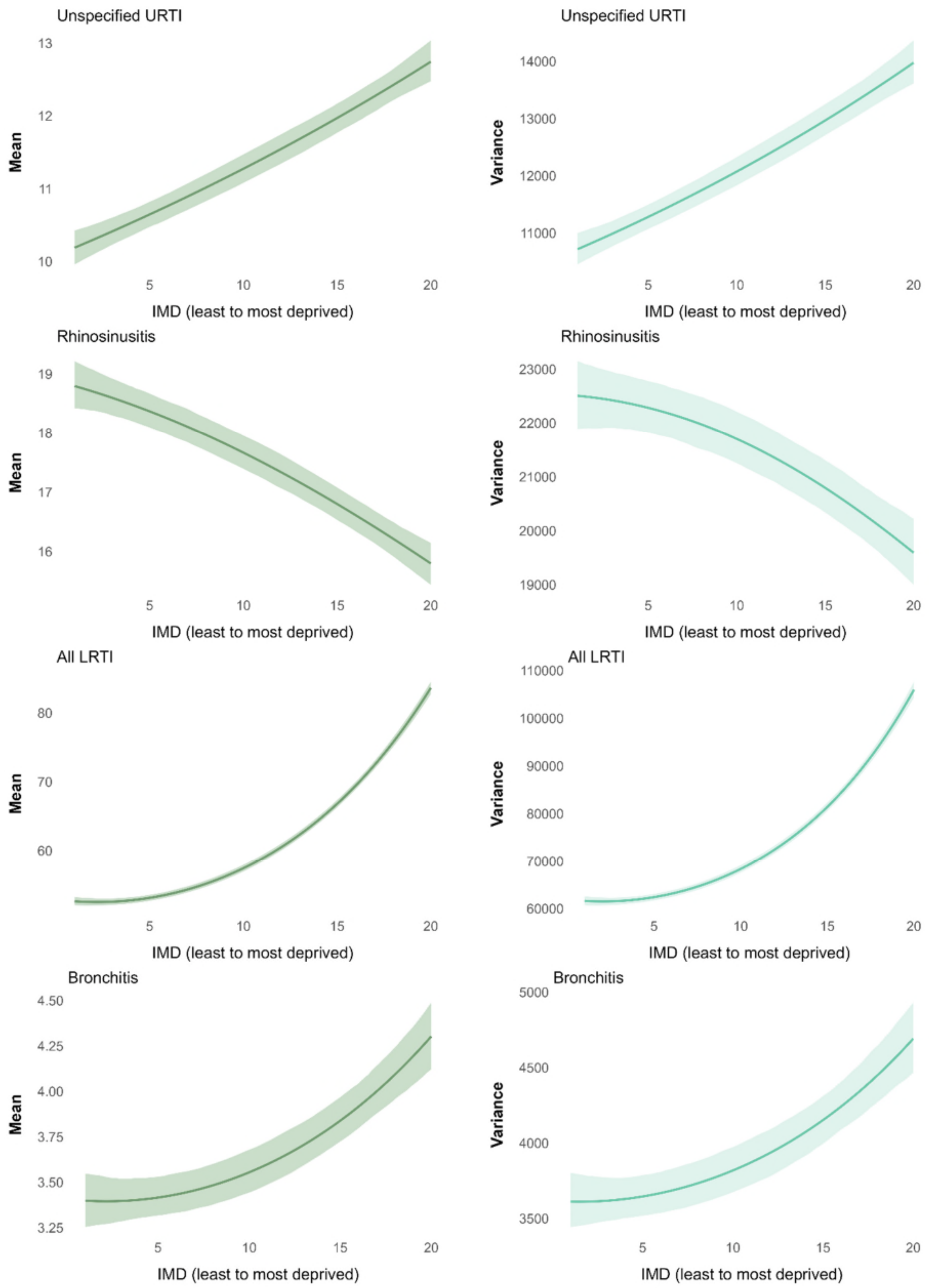

*Figure S11 (b). Prescribing rate (per 1000 person-years) in adults - mean and variance (continued)*

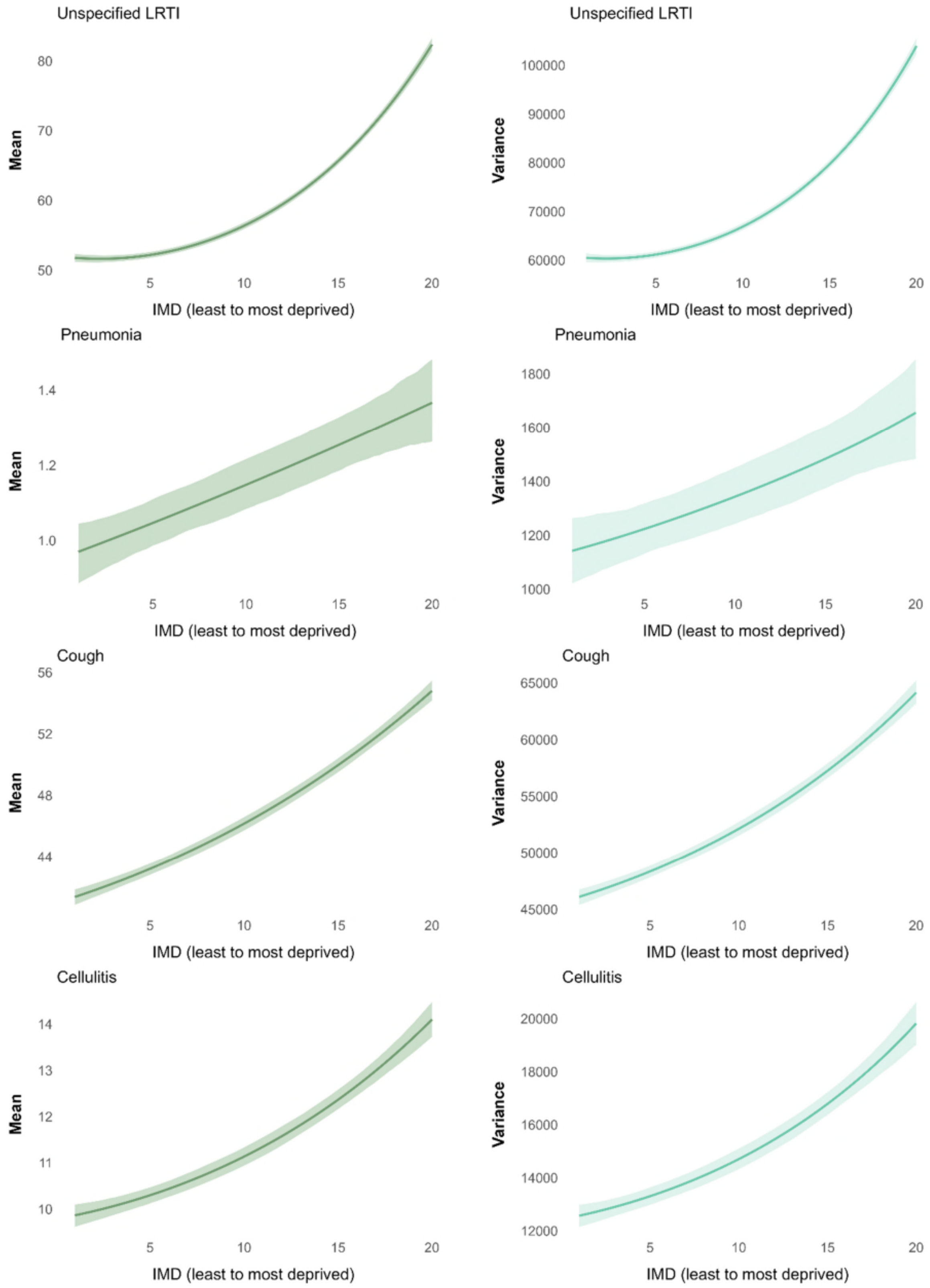

*Figure S11 (b). Prescribing rate (per 1000 person-years) in adults - mean and variance (continued)*

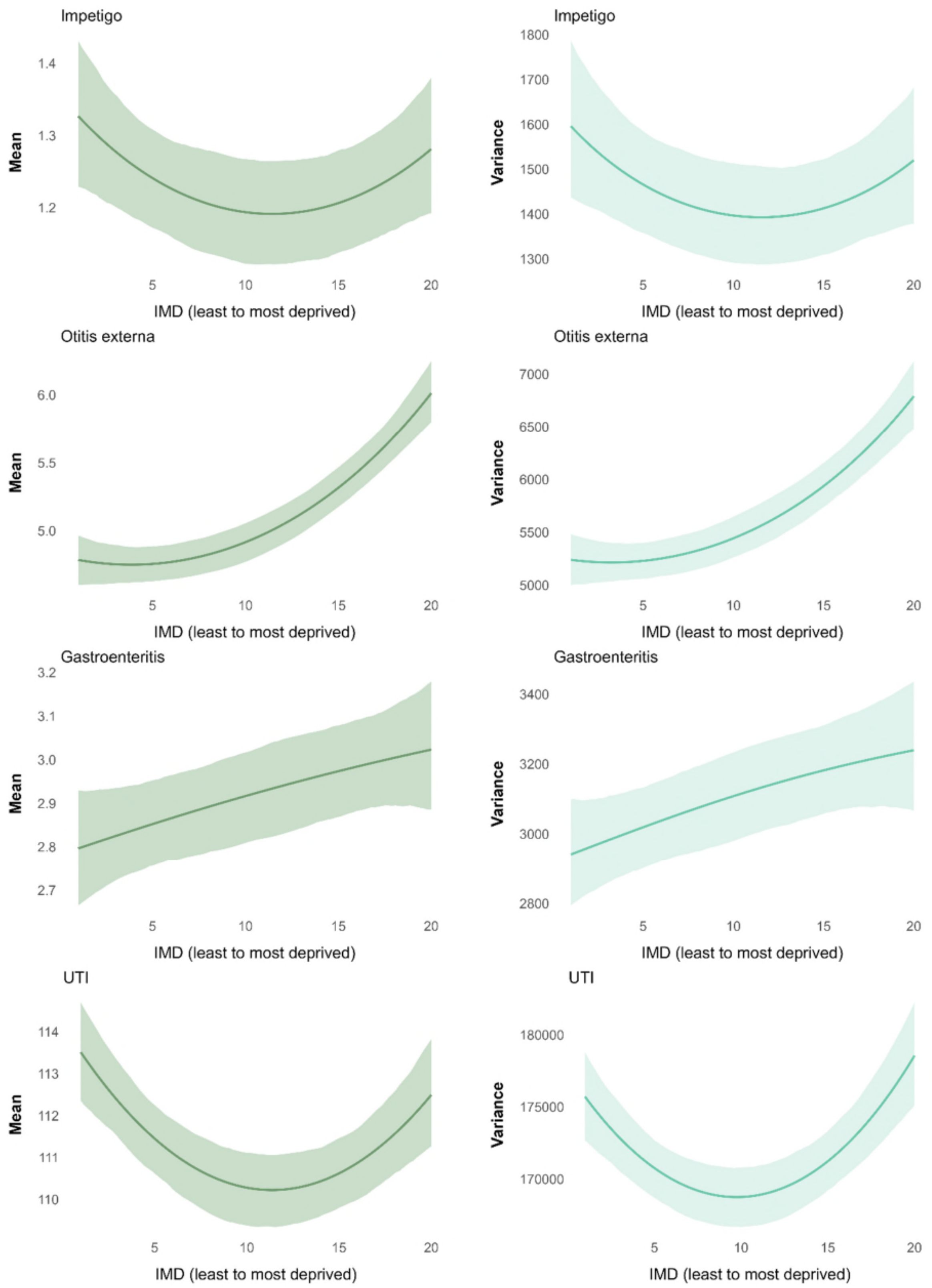

*Figure S11 (b). Prescribing rate (per 1000 person-years) in adults - mean and variance (continued)*

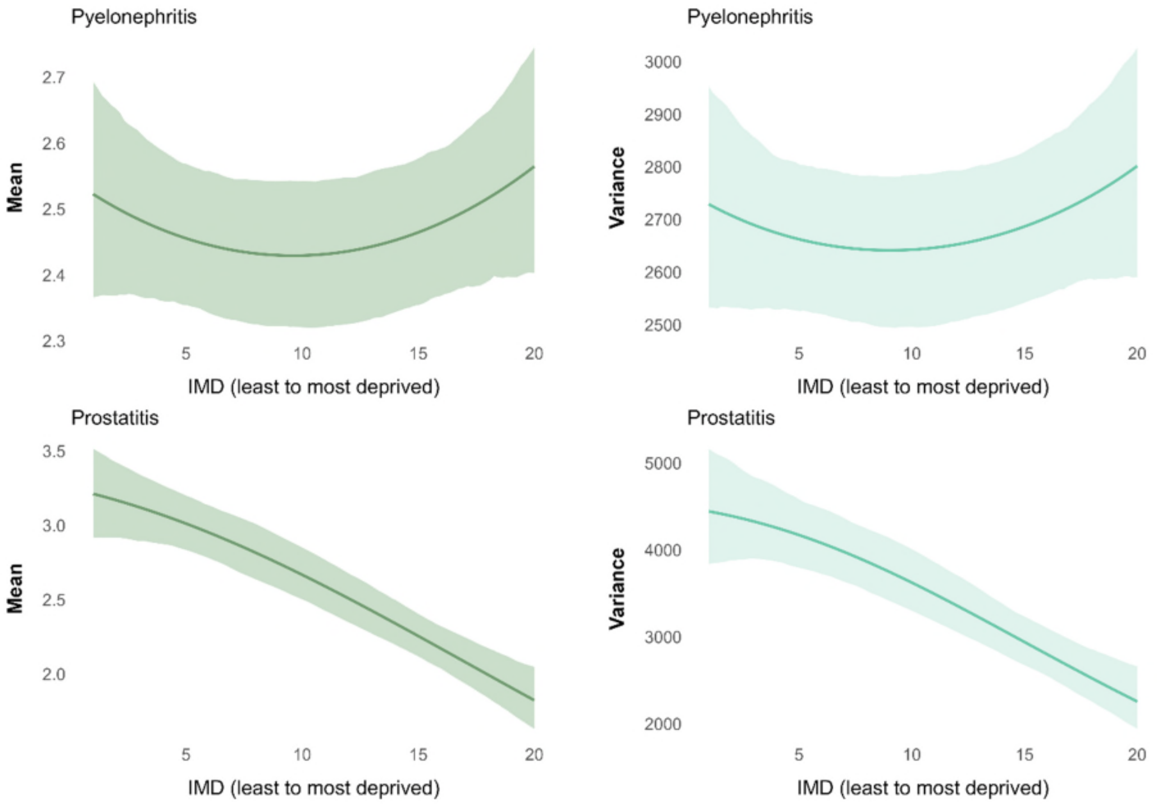


Notes: All models were adjusted for age, sex, and ethnicity. Abbreviations: IMD index of multiple deprivation, COPD chronic obstructive pulmonary disease, LRTI lower respiratory tract infections, URTI upper respiratory tract infections, UTI urinary tract infections. “All URTI” includes sore throat, unspecified upper respiratory tract infection, rhinosinusitis whereas “All LRTI” includes bronchitis, unspecified lower respiratory tract infection, pneumonia.

*Figure S12 Association between IMD and consultation rates (consultations per 1000 person-years) in the observed population - mean and variance measure with shaded 95% confidence intervals reported*

Panel (a) Children

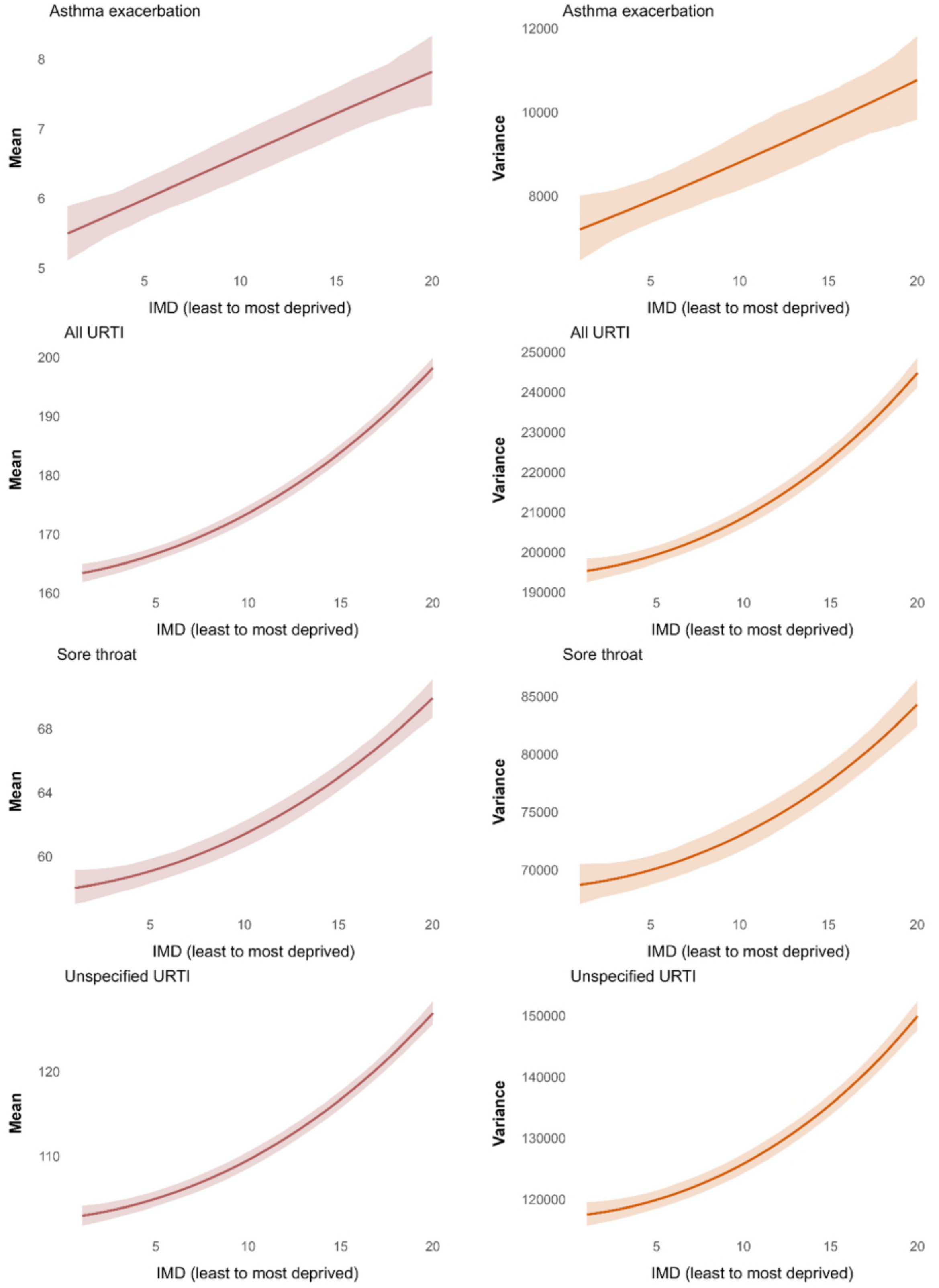

*Figure S12 (a). Consultation rate (per 1000 person-years) in children - mean and variance (continued)*

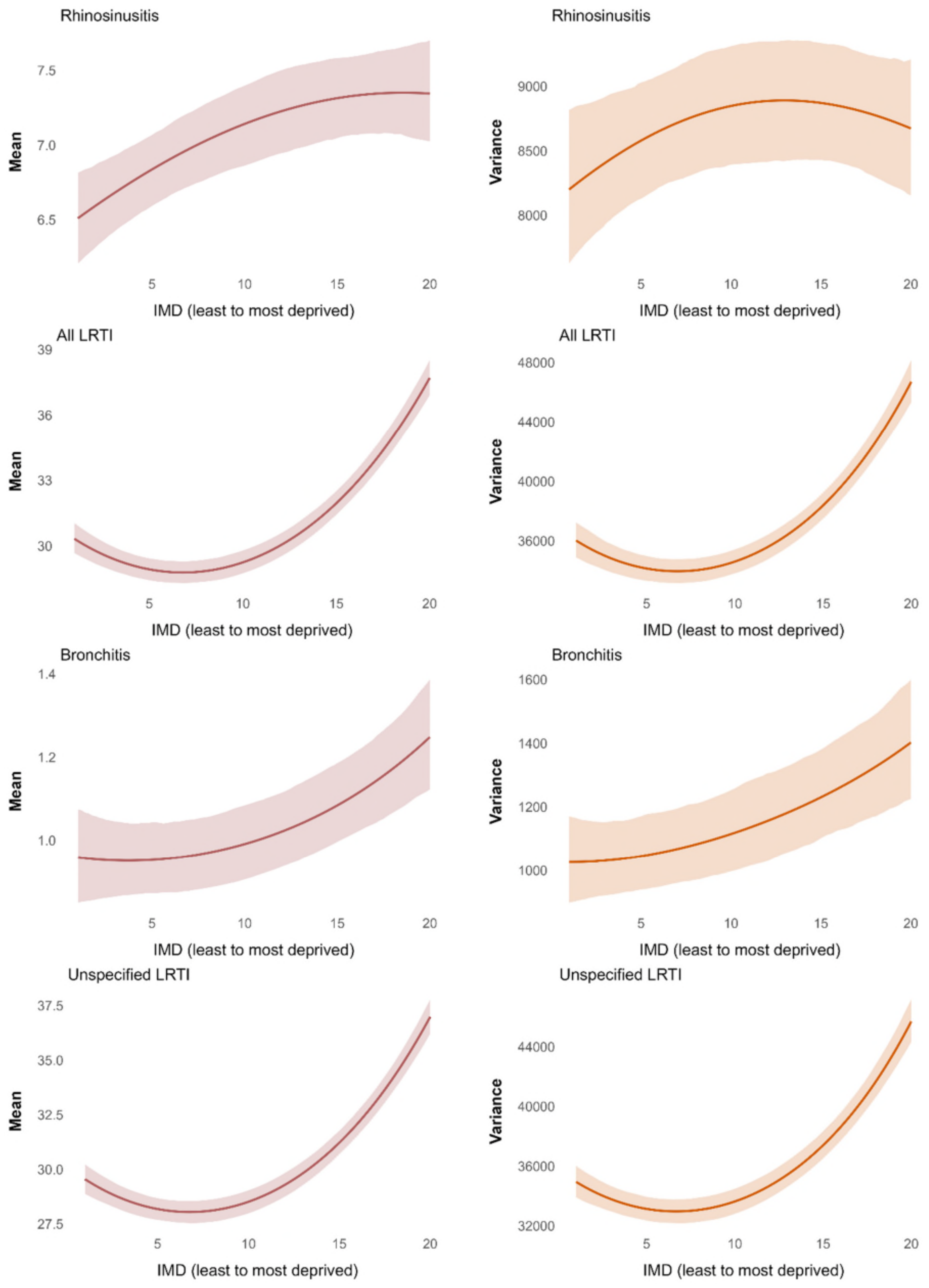

*Figure S12 (a). Consultation rate (per 1000 person-years) in children - mean and variance (continued)*

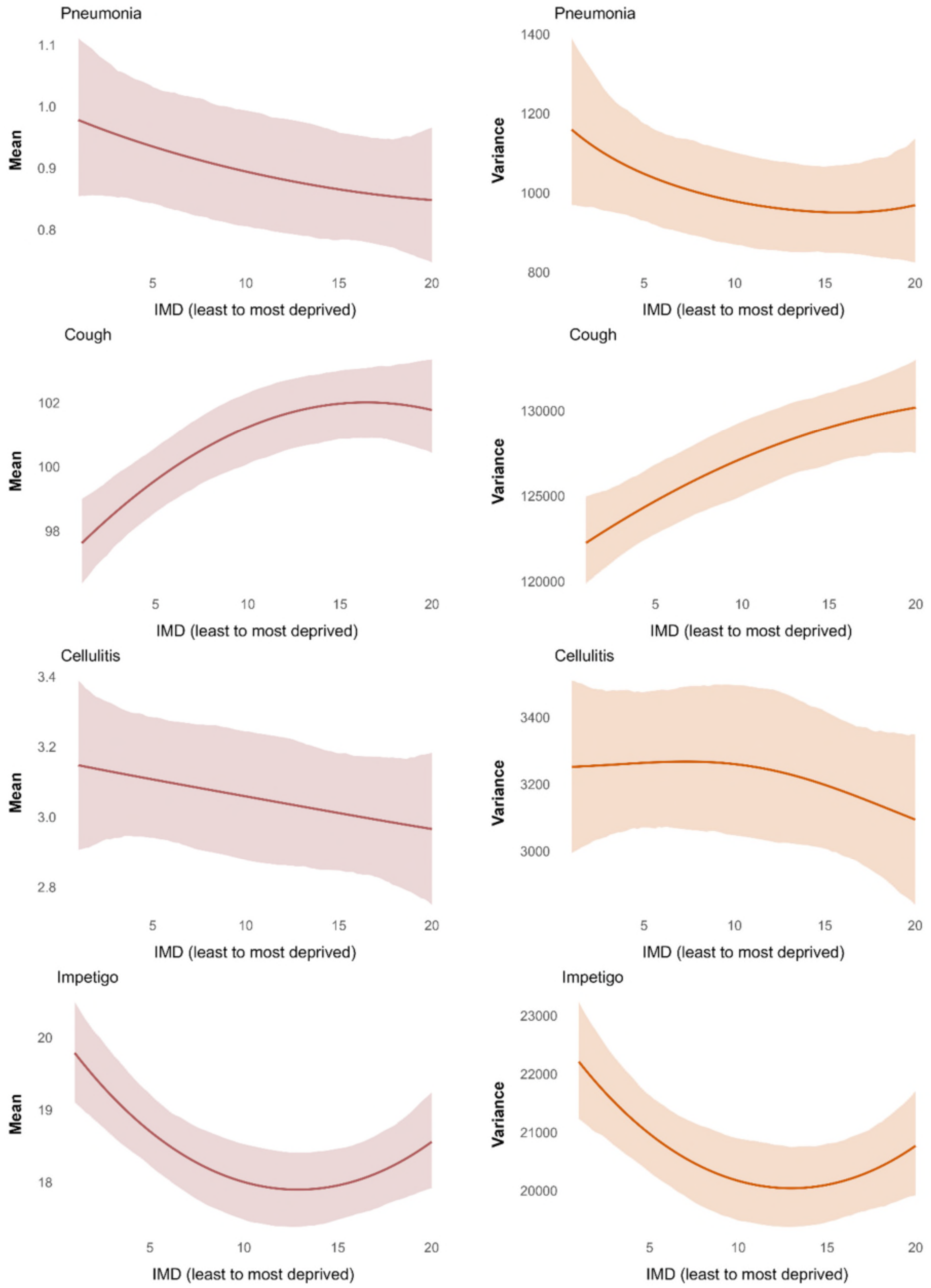

*Figure S12 (a). Consultation rate (per 1000 person-years) in children - mean and variance (continued)*

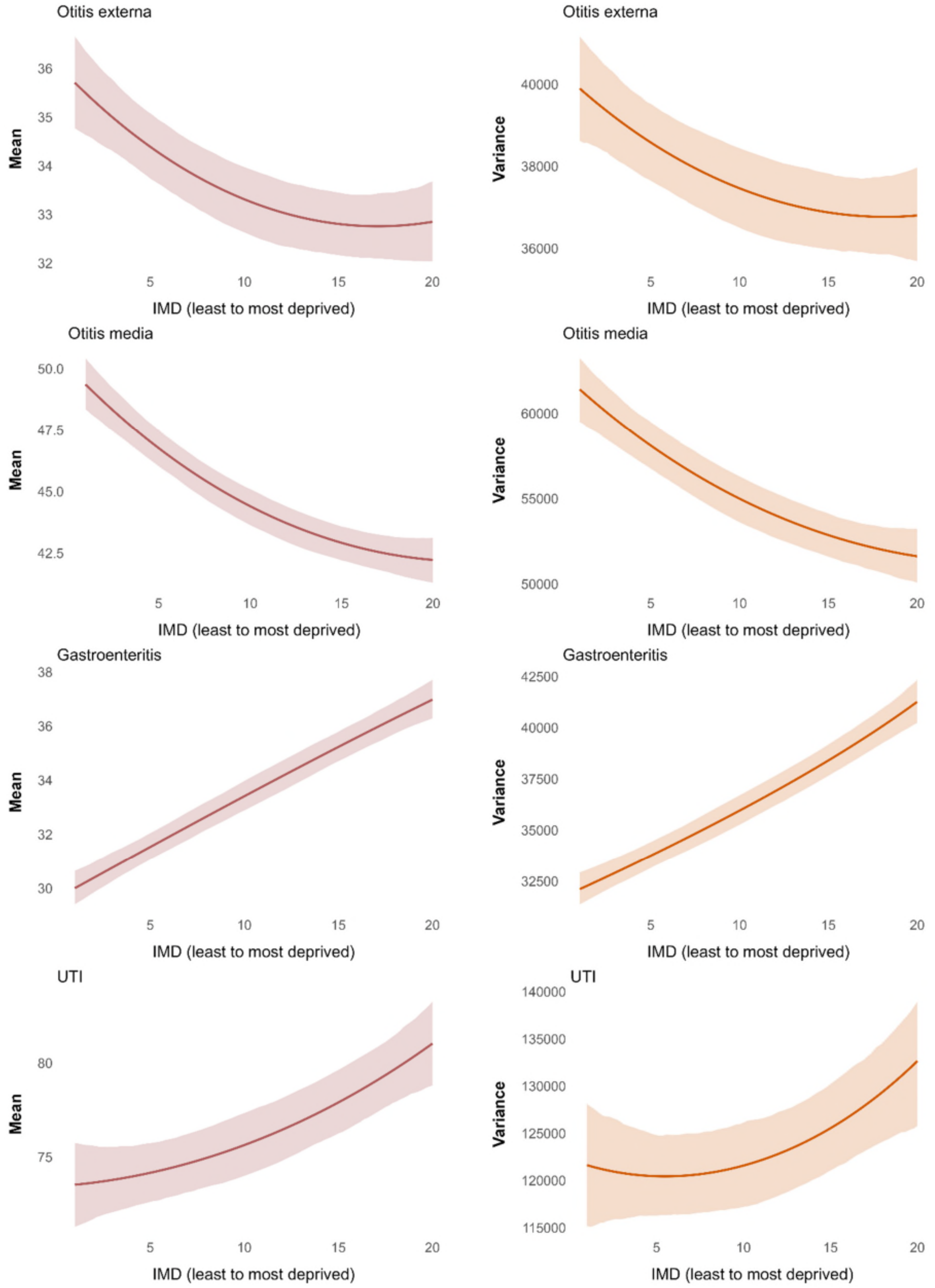

Panel (b) Adults

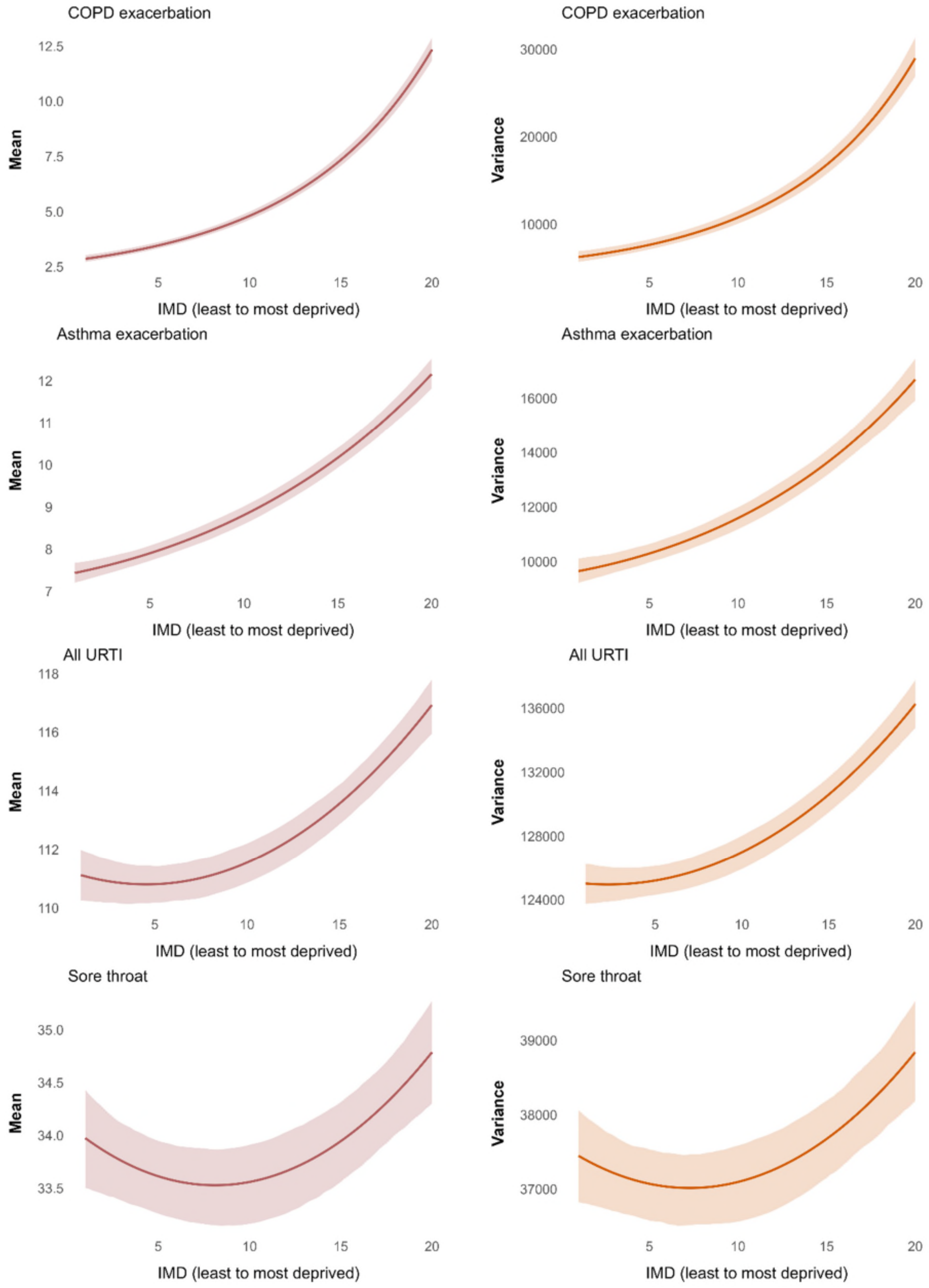

*Figure S12 (b). Consultation rate (per 1000 person-years) in adults - mean and variance (continued)*

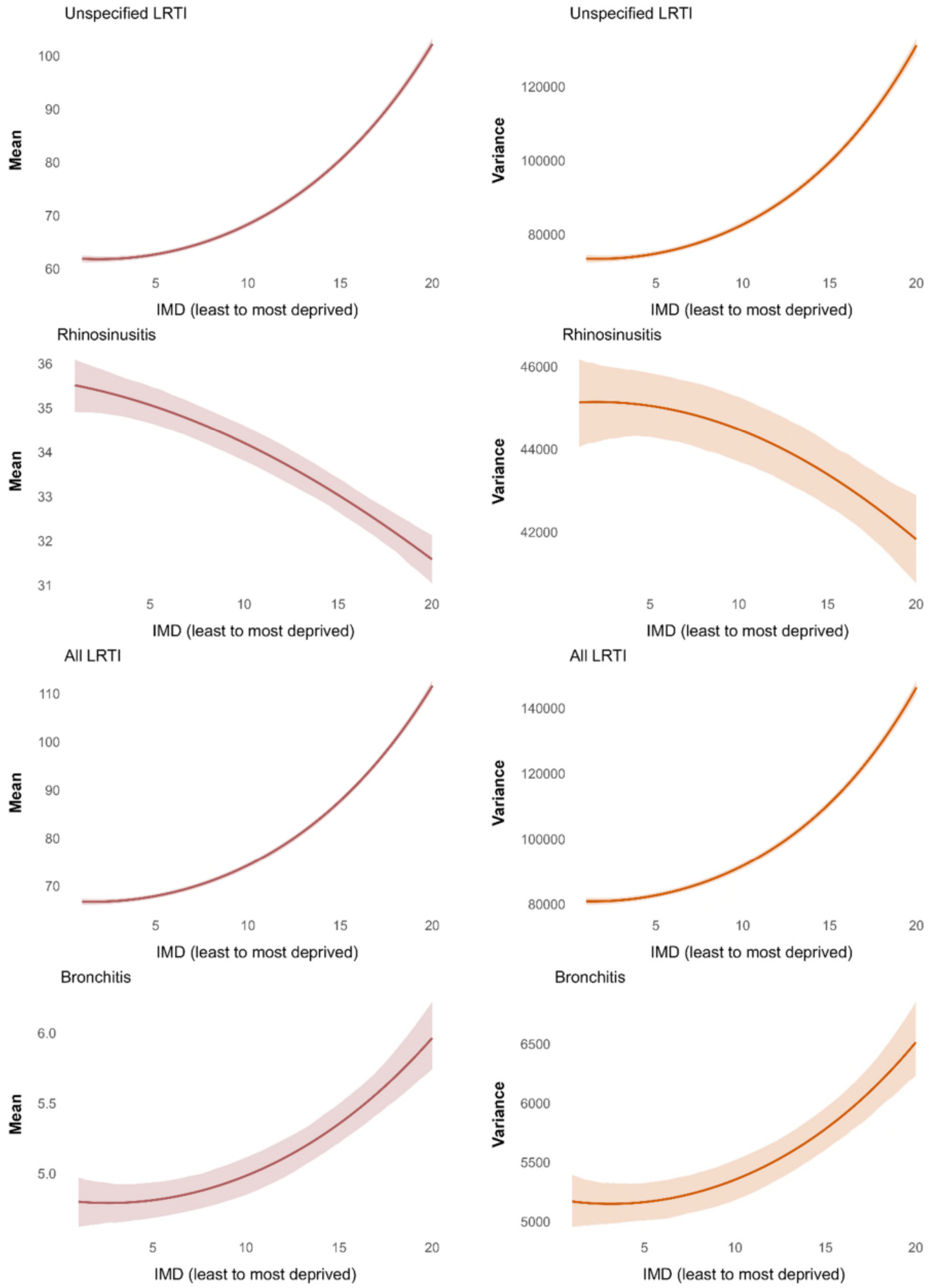

*Figure S12 (b). Consultation rate (per 1000 person-years) in adults - mean and variance (continued)*

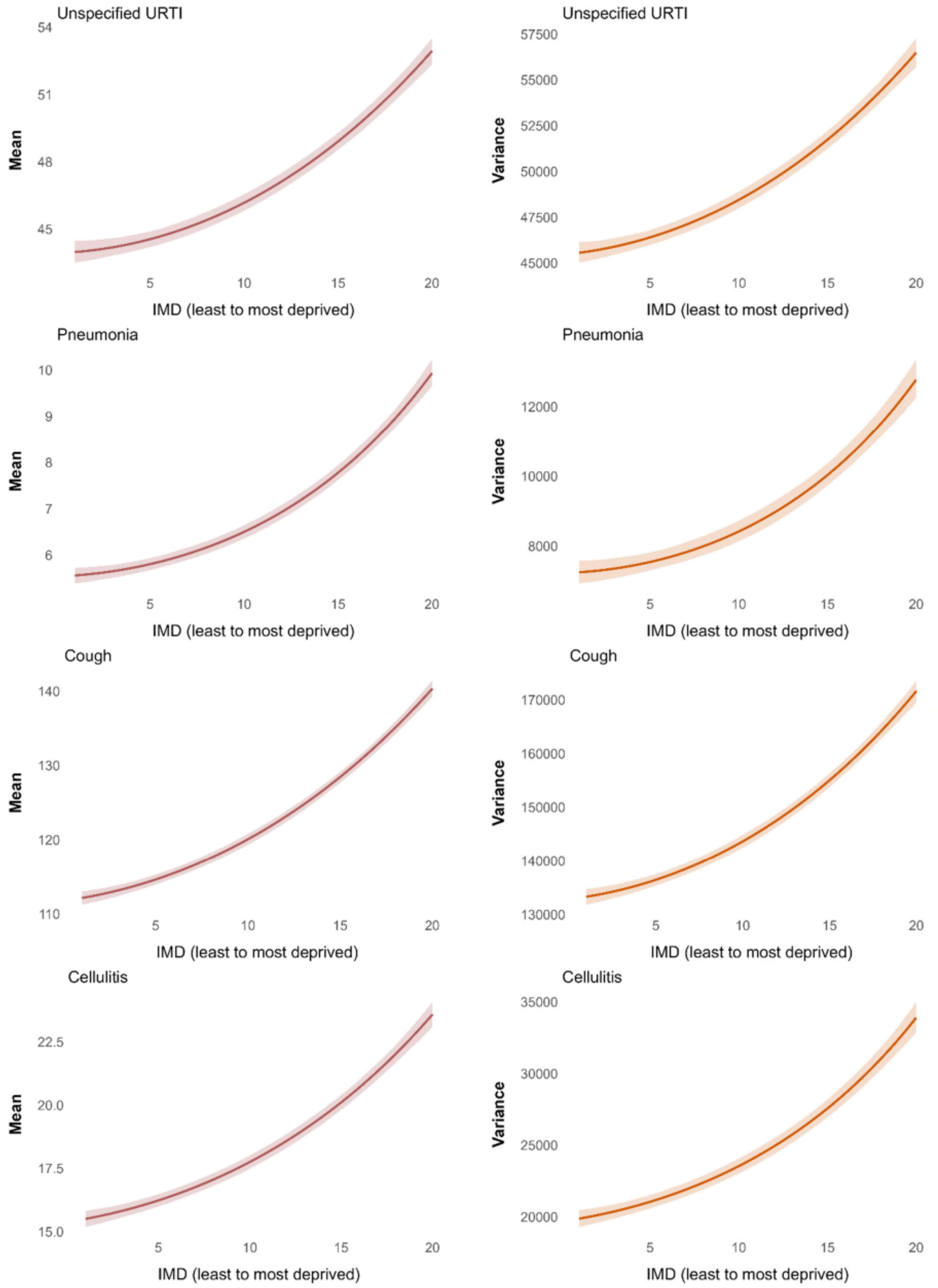

*Figure S12 (b). Consultation rate (per 1000 person-years) in adults - mean and variance (continued)*

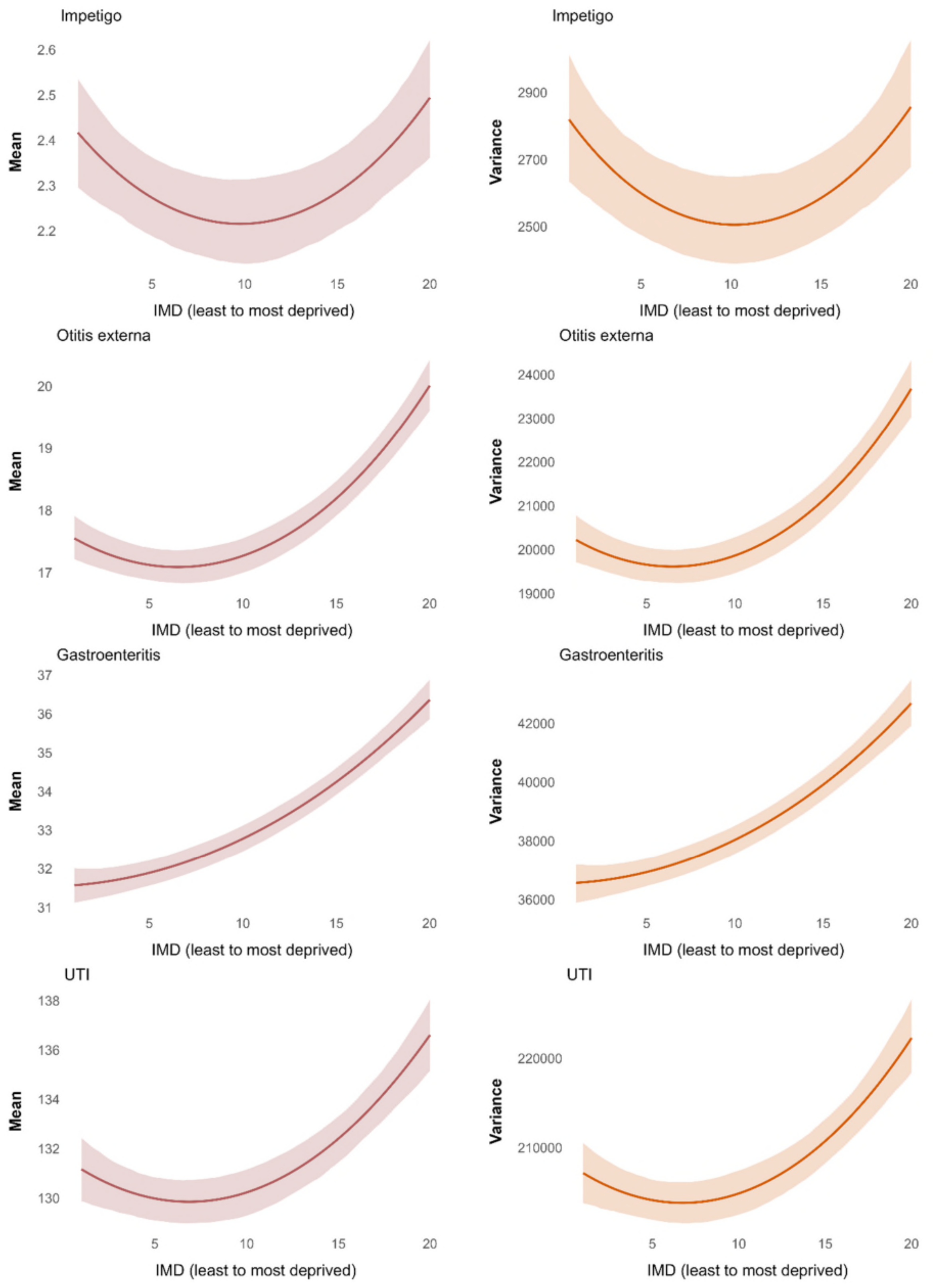

*Figure S12 (b). Consultation rate (per 1000 person-years) in adults - mean and variance (continued)*

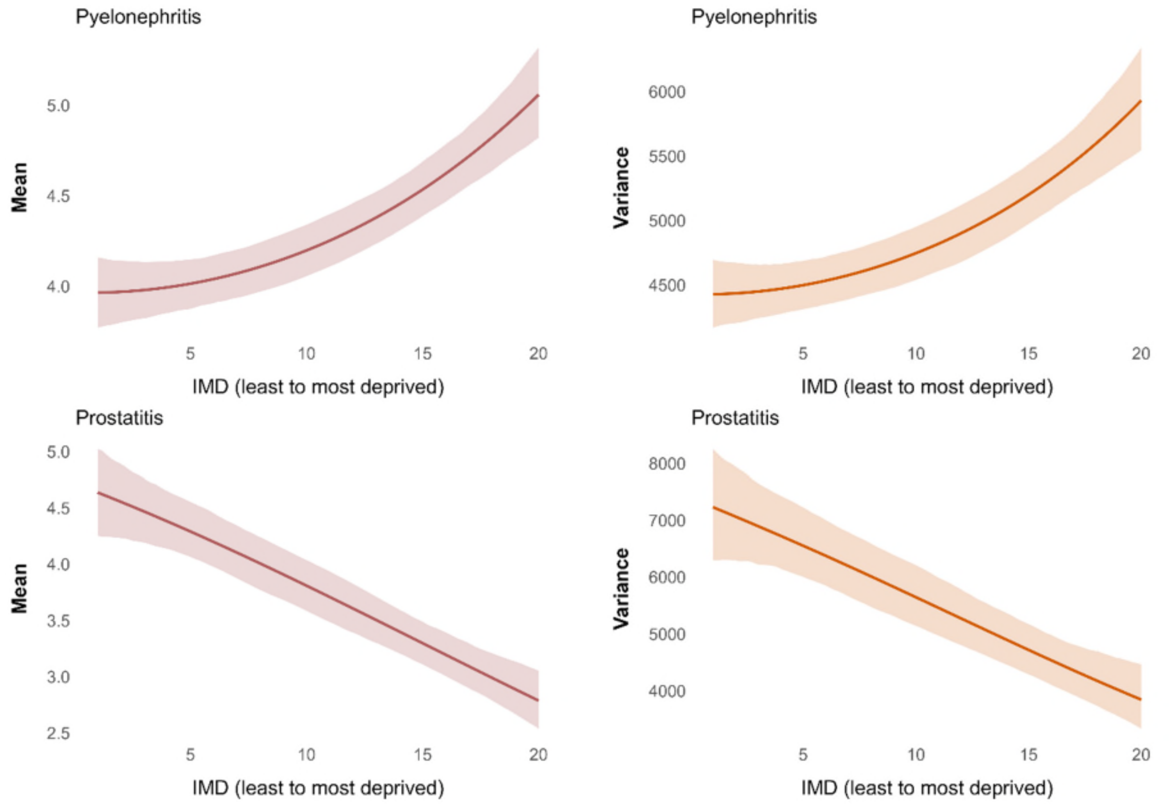


Notes: All models were adjusted for age, sex, and ethnicity. Abbreviations: IMD index of multiple deprivation, COPD chronic obstructive pulmonary disease, LRTI lower respiratory tract infections, URTI upper respiratory tract infections, UTI urinary tract infections. “All URTI” includes sore throat, unspecified upper respiratory tract infection, rhinosinusitis whereas “All LRTI” includes bronchitis, unspecified lower respiratory tract infection, pneumonia.

*Figure S13 Association between IMD and prescribing rates, consultation rates, and probability of receiving antibiotics once consulted in the observed population (mean measure with shaded 95% confidence intervals reported).*

Panel (a) Children

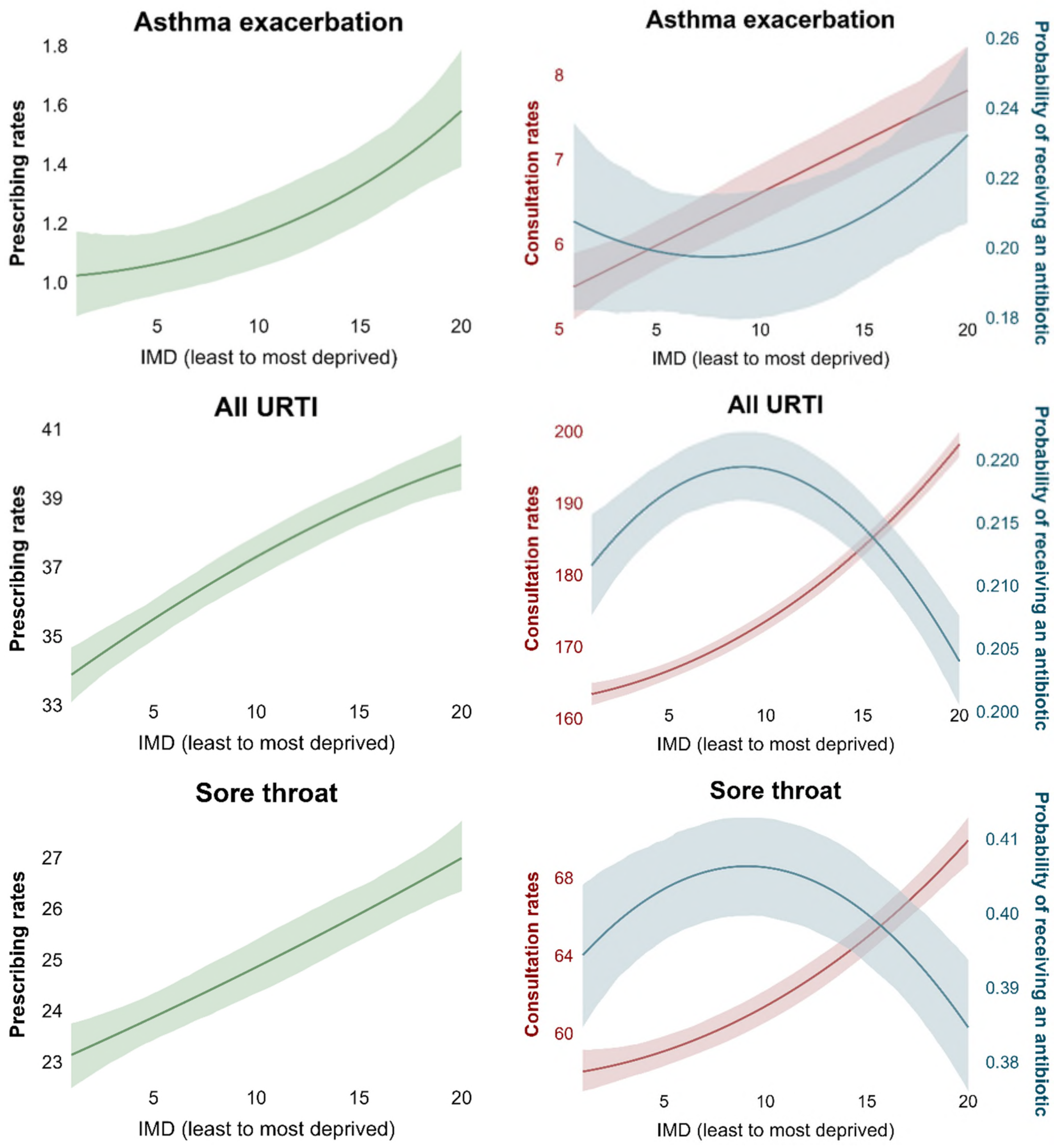

*Figure S13 (a). Mean prescribing rates, consultation rates, and probability of receiving antibiotics once consulted in children (continued)*

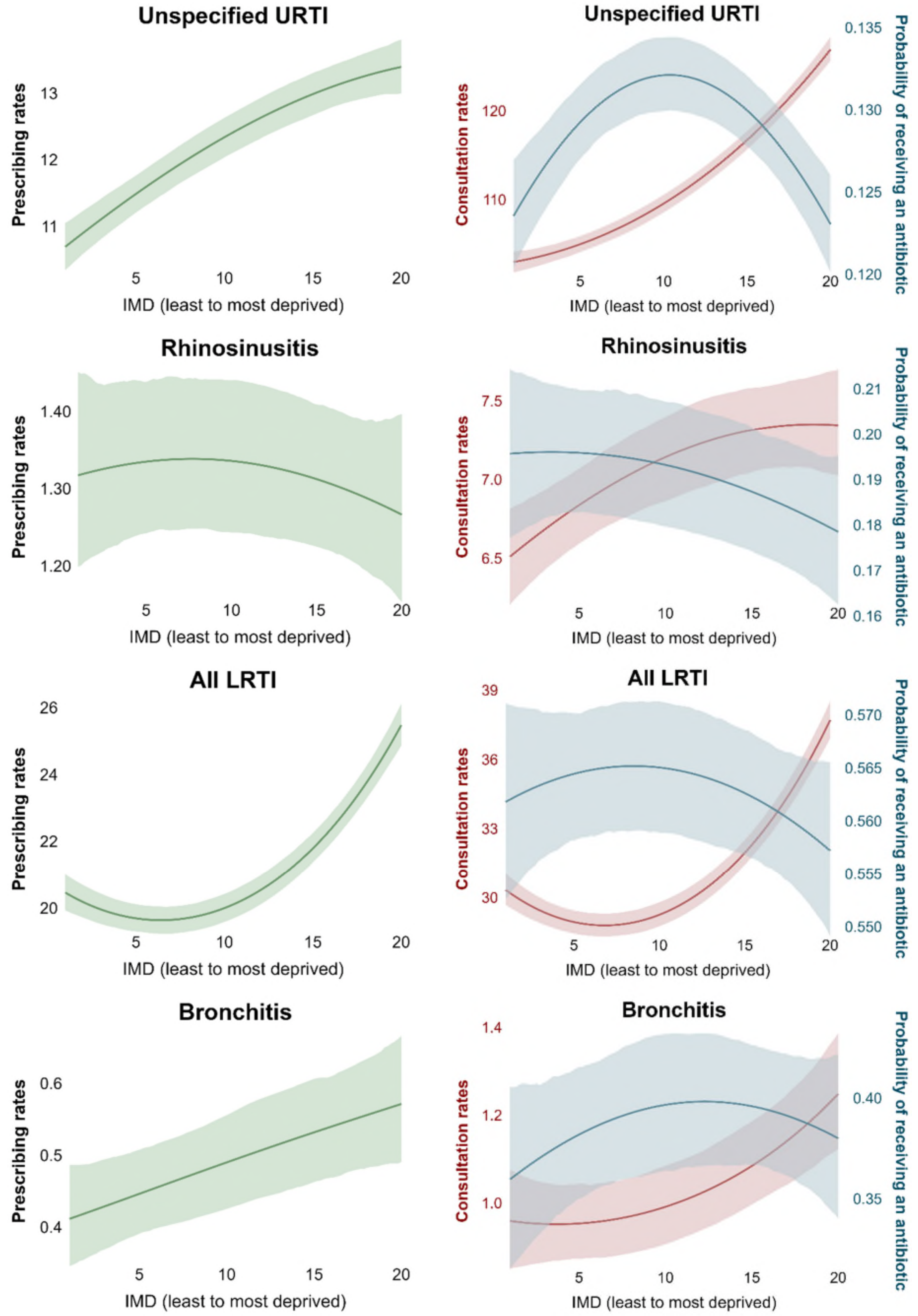

*Figure S13 (a). Mean prescribing rates, consultation rates, and probability of receiving antibiotics once consulted in children (continued)*

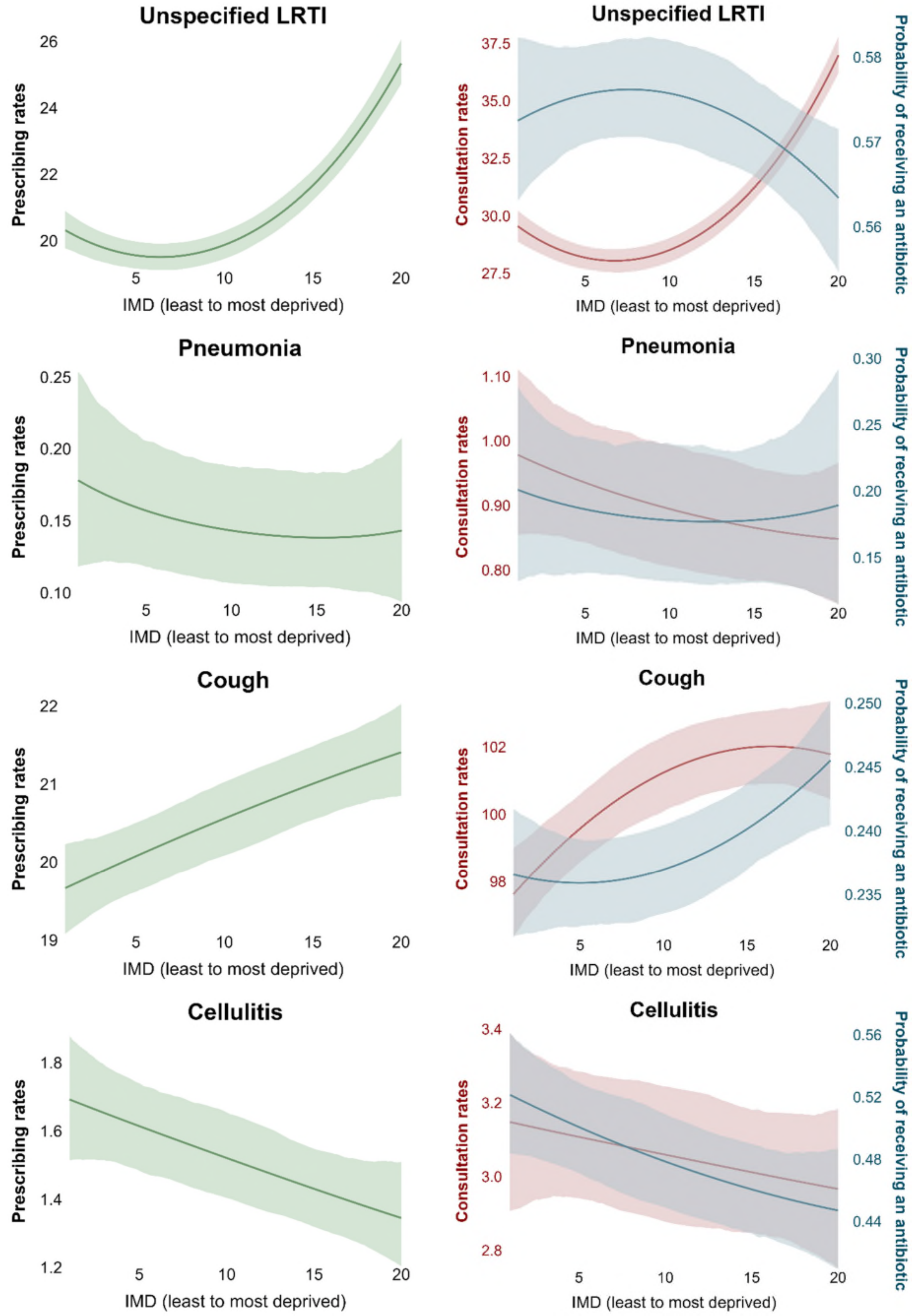

*Figure S13 (a). Mean prescribing rates, consultation rates, and probability of receiving antibiotics once consulted in children (continued)*

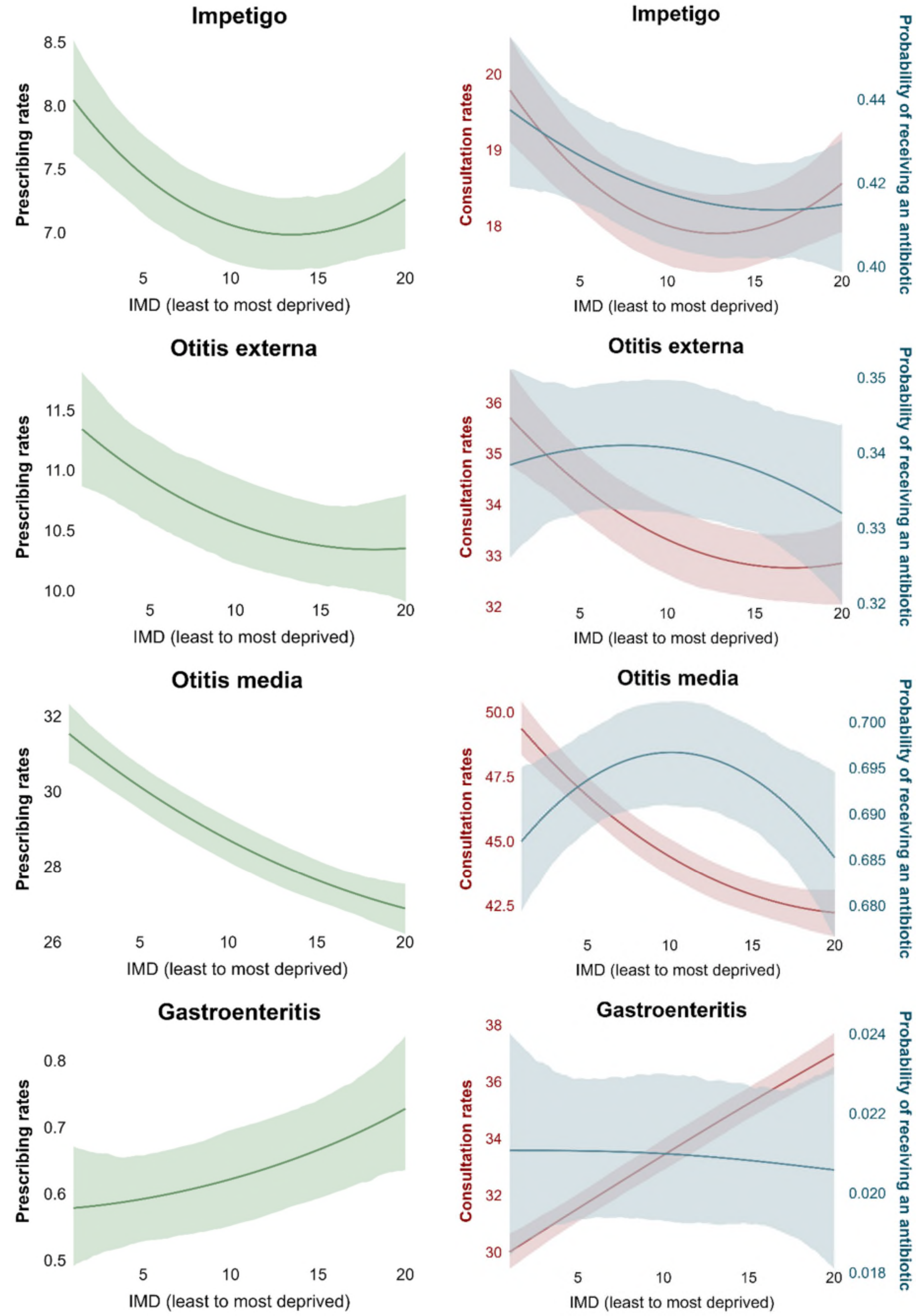

*Figure S13 (a). Mean prescribing rates, consultation rates, and probability of receiving antibiotics once consulted in children (continued)*

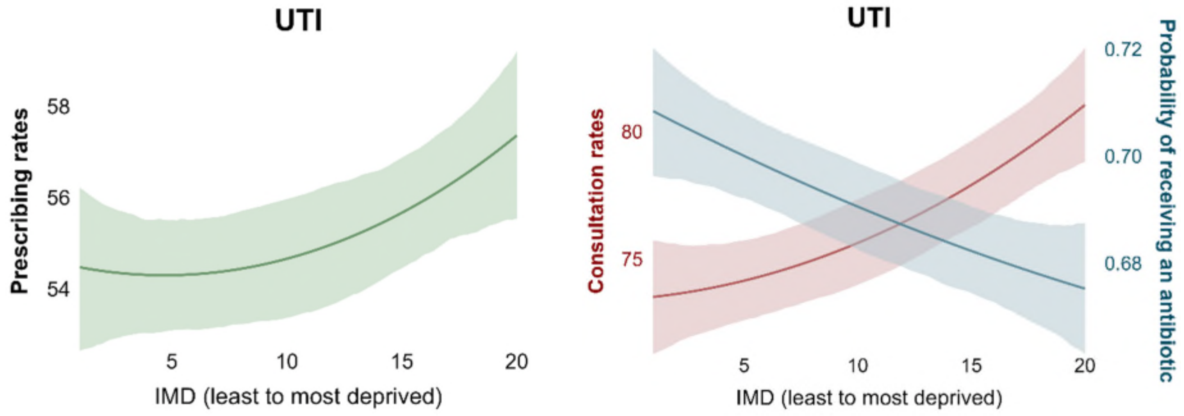


Panel (b) Adults

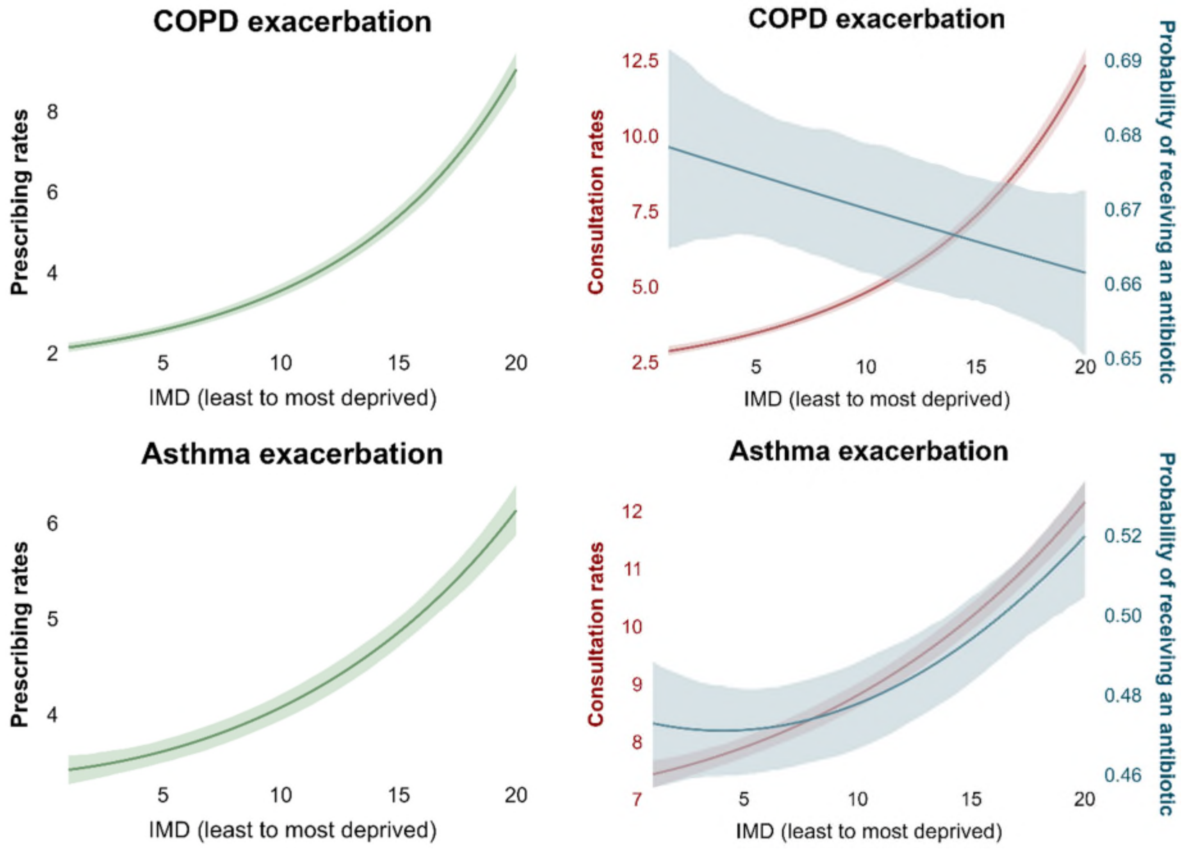

*Figure S13 (b). Mean prescribing rates, consultation rates, and probability of receiving antibiotics once consulted in adults (continued)*

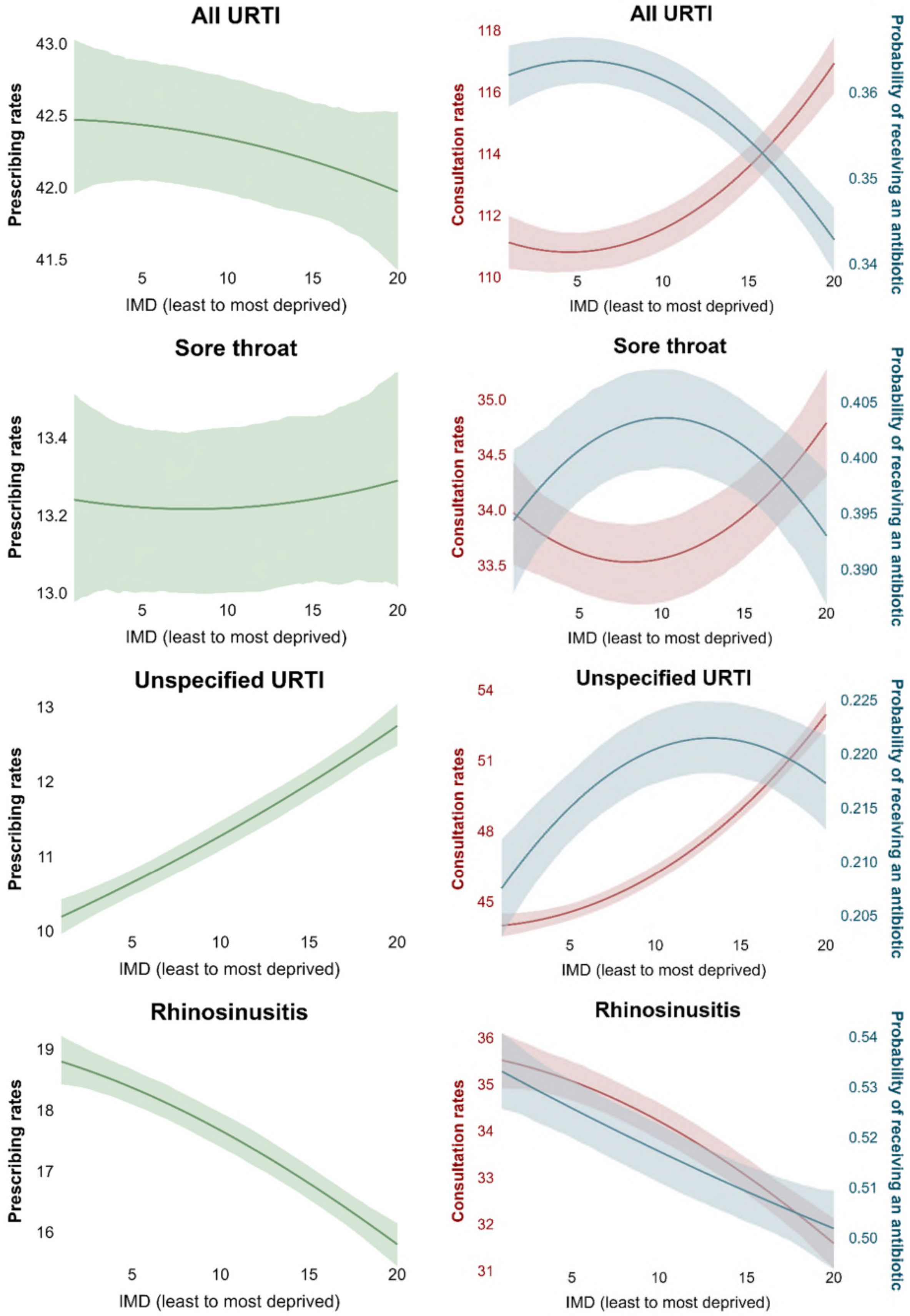

*Figure S13 (b). Mean prescribing rates, consultation rates, and probability of receiving antibiotics once consulted in adults (continued)*

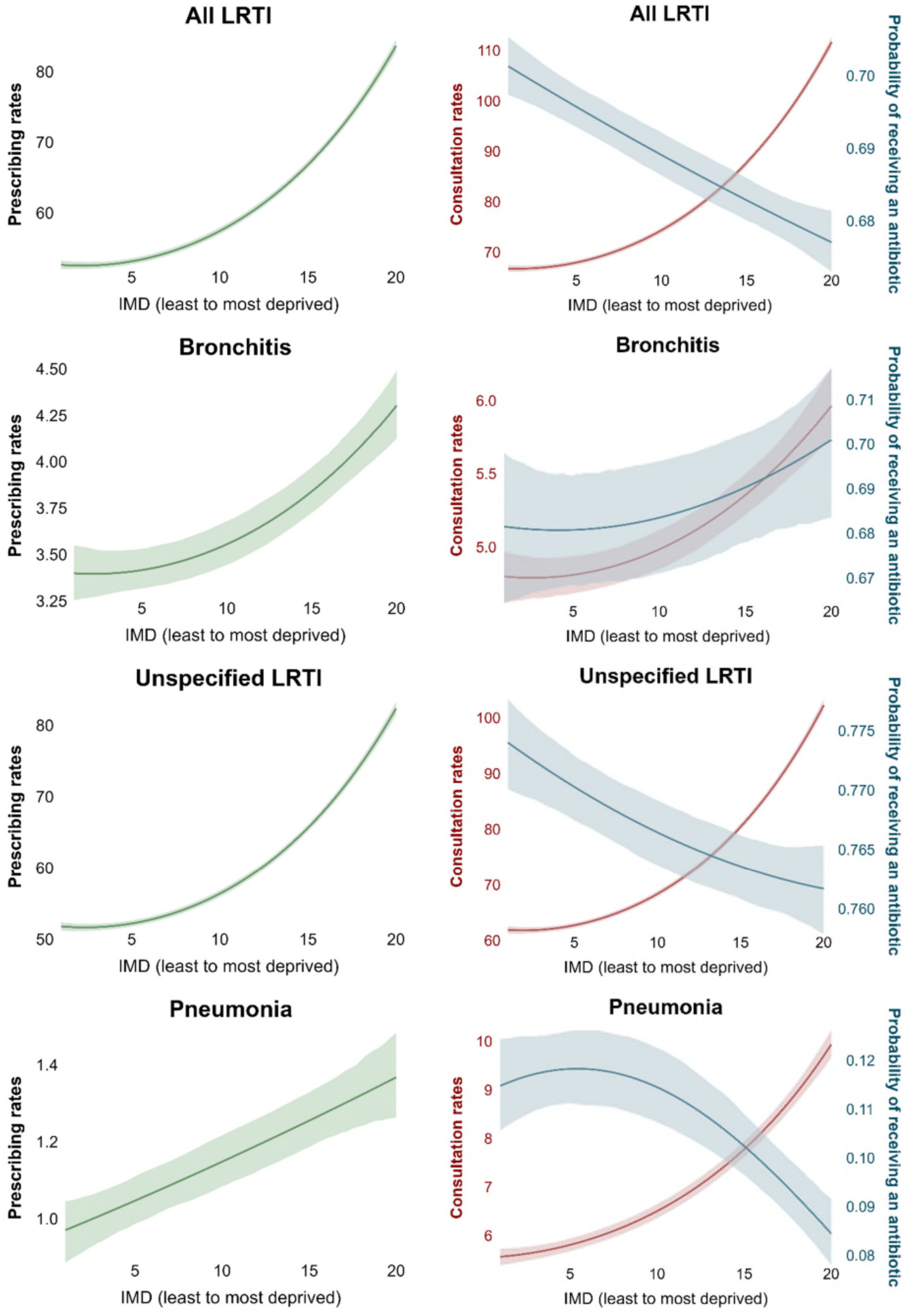

*Figure S13 (b). Mean prescribing rates, consultation rates, and probability of receiving antibiotics once consulted in adults (continued)*

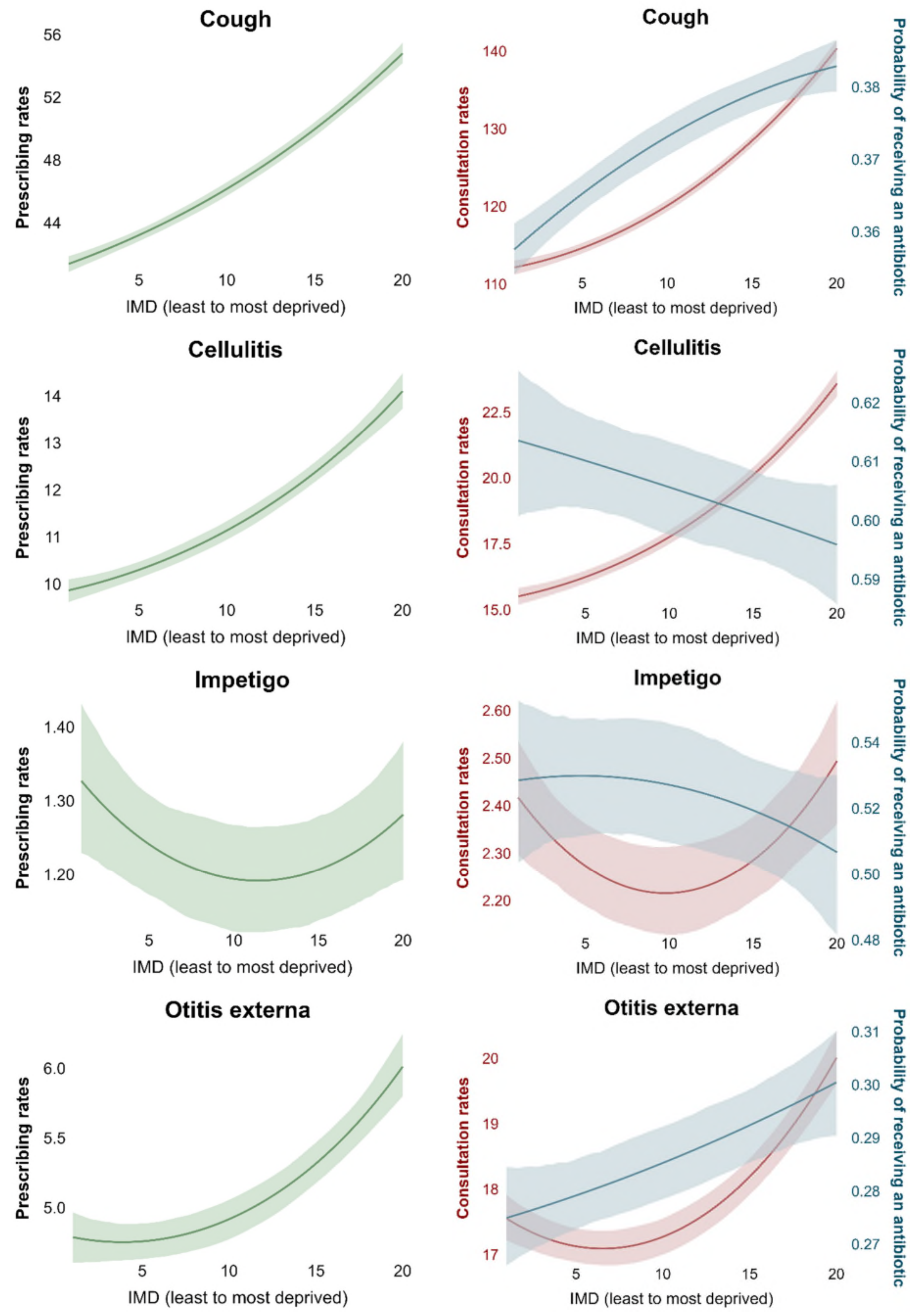

*Figure S13 (b). Mean prescribing rates, consultation rates, and probability of receiving antibiotics once consulted in adults (continued)*

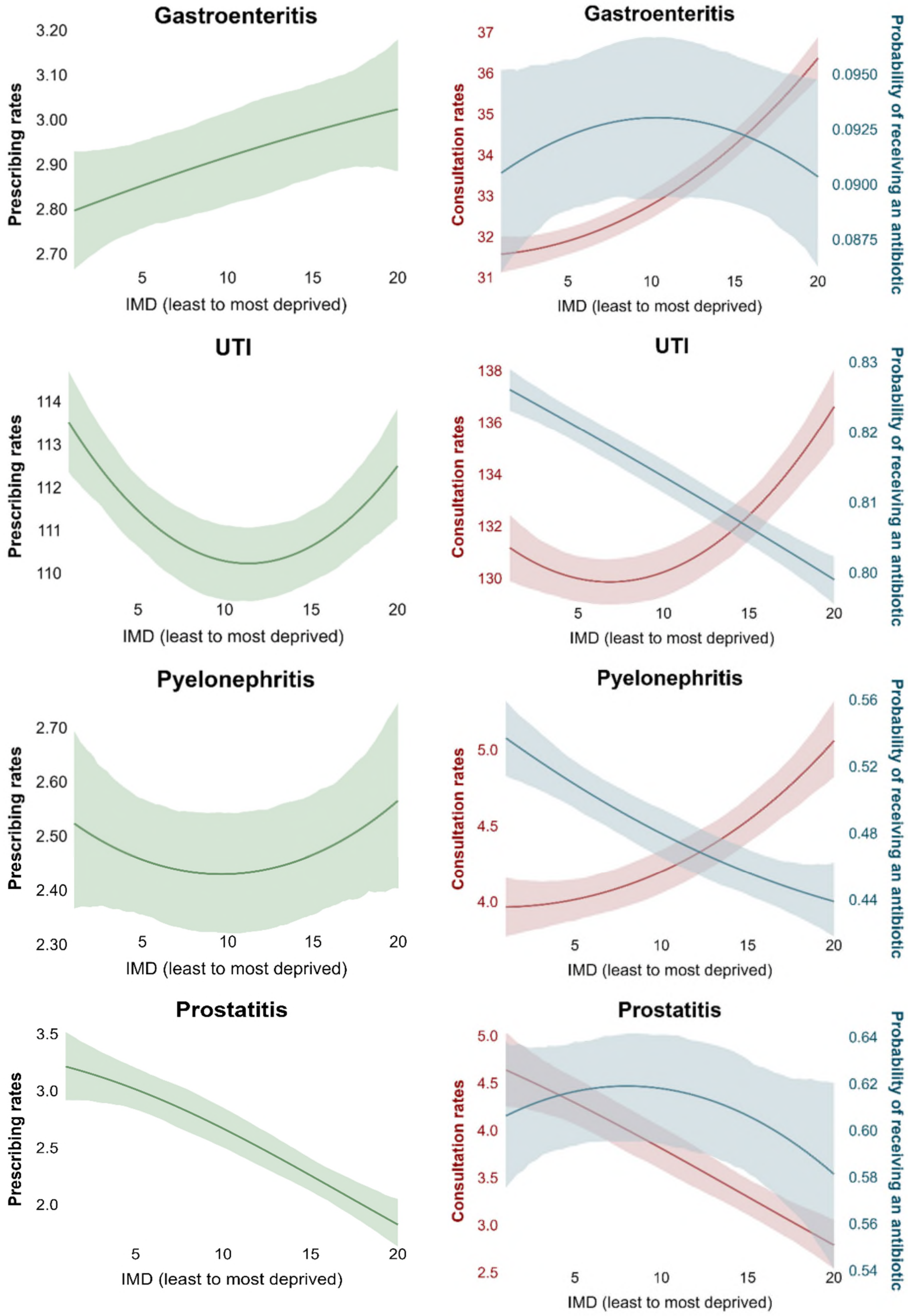

Notes: All models were adjusted for age, sex, and ethnicity. Abbreviations: IMD index of multiple deprivation, COPD chronic obstructive pulmonary disease, LRTI lower respiratory tract infections, URTI upper respiratory tract infections, UTI urinary tract infections. “All URTI” includes sore throat, unspecified upper respiratory tract infection, rhinosinusitis whereas “All LRTI” includes bronchitis, unspecified lower respiratory tract infection, pneumonia.

##### S3.3.4 Role of contributors in explaining inequalities

*Figure S14 Role of contributors in explaining inequalities in consultations in the observed population*

Panel (a) Children

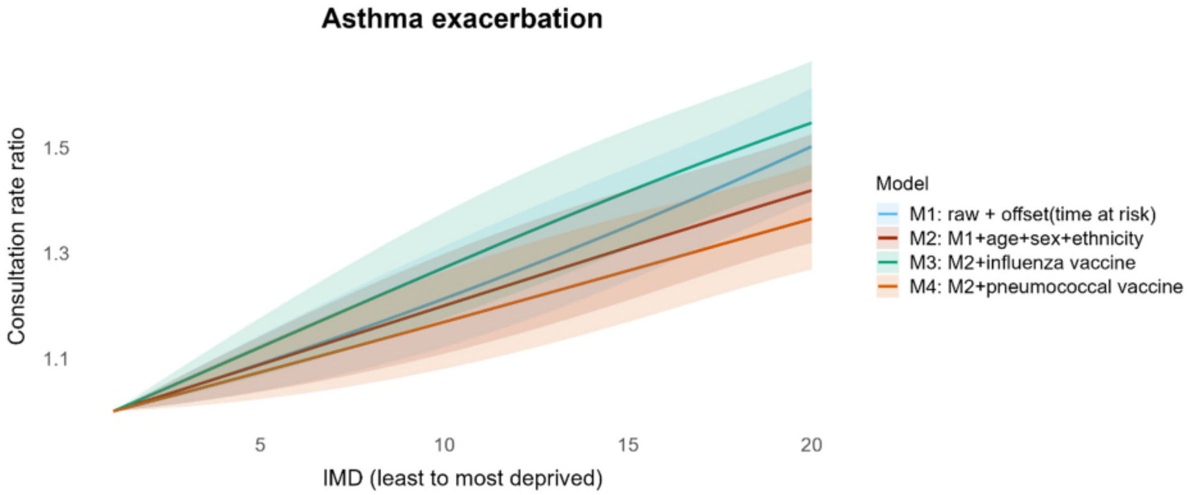


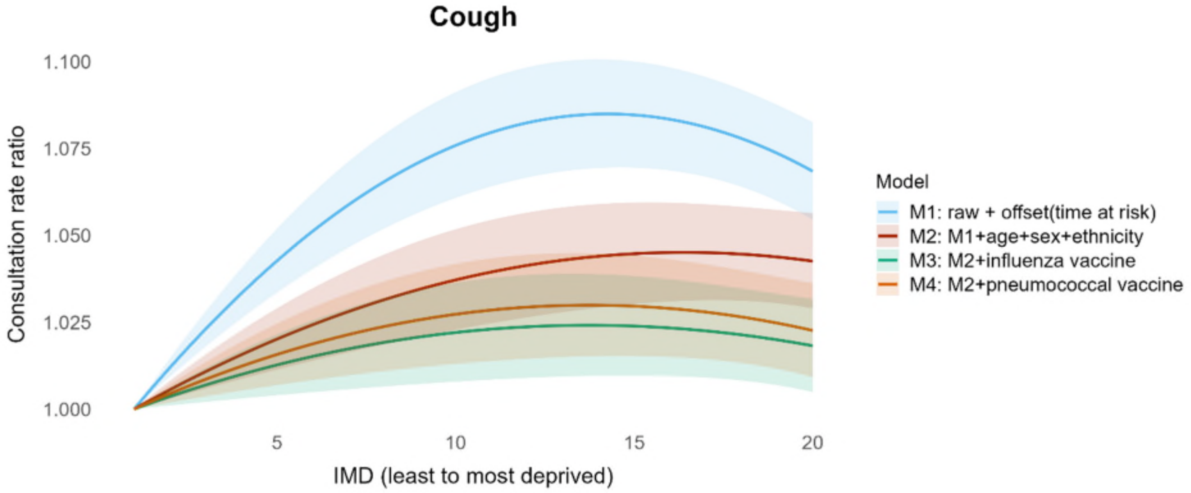


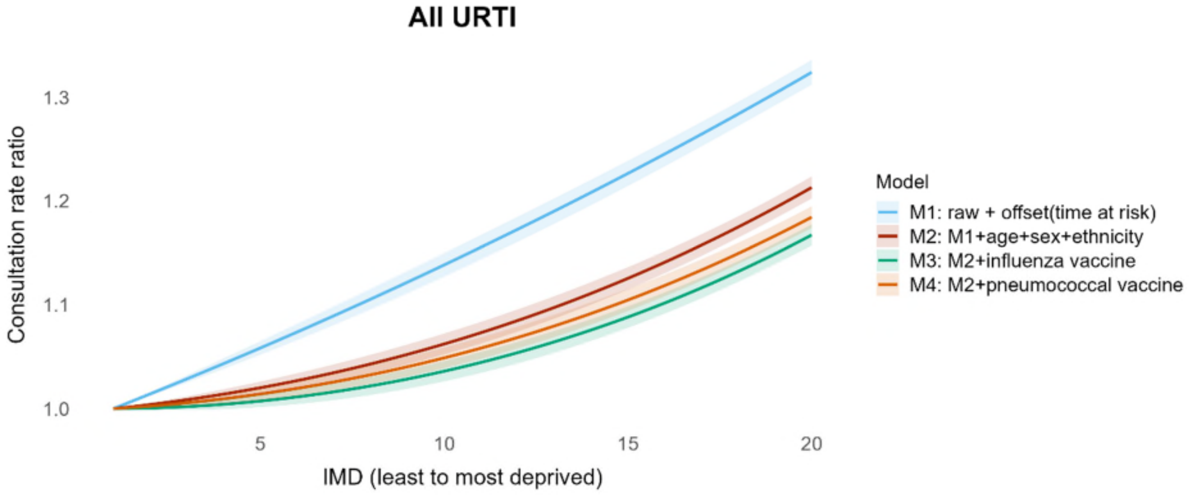

*Figure S6 (a).* Role of contributors in explaining inequalities in consultations *in children (continued)*

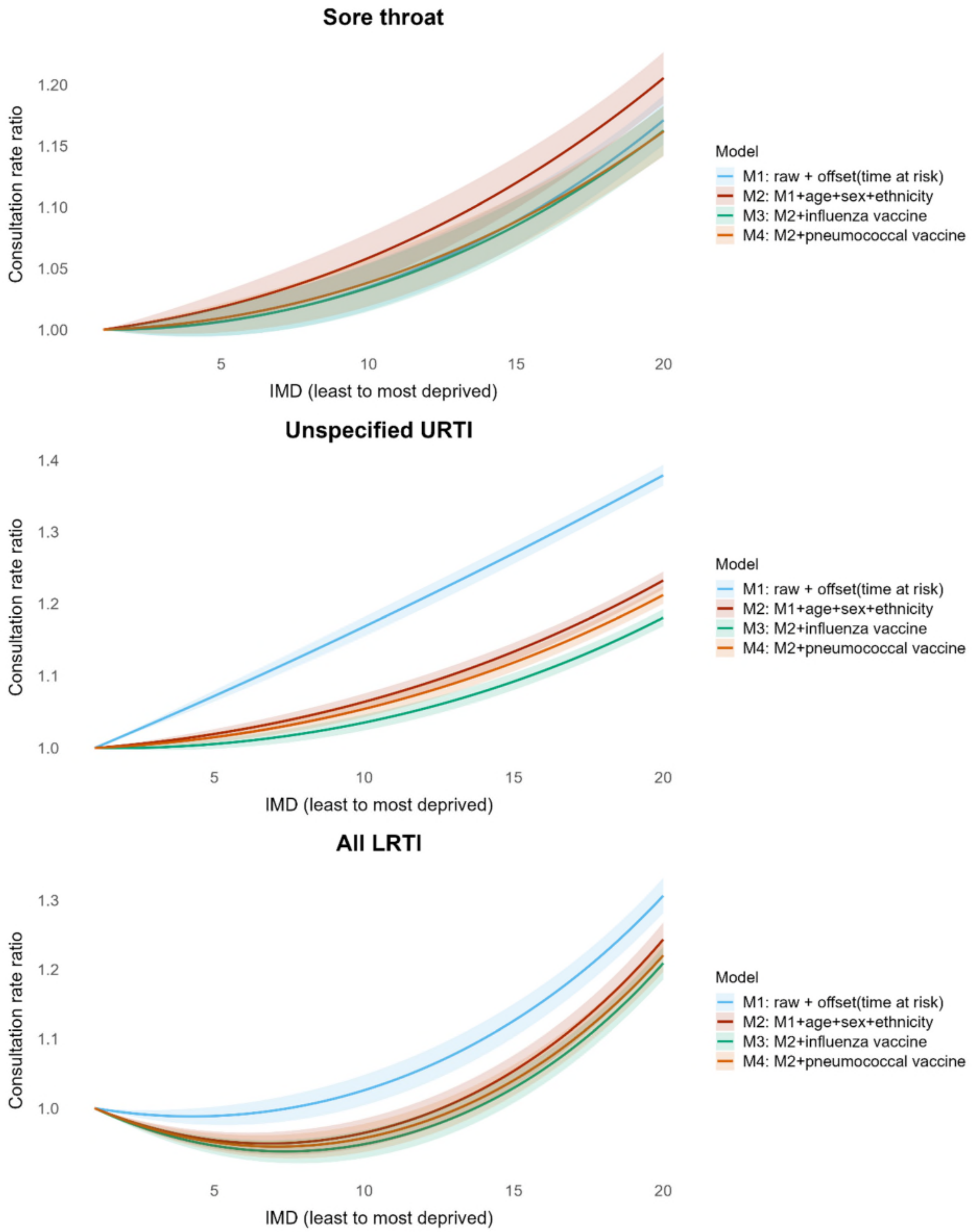

*Figure S6 (a).* Role of contributors in explaining inequalities in consultations *in children (continued)*

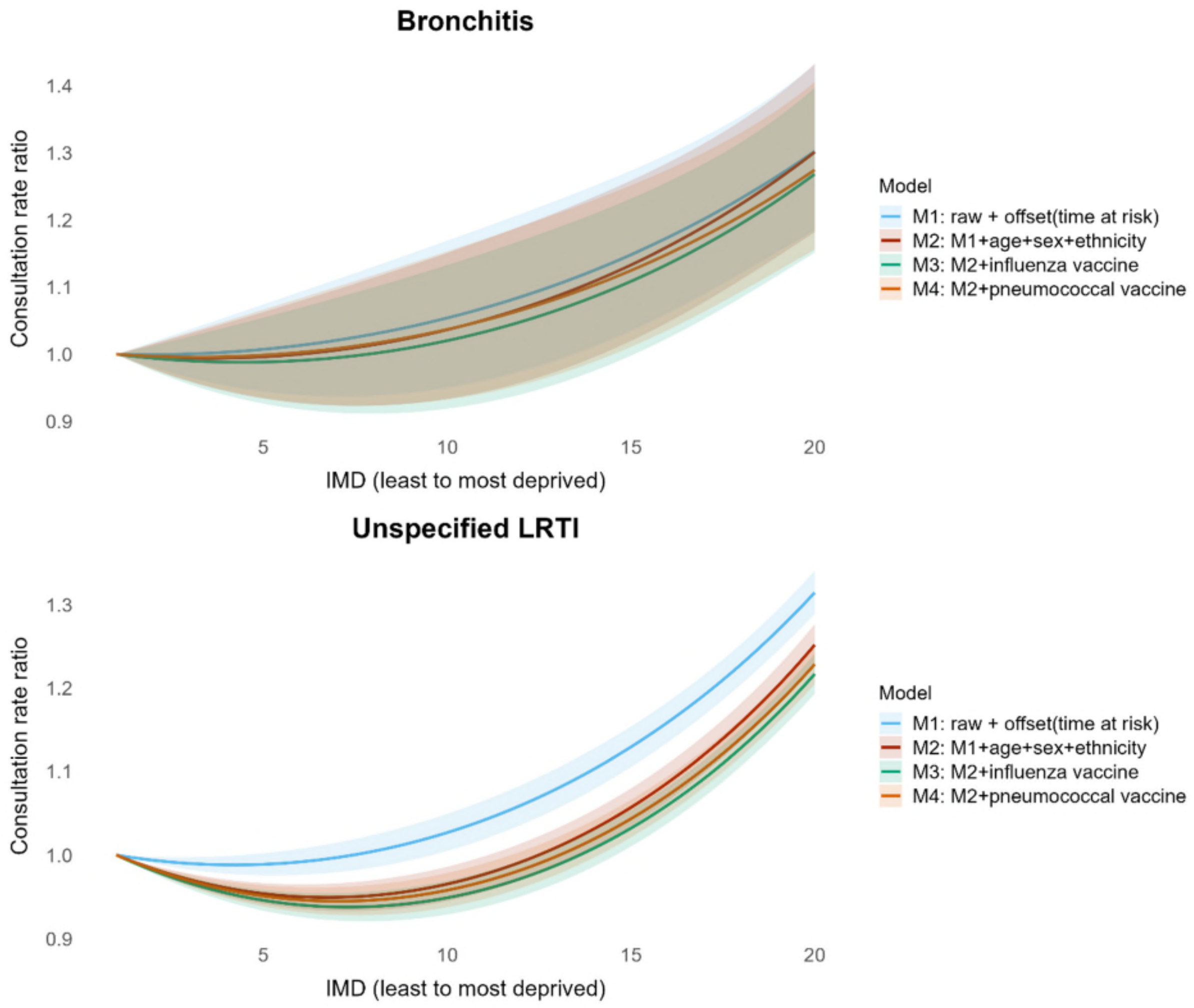

Panel (b) Adults

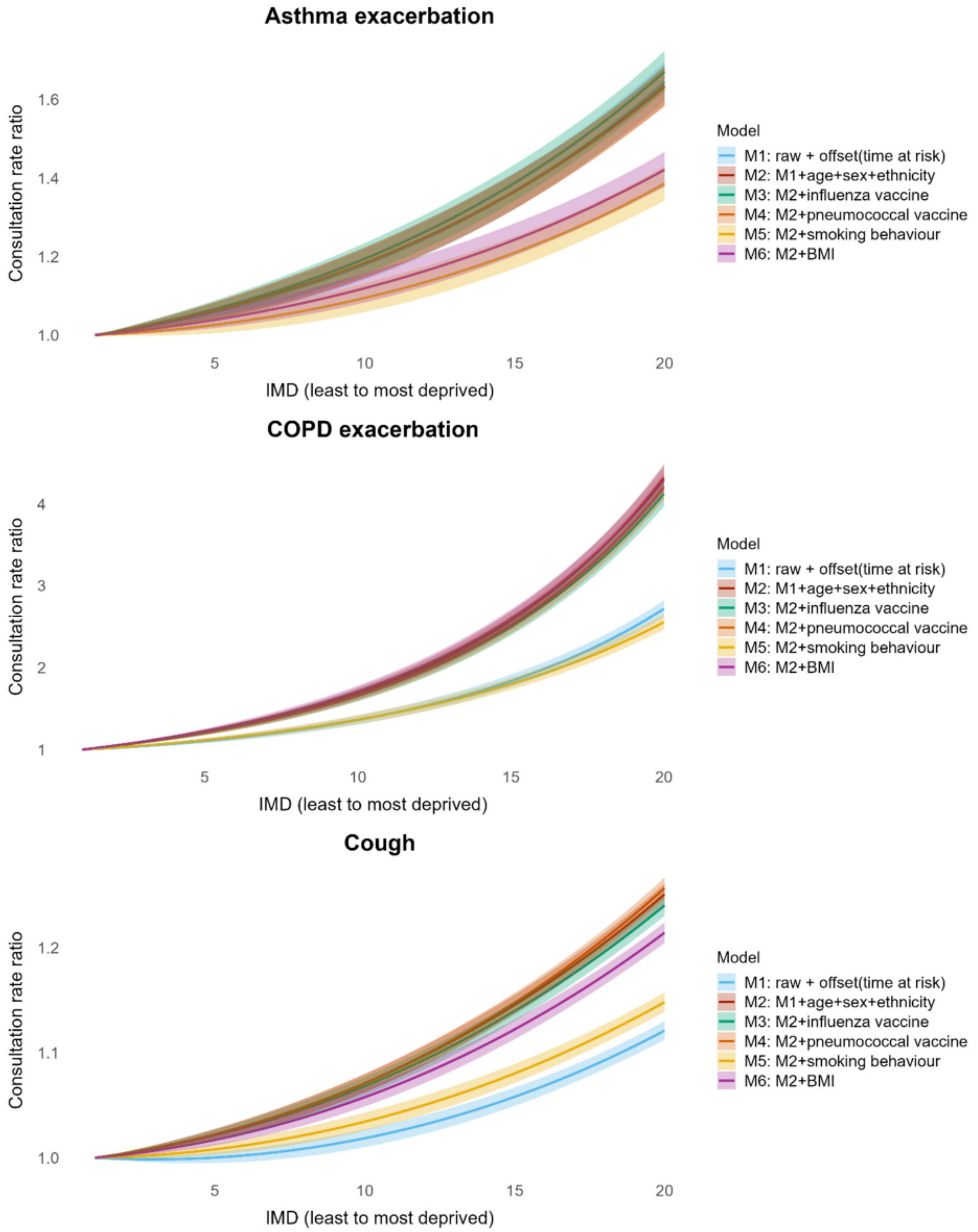

*Figure S14 (b).* Role of contributors in explaining inequalities in consultations *in adults (continued)*

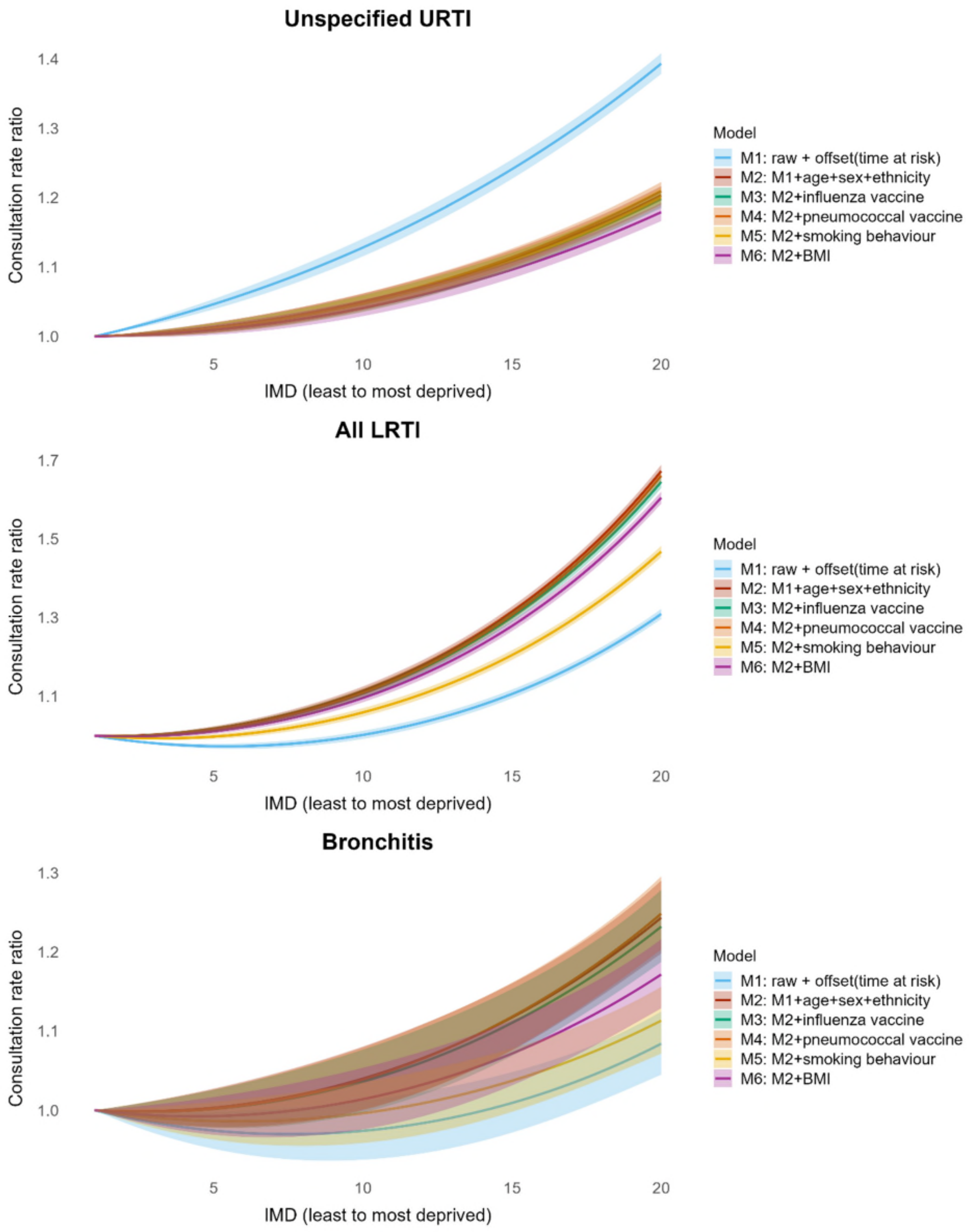

*Figure S14 (b).* Role of contributors in explaining inequalities in consultations *in adults (continued)*

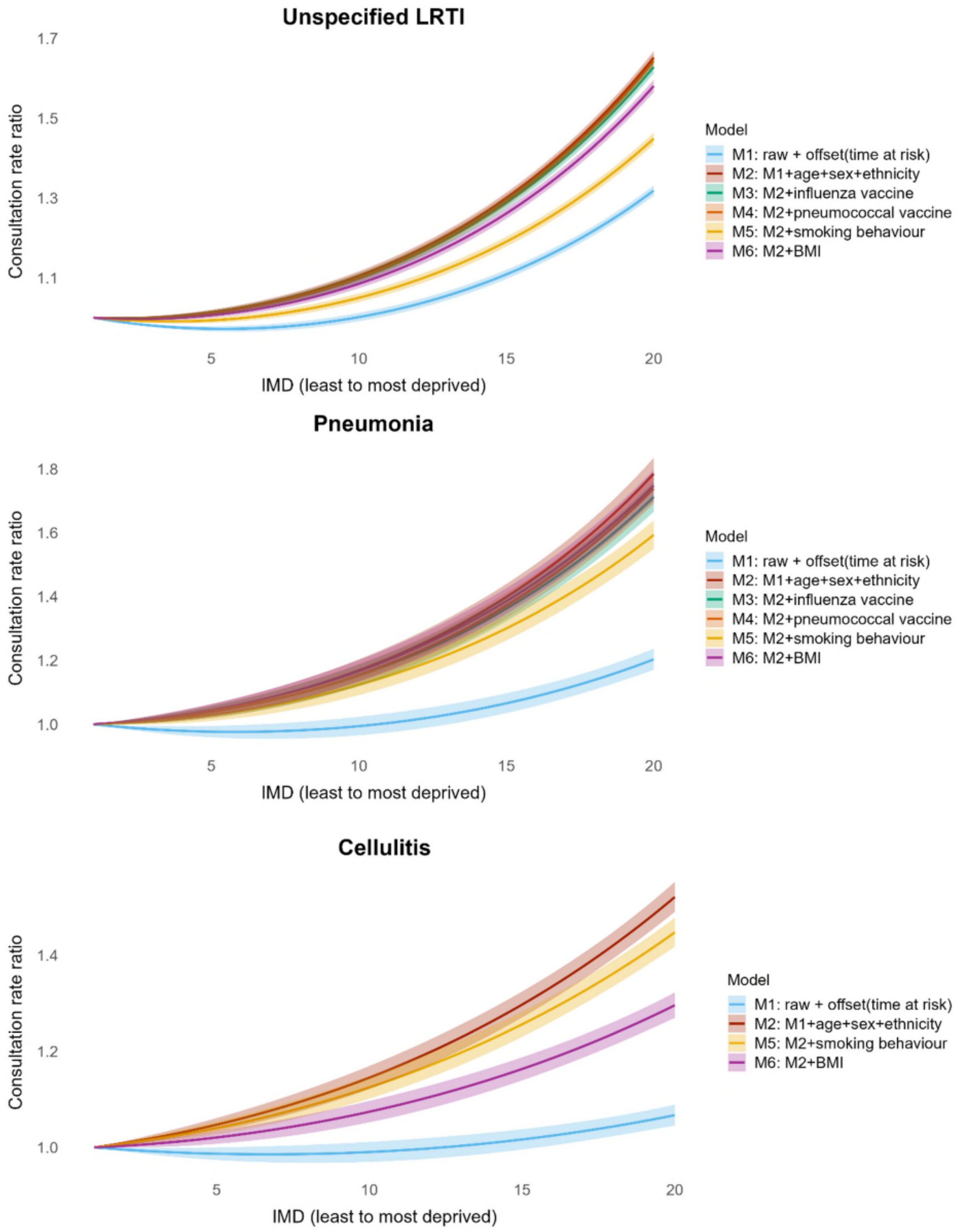

*Figure S14 (b).* Role of contributors in explaining inequalities in consultations *in adults (continued)*

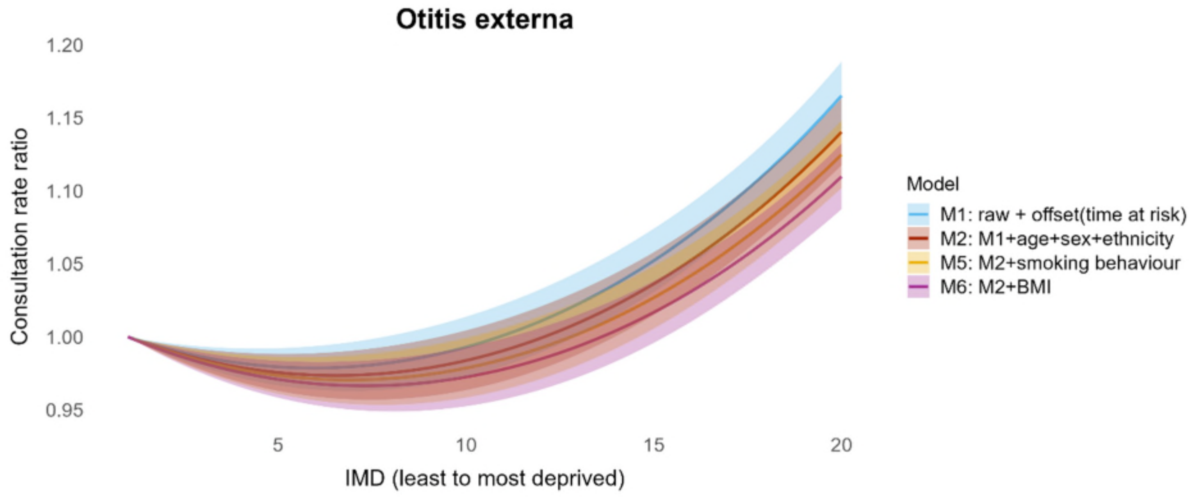


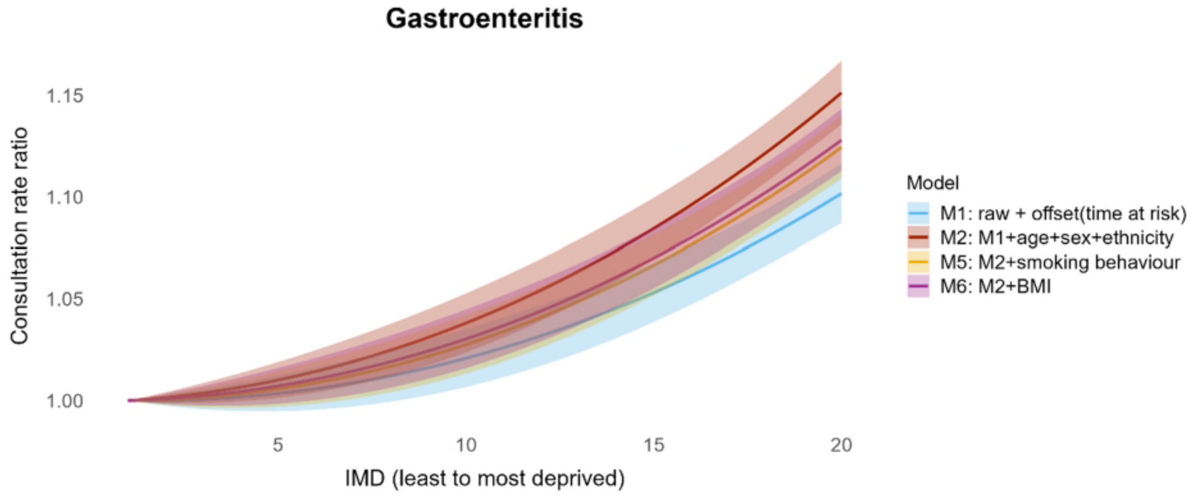

#### S3.4 Sensitivity analysis: complete case analyses in the observed population

**Figure S15 Distribution of continuous variables with missing data in the complete case and imputed dataset**

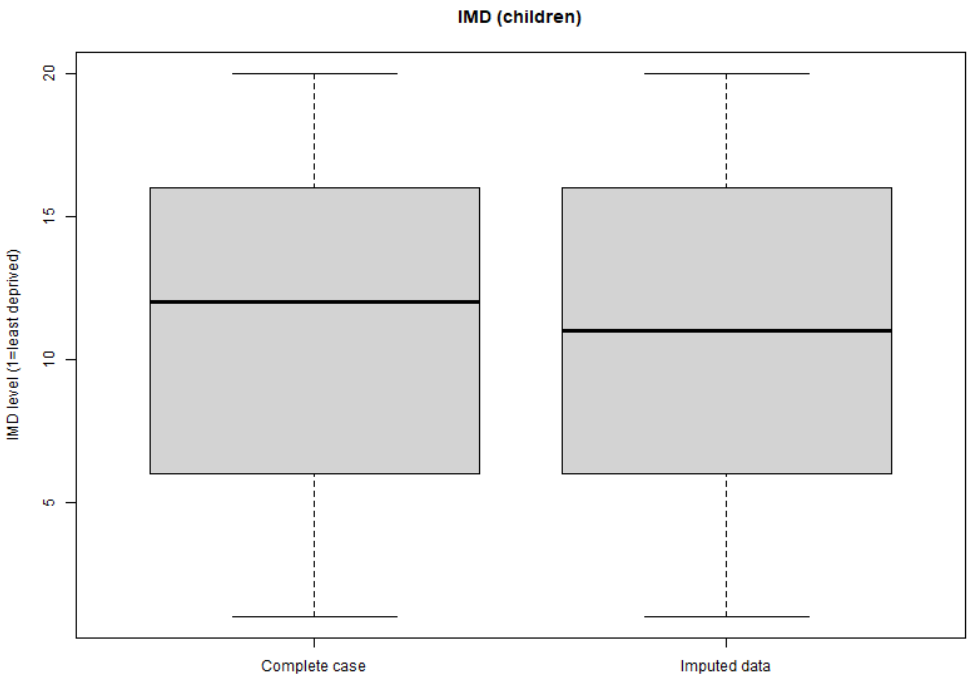


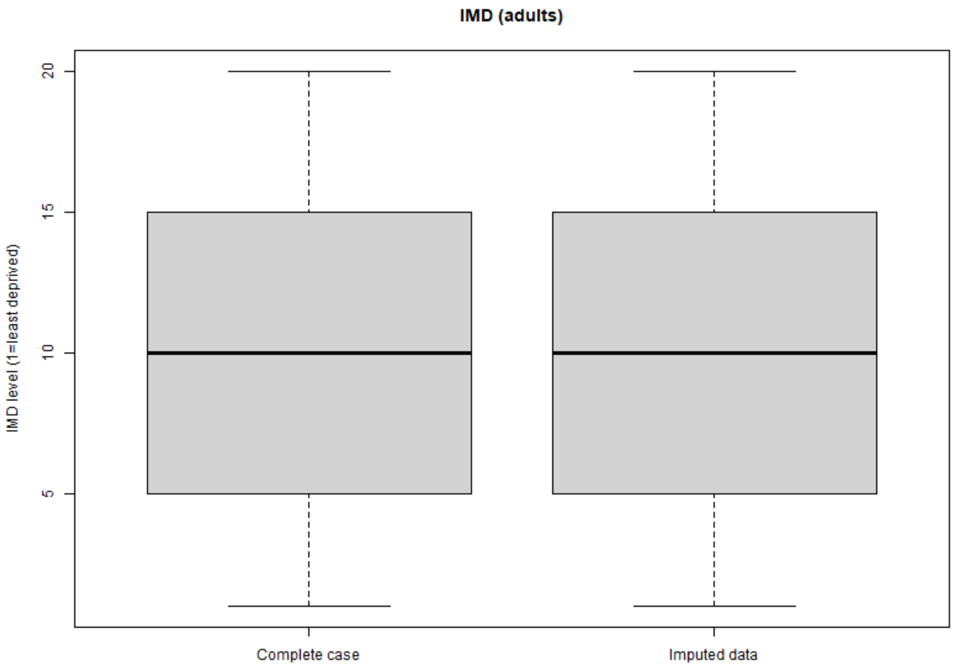

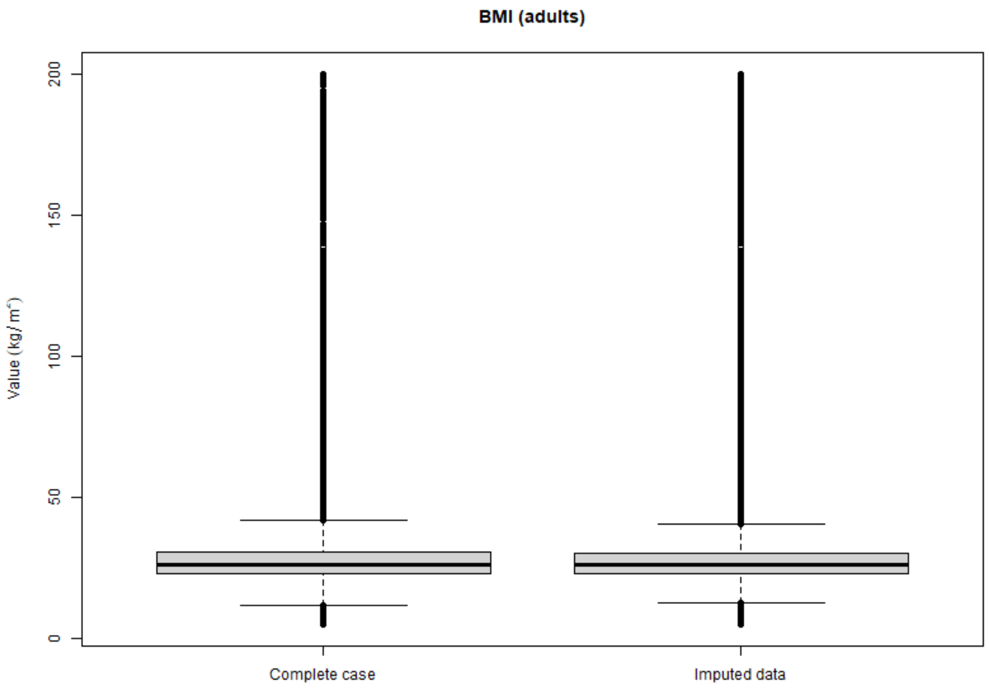


Abbreviations: IMD index of multiple deprivation, BMI body mass index. Details of missing data are shown in Table S10.

### Supplementary file 4: supplementary tables

#### S4.1 Main analysis using total population

##### S4.1.1 Descriptive results

*Table S1 Antibiotic prescribing rates (number of prescriptions per 1000 person-years) in children (<18 years) and adults (≥18 years) in the total population*

| | Prescribing rate | (95% CI) |
|---|---|---|
| **Children** | | |
| All URTI | 35.1 | (34.9 to 35.3) |
| UTI | 25.9 | (25.6 to 26.1) |
| Otitis media | 24.3 | (24.2 to 24.5) |
| All LRTI | 22.6 | (22.4 to 22.8) |
| Unspecified LRTI | 22.5 | (22.4 to 22.7) |
| Cough | 21.1 | (21.0 to 21.3) |
| Sore throat | 17.4 | (17.3 to 17.6) |
| Unspecified URTI | 17.4 | (17.2 to 17.5) |
| Otitis externa | 6.3 | (6.3 to 6.4) |
| Impetigo | 3.9 | (3.9 to 4.0) |
| Rhinosinusitis | 1.7 | (1.6 to 1.7) |
| Cellulitis | 1.0 | (1.0 to 1.1) |
| Gastroenteritis | 0.8 | (0.8 to 0.8) |
| Asthma exacerbation | 0.7 | (0.7 to 0.7) |
| Bronchitis | 0.5 | (0.5 to 0.6) |
| Pneumonia | 0.1 | (0.1 to 0.1) |
| **Adults** | | |
| UTI | 81.4 | (81.2 to 81.6) |
| All LRTI | 33.6 | (33.5 to 33.7) |
| Unspecified LRTI | 33.0 | (32.9 to 33.1) |
| Cough | 26.2 | (26.1 to 26.3) |
| All URTI | 24.3 | (24.2 to 24.3) |
| Rhinosinusitis | 9.1 | (9.0 to 9.1) |
| Sore throat | 8.9 | (8.9 to 9.0) |
| Cellulitis | 7.7 | (7.7 to 7.7) |
| Unspecified URTI | 6.9 | (6.9 to 7.0) |
| COPD exacerbation | 4.1 | (4.1 to 4.2) |
| Otitis externa | 2.5 | (2.4 to 2.5) |
| Asthma exacerbation | 2.1 | (2.1 to 2.1) |
| Bronchitis | 1.8 | (1.8 to 1.8) |
| Gastroenteritis | 1.5 | (1.5 to 1.5) |
| Pyelonephritis | 1.4 | (1.4 to 1.4) |
| Prostatitis | 0.7 | (0.7 to 0.7) |
| Impetigo | 0.6 | (0.6 to 0.7) |
| Pneumonia | 0.6 | (0.6 to 0.6) |

URTI upper respiratory tract infection, LRTI lower respiratory tract infection, UTI urinary tract infection, COPD chronic obstructive pulmonary disease, CI confidence interval. “All URTI” includes sore throat, unspecified upper respiratory tract infection, and rhinosinusitis, whereas “All LRTI” includes bronchitis, unspecified lower respiratory tract infection, pneumonia.

##### S4.1.2 Association between IMD and indication-related outcomes

*Table S2 Mean prescribing rates and differences between the most deprived (IMD=20) and least deprived (IMD=1, reference)*

| | | | Children | | Adults | |
|---|---|---|---|---|---|---|
| | Indication | Deprivation level/Δ gap | Mean | (95% CI) | Mean | (95% CI) |
| Respiratory | Asthma exacerbation | IMD=20 | 1.4 | (1.3 to 1.6) | 2.2 | (2.1 to 2.3) |
| | | IMD=1 | 0.8 | (0.6 to 0.9) | 1.5 | (1.4 to 1.5) |
| | | Δ Absolute | 0.7 | (0.5 to 0.8) | 0.8 | (0.7 to 0.8) |
| | | Δ Percentage (%) | 89.0 | (64.5 to 117.9) | 51.5 | (45.3 to 58.1) |
| | COPD exacerbation | IMD=20 | - | - | 2.2 | (2.1 to 2.3) |
| | | IMD=1 | - | - | 0.9 | (0.8 to 0.9) |
| | | Δ Absolute | - | - | 1.3 | (1.3 to 1.4) |
| | | Δ Percentage (%) | - | - | 151.1 | (140.1 to 162.2) |
| | Cough | IMD=20 | 13.9 | (13.5 to 14.3) | 19.5 | (19.2 to 19.7) |
| | | IMD=1 | 10.6 | (10.3 to 11.0) | 18.2 | (17.9 to 18.4) |
| | | Δ Absolute | 3.2 | (2.9 to 3.6) | 1.3 | (1.1 to 1.6) |
| | | Δ Percentage (%) | 30.4 | (27.1 to 33.8) | 7.3 | (6.1 to 8.8) |
| | All URTI | IMD=20 | 28.2 | (27.7 to 28.8) | 17.0 | (16.8 to 17.2) |
| | | IMD=1 | 19.5 | (19.1 to 20.0) | 17.6 | (17.4 to 17.9) |
| | | Δ Absolute | 8.7 | (8.3 to 9.2) | -0.7 | (-0.9 to -0.4) |
| | | Δ Percentage (%) | 44.8 | (41.9 to 47.7) | -3.7 | (-4.9 to -2.6) |
| | Sore throat | IMD=20 | 19.5 | (19.0 to 20.1) | 5.7 | (5.6 to 5.8) |
| | | IMD=1 | 14.5 | (14.1 to 15.0) | 5.4 | (5.3 to 5.5) |
| | | Δ Absolute | 5.0 | (4.5 to 5.4) | 0.2 | (0.1 to 0.3) |
| | | Δ Percentage (%) | 34.2 | (30.5 to 37.9) | 4.2 | (2.1 to 6.3) |
| | Rhinosinusitis | IMD=20 | 1.1 | (1.0 to 1.2) | 6.1 | (6.0 to 6.2) |
| | | IMD=1 | 0.9 | (0.9 to 1.0) | 8.1 | (7.9 to 8.2) |
| | | Δ Absolute | 0.1 | (0.0 to 0.2) | -2.0 | (-2.1 to -1.8) |
| | | Δ Percentage (%) | 14.1 | (4.3 to 24.8) | -24.4 | (-25.9 to -22.8) |
| | Unspecified URTI | IMD=20 | 8.7 | (8.5 to 9.0) | 5.2 | (5.1 to 5.3) |
| | | IMD=1 | 5.3 | (5.1 to 5.5) | 4.2 | (4.1 to 4.3) |
| | | Δ Absolute | 3.5 | (3.2 to 3.7) | 1.0 | (0.9 to 1.1) |
| | | Δ Percentage (%) | 65.3 | (60.6 to 70.0) | 23.3 | (20.5 to 26.0) |
| | All LRTI | IMD=20 | 16.2 | (15.8 to 16.6) | 28.9 | (28.6 to 29.2) |
| | | IMD=1 | 11.0 | (10.7 to 11.3) | 23.5 | (23.3 to 23.8) |
| | | Δ Absolute | 5.2 | (4.9 to 5.6) | 5.4 | (5.1 to 5.7) |
| | | Δ Percentage (%) | 47.6 | (44.2 to 51.3) | 22.7 | (21.4 to 24.1) |
| | Bronchitis | IMD=20 | 0.4 | (0.3 to 0.5) | 1.5 | (1.4 to 1.6) |
| | | IMD=1 | 0.2 | (0.2 to 0.3) | 1.5 | (1.4 to 1.6) |
| | | Δ Absolute | 0.1 | (0.1 to 0.2) | 0.0 | (-0.1 to 0.1) |
| | | Δ Percentage (%) | 60.9 | (37.0 to 87.1) | -0.2 | (-4.6 to 4.4) |
| | Unspecified LRTI | IMD=20 | 16.2 | (15.7 to 16.6) | 28.5 | (28.2 to 28.8) |
| | | IMD=1 | 10.9 | (10.6 to 11.3) | 23.2 | (22.9 to 23.4) |
| | | Δ Absolute | 5.2 | (4.9 to 5.6) | 5.3 | (5.0 to 5.6) |
| | | Δ Percentage (%) | 47.9 | (44.4 to 51.4) | 23.0 | (21.6 to 24.3) |
| | Pneumonia | IMD=20 | 0.1 | (0.1 to 0.1) | 0.5 | (0.4 to 0.5) |
| | | IMD=1 | 0.1 | (0.1 to 0.2) | 0.5 | (0.4 to 0.5) |

| | | | | | | |
|---|---|---|---|---|---|---|
| | | Δ Absolute | 0.0 | (-0.1 to 0.0) | 0.0 | (0.0 to 0.0) |
| | | Δ Percentage (%) | -8.5 | (-39.5 to 36.4) | 0.0 | (-7.6 to 8.1) |
| Skin | Cellulitis | IMD=20 | 1.1 | (1.0 to 1.2) | 4.5 | (4.4 to 4.6) |
| | | IMD=1 | 1.2 | (1.1 to 1.3) | 4.6 | (4.5 to 4.7) |
| | | Δ Absolute | -0.1 | (-0.2 to 0.0) | -0.1 | (-0.2 to 0.0) |
| | | Δ Percentage (%) | -10.0 | (-19.2 to -0.5) | -2.6 | (-4.9 to 0.1) |
| | Impetigo | IMD=20 | 4.8 | (4.5 to 5.0) | 0.5 | (0.5 to 0.6) |
| | | IMD=1 | 5.1 | (4.8 to 5.4) | 0.6 | (0.5 to 0.6) |
| | | Δ Absolute | -0.3 | (-0.6 to -0.1) | -0.1 | (-0.1 to 0.0) |
| | | Δ Percentage (%) | -6.3 | (-11.4 to -1.3) | -12.1 | (-18.1 to -5.5) |
| Ear | Otitis externa | IMD=20 | 7.0 | (6.6 to 7.3) | 2.3 | (2.2 to 2.4) |
| | | IMD=1 | 7.0 | (6.7 to 7.3) | 2.0 | (2.0 to 2.1) |
| | | Δ Absolute | 0.0 | (-0.3 to 0.3) | 0.3 | (0.2 to 0.4) |
| | | Δ Percentage (%) | -0.5 | (-4.7 to 3.9) | 14.5 | (10.2 to 18.6) |
| | Otitis media | IMD=20 | 17.0 | (16.6 to 17.4) | - | - |
| | | IMD=1 | 17.9 | (17.5 to 18.4) | - | - |
| | | Δ Absolute | -0.9 | (-1.3 to -0.6) | - | - |
| | | Δ Percentage (%) | -5.2 | (-7.4 to -3.1) | - | - |
| Gastrointestinal | Gastroenteritis | IMD=20 | 0.6 | (0.5 to 0.6) | 1.1 | (1.1 to 1.2) |
| | | IMD=1 | 0.4 | (0.3 to 0.4) | 1.2 | (1.2 to 1.3) |
| | | Δ Absolute | 0.2 | (0.2 to 0.3) | -0.1 | (-0.1 to 0.0) |
| | | Δ Percentage (%) | 63.1 | (43.7 to 82.9) | -6.5 | (-10.8 to -2.0) |
| Urinary | UTI | IMD=20 | 41.8 | (40.4 to 43.2) | 61.4 | (60.7 to 62.1) |
| | | IMD=1 | 36.7 | (35.4 to 37.9) | 70.5 | (69.7 to 71.2) |
| | | Δ Absolute | 5.1 | (3.7 to 6.5) | -9.1 | (-9.9 to -8.4) |
| | | Δ Percentage (%) | 14.0 | (10.0 to 18.3) | -12.9 | (-14.0 to -11.9) |
| | Pyelonephritis | IMD=20 | - | - | 1.5 | (1.4 to 1.6) |
| | | IMD=1 | - | - | 1.5 | (1.4 to 1.6) |
| | | Δ Absolute | - | - | -0.1 | (-0.2 to 0.0) |
| | | Δ Percentage (%) | - | - | -4.3 | (-10.5 to 2.6) |
| | Prostatitis | IMD=20 | - | - | 0.6 | (0.5 to 0.7) |
| | | IMD=1 | - | - | 1.3 | (1.2 to 1.4) |
| | | Δ Absolute | - | - | -0.7 | (-0.8 to -0.6) |
| | | Δ Percentage (%) | - | - | -53.4 | (-58.6 to -47.8) |

Mean prescribing rates (prescriptions per 1000 person-years) at IMD=20 and IMD=1 were derived from baseline models adjusted for age, sex, and ethnicity. Δ absolute represents the absolute difference in mean prescribing rates between the most deprived (IMD=20) and least deprived (IMD=1, reference), and Δ Percentage represents the percentage difference, calculated by Δ absolute divided by mean prescribing rate at IMD=1. Abbreviations: IMD index of multiple deprivation, URTI upper respiratory tract infections, LRTI lower respiratory tract infections, UTI urinary tract infection, COPD chronic obstructive pulmonary disease. "All URTI" includes sore throat, unspecified upper respiratory tract infection, rhinosinusitis whereas "All LRTI" includes bronchitis, unspecified lower respiratory tract infection, pneumonia.

*Table S3 Sex-specific analyses: mean prescribing rates and differences between the most deprived (IMD=20) and least deprived (IMD=1, reference) for children*

| | | Male | | Female | | |
|---|---|---|---|---|---|---|
| Infection | Deprivation level/Δ gap | Mean | (95% CI) | Mean | (95% CI) | p value |
| *Respiratory* | | | | | | |
| Asthma exacerbation | IMD=20 | 1.4 | (1.2 to 1.6) | 1.0 | (0.9 to 1.2) | |
| | IMD=1 | 0.8 | (0.6 to 0.9) | 0.5 | (0.4 to 0.6) | |
| | Δ Absolute | 0.6 | (0.4 to 0.8) | 0.5 | (0.4 to 0.7) | |
| | Δ Percentage (%) | 81.5 | (49.9 to 113.8) | 103.2 | (63.7 to 150.2) | 0.429 |
| Cough | IMD=20 | 13.4 | (12.9 to 13.9) | 13.6 | (13.1 to 14.1) | |
| | IMD=1 | 10.3 | (9.9 to 10.7) | 10.4 | (10.0 to 10.9) | |
| | Δ Absolute | 3.1 | (2.7 to 3.5) | 3.2 | (2.8 to 3.6) | |
| | Δ Percentage (%) | 30.2 | (25.9 to 34.8) | 30.5 | (25.9 to 35.0) | 0.944 |
| **All URTI** | IMD=20 | 25.6 | (24.9 to 26.3) | 33.5 | (32.6 to 34.5) | |
| | IMD=1 | 18.2 | (17.6 to 18.8) | 22.6 | (21.9 to 23.2) | |
| | Δ Absolute | 7.4 | (6.7 to 8.1) | 10.9 | (10.2 to 11.7) | |
| | **Δ Percentage (%)** | **40.7** | **(36.7 to 44.7)** | **48.5** | **(44.5 to 52.3)** | **0.006** |
| **Sore throat** | IMD=20 | 18.0 | (17.3 to 18.6) | 25.3 | (24.5 to 26.2) | |
| | IMD=1 | 13.9 | (13.3 to 14.4) | 18.4 | (17.7 to 19.1) | |
| | Δ Absolute | 4.1 | (3.5 to 4.8) | 6.9 | (6.2 to 7.7) | |
| | **Δ Percentage (%)** | **29.7** | **(24.5 to 35.1)** | **37.8** | **(33.0 to 43.0)** | **0.030** |
| Rhinosinusitis | IMD=20 | 1.0 | (0.9 to 1.1) | 1.3 | (1.2 to 1.5) | |
| | IMD=1 | 0.9 | (0.8 to 1.0) | 1.1 | (1.0 to 1.3) | |
| | Δ Absolute | 0.1 | (0.0 to 0.2) | 0.2 | (0.0 to 0.3) | |
| | Δ Percentage (%) | 10.9 | (-1.8 to 24.3) | 17.1 | (2.3 to 32.3) | 0.540 |
| Unspecified URTI | IMD=20 | 8.1 | (7.7 to 8.4) | 9.0 | (8.6 to 9.4) | |
| | IMD=1 | 5.0 | (4.8 to 5.3) | 5.3 | (5.0 to 5.6) | |
| | Δ Absolute | 3.1 | (2.8 to 3.3) | 3.7 | (3.4 to 4.0) | |
| | Δ Percentage (%) | 60.9 | (54.5 to 67.5) | 70.1 | (63.4 to 77.1) | 0.058 |
| All LRTI | IMD=20 | 15.7 | (15.1 to 16.3) | 15.2 | (14.6 to 15.8) | |
| | IMD=1 | 10.9 | (10.5 to 11.3) | 10.0 | (9.6 to 10.4) | |
| | Δ Absolute | 4.8 | (4.4 to 5.3) | 5.1 | (4.7 to 5.6) | |
| | Δ Percentage (%) | 44.3 | (39.8 to 49.4) | 51.3 | (46.0 to 56.7) | 0.057 |
| Bronchitis | IMD=20 | 0.4 | (0.3 to 0.5) | 0.3 | (0.3 to 0.4) | |
| | IMD=1 | 0.2 | (0.2 to 0.3) | 0.2 | (0.2 to 0.3) | |
| | Δ Absolute | 0.2 | (0.1 to 0.2) | 0.1 | (0.1 to 0.2) | |
| | Δ Percentage (%) | 69.9 | (38.4 to 107.6) | 50.0 | (20.4 to 82.0) | 0.401 |
| Unspecified LRTI | IMD=20 | 15.6 | (15.1 to 16.2) | 15.1 | (14.5 to 15.6) | |
| | IMD=1 | 10.8 | (10.4 to 11.2) | 10.0 | (9.5 to 10.4) | |
| | Δ Absolute | 4.8 | (4.4 to 5.3) | 5.1 | (4.7 to 5.5) | |
| | Δ Percentage (%) | 44.9 | (40.3 to 49.6) | 51.3 | (45.8 to 56.4) | 0.078 |
| **Pneumonia** | IMD=20 | 0.1 | (0.0 to 0.1) | 0.2 | (0.1 to 0.3) | |
| | IMD=1 | 0.1 | (0.1 to 0.2) | 0.1 | (0.1 to 0.2) | |
| | Δ Absolute | -0.1 | (-0.1 to 0.0) | 0.1 | (0.0 to 0.2) | |
| | **Δ Percentage (%)** | **-43.2** | **(-65.0 to -11.3)** | **77.0** | **(-5.8 to 209.4)** | **0.034** |
| **Cellulitis** | IMD=20 | 1.2 | (1.0 to 1.4) | 1.0 | (0.8 to 1.1) | |
| | IMD=1 | 1.2 | (1.0 to 1.4) | 1.2 | (1.1 to 1.4) | |

| | | | | | | |
|---|---|---|---|---|---|---|
| | Δ Absolute | 0.0 | (-0.2 to 0.2) | -0.3 | (-0.4 to -0.1) | |
| | **Δ Percentage (%)** | **2.1** | **(-11.7 to 18.4)** | **-21.5** | **(-32.6 to -9.6)** | **0.015** |
| *Skin* | | | | | | |
| **Impetigo** | IMD=20 | 4.4 | (4.1 to 4.8) | 5.1 | (4.7 to 5.5) | |
| | IMD=1 | 5.0 | (4.6 to 5.4) | 5.1 | (4.7 to 5.5) | |
| | Δ Absolute | -0.6 | (-0.9 to -0.2) | 0.0 | (-0.4 to 0.4) | |
| | **Δ Percentage (%)** | **-11.5** | **(-17.7 to -4.7)** | **-0.4** | **(-7.8 to 7.9)** | **0.032** |
| Otitis externa | IMD=20 | 6.4 | (6.0 to 6.9) | 8.0 | (7.5 to 8.5) | |
| | IMD=1 | 6.7 | (6.3 to 7.1) | 7.8 | (7.3 to 8.3) | |
| | Δ Absolute | -0.3 | (-0.7 to 0.1) | 0.3 | (-0.2 to 0.8) | |
| | Δ Percentage (%) | -4.4 | (-10.1 to 1.6) | 3.5 | (-2.9 to 10.0) | 0.077 |
| *Ear* | | | | | | |
| **Otitis media** | IMD=20 | 14.6 | (14.1 to 15.2) | 19.0 | (18.4 to 19.7) | |
| | IMD=1 | 16.0 | (15.4 to 16.6) | 19.3 | (18.6 to 20.0) | |
| | Δ Absolute | -1.4 | (-1.9 to -0.8) | -0.3 | (-0.9 to 0.4) | |
| | **Δ Percentage (%)** | **-8.5** | **(-11.6 to -5.3)** | **-1.4** | **(-4.6 to 2.0)** | **0.002** |
| *Gastrointestinal* | | | | | | |
| Gastroenteritis | IMD=20 | 0.4 | (0.3 to 0.5) | 0.6 | (0.5 to 0.8) | |
| | IMD=1 | 0.3 | (0.2 to 0.3) | 0.4 | (0.3 to 0.5) | |
| | Δ Absolute | 0.1 | (0.1 to 0.2) | 0.3 | (0.2 to 0.4) | |
| | Δ Percentage (%) | 52.5 | (28.9 to 78.3) | 71.3 | (42.8 to 105.1) | 0.354 |

Mean prescribing rates (prescriptions per 1000 person-years) at IMD=20 and IMD=1 were derived from baseline models adjusted for age, sex, and ethnicity. Δ absolute represents the absolute difference in mean prescribing rates between the most deprived (IMD=20) and least deprived (IMD=1, reference), and Δ Percentage represents the percentage difference, calculated by Δ absolute divided by mean prescribing rate at IMD=1. P-values are from t-test comparing percentage difference between males and females. Values in bold indicate statistically significant differences at the 5% level. Abbreviations: IMD index of multiple deprivation, URTI upper respiratory tract infections, LRTI lower respiratory tract infections, UTI urinary tract infection, COPD chronic obstructive pulmonary disease. "All URTI" includes sore throat, unspecified upper respiratory tract infection, rhinosinusitis whereas "All LRTI" includes bronchitis, unspecified lower respiratory tract infection, pneumonia.

*Table S4 Sex-specific analyses: Mean prescribing rates and differences between the most deprived (IMD=20) and least deprived (IMD=1, reference) for adults*

| | | Male | | Female | | |
|---|---|---|---|---|---|---|
| Infection | Deprivation level/Δ gap | Mean | (95% CI) | Mean | (95% CI) | p value |
| *Respiratory* | | | | | | |
| **Asthma exacerbation** | IMD=20 | 1.8 | (1.7 to 2.0) | 4.9 | (4.7 to 5.1) | |
| | IMD=1 | 1.5 | (1.4 to 1.6) | 2.9 | (2.8 to 3.1) | |
| | Δ Absolute | 0.3 | (0.2 to 0.5) | 2.0 | (1.8 to 2.2) | |
| | **Δ Percentage (%)** | **22.0** | **(13.3 to 31.1)** | **68.5** | **(59.5 to 77.3)** | **0.000** |
| **COPD exacerbation** | IMD=20 | 1.6 | (1.5 to 1.7) | 3.2 | (2.9 to 3.4) | |
| | IMD=1 | 0.8 | (0.7 to 0.9) | 1.1 | (1.0 to 1.2) | |
| | Δ Absolute | 0.8 | (0.7 to 0.9) | 2.1 | (1.9 to 2.3) | |
| | **Δ Percentage (%)** | **104.8** | **(91.4 to 118.1)** | **197.9** | **(179.7 to 216.0)** | **0.000** |
| **Cough** | IMD=20 | 16.0 | (15.7 to 16.3) | 29.8 | (29.3 to 30.2) | |
| | IMD=1 | 16.9 | (16.6 to 17.2) | 25.5 | (25.1 to 25.8) | |
| | Δ Absolute | -0.9 | (-1.3 to -0.6) | 4.3 | (3.9 to 4.7) | |
| | **Δ Percentage (%)** | **-5.5** | **(-7.3 to -3.7)** | **16.9** | **(15.1 to 18.7)** | **0.000** |
| **All URTI** | IMD=20 | 14.8 | (14.5 to 15.1) | 37.7 | (37.1 to 38.2) | |
| | IMD=1 | 16.6 | (16.3 to 17.0) | 37.6 | (37.1 to 38.1) | |

| | | | | | | |
|---|---|---|---|---|---|---|
| | Δ Absolute | -1.9 | (-2.2 to -1.5) | 0.1 | (-0.5 to 0.6) | |
| | **Δ Percentage (%)** | **-11.1** | **(-13.0 to -9.4)** | **0.2** | **(-1.3 to 1.7)** | **0.000** |
| **Sore throat** | IMD=20 | 5.3 | (5.1 to 5.4) | 11.5 | (11.2 to 11.7) | |
| | IMD=1 | 5.4 | (5.2 to 5.6) | 10.7 | (10.4 to 10.9) | |
| | Δ Absolute | -0.1 | (-0.3 to 0.0) | 0.8 | (0.5 to 1.1) | |
| | **Δ Percentage (%)** | **-2.4** | **(-5.7 to 0.7)** | **7.6** | **(5.0 to 10.1)** | **0.000** |
| **Rhinosinusitis** | IMD=20 | 5.2 | (5.0 to 5.4) | 16.2 | (15.9 to 16.6) | |
| | IMD=1 | 7.9 | (7.6 to 8.1) | 20.4 | (19.9 to 20.8) | |
| | Δ Absolute | -2.6 | (-2.9 to -2.4) | -4.1 | (-4.6 to -3.7) | |
| | **Δ Percentage (%)** | **-33.6** | **(-36.3 to -30.9)** | **-20.3** | **(-22.4 to -18.2)** | **0.000** |
| **Unspecified URTI** | IMD=20 | 4.4 | (4.3 to 4.6) | 9.9 | (9.7 to 10.2) | |
| | IMD=1 | 3.9 | (3.8 to 4.1) | 7.7 | (7.5 to 7.9) | |
| | Δ Absolute | 0.5 | (0.3 to 0.6) | 2.3 | (2.0 to 2.5) | |
| | **Δ Percentage (%)** | **12.6** | **(8.6 to 16.9)** | **29.5** | **(25.8 to 33.0)** | **0.000** |
| **All LRTI** | IMD=20 | 24.1 | (23.7 to 24.5) | 43.4 | (42.8 to 43.9) | |
| | IMD=1 | 22.2 | (21.8 to 22.5) | 32.5 | (32.1 to 32.9) | |
| | Δ Absolute | 2.0 | (1.5 to 2.4) | 10.9 | (10.3 to 11.4) | |
| | **Δ Percentage (%)** | **8.8** | **(6.9 to 10.7)** | **33.4** | **(31.5 to 35.3)** | **0.000** |
| **Bronchitis** | IMD=20 | 1.3 | (1.3 to 1.4) | 2.4 | (2.3 to 2.5) | |
| | IMD=1 | 1.5 | (1.4 to 1.6) | 2.3 | (2.1 to 2.4) | |
| | Δ Absolute | -0.1 | (-0.2 to 0.0) | 0.1 | (0.0 to 0.2) | |
| | **Δ Percentage (%)** | **-8.4** | **(-14.6 to -1.8)** | **5.2** | **(-0.8 to 11.0)** | **0.002** |
| **Unspecified LRTI** | IMD=20 | 23.8 | (23.4 to 24.2) | 42.8 | (42.2 to 43.4) | |
| | IMD=1 | 21.9 | (21.5 to 22.3) | 32.0 | (31.6 to 32.4) | |
| | Δ Absolute | 1.9 | (1.5 to 2.3) | 10.8 | (10.2 to 11.4) | |
| | **Δ Percentage (%)** | **8.7** | **(6.6 to 10.6)** | **33.7** | **(31.8 to 35.8)** | **0.000** |
| Pneumonia | IMD=20 | 0.4 | (0.3 to 0.4) | 0.6 | (0.5 to 0.7) | |
| | IMD=1 | 0.4 | (0.3 to 0.4) | 0.6 | (0.5 to 0.6) | |
| | Δ Absolute | 0.0 | (-0.1 to 0.0) | 0.0 | (0.0 to 0.1) | |
| | Δ Percentage (%) | -5.1 | (-15.7 to 7.1) | 4.2 | (-6.5 to 15.8) | 0.255 |
| *Skin* | | | | | | |
| **Cellulitis** | IMD=20 | 4.7 | (4.5 to 4.8) | 4.4 | (4.3 to 4.6) | |
| | IMD=1 | 5.2 | (5.0 to 5.4) | 4.3 | (4.1 to 4.4) | |
| | Δ Absolute | -0.5 | (-0.7 to -0.4) | 0.2 | (0.0 to 0.3) | |
| | **Δ Percentage (%)** | **-10.0** | **(-13.1 to -6.9)** | **3.6** | **(0.5 to 6.9)** | **0.000** |
| Impetigo | IMD=20 | 0.5 | (0.4 to 0.5) | 0.8 | (0.8 to 0.9) | |
| | IMD=1 | 0.6 | (0.5 to 0.6) | 0.9 | (0.8 to 1.0) | |
| | Δ Absolute | -0.1 | (-0.2 to 0.0) | -0.1 | (-0.2 to 0.0) | |
| | Δ Percentage (%) | -16.4 | (-26.2 to -6.3) | -9.6 | (-18.0 to -0.4) | 0.311 |
| *Ear* | | | | | | |
| **Otitis externa** | IMD=20 | 1.9 | (1.8 to 2.1) | 4.4 | (4.2 to 4.6) | |
| | IMD=1 | 2.1 | (1.9 to 2.2) | 3.4 | (3.3 to 3.6) | |
| | Δ Absolute | -0.1 | (-0.2 to 0.0) | 0.9 | (0.7 to 1.1) | |
| | **Δ Percentage (%)** | **-5.5** | **(-11.3 to 0.2)** | **26.9** | **(20.8 to 32.6)** | **0.000** |
| *Gastrointestinal* | | | | | | |
| **Gastroenteritis** | IMD=20 | 1.1 | (1.0 to 1.2) | 1.6 | (1.5 to 1.6) | |
| | IMD=1 | 1.3 | (1.2 to 1.4) | 1.5 | (1.5 to 1.6) | |
| | Δ Absolute | -0.2 | (-0.3 to -0.1) | 0.0 | (-0.1 to 0.1) | |

| | Δ Percentage (%) | -16.0 | (-21.9 to -9.6) | 0.9 | (-5.3 to 7.1) | 0.000 |
|---|---|---|---|---|---|---|

Mean prescribing rates (prescriptions per 1000 person-years) at IMD=20 and IMD=1 were derived from baseline models adjusted for age, sex, and ethnicity. Δ absolute represents the absolute difference in mean prescribing rates between the most deprived (IMD=20) and least deprived (IMD=1, reference), and Δ Percentage represents the percentage difference, calculated by Δ absolute divided by mean prescribing rate at IMD=1. P-values are from t-test comparing percentage difference between males and females. Values in bold indicate statistically significant differences at the 5% level. Abbreviations: IMD index of multiple deprivation, URTI upper respiratory tract infections, LRTI lower respiratory tract infections, UTI urinary tract infection, COPD chronic obstructive pulmonary disease. "All URTI" includes sore throat, unspecified upper respiratory tract infection, rhinosinusitis whereas "All LRTI" includes bronchitis, unspecified lower respiratory tract infection, pneumonia.

*Table S5 Absolute and relative contribution of modifiable health factors to consultation rate differences between the most and least deprived*

Panel (a) Children

| | **Δ Observed (95% CI)** | | **Δ Counterfactual (95% CI)** | | **Absolute contribution (95% CI)** | | **% Relative contribution (95% CI)** | |
|---|---|---|---|---|---|---|---|---|
| | Influenza vaccination | | | | | | | |
| Asthma exacerbation | 1.9 | (1.7 to 2.2) | 2.3 | (2.0 to 2.6) | -0.4 | (-0.4 to -0.3) | -19.0 | (-21.6 to -16.8) |
| Cough | 19.4 | (18.1 to 20.6) | 17.5 | (16.3 to 18.8) | 1.8 | (1.7 to 2.0) | 9.5 | (8.7 to 10.4) |
| All URTI | 84.2 | (82.3 to 86.2) | 75.8 | (74.0 to 77.9) | 8.4 | (8.2 to 8.7) | 10.0 | (9.6 to 10.3) |
| Sore throat | 15.4 | (14.5 to 16.3) | 14.1 | (13.3 to 15.0) | 1.3 | (1.2 to 1.4) | 8.3 | (7.7 to 9.0) |
| Rhinosinusitis | 4.1 | (3.7 to 4.5) | 4.0 | (3.6 to 4.3) | 0.2 | (0.1 to 0.2) | 3.7 | (2.9 to 4.5) |
| Unspecified URTI | 70.9 | (69.1 to 72.6) | 63.3 | (61.5 to 65.0) | 7.6 | (7.4 to 7.9) | 10.8 | (10.4 to 11.1) |
| All LRTI | 19.8 | (18.8 to 20.8) | 18.4 | (17.4 to 19.3) | 1.4 | (1.3 to 1.5) | 7.1 | (6.5 to 7.6) |
| Bronchitis | 0.7 | (0.6 to 0.9) | 0.7 | (0.5 to 0.8) | 0.0 | (0.0 to 0.1) | 7.1 | (4.5 to 9.9) |
| Unspecified LRTI | 19.8 | (18.9 to 20.8) | 18.4 | (17.5 to 19.4) | 1.4 | (1.3 to 1.5) | 7.1 | (6.5 to 7.6) |
| | Pneumococcal vaccination | | | | | | | |
| Asthma exacerbation | 1.9 | (1.7 to 2.2) | 1.9 | (1.6 to 2.1) | 0.0 | (0.0 to 0.0) | 1.6 | (1.3 to 2.0) |
| Cough | 19.6 | (18.4 to 20.8) | 18.9 | (17.7 to 20.1) | 0.7 | (0.7 to 0.8) | 3.7 | (3.3 to 4.1) |
| All URTI | 85.3 | (83.2 to 87.3) | 83.2 | (81.1 to 85.2) | 2.1 | (1.8 to 2.3) | 2.4 | (2.2 to 2.7) |
| Sore throat | 15.6 | (14.7 to 16.5) | 15.0 | (14.1 to 16.0) | 0.6 | (0.5 to 0.6) | 3.8 | (3.4 to 4.1) |
| Rhinosinusitis | 4.2 | (3.9 to 4.6) | 4.0 | (3.6 to 4.3) | 0.3 | (0.3 to 0.3) | 6.7 | (5.9 to 7.5) |
| Unspecified URTI | 72.2 | (70.5 to 74.0) | 70.8 | (69.1 to 72.5) | 1.4 | (1.2 to 1.6) | 2.0 | (1.7 to 2.3) |
| All LRTI | 20.0 | (19.0 to 21.0) | 19.4 | (18.4 to 20.4) | 0.6 | (0.5 to 0.7) | 3.0 | (2.7 to 3.4) |
| Bronchitis | 0.7 | (0.6 to 0.9) | 0.7 | (0.5 to 0.8) | 0.0 | (0.0 to 0.0) | 4.2 | (3.2 to 5.5) |
| Unspecified LRTI | 20.0 | (19.1 to 21.0) | 19.4 | (18.5 to 20.4) | 0.6 | (0.5 to 0.7) | 3.0 | (2.7 to 3.4) |

Panel (b) Adults

| | **Δ Observed (95% CI)** | | **Δ Counterfactual (95% CI)** | | **Absolute contribution (95% CI)** | | **% Relative contribution (95% CI)** | |
|---|---|---|---|---|---|---|---|---|
| | | | | | **Influenza vaccination** | | | |
| Asthma exacerbation | 1.8 | (1.7 to 1.9) | 1.8 | (1.7 to 2.0) | -0.1 | (-0.1 to 0.0) | -2.9 | (-3.2 to -2.6) |
| COPD exacerbation | 5.8 | (5.6 to 6.1) | 5.7 | (5.4 to 5.9) | 0.2 | (0.1 to 0.2) | 2.7 | (2.5 to 3.0) |
| Cough | 0.1 | (-0.4 to 0.6) | -0.4 | (-0.9 to 0.1) | 0.5 | (0.5 to 0.5) | 167.8 | (-2278.0 to 2787.8) |
| Sore throat | 1.4 | (1.1 to 1.7) | 1.4 | (1.1 to 1.7) | 0.1 | (0.1 to 0.1) | 4.2 | (3.4 to 5.3) |
| Unspecified URTI | 5.5 | (5.2 to 5.9) | 5.4 | (5.1 to 5.7) | 0.1 | (0.1 to 0.1) | 2.1 | (1.9 to 2.3) |
| All LRTI | 9.2 | (8.7 to 9.6) | 8.6 | (8.1 to 9.0) | 0.6 | (0.6 to 0.6) | 6.4 | (5.9 to 6.9) |
| Unspecified LRTI | 8.3 | (7.9 to 8.8) | 7.9 | (7.4 to 8.3) | 0.5 | (0.4 to 0.5) | 5.6 | (5.1 to 6.0) |
| | | | | | **Pneumococcal vaccination** | | | |
| Asthma exacerbation | 1.7 | (1.5 to 1.8) | 1.7 | (1.5 to 1.8) | 0.0 | (0.0 to 0.0) | 1.3 | (1.1 to 1.5) |

| | | | | | | | | |
|---|---|---|---|---|---|---|---|---|
| COPD exacerbation | 6.1 | (5.8 to 6.3) | 6.0 | (5.8 to 6.3) | 0.0 | (0.0 to 0.0) | 0.4 | (0.3 to 0.5) |
| Cough | 0.3 | (-0.2 to 0.9) | 0.3 | (-0.3 to 0.8) | 0.1 | (0.0 to 0.1) | -19.6 | (-226.7 to 214.6) |
| Sore throat | 1.4 | (1.2 to 1.8) | 1.4 | (1.2 to 1.8) | 0.0 | (0.0 to 0.0) | 0.0 | (-0.2 to 0.1) |
| Unspecified URTI | 5.6 | (5.2 to 5.9) | 5.6 | (5.3 to 5.9) | 0.0 | (0.0 to 0.0) | -0.4 | (-0.5 to -0.4) |
| All LRTI | 9.1 | (8.6 to 9.6) | 9.0 | (8.5 to 9.5) | 0.1 | (0.1 to 0.2) | 1.6 | (1.1 to 2.1) |
| Unspecified LRTI | 8.3 | (7.9 to 8.8) | 8.2 | (7.8 to 8.7) | 0.1 | (0.1 to 0.1) | 1.4 | (0.9 to 1.8) |
| | | | | **Smoking status** | | | | |
| Asthma exacerbation | 1.8 | (1.6 to 1.9) | 0.7 | (0.5 to 0.8) | 1.1 | (1.1 to 1.1) | 62.6 | (58.5 to 67.4) |
| COPD exacerbation | 7.2 | (6.9 to 7.5) | 1.9 | (1.7 to 2.1) | 5.3 | (5.1 to 5.4) | 73.2 | (71.5 to 75.0) |
| Cough | -0.1 | (-0.6 to 0.4) | -8.3 | (-8.8 to -7.7) | 8.2 | (8.1 to 8.3) | 3605.5 | (-31940.0 to 62234.3) |
| Sore throat | 1.5 | (1.2 to 1.8) | 0.5 | (0.2 to 0.8) | 1.0 | (0.9 to 1.0) | 65.0 | (53.5 to 80.3) |
| Unspecified URTI | 5.7 | (5.3 to 6.0) | 4.0 | (3.7 to 4.3) | 1.6 | (1.6 to 1.7) | 28.7 | (27.1 to 30.7) |
| All LRTI | 9.4 | (8.9 to 9.9) | 0.6 | (0.2 to 1.1) | 8.7 | (8.6 to 8.9) | 93.5 | (88.8 to 98.0) |
| Unspecified LRTI | 8.6 | (8.2 to 9.0) | 0.6 | (0.2 to 1.1) | 7.9 | (7.8 to 8.1) | 92.7 | (88.0 to 97.1) |
| Otitis externa | 0.3 | (0.1 to 0.5) | -0.4 | (-0.6 to -0.2) | 0.7 | (0.6 to 0.7) | 271.7 | (133.2 to 659.0) |
| | | | | **Body mass index** | | | | |
| Asthma exacerbation | 1.7 | (1.6 to 1.9) | 1.2 | (1.0 to 1.3) | 1.2 | (1.0 to 1.3) | 31.8 | (26.0 to 35.1) |
| COPD exacerbation | 6.6 | (6.2 to 7.2) | 5.8 | (5.5 to 6.1) | 5.8 | (5.5 to 6.1) | 12.8 | (10.4 to 18.6) |
| Cough | 0.3 | (-0.3 to 0.9) | -0.1 | (-0.6 to 0.5) | -0.1 | (-0.6 to 0.5) | 13.0 | (-1163.9 to 1296.4) |
| Sore throat | 1.4 | (1.1 to 1.7) | 2.1 | (1.8 to 2.4) | 2.1 | (1.8 to 2.4) | -52.2 | (-67.0 to -31.8) |
| Unspecified URTI | 5.5 | (5.2 to 5.9) | 6.1 | (5.7 to 6.4) | 6.1 | (5.7 to 6.4) | -10.2 | (-11.8 to -5.6) |
| All LRTI | 9.7 | (9.2 to 10.2) | 8.3 | (7.8 to 8.8) | 8.3 | (7.8 to 8.8) | 15.0 | (13.3 to 16.4) |
| Unspecified LRTI | 8.8 | (8.3 to 9.3) | 7.5 | (7.1 to 8.0) | 1.2 | (1.1 to 1.3) | 14.1 | (12.8 to 15.2) |
| Otitis externa | 0.2 | (0.0 to 0.4) | 0.4 | (0.2 to 0.6) | -0.1 | (-0.2 to 0.0) | -49.4 | (-203.4 to -11.5) |

Notes: Δ represents the absolute difference in mean consultation rates (consultations per 1000 person-years) between the most deprived (IMD=20) and least deprived (IMD=1, reference), adjusted for age, sex, ethnicity, and the contributor of interest. Δ observed uses the actual contributor distributions, and Δ counterfactual replaces the contributor distribution of the most deprived with that of the least deprived. Absolute contribution is calculated as the difference between Δ observed and Δ counterfactual. Relative contribution is calculated as the absolute contribution divided by Δ observed. Positive values indicate the contributor reduces the deprivation gap, and negative values indicate that the contributor increases the gap. Abbreviations: IMD index of multiple deprivation, URTI upper respiratory tract infection, LRTI lower respiratory tract infection, UTI urinary tract infection, COPD chronic obstructive pulmonary disease, CI confidence interval. "All URTI" includes sore throat, unspecified upper respiratory tract infection, rhinosinusitis whereas "All LRTI" includes bronchitis, unspecified lower respiratory tract infection, pneumonia.

#### S4.2 Sensitivity analysis using observed population

##### S4.2.1 Descriptive results

*Table S6 Characteristics of observed population by deprivation quintile. Data are median (interquartile range) for continuous variables and % for categorical variables*

| Characteristic | Q1 (least deprived) | Q2 | Q3 | Q4 | Q5 (most deprived) |
|---|---|---|---|---|---|
| **Children** | | | | | |
| Age (years) | 7.0 (4.0-12.0) | 7.0 (3.0-12.0) | 7.0 (3.0-11.0) | 7.0 (3.0-11.0) | 7.0 (3.0-11.0) |
| Age group | | | | | |
| 0-4 | 32.3 | 33.7 | 34.2 | 34.3 | 34.0 |
| 5-11 | 26.7 | 25.9 | 24.9 | 23.9 | 24.7 |
| 12-17 | 41.1 | 40.4 | 40.9 | 41.8 | 41.3 |
| Female | 50.3 | 50.1 | 50.2 | 49.7 | 49.9 |
| Ethnic minority group | 13.1 | 14.9 | 22.2 | 32.5 | 33.2 |
| Influenza vaccination | 35.5 | 32.5 | 28.4 | 24.4 | 23.4 |

| | | | | | |
|---|---|---|---|---|---|
| Pneumococcal vaccination | 81.3 | 81.3 | 80.7 | 80.7 | 79.5 |
| N | 433,721 | 418,998 | 422,143 | 485,314 | 563,279 |
| **Adults** | | | | | |
| Age (years) | 52.0 (35.0-70.0) | 51.0 (33.0-69.0) | 49.0 (32.0-66.0) | 46.0 (31.0-63.0) | 45.0 (30.0-61.0) |
| Age group | | | | | |
| 18-64 | 68.1 | 69.8 | 72.7 | 76.5 | 78.7 |
| 65 and above | 31.9 | 30.2 | 27.3 | 23.5 | 21.3 |
| Female | 60.2 | 60.3 | 60.5 | 60.4 | 60.2 |
| Ethnic minority group | 6.7 | 8.4 | 12.7 | 19.2 | 20.6 |
| Influenza vaccination | 30.8 | 29.2 | 27.3 | 24.7 | 23.2 |
| Pneumococcal vaccination | 26.5 | 25.4 | 23.7 | 21.6 | 20.7 |
| Smoking status | | | | | |
| Non-smoker | 69.5 | 64.6 | 60.6 | 55.2 | 48.8 |
| Ex-smoker | 14.0 | 15.2 | 15.5 | 15.9 | 15.6 |
| Current smoker | 16.5 | 20.2 | 23.9 | 28.9 | 35.7 |
| Body mass index | 25.6 (22.8-29.1) | 25.9 (23.0-29.5) | 26.1 (23.1-29.8) | 26.3 (23.1-30.3) | 26.7 (23.4-31.1) |
| N | 1,444,387 | 1,458,853 | 1,385,394 | 1,431,266 | 1,383,480 |

*Table S7 Antibiotic prescribing rates (number of prescriptions per 1000 person-years) in children (<18 years) and adults (≥18 years) in the observed population*

| | Prescribing rate | (95% CI) |
|---|---|---|
| **Children** | | |
| All URTI | 53.7 | (53.4 to 54.0) |
| UTI | 40.5 | (40.2 to 40.9) |
| Otitis media | 35.0 | (34.8 to 35.2) |
| All LRTI | 33.0 | (32.7 to 33.2) |
| Unspecified LRTI | 32.8 | (32.6 to 33.0) |
| Cough | 30.9 | (30.7 to 31.1) |
| Sore throat | 27.8 | (27.6 to 28.0) |
| Unspecified URTI | 25.0 | (24.8 to 25.2) |
| Otitis externa | 9.7 | (9.5 to 9.8) |
| Impetigo | 6.0 | (5.9 to 6.1) |
| Rhinosinusitis | 2.8 | (2.7 to 2.9) |
| Cellulitis | 1.7 | (1.6 to 1.8) |
| Asthma exacerbation | 1.2 | (1.1 to 1.2) |
| Gastroenteritis | 1.2 | (1.1 to 1.2) |
| Bronchitis | 0.8 | (0.8 to 0.9) |
| Pneumonia | 0.2 | (0.1 to 0.2) |
| **Adults** | | |
| UTI | 132.1 | (131.8 to 132.4) |
| All LRTI | 61.4 | (61.3 to 61.6) |
| Unspecified LRTI | 60.4 | (60.3 to 60.6) |
| Cough | 48.5 | (48.3 to 48.7) |
| All URTI | 47.7 | (47.5 to 47.8) |
| Rhinosinusitis | 18.0 | (17.9 to 18.1) |
| Sore throat | 17.5 | (17.4 to 17.6) |
| Unspecified URTI | 13.4 | (13.3 to 13.4) |

| | | |
|---|---|---|
| Cellulitis | 13.3 | (13.2 to 13.3) |
| COPD exacerbation | 7.2 | (7.2 to 7.3) |
| Otitis externa | 4.8 | (4.8 to 4.9) |
| Asthma exacerbation | 4.1 | (4.0 to 4.1) |
| Bronchitis | 3.4 | (3.4 to 3.4) |
| Gastroenteritis | 2.9 | (2.8 to 2.9) |
| Pyelonephritis | 2.3 | (2.3 to 2.3) |
| Prostatitis | 1.7 | (1.6 to 1.7) |
| Impetigo | 1.3 | (1.2 to 1.3) |
| Pneumonia | 1.1 | (1.1 to 1.1) |

Abbreviations: IMD index of multiple deprivation, URTI upper respiratory tract infections, LRTI lower respiratory tract infections, UTI urinary tract infection, COPD chronic obstructive pulmonary disease, CI confidence interval. "All URTI" includes sore throat, unspecified upper respiratory tract infection, rhinosinusitis whereas "All LRTI" includes bronchitis, unspecified lower respiratory tract infection, pneumonia.

##### S4.2.2 Association between deprivation and indication-related outcomes

*Table S8 Mean prescribing rates and differences between the most deprived (IMD=20) and least deprived (IMD=1, reference)*

| | | | Children | | Adults | |
|---|---|---|---|---|---|---|
| | Indication | Deprivation level/Δ gap | Mean | (95% CI) | Mean | (95% CI) |
| Respiratory | Asthma exacerbation | IMD=20 | 1.6 | (1.4 to 1.8) | 6.1 | (5.9 to 6.4) |
| | | IMD=1 | 1.0 | (0.9 to 1.2) | 3.4 | (3.3 to 3.6) |
| | | Δ Absolute | 0.6 | (0.4 to 0.7) | 2.7 | (2.5 to 2.9) |
| | | Δ Percentage (%) | 54.9 | (35.1 to 77.2) | 79.3 | (72.2 to 86.8) |
| | COPD exacerbation | IMD=20 | - | - | 9.1 | (8.6 to 9.5) |
| | | IMD=1 | - | - | 2.1 | (2.0 to 2.3) |
| | | Δ Absolute | - | - | 6.9 | (6.6 to 7.3) |
| | | Δ Percentage (%) | - | - | 323.7 | (305.9 to 342.2) |
| | Cough | IMD=20 | 21.4 | (20.8 to 21.9) | 54.8 | (54.1 to 55.5) |
| | | IMD=1 | 19.7 | (19.1 to 20.2) | 41.4 | (40.9 to 41.9) |
| | | Δ Absolute | 1.7 | (1.3 to 2.2) | 13.4 | (12.8 to 14.0) |
| | | Δ Percentage (%) | 8.8 | (6.3 to 11.5) | 32.4 | (30.8 to 34.0) |
| | All URTI | IMD=20 | 40.0 | (39.2 to 40.8) | 42.0 | (41.5 to 42.6) |
| | | IMD=1 | 33.9 | (33.2 to 34.6) | 42.5 | (42.0 to 43.0) |
| | | Δ Absolute | 6.1 | (5.4 to 6.8) | -0.5 | (-1.0 to 0.0) |
| | | Δ Percentage (%) | 18.0 | (15.8 to 20.2) | -1.2 | (-2.3 to 0.0) |
| | Sore throat | IMD=20 | 27.0 | (26.3 to 27.8) | 13.3 | (13.0 to 13.6) |
| | | IMD=1 | 23.1 | (22.5 to 23.8) | 13.2 | (13.0 to 13.5) |
| | | Δ Absolute | 3.9 | (3.2 to 4.5) | 0.1 | (-0.2 to 0.3) |
| | | Δ Percentage (%) | 16.7 | (13.6 to 19.9) | 0.4 | (-1.6 to 2.5) |
| | Rhinosinusitis | IMD=20 | 1.3 | (1.2 to 1.4) | 15.8 | (15.5 to 16.2) |
| | | IMD=1 | 1.3 | (1.2 to 1.5) | 18.8 | (18.4 to 19.2) |
| | | Δ Absolute | -0.1 | (-0.2 to 0.1) | -3.0 | (-3.4 to -2.7) |
| | | Δ Percentage (%) | -3.9 | (-12.1 to 5.3) | -16.0 | (-17.7 to -14.3) |
| | Unspecified URTI | IMD=20 | 13.4 | (13.0 to 13.8) | 12.7 | (12.4 to 13.0) |
| | | IMD=1 | 10.7 | (10.3 to 11.0) | 10.2 | (10.0 to 10.4) |

| | | | | | | |
|---|---|---|---|---|---|---|
| | | Δ Absolute | 2.7 | (2.4 to 3.1) | 2.6 | (2.3 to 2.8) |
| | | Δ Percentage (%) | 25.4 | (22.0 to 29.1) | 25.1 | (22.4 to 28.0) |
| | All LRTI | IMD=20 | 25.5 | (24.9 to 26.1) | 83.6 | (82.7 to 84.5) |
| | | IMD=1 | 20.5 | (19.9 to 21.0) | 52.6 | (52.1 to 53.2) |
| | | Δ Absolute | 5.0 | (4.5 to 5.6) | 31.0 | (30.2 to 31.8) |
| | | Δ Percentage (%) | 24.6 | (21.7 to 27.7) | 59.0 | (57.1 to 60.7) |
| | Bronchitis | IMD=20 | 0.6 | (0.5 to 0.7) | 4.3 | (4.1 to 4.5) |
| | | IMD=1 | 0.4 | (0.3 to 0.5) | 3.4 | (3.3 to 3.6) |
| | | Δ Absolute | 0.2 | (0.1 to 0.2) | 0.9 | (0.8 to 1.1) |
| | | Δ Percentage (%) | 39.4 | (20.4 to 61.1) | 26.7 | (21.6 to 32.1) |
| | Unspecified LRTI | IMD=20 | 25.3 | (24.7 to 26.0) | 82.3 | (81.4 to 83.2) |
| | | IMD=1 | 20.3 | (19.8 to 20.9) | 51.8 | (51.2 to 52.3) |
| | | Δ Absolute | 5.0 | (4.5 to 5.6) | 30.6 | (29.8 to 31.4) |
| | | Δ Percentage (%) | 24.7 | (21.9 to 27.8) | 59.1 | (57.3 to 60.7) |
| | Pneumonia | IMD=20 | 0.1 | (0.1 to 0.2) | 1.4 | (1.3 to 1.5) |
| | | IMD=1 | 0.2 | (0.1 to 0.3) | 1.0 | (0.9 to 1.1) |
| | | Δ Absolute | 0.0 | (-0.1 to 0.0) | 0.4 | (0.3 to 0.5) |
| | | Δ Percentage (%) | -18.6 | (-45.6 to 17.7) | 40.9 | (30.1 to 52.0) |
| Skin | Cellulitis | IMD=20 | 1.4 | (1.2 to 1.5) | 14.1 | (13.8 to 14.4) |
| | | IMD=1 | 1.7 | (1.5 to 1.9) | 9.9 | (9.6 to 10.1) |
| | | Δ Absolute | -0.3 | (-0.5 to -0.2) | 4.2 | (3.9 to 4.5) |
| | | Δ Percentage (%) | -20.2 | (-28.2 to -12.1) | 43.0 | (39.5 to 46.3) |
| | Impetigo | IMD=20 | 7.3 | (6.9 to 7.7) | 1.3 | (1.2 to 1.4) |
| | | IMD=1 | 8.1 | (7.6 to 8.6) | 1.3 | (1.2 to 1.4) |
| | | Δ Absolute | -0.8 | (-1.2 to -0.4) | -0.1 | (-0.2 to 0.1) |
| | | Δ Percentage (%) | -9.6 | (-14.2 to -4.8) | -3.4 | (-10.6 to 4.3) |
| Ear | Otitis externa | IMD=20 | 10.4 | (9.9 to 10.8) | 6.0 | (5.8 to 6.2) |
| | | IMD=1 | 11.3 | (10.8 to 11.9) | 4.8 | (4.6 to 5.0) |
| | | Δ Absolute | -1.0 | (-1.5 to -0.5) | 1.2 | (1.0 to 1.4) |
| | | Δ Percentage (%) | -8.7 | (-12.7 to -4.6) | 25.5 | (20.4 to 30.1) |
| | Otitis media | IMD=20 | 26.9 | (26.2 to 27.5) | - | - |
| | | IMD=1 | 31.5 | (30.7 to 32.3) | - | - |
| | | Δ Absolute | -4.6 | (-5.3 to -3.9) | - | - |
| | | Δ Percentage (%) | -14.7 | (-16.6 to -12.6) | - | - |
| Gastrointestinal | Gastroenteritis | IMD=20 | 0.7 | (0.6 to 0.8) | 3.0 | (2.9 to 3.2) |
| | | IMD=1 | 0.6 | (0.5 to 0.7) | 2.8 | (2.7 to 2.9) |
| | | Δ Absolute | 0.2 | (0.1 to 0.2) | 0.2 | (0.1 to 0.4) |
| | | Δ Percentage (%) | 26.5 | (11.2 to 43.6) | 8.0 | (3.0 to 13.5) |
| Urinary | UTI | IMD=20 | 57.4 | (55.5 to 59.3) | 112.5 | (111.3 to 113.7) |
| | | IMD=1 | 54.5 | (52.7 to 56.3) | 113.5 | (112.3 to 114.7) |
| | | Δ Absolute | 2.9 | (1.0 to 5.0) | -1.0 | (-2.3 to 0.3) |
| | | Δ Percentage (%) | 5.4 | (1.9 to 9.3) | -0.9 | (-2.0 to 0.2) |
| | Pyelonephritis | IMD=20 | - | - | 2.6 | (2.4 to 2.8) |
| | | IMD=1 | - | - | 2.5 | (2.4 to 2.7) |
| | | Δ Absolute | - | - | 0.0 | (-0.1 to 0.2) |
| | | Δ Percentage (%) | - | - | 1.8 | (-4.8 to 9.0) |
| | Prostatitis | IMD=20 | - | - | 1.8 | (1.6 to 2.1) |
| | | IMD=1 | - | - | 3.2 | (2.9 to 3.5) |

| | | | | |
|---|---|---|---|---|
| Δ Absolute | - | - | -1.4 | (-1.7 to -1.1) |
| Δ Percentage (%) | - | - | -42.8 | (-49.1 to -35.9) |

Abbreviations: IMD index of multiple deprivation, URTI upper respiratory tract infections, LRTI lower respiratory tract infections, UTI urinary tract infection, COPD chronic obstructive pulmonary disease, CI confidence interval. “All URTI” includes sore throat, unspecified upper respiratory tract infection, rhinosinusitis whereas “All LRTI” includes bronchitis, unspecified lower respiratory tract infection, pneumonia.

*Table S9 Absolute and relative contribution of modifiable health factors to consultation rate differences between the most and least deprived*

Panel (a) Children

| | **Δ Observed (95% CI)** | | **Δ Counterfactual (95% CI)** | | **Absolute contribution (95% CI)** | | **% Relative contribution (95% CI)** | |
|---|---|---|---|---|---|---|---|---|
| | | | | **Influenza vaccination** | | | | |
| Asthma exacerbation | 1.8 | (1.4 to 2.2) | 2.3 | (1.9 to 2.6) | -0.5 | (-0.5 to -0.4) | **-26.2** | (-31.9 to -21.9) |
| Cough | 11.4 | (9.8 to 13.1) | 8.0 | (6.4 to 9.7) | 3.4 | (3.3 to 3.6) | **30.1** | (25.9 to 35.2) |
| All URTI | 78.8 | (76.3 to 81.4) | 65.2 | (62.7 to 67.7) | 13.7 | (13.3 to 14.0) | **17.3** | (16.7 to 18.0) |
| Sore throat | 11.4 | (10.1 to 12.6) | 8.8 | (7.6 to 10.0) | 2.5 | (2.4 to 2.7) | **22.4** | (20.2 to 25.0) |
| Unspecified URTI | 70.6 | (68.4 to 72.9) | 58.7 | (56.5 to 60.9) | 11.9 | (11.6 to 12.2) | **16.9** | (16.2 to 17.5) |
| All LRTI | 21.0 | (19.7 to 22.3) | 18.8 | (17.6 to 20.0) | 2.2 | (2.0 to 2.3) | **10.4** | (9.6 to 11.3) |
| Bronchitis | 0.8 | (0.5 to 1.0) | 0.7 | (0.5 to 0.9) | 0.1 | (0.1 to 0.1) | **10.0** | (6.6 to 14.3) |
| Unspecified LRTI | 21.1 | (19.9 to 22.4) | 18.9 | (17.7 to 20.2) | 2.2 | (2.0 to 2.3) | **10.3** | (9.5 to 11.2) |
| | | | | **Pneumococcal vaccination** | | | | |
| Asthma exacerbation | 1.8 | (1.5 to 2.1) | 1.6 | (1.3 to 1.9) | 0.2 | (0.2 to 0.2) | **12.3** | (10.3 to 14.8) |
| Cough | 12.9 | (11.1 to 14.8) | 9.4 | (7.6 to 11.2) | 3.5 | (3.3 to 3.7) | **27.3** | (23.6 to 31.5) |
| All URTI | 82.8 | (80.3 to 85.6) | 73.4 | (70.8 to 76.1) | 9.5 | (8.9 to 10.1) | **11.4** | (10.7 to 12.2) |
| Sore throat | 12.4 | (11.1 to 13.6) | 9.6 | (8.4 to 10.9) | 2.7 | (2.6 to 2.9) | **22.2** | (20.0 to 24.7) |
| Unspecified URTI | 74.1 | (71.8 to 76.4) | 68.0 | (65.7 to 70.3) | 6.1 | (5.7 to 6.5) | **8.2** | (7.6 to 8.8) |
| All LRTI | 22.2 | (20.9 to 23.5) | 19.9 | (18.7 to 21.2) | 2.2 | (2.1 to 2.4) | **10.2** | (9.3 to 11.1) |
| Bronchitis | 0.8 | (0.6 to 1.0) | 0.7 | (0.5 to 0.9) | 0.1 | (0.1 to 0.1) | **12.4** | (9.0 to 16.9) |
| Unspecified LRTI | 22.3 | (21.1 to 23.7) | 20.1 | (18.9 to 21.4) | 2.2 | (2.1 to 2.4) | **10.0** | (9.2 to 10.9) |

Panel (b) Adults

| | **Δ Observed (95% CI)** | | **Δ Counterfactual (95% CI)** | | **Absolute contribution (95% CI)** | | **% Relative contribution (95% CI)** | |
|---|---|---|---|---|---|---|---|---|
| | | | | **Influenza vaccination** | | | | |
| Asthma exacerbation | 4.6 | (4.3 to 4.9) | 4.6 | (4.3 to 4.9) | 0.0 | (-0.1 to 0.0) | **-1.0** | (-1.3 to -0.8) |
| COPD exacerbation | 15.4 | (14.8 to 16.0) | 14.9 | (14.4 to 15.5) | 0.5 | (0.4 to 0.5) | **3.2** | (2.9 to 3.4) |
| Cough | 15.9 | (15.0 to 16.9) | 14.8 | (13.8 to 15.8) | 1.1 | (1.1 to 1.2) | **7.2** | (6.6 to 7.8) |
| Unspecified URTI | 15.8 | (15.3 to 16.5) | 15.6 | (15.0 to 16.3) | 0.2 | (0.2 to 0.2) | **1.3** | (1.2 to 1.5) |
| All LRTI | 30.7 | (29.8 to 31.7) | 29.2 | (28.3 to 30.1) | 1.6 | (1.5 to 1.6) | **5.1** | (4.8 to 5.4) |
| Bronchitis | 0.6 | (0.4 to 0.8) | 0.5 | (0.4 to 0.7) | 0.0 | (0.0 to 0.1) | **8.3** | (6.1 to 11.6) |
| Unspecified LRTI | 28.0 | (27.1 to 28.9) | 26.7 | (25.9 to 27.6) | 1.2 | (1.2 to 1.3) | **4.3** | (4.1 to 4.6) |
| Pneumonia | 2.9 | (2.6 to 3.3) | 2.4 | (2.1 to 2.7) | 0.5 | (0.5 to 0.5) | **17.2** | (15.6 to 19.0) |
| | | | | **Pneumococcal vaccination** | | | | |
| Asthma exacerbation | 4.5 | (4.2 to 4.7) | 4.4 | (4.1 to 4.7) | 0.0 | (0.0 to 0.0) | **0.9** | (0.6 to 1.1) |
| COPD exacerbation | 15.5 | (14.9 to 16.0) | 15.5 | (14.9 to 16.0) | 0.0 | (0.0 to 0.0) | **0.2** | (0.1 to 0.3) |
| Cough | 16.2 | (15.2 to 17.2) | 17.1 | (16.0 to 18.0) | -0.8 | (-0.9 to -0.7) | **-5.0** | (-5.7 to -4.4) |
| Unspecified URTI | 15.9 | (15.3 to 16.5) | 16.3 | (15.7 to 16.9) | -0.5 | (-0.5 to -0.4) | **-2.9** | (-3.1 to -2.7) |

| | | | | | | | | |
|---|---|---|---|---|---|---|---|---|
| All LRTI | 30.7 | (29.8 to 31.6) | 31.0 | (30.1 to 32.0) | -0.3 | (-0.4 to -0.2) | **-1.1** | (-1.5 to -0.7) |
| Bronchitis | 0.6 | (0.4 to 0.8) | 0.6 | (0.4 to 0.8) | 0.0 | (0.0 to 0.0) | **-6.4** | (-9.2 to -4.8) |
| Unspecified LRTI | 27.9 | (27.1 to 28.7) | 28.3 | (27.4 to 29.1) | -0.4 | (-0.4 to -0.3) | **-1.3** | (-1.6 to -0.9) |
| Pneumonia | 3.0 | (2.7 to 3.3) | 2.7 | (2.4 to 3.0) | 0.2 | (0.2 to 0.3) | **7.5** | (6.3 to 8.9) |
| **Smoking status** | | | | | | | | |
| Asthma exacerbation | 4.6 | (4.3 to 4.9) | 2.7 | (2.5 to 3.0) | 1.9 | (1.8 to 2.0) | **41.1** | (38.9 to 43.5) |
| COPD exacerbation | 18.1 | (17.5 to 18.7) | 5.3 | (4.9 to 5.6) | 12.8 | (12.5 to 13.1) | **70.9** | (69.7 to 72.1) |
| Cough | 16.5 | (15.5 to 17.5) | 4.7 | (3.7 to 5.6) | 11.8 | (11.6 to 12.1) | **71.8** | (67.6 to 76.3) |
| Unspecified URTI | 15.9 | (15.3 to 16.5) | 15.7 | (15.1 to 16.3) | 0.2 | (0.0 to 0.3) | **1.0** | (0.2 to 1.7) |
| All LRTI | 31.9 | (30.9 to 32.8) | 16.6 | (15.7 to 17.5) | 15.3 | (15.1 to 15.5) | **48.0** | (46.7 to 49.4) |
| Bronchitis | 0.6 | (0.5 to 0.8) | 0.0 | (-0.2 to 0.2) | 0.6 | (0.6 to 0.7) | **99.6** | (76.2 to 133.0) |
| Unspecified LRTI | 29.1 | (28.2 to 30.0) | 15.2 | (14.4 to 16.1) | 13.8 | (13.6 to 14.1) | **47.6** | (46.2 to 49.0) |
| Pneumonia | 3.1 | (2.8 to 3.4) | 1.5 | (1.2 to 1.7) | 1.6 | (1.5 to 1.7) | **52.8** | (48.1 to 58.1) |
| Cellulitis | 3.5 | (3.0 to 3.9) | 2.1 | (1.6 to 2.5) | 1.4 | (1.3 to 1.5) | **40.7** | (35.4 to 46.6) |
| Otitis externa | 2.9 | (2.5 to 3.3) | 2.6 | (2.2 to 3.0) | 0.3 | (0.2 to 0.4) | **10.5** | (7.3 to 13.6) |
| Gastroenteritis | 4.3 | (3.8 to 4.9) | 3.2 | (2.7 to 3.8) | 1.1 | (1.0 to 1.2) | **25.2** | (21.5 to 29.4) |
| **Body mass index** | | | | | | | | |
| Asthma exacerbation | 4.6 | (4.3 to 4.9) | 3.2 | (2.9 to 3.5) | 1.4 | (1.3 to 1.4) | **29.6** | (27.5 to 31.5) |
| COPD exacerbation | 16.1 | (15.6 to 16.7) | 14.1 | (13.6 to 14.7) | 2.0 | (1.9 to 2.2) | **12.4** | (11.6 to 13.2) |
| Cough | 15.9 | (14.8 to 16.8) | 12.2 | (11.2 to 13.3) | 3.7 | (3.4 to 3.9) | **23.2** | (21.3 to 25.1) |
| Unspecified URTI | 15.9 | (15.3 to 16.5) | 15.0 | (14.4 to 15.6) | 0.9 | (0.8 to 1.0) | **5.7** | (5.1 to 6.3) |
| All LRTI | 31.1 | (30.1 to 32.0) | 25.9 | (25.0 to 27.0) | 5.2 | (4.9 to 5.4) | **16.6** | (15.6 to 17.4) |
| Bronchitis | 0.6 | (0.4 to 0.8) | 0.3 | (0.2 to 0.5) | 0.2 | (0.2 to 0.3) | **42.5** | (30.7 to 58.8) |
| Unspecified LRTI | 28.3 | (27.4 to 29.2) | 23.6 | (22.8 to 24.7) | 4.6 | (4.4 to 4.8) | **16.4** | (15.4 to 17.1) |
| Pneumonia | 3.0 | (2.7 to 3.3) | 2.3 | (2.0 to 2.6) | 0.7 | (0.6 to 0.7) | **23.3** | (20.6 to 26.0) |
| Cellulitis | 3.7 | (3.3 to 4.2) | 0.3 | (-0.2 to 0.8) | 3.5 | (3.3 to 3.6) | **93.6** | (81.7 to 106.0) |
| Otitis externa | 2.9 | (2.5 to 3.2) | 2.5 | (2.2 to 2.9) | 0.3 | (0.3 to 0.4) | **11.6** | (9.2 to 13.9) |
| Gastroenteritis | 4.3 | (3.7 to 4.9) | 3.4 | (2.8 to 3.9) | 0.9 | (0.8 to 1.0) | **21.2** | (17.8 to 24.8) |

Abbreviations: URTI upper respiratory tract infections, LRTI lower respiratory tract infections, CI confidence interval. “All URTI” includes sore throat, unspecified upper respiratory tract infection, rhinosinusitis whereas “All LRTI” includes bronchitis, unspecified lower respiratory tract infection, pneumonia.

#### S4.3 Sensitivity analysis using complete case data in the observed population

##### S4.3.1 Descriptive statistics

*Table S10 Summary statistics of covariates with missing data in the complete case and imputed dataset in the observed population*

| | Complete case | Imputed data |
|---|---|---|
| **Children** | **N=1,736,753** | **N=2,323,455** |
| IMD - Median (interquartile range) | 12 (6-16) | 11 (6-16) |
| *Missing - n (%)* | *540,486 (23.3)* | |
| Ethnicity - % | | |
| White | 74.8 | 76.0 |
| Ethnic minority group | 25.2 | 24.0 |
| *Missing - n (%)* | *113,565 (4.9)* | |
| **Adults** | **N=4,972,709** | **N=7,103,380** |

| | | |
|---|---|---|
| IMD - Median (interquartile range) | 10 (5-15) | 10 (5-15) |
| *Missing - n (%)* | *1,604,482 (22.6)* | |
| Ethnicity - % | | |
| White | 86.3 | 86.6 |
| Ethnic minority group | 13.7 | 13.4 |
| *Missing - n (%)* | *217,705 (3.1)* | |
| Smoking status | | |
| Non-smoker | 58.2 | 59.8 |
| Ex-smoker | 16.5 | 15.2 |
| Current smoker | 25.3 | 24.9 |
| *Missing - n (%)* | *2,363(0.04)* | |
| BMI - Median (interquartile range) | 26.2 (22.9-30.4) | 26.1 (23.1-29.9) |
| *Missing - n (%)* | *599,863 (8.4)* | |

Abbreviations: IMD index of multiple deprivation, BMI body mass index.

##### S4.3.2 Association between IMD and antibiotic prescribing rates

*Table S11 Mean prescribing rates and differences between the most deprived (IMD=20) and least deprived (IMD=1, reference) – complete case vs imputed data – children*

| | | | **Complete case** N=1,736,753 | | **Imputed data** N=2,323,455 | |
|---|---|---|---|---|---|---|
| | Indication | Deprivation level/Δ gap | Mean | (95% CI) | Mean | (95% CI) |
| Respiratory | Asthma exacerbation | IMD=20 | 1.7 | (1.4 to 1.9) | 1.6 | (1.4 to 1.8) |
| | | IMD=1 | 1.0 | (0.8 to 1.1) | 1.0 | (0.9 to 1.2) |
| | | Δ Absolute | 0.7 | (0.5 to 0.9) | 0.6 | (0.4 to 0.7) |
| | | Δ Percentage (%) | 74.8 | (46.5 to 104.5) | 54.9 | (35.1 to 77.2) |
| | Cough | IMD=20 | 19.6 | (19.0 to 20.2) | 21.4 | (20.8 to 21.9) |
| | | IMD=1 | 17.5 | (16.9 to 18.1) | 19.7 | (19.1 to 20.2) |
| | | Δ Absolute | 2.1 | (1.5 to 2.6) | 1.7 | (1.3 to 2.2) |
| | | Δ Percentage (%) | 12.0 | (8.2 to 15.1) | 8.8 | (6.3 to 11.5) |
| | All URTI | IMD=20 | 37.1 | (36.2 to 37.9) | 40.0 | (39.2 to 40.8) |
| | | IMD=1 | 29.5 | (28.8 to 30.3) | 33.9 | (33.2 to 34.6) |
| | | Δ Absolute | 7.5 | (6.7 to 8.3) | 6.1 | (5.4 to 6.8) |
| | | Δ Percentage (%) | 25.5 | (22.4 to 28.5) | 18.0 | (15.8 to 20.2) |
| | Sore throat | IMD=20 | 25.3 | (24.5 to 26.1) | 27.0 | (26.3 to 27.8) |
| | | IMD=1 | 20.5 | (19.8 to 21.2) | 23.1 | (22.5 to 23.8) |
| | | Δ Absolute | 4.8 | (4.1 to 5.6) | 3.9 | (3.2 to 4.5) |
| | | Δ Percentage (%) | 23.6 | (19.5 to 27.8) | 16.7 | (13.6 to 19.9) |
| | Rhinosinusitis | IMD=20 | 1.1 | (1.0 to 1.2) | 1.3 | (1.2 to 1.4) |
| | | IMD=1 | 1.2 | (1.1 to 1.4) | 1.3 | (1.2 to 1.5) |
| | | Δ Absolute | -0.1 | (-0.2 to 0.0) | -0.1 | (-0.2 to 0.1) |
| | | Δ Percentage (%) | -7.3 | (-17.0 to 2.8) | -3.9 | (-12.1 to 5.3) |
| | Unspecified URTI | IMD=20 | 12.5 | (12.1 to 13.0) | 13.4 | (13.0 to 13.8) |
| | | IMD=1 | 9.2 | (8.9 to 9.6) | 10.7 | (10.3 to 11.0) |
| | | Δ Absolute | 3.3 | (2.9 to 3.7) | 2.7 | (2.4 to 3.1) |
| | | Δ Percentage (%) | 36.0 | (31.2 to 40.8) | 25.4 | (22.0 to 29.1) |
| | All LRTI | IMD=20 | 25.0 | (24.2 to 25.8) | 25.5 | (24.9 to 26.1) |
| | | IMD=1 | 18.5 | (18.0 to 19.2) | 20.5 | (19.9 to 21.0) |
| | | Δ Absolute | 6.4 | (5.8 to 7.0) | 5.0 | (4.5 to 5.6) |

| | | | | | | |
|---|---|---|---|---|---|---|
| | | Δ Percentage (%) | 34.6 | (30.8 to 38.5) | 24.6 | (21.7 to 27.7) |
| | Bronchitis | IMD=20 | 0.6 | (0.5 to 0.7) | 0.6 | (0.5 to 0.7) |
| | | IMD=1 | 0.4 | (0.3 to 0.4) | 0.4 | (0.3 to 0.5) |
| | | Δ Absolute | 0.2 | (0.1 to 0.3) | 0.2 | (0.1 to 0.2) |
| | | Δ Percentage (%) | 58.1 | (33.3 to 87.1) | 39.4 | (20.4 to 61.1) |
| | Unspecified LRTI | IMD=20 | 24.3 | (23.6 to 25) | 25.3 | (24.7 to 26.0) |
| | | IMD=1 | 18.1 | (17.5 to 18.7) | 20.3 | (19.8 to 20.9) |
| | | Δ Absolute | 6.2 | (5.6 to 6.8) | 5.0 | (4.5 to 5.6) |
| | | Δ Percentage (%) | 34.3 | (30.5 to 38.1) | 24.7 | (21.9 to 27.8) |
| | Pneumonia | IMD=20 | 0.15 | (0.1 to 0.2) | 0.1 | (0.1 to 0.2) |
| | | IMD=1 | 0.18 | (0.1 to 0.3) | 0.2 | (0.1 to 0.3) |
| | | Δ Absolute | -0.03 | (-0.1 to 0.1) | 0.0 | (-0.1 to 0.0) |
| | | Δ Percentage (%) | -16.1 | (-48.3 to 30.5) | -18.6 | (-45.6 to 17.7) |
| Skin | Cellulitis | IMD=20 | 1.2 | (1.1 to 1.4) | 1.4 | (1.2 to 1.5) |
| | | IMD=1 | 1.5 | (1.4 to 1.8) | 1.7 | (1.5 to 1.9) |
| | | Δ Absolute | -0.3 | (-0.5 to -0.2) | -0.3 | (-0.5 to -0.2) |
| | | Δ Percentage (%) | -21.8 | (-31.4 to -10.7) | -20.2 | (-28.2 to -12.1) |
| | Impetigo | IMD=20 | 7.0 | (6.5 to 7.5) | 7.3 | (6.9 to 7.7) |
| | | IMD=1 | 7.9 | (7.4 to 8.4) | 8.1 | (7.6 to 8.6) |
| | | Δ Absolute | -0.9 | (-1.4 to -0.5) | -0.8 | (-1.2 to -0.4) |
| | | Δ Percentage (%) | -11.3 | (-16.6 to -5.9) | -9.6 | (-14.2 to -4.8) |
| Ear | Otitis externa | IMD=20 | 9.2 | (8.8 to 9.7) | 10.4 | (9.9 to 10.8) |
| | | IMD=1 | 10.5 | (10 to 11.1) | 11.3 | (10.8 to 11.9) |
| | | Δ Absolute | -1.3 | (-1.8 to -0.8) | -1.0 | (-1.5 to -0.5) |
| | | Δ Percentage (%) | -12.4 | (-16.5 to -7.9) | -8.7 | (-12.7 to -4.6) |
| | Otitis media | IMD=20 | 27.4 | (26.6 to 28.2) | 26.9 | (26.2 to 27.5) |
| | | IMD=1 | 33.3 | (32.4 to 34.3) | 31.5 | (30.7 to 32.3) |
| | | Δ Absolute | -6.0 | (-6.7 to -5.1) | -4.6 | (-5.3 to -3.9) |
| | | Δ Percentage (%) | -17.9 | (-19.8 to -15.6) | -14.7 | (-16.6 to -12.6) |
| Gastrointestinal | Gastroenteritis | IMD=20 | 0.8 | (0.6 to 0.9) | 0.7 | (0.6 to 0.8) |
| | | IMD=1 | 0.6 | (0.5 to 0.7) | 0.6 | (0.5 to 0.7) |
| | | Δ Absolute | 0.2 | (0.1 to 0.3) | 0.2 | (0.1 to 0.2) |
| | | Δ Percentage (%) | 37.4 | (17.9 to 59.9) | 26.5 | (11.2 to 43.6) |
| Urinary | UTI | IMD=20 | 53.2 | (51.3 to 55.3) | 57.4 | (55.5 to 59.3) |
| | | IMD=1 | 49.5 | (47.6 to 51.3) | 54.5 | (52.7 to 56.3) |
| | | Δ Absolute | 3.8 | (1.8 to 5.8) | 2.9 | (1.0 to 5.0) |
| | | Δ Percentage (%) | 7.6 | (3.5 to 11.9) | 5.4 | (1.9 to 9.3) |

Abbreviations: IMD index of multiple deprivation, URTI upper respiratory tract infections, LRTI lower respiratory tract infections, UTI urinary tract infection, COPD chronic obstructive pulmonary disease, CI confidence interval. “All URTI” includes sore throat, unspecified upper respiratory tract infection, rhinosinusitis whereas “All LRTI” includes bronchitis, unspecified lower respiratory tract infection, pneumonia.

*Table S12 Mean prescribing rates and differences between the most deprived (IMD=20) and least deprived (IMD=1, reference) – complete case vs imputed data – adults*

| | | | Complete case N=4,972,709 | | Imputed data N=7,103,380 | |
|---|---|---|---|---|---|---|
| | Indication | Deprivation level/Δ gap | Mean | (95% CI) | Mean | (95% CI) |

| | | | | | | |
|---|---|---|---|---|---|---|
| Respiratory | Asthma exacerbation | IMD=20 | 7.0 | (6.7 to 7.4) | 6.1 | (5.9 to 6.4) |
| | | IMD=1 | 3.7 | (3.5 to 3.9) | 3.4 | (3.3 to 3.6) |
| | | Δ Absolute | 3.3 | (3.0 to 3.6) | 2.7 | (2.5 to 2.9) |
| | | Δ Percentage (%) | 88.7 | (79.6 to 98) | 79.3 | (72.2 to 86.8) |
| | COPD exacerbation | IMD=20 | 11.7 | (11.2 to 12.3) | 9.1 | (8.6 to 9.5) |
| | | IMD=1 | 2.3 | (2.2 to 2.4) | 2.1 | (2.0 to 2.3) |
| | | Δ Absolute | 9.4 | (8.9 to 9.9) | 6.9 | (6.6 to 7.3) |
| | | Δ Percentage (%) | 409.6 | (384.5 to 436.1) | 323.7 | (305.9 to 342.2) |
| | Cough | IMD=20 | 54.5 | (53.7 to 55.3) | 54.8 | (54.1 to 55.5) |
| | | IMD=1 | 40.0 | (39.4 to 40.6) | 41.4 | (40.9 to 41.9) |
| | | Δ Absolute | 14.5 | (13.8 to 15.1) | 13.4 | (12.8 to 14.0) |
| | | Δ Percentage (%) | 36.1 | (34.2 to 38) | 32.4 | (30.8 to 34.0) |
| | All URTI | IMD=20 | 36.9 | (36.3 to 37.5) | 42.0 | (41.5 to 42.6) |
| | | IMD=1 | 37.6 | (37.1 to 38.2) | 42.5 | (42.0 to 43.0) |
| | | Δ Absolute | -0.7 | (-1.3 to -0.1) | -0.5 | (-1.0 to 0.0) |
| | | Δ Percentage (%) | -1.9 | (-3.4 to -0.4) | -1.2 | (-2.3 to 0.0) |
| | Sore throat | IMD=20 | 11.3 | (11.0 to 11.6) | 13.3 | (13.0 to 13.6) |
| | | IMD=1 | 11.2 | (10.9 to 11.5) | 13.2 | (13.0 to 13.5) |
| | | Δ Absolute | 0.1 | (-0.2 to 0.4) | 0.1 | (-0.2 to 0.3) |
| | | Δ Percentage (%) | 1.1 | (-1.4 to 3.7) | 0.4 | (-1.6 to 2.5) |
| | Rhinosinusitis | IMD=20 | 13.7 | (13.3 to 14.0) | 15.8 | (15.5 to 16.2) |
| | | IMD=1 | 17.1 | (16.7 to 17.5) | 18.8 | (18.4 to 19.2) |
| | | Δ Absolute | -3.4 | (-3.8 to -3.0) | -3.0 | (-3.4 to -2.7) |
| | | Δ Percentage (%) | -20.1 | (-22.0 to -18.0) | -16.0 | (-17.7 to -14.3) |
| | Unspecified URTI | IMD=20 | 12.3 | (12.0 to 12.7) | 12.7 | (12.4 to 13.0) |
| | | IMD=1 | 9.5 | (9.2 to 9.7) | 10.2 | (10.0 to 10.4) |
| | | Δ Absolute | 2.9 | (2.6 to 3.2) | 2.6 | (2.3 to 2.8) |
| | | Δ Percentage (%) | 30.5 | (27.1 to 34.1) | 25.1 | (22.4 to 28.0) |
| | All LRTI | IMD=20 | 87.7 | (86.6 to 88.8) | 83.6 | (82.7 to 84.5) |
| | | IMD=1 | 53.3 | (52.6 to 54.0) | 52.6 | (52.1 to 53.2) |
| | | Δ Absolute | 34.4 | (33.5 to 35.4) | 31.0 | (30.2 to 31.8) |
| | | Δ Percentage (%) | 64.7 | (62.5 to 66.7) | 59.0 | (57.1 to 60.7) |
| | Bronchitis | IMD=20 | 4.1 | (3.8 to 4.3) | 4.3 | (4.1 to 4.5) |
| | | IMD=1 | 3.1 | (3.0 to 3.3) | 3.4 | (3.3 to 3.6) |
| | | Δ Absolute | 1.0 | (0.8 to 1.1) | 0.9 | (0.8 to 1.1) |
| | | Δ Percentage (%) | 30.5 | (23.9 to 37.6) | 26.7 | (21.6 to 32.1) |
| | Unspecified LRTI | IMD=20 | 82.5 | (81.5 to 83.7) | 82.3 | (81.4 to 83.2) |
| | | IMD=1 | 49.4 | (48.8 to 50.1) | 51.8 | (51.2 to 52.3) |
| | | Δ Absolute | 33.1 | (32.2 to 34.0) | 30.6 | (29.8 to 31.4) |
| | | Δ Percentage (%) | 66.9 | (64.8 to 69) | 59.1 | (57.3 to 60.7) |
| | Pneumonia | IMD=20 | 1.1 | (1.0 to 1.3) | 1.4 | (1.3 to 1.5) |
| | | IMD=1 | 0.9 | (0.8 to 1.0) | 1.0 | (0.9 to 1.1) |
| | | Δ Absolute | 0.2 | (0.1 to 0.3) | 0.4 | (0.3 to 0.5) |
| | | Δ Percentage (%) | 23.9 | (12.6 to 36.2) | 40.9 | (30.1 to 52.0) |
| Skin | Cellulitis | IMD=20 | 14.2 | (13.8 to 14.6) | 14.1 | (13.8 to 14.4) |
| | | IMD=1 | 9.8 | (9.5 to 10.1) | 9.9 | (9.6 to 10.1) |
| | | Δ Absolute | 4.5 | (4.1 to 4.8) | 4.2 | (3.9 to 4.5) |
| | | Δ Percentage (%) | 45.6 | (41.6 to 49.8) | 43.0 | (39.5 to 46.3) |

| | | | | | | |
|---|---|---|---|---|---|---|
| | Impetigo | IMD=20 | 1.1 | (1.0 to 1.3) | 1.3 | (1.2 to 1.4) |
| | | IMD=1 | 1.1 | (1.0 to 1.3) | 1.3 | (1.2 to 1.4) |
| | | Δ Absolute | 0.0 | (-0.1 to 0.1) | -0.1 | (-0.2 to 0.1) |
| | | Δ Percentage (%) | 0.1 | (-9.1 to 10.1) | -3.4 | (-10.6 to 4.3) |
| Ear | Otitis externa | IMD=20 | 5.6 | (5.4 to 5.9) | 6.0 | (5.8 to 6.2) |
| | | IMD=1 | 4.3 | (4.1 to 4.5) | 4.8 | (4.6 to 5.0) |
| | | Δ Absolute | 1.3 | (1.1 to 1.6) | 1.2 | (1.0 to 1.4) |
| | | Δ Percentage (%) | 30.7 | (24.8 to 36.8) | 25.5 | (20.4 to 30.1) |
| Gastrointestinal | Gastroenteritis | IMD=20 | 2.9 | (2.7 to 3.1) | 3.0 | (2.9 to 3.2) |
| | | IMD=1 | 2.6 | (2.5 to 2.8) | 2.8 | (2.7 to 2.9) |
| | | Δ Absolute | 0.3 | (0.1 to 0.4) | 0.2 | (0.1 to 0.4) |
| | | Δ Percentage (%) | 10.5 | (4.3 to 16.9) | 8.0 | (3.0 to 13.5) |
| Urinary | UTI | IMD=20 | 102.9 | (101.5 to 104.2) | 112.5 | (111.3 to 113.7) |
| | | IMD=1 | 103.7 | (102.5 to 105) | 113.5 | (112.3 to 114.7) |
| | | Δ Absolute | -0.8 | (-2.2 to 0.6) | -1.0 | (-2.3 to 0.3) |
| | | Δ Percentage (%) | -0.8 | (-2.1 to 0.6) | -0.9 | (-2.0 to 0.2) |
| | Pyelonephritis | IMD=20 | 2.5 | (2.3 to 2.7) | 2.6 | (2.4 to 2.8) |
| | | IMD=1 | 2.4 | (2.3 to 2.6) | 2.5 | (2.4 to 2.7) |
| | | Δ Absolute | 0.1 | (-0.1 to 0.3) | 0.0 | (-0.1 to 0.2) |
| | | Δ Percentage (%) | 2.3 | (-5.5 to 11.3) | 1.8 | (-4.8 to 9.0) |
| | Prostatitis | IMD=20 | 1.7 | (1.5 to 1.9) | 1.8 | (1.6 to 2.1) |
| | | IMD=1 | 3.4 | (3.0 to 3.7) | 3.2 | (2.9 to 3.5) |
| | | Δ Absolute | -1.7 | (-2.0 to -1.4) | -1.4 | (-1.7 to -1.1) |
| | | Δ Percentage (%) | -49.7 | (-56.1 to -43.3) | -42.8 | (-49.1 to -35.9) |

Abbreviations: IMD index of multiple deprivation, URTI upper respiratory tract infections, LRTI lower respiratory tract infections, UTI urinary tract infection, COPD chronic obstructive pulmonary disease, CI confidence interval. “All URTI” includes sore throat, unspecified upper respiratory tract infection, rhinosinusitis whereas “All LRTI” includes bronchitis, unspecified lower respiratory tract infection, pneumonia.
